# Description of the updated nutrition calculation of the Oxford WebQ questionnaire and comparison with the previous version among 207,144 participants in UK Biobank

**DOI:** 10.1101/2020.11.30.20240713

**Authors:** Aurora Perez-Cornago, Zoe Pollard, Heather Young, Marloes van Uden, Colm Andrews, Carmen Piernas, Tim J Key, Angela Mulligan, Marleen Lentjes

## Abstract

**Purpose:** The Oxford WebQ is a web-based 24-hour dietary assessment method which has been used in UK Biobank and other large prospective studies. The food composition table used to calculate nutrient intakes has recently been replaced with the UK Nutrient Databank, which has food composition data closer in time to when participants completed the questionnaire, and new dietary variables were incorporated. Here we describe the updated version of the Oxford WebQ questionnaire nutrient calculation, and compare nutrient intakes with the previous version used.

**Methods:** 207,144 UK Biobank participants completed ≥1 Oxford WebQs, and means and standard deviations of nutrient intakes were averaged for all completed 24-h dietary assessments. Spearman correlations and weighted kappa statistics were used to compare the re-classification and agreement of nutrient intakes between the two versions.

**Results:** 35 new nutrients were incorporated in the updated version. Compared to the previous version, most nutrients were very similar in the updated version except for a few nutrients which showed a difference of >10%: lower with the new version for trans-fat (−20%), and vitamin C (−15%), but higher for retinol (+42%), vitamin D (+26%) and vitamin E (+20%). Most participants were in the same (>60%) or adjacent (>90%) quintile of intake for the two versions. Except for trans-fat (r=0.58, κ=0.42), very high correlations were found between the nutrients calculated using the two versions (r>0.79 and κ>0.60).

**Conclusion:** Small absolute differences in nutrient intakes were observed between the two versions, and the ranking of individuals was minimally affected, except for trans-fat.

## Introduction

Traditional methods to determine dietary intake in large prospective studies, such us paper-based food frequency questionnaires (FFQ) and/or interviewer administered 24-h recalls, are costly and time-consuming. Recently, self-administered online 24-h dietary assessments have been incorporated in some large prospective studies and been shown to facilitate data analyses and decrease the researcher burden, including data entry and data coding, by automatically calculating nutrient intakes (1).

The Oxford WebQ is a fully automated web-based 24-hour dietary assessment tool which seeks information from participants about their consumption of food and drink during the previous 24 hours (2). This online questionnaire has already been used by several large-scale cohort studies, such us the UK Biobank (3) and the Million Women Study (4), as it is easy and quick (∼12 minutes) to self-complete and suitable for repeated use in large-scale prospective studies. Moreover, nutrients are automatically estimated via built-in algorithms and food composition data. Until now, the food composition table (FCT) used for the Oxford WebQ has been the UK McCance and Widdowson’s “The Composition of Foods 6^th^ edition (2002) and its supplements (5-15), of which 550 of 1,200 foods were incorporated into the Oxford WebQ. This FCT has now been replaced by the UK Nutrient Databank (UKNDB) (2013), which provides food composition data measured closer in time to when participants completed the questionnaire in UK Biobank (2009-2012) and contains over 5,600 foods, of which 681 food codes have been incorporated (16, 17). The UKNDB is commissioned by Public Health England as part of the National Diet and Nutrition Survey (NDNS), and is available in electronic format as an integrated dataset, and contains up-to-date nutrient composition data. Data in the UKNDB is very similar to the UK McCance and Widdowson’s FCT but includes a larger range of processed foods and composite dishes. As well as replacing the FCT used to calculate nutrient intakes, we have made other changes such as some changes in portion sizes, personalisation of fats used in cooking, and updating the underlying program code for the nutrient calculation, and new dietary variables such as energy density, and animal and plant fats and proteins, have been incorporated. This paper describes the main changes made to nutrient estimation for the Oxford WebQ questionnaire, and compares the two versions of obtained nutrient intakes in over 200,000 UK Biobank participants.

## Methods

### Study design

UK Biobank includes a total 211,031 participants aged 40-69 years who have completed the Oxford WebQ dietary assessment at least once between 2009 and 2012. Details about the UK Biobank study can be found elsewhere (3). Briefly, participants provided detailed information on a range of sociodemographic, physical, lifestyle, and health-related factors via self-completed touch-screen questionnaires and a computer assisted personal interview at recruitment (3).

The study protocol and information about data access are available online (http://www.ukbiobank.ac.uk/wp-content/uploads/2011/11/UK-Biobank-Protocol.pdf) and in the literature (18).

### Dietary assessment – The Oxford WebQ questionnaire

The Oxford WebQ questionnaire was developed to obtain information on the quantities of up to 206 types of foods and 32 types of drinks consumed over the previous day (24 hours; https://biobank.ctsu.ox.ac.uk/crystal/crystal/docs/DietWebQ.pdf) (2). The quantity of each food or drink consumed is calculated by multiplying the assigned portion size (**Supplementary Table 1**) of each food or beverage by the amount consumed (19). This questionnaire has recently been validated; compared to recovery biomarkers for energy, protein and potassium, and was considered to perform well in approximating true dietary intake (20). This questionnaire also provided similar mean estimates of energy and nutrient intakes when compared with an interviewer administered 24-h dietary recall(2). Further information about the Oxford WebQ can be found here http://www.ceu.ox.ac.uk/research/oxford-webq

**Table 1.** Major changes between the previous (McCance and Widdowson) version and the updated (Nutrient databank + other changes) version.

| Item | Changes made to the updated version (Nutrient databank + other changes) |
| --- | --- |
| Portion size | Some food items had their serving size changed to better reflect what an average portion size would be, taking into account how the question was asked (e.g. Yorkshire pudding). Some portion sizes were revised based on published data (e.g. spreads). Some portion sizes were changed to reflect the state of the food item (e.g. edible part of fruit, or inclusion of liquid for powdered items). These changes can be found in Supplementary Table 2. |
| Milk type | We have now taken into account each milk type beyond fat content, including cholesterol lowering milk, goat's or sheep's milk, powdered milk, rice, oat, almond, coconut milk, fortified soya milk, unfortified soya milk, other milk (e.g. lactose free) as well as skimmed, semi skimmed and whole milk.<br>This is now applied to all hot drinks where milk is added (i.e. tea, coffee, cappuccino, latte, hot chocolate), milk-based sauces, porridge, crepes and pancakes/blinis. |
| Type of fat used in cooking vegetables | Participants were asked to select the type of fat/oil, if any, they use in the cooking, and a total 40 different types of fat/oils were available. We have now added an amount of fat/oils in certain vegetables such as onion, mushroom, mixed veg, peppers, courgette, leek, parsnip, veg other and mashed potato which are likely to be cooked with oils/fats. These fats/oils include:<br>Butters, spreadable butters, hard margarine, lard, dairy spreads, polyunsaturated margarines, cholesterol lowering margarines, olive oil-based spreads, soya spreads, olive oil, rapeseed oil, sunflower oil, vegetable oil. |
| Gluten free versions | We have added a gluten free version where available (e.g. for baguettes, bread rolls, sliced bread, and pasta). |
| Powdered milk | A water code was added to powdered milk codes so the food volume fits with the way the food is served (important in relation to e.g. energy density). |
| 'Other' items | We studied the free text entered by the UK Biobank participants and where possible mapped the 'other' items against commonly entered foods (i.e. according to the participants' understanding of the questions). Whereas previously, these were mapped against a more generic item or a selection of items which were truly different from the specific items listed due to lack of a suitable food code. |

Further details about these changes can be found in Supplementary Methods and Supplementary Table 1.

For the previous version of calculating nutrient intakes for the Oxford WebQ, the UK McCance and Widdowson’s 6^th^ edition (2002) FCT and its supplements were used(2). The nutrients determined were total energy intake, total protein, total fat, saturated fatty acids (SFA), monounsaturated fatty acids (MUFA), polyunsaturated fatty acids (PUFA), cholesterol, carbohydrates, total sugars, fibre, alcohol, calcium, iron, magnesium, potassium, carotene, vitamin B6, vitamin B12, vitamin C, vitamin D and vitamin E. Details about the nutrient calculation can be found in **Supplementary Table 2**. Trans fatty acids (TFA) and retinol in the previous version of the nutrient calculation were excluded since there were multiple food codes with missing values; for the purpose of comparison, illustration of the consequences of missing data, and because TFA have a public health impact, we are however presenting the results from the previous calculation here.

**Table 2.** Comparison of total energy and nutrient intake between previous (McCance & Widdowson) and updated (Nutrient databank + other updates) datasources in 207,144 participants from UK Biobank.

| Nutrient | Previous version: McCance & Widdowson |  |  |  | Updated version: Nutrient databank |  |  |  | Mean difference<sup>1</sup> | Percentage change, %<sup>2</sup> |
| --- | --- | --- | --- | --- | --- | --- | --- | --- | --- | --- |
|  | Mean (SD) | Median | 5<sup>th</sup> percentile | 95<sup>th</sup> percentile | Mean (SD) | Median | 5<sup>th</sup> percentile | 95<sup>th</sup> percentile |  |  |
| Energy (kJ/day) | 8,675 (2,238) | 8,479 | 5,311 | 12,708 | 8,547 (2,204) | 8,350 | 5,250 | 12,519 | -128.6 | -1.48 |
| Protein (g/day) | 81.3 (23.3) | 79.6 | 46.4 | 121.3 | 80.1 (22.9) | 78.30 | 46.07 | 119.72 | -1.18 | -1.45 |
| Total fat (g/day) | 76.5 (27.5) | 73.7 | 36.6 | 126.0 | 72.1 (26.3) | 69.27 | 34.27 | 119.49 | -4.38 | -5.72 |
| SFA (g/day) | 29.2 (11.7) | 27.8 | 12.7 | 50.7 | 26.7 (11.2) | 25.22 | 11.03 | 47.19 | -2.57 | -8.80 |
| PUFA (g/day) | 14.1 (6.88) | 13.1 | 5.1 | 26.8 |  |  |  |  |  |  |
| TFA (g/day) | 1.47 (0.794) | 1.34 | 0.43 | 2.93 | 1.20 (0.649) | 1.08 | 0.34 | 2.36 | -0.29 | -19.8 |
| Carbohydrates (g/day) | 249 (74.4) | 243 | 139 | 381 | 252 (72.7) | 246 | 143.30 | 380.44 | 2.45 | 0.98 |
| Total sugars (g/day) | 119 (45.5) | 113 | 54 | 201 | 124 (46.3) | 119 | 58.32 | 207.25 | 5.44 | 4.59 |
| Englyst fibre (g/day) | 16.3 (6.34) | 15.6 | 7.3 | 27.5 | 17.7 (6.40) | 17.11 | 8.44 | 28.98 | 1.42 | 8.70 |
| Alcohol (g/day) | 16.1 (21.0) | 8.7 | 0.0 | 58.5 | 16.9 (21.7) | 9.05 | 0.00 | 59.91 | 0.74 | 4.62 |
| Calcium (mg/day) | 959 (329) | 923 | 492 | 1,545 | 975 (325) | 942 | 509 | 1,550 | 16.33 | 1.70 |
| Iron (mg/day) | 13.6 (4.14) | 13.2 | 7.3 | 20.8 | 12.3 (3.60) | 11.98 | 6.85 | 18.55 | -1.29 | -9.55 |
| Magnesium (mg/day) | 343 (95.5) | 335 | 203 | 513 | 330 (88.4) | 323 | 199 | 486 | -13.03 | -3.80 |
| Potassium (mg/day) | 3,695 (1,064) | 3,609 | 2,121 | 5,553 | 3,640 (1,007) | 3,571 | 2,115 | 5,384 | -55.20 | -1.49 |
| Retinol (µg/day) | 323 (169) | 300 | 89 | 641 | 461 (884) | 305 | 89 | 880 | 137.2 | 42.0 |
| Total carotene (µg/day) | 3,091 (2,583) | 2,522 | 316 | 7,876 | 2,959 (2,791) | 2,095 | 334 | 8,063 | -131.8 | -4.26 |
| Folate (µg/day) | 301 (107.2) | 287 | 152 | 496 | 310 (104) | 300 | 160 | 493 | 9.79 | 3.26 |
| Vitamin B6 (mg/day) | 2.16 (0.691) | 2.11 | 1.13 | 3.36 | 2.05 (0.664) | 1.99 | 1.09 | 3.23 | -0.11 | -5.07 |
| Vitamin B12 (µg/day) | 6.47 (4.49) | 5.39 | 1.64 | 14.90 | 6.11 (3.25) | 5.56 | 2.21 | 11.66 | -0.36 | -5.61 |
| Vitamin C (mg/day) | 150 (102) | 131 | 30 | 338 | 127 (76.5) | 115 | 29 | 266 | -23.35 | -15.53 |
| Vitamin D (µg/day) | 2.85 (2.71) | 2.02 | 0.34 | 8.92 | 3.60 (2.866) | 2.83 | 0.66 | 9.45 | 0.75 | 26.24 |
| Vitamin E (mg/day) | 9.01 (3.98) | 8.47 | 3.60 | 16.27 | 10.8 (4.26) | 10.30 | 4.85 | 18.54 | 1.80 | 19.99 |

All mean differences were statistically significant from zero when using paired t-tests or Wilcoxon's rank sum test (P<0.05).
<sup>1</sup> Calculated as the difference of the mean (UK Nutrient Databank - McCance & Widdowson).
<sup>2</sup> Calculated as the difference of the mean (UK Nutrient Databank - McCance & Widdowson) divided by their mean and multiplied by 100.
Abbreviations: PUFA, polyunsaturated fatty acids; SFA, saturated fatty acids; TFA, trans fatty acids.

For the updated version of the nutrient calculation of the Oxford WebQ, nutrient intakes were calculated using the UKNDB FCT from survey year 6, which includes FCT for years 2012-2013 and 2013-2014. Moreover, changes in allocated portion sizes, personalisation of milk types and fats used in cooking, gluten free versions and the underlying code for nutrient calculation were revised and updated (details in **Supplementary Methods, Table 1**, and **Supplementary Tables 2 and 3**). Except for total PUFA, all the nutrients available in the previous version are also available in the UKNDB (and total PUFA can be calculated by adding n-3 and n-6 PUFA). Moreover, the following further dietary variables are now available: energy density, animal protein, plant protein, animal fat, plant fat, MUFA, n-3 PUFA, n-6 PUFA, free sugars, non-free sugars, non-milk extrinsic sugars, intrinsic and milk sugars, fructose, glucose, sucrose, lactose, maltose, other sugars, alpha-carotene, beta-carotene, beta cryptoxanthin, vitamin a (retinol equivalents), biotin, chloride, copper, haem iron, non-haem iron, iodine, manganese, sodium, niacin equivalent, pantothenic acid, selenium, total nitrogen and zinc.

**Table 3.**
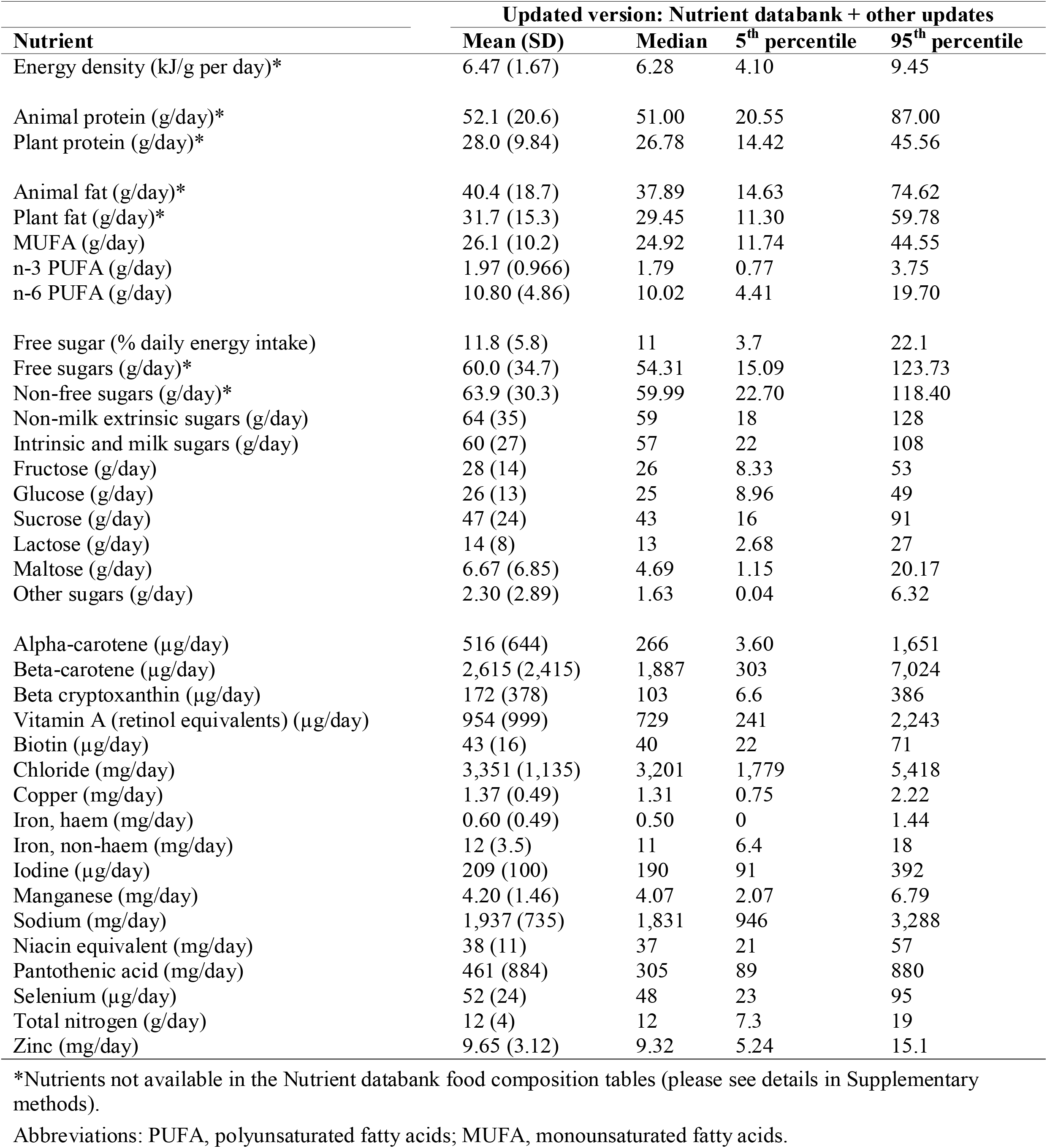
New nutrients incorporated in the updated version (Nutrient databank + other updates) data source in 207,144 participants from UK Biobank.

### Participants

A subsample of UK Biobank participants recruited towards the end of the recruitment period (from April 2009 to September 2010) was invited to complete the Oxford WebQ questionnaire. Moreover, those who provided email addresses were invited to complete the Oxford WebQ a total of four times every 3-4 months on variable days of the week during the follow-up period (online cycle 1, February 2011 to April 2011; online cycle 2, June 2011 to September 2011; online cycle 3, October 2011 to December 2011; online cycle 4, April 2012 to June 2012). 24-h dietary assessments with extreme energy intakes (men: <3,347 or >17,573 kJ/d or <800 or >4200 kcal/d); women: <2,092 or >14,644 kJ/d or <600 or >3500 kcal/d)(21) as calculated with either version of the FCT, were excluded. For this reason, 3887 participants were excluded because they did not have a valid WebQ. In this analysis, we are not interested in usual intakes for individuals but in comparing the estimates of intakes of the participants in the completed 24-h dietary assessments, therefore we have not excluded participants with only one dietary assessment. However, researchers using this dietary assessment tool for diet-disease associations are advised to use at least two 24-h dietary assessments(but more if possible), since intakes from one 24-h dietary assessment are unlikely to reflect usual intakes(20). A total of 207,144 (out of 211,031, 98%) participants were included in this study.

### Statistical analyses

The WebQ results were averaged for all completed 24-h dietary questionnaire for each participant. Means, standard deviations (SDs), and the 5^th^ and 95^th^ percentiles of nutrient intakes are given. The differences and percentage difference (see equation) in nutrient intakes between the previous and the updated version of the nutrient calculation were determined, and means were compared using paired t-tests or Wilcoxon’s rank sum test, depending on the normality of the distribution.

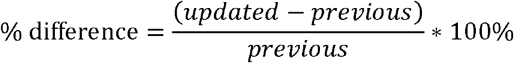

The Spearman correlations of the nutrient data were calculated. Participants were divided into fifths of intake for each nutrient in the two versions of the nutrient calculation and weighted kappa statistics and the percentage of participants who were categorised into the same or adjacent fifth were calculated, since most prospective studies on diet and disease risk examine associations by comparing disease incidence in categories of the dietary factor of interest. Weighted kappas should be interpreted as follows: values ≤ 0 indicates no agreement, 0.01–0.20 as none to slight, 0.21–0.40 as fair, 0.41– 0.60 as moderate, 0.61–0.80 as substantial, and 0.81–1.00 as almost perfect agreement (22).

All analyses were conducted using the STATA statistical software package version 14 (Stata Corporation, College Station, TX, USA).

## Results

The mean age at recruitment was 56 years (SD=8) and 55% were women. Participants completed on average 2.14 (SD=1.16) 24h dietary assessments. **Table 2** shows the mean, median, percentiles, and mean differences of energy and nutrient intakes in the two versions. There were small but significant differences (likely due to the large sample size) in the mean nutrient intakes between the existing version and the updated version. Compared to the previous version, intakes in the updated version were >10% different for the following nutrients: lower for TFA (−20%), vitamin C (−15%) and iron (−9.5%), but higher for retinol (+42%), vitamin D (+26%) and vitamin E (+20%). SFA and TFA intakes provided 12.4% and 0.63% from total energy intake in the previous version of the nutrient calculation, while they provided 11.6% and 0.52% respectively in the updated version.

A total of 35 new nutrients and exposures of interest were available in the UKNDB, and intakes of these nutrients in this population are displayed in **Table 3**.

**Table 4** shows the correlations and the strengths of agreement on ranking nutrient intakes between the previous and the updated version. Except for TFA (r=0.58) and some of the fat-soluble vitamins, high correlations (r>0.90) were found between nutrients calculated using the two versions: energy (r=0.96), protein (r=0.97), total fat (r=0.95), carbohydrates (r=0.95), saturated fat (r=0.91), total sugars (r=0.96), and fibre (r=0.94), with the strongest correlation being for alcohol intake (r=0.99). The percentage of agreement between the two versions was generally good, with the majority of the nutrients classified into the same or adjacent fifth ranging from 90.7 % for retinol (κ= 0.64) to 99.3% for protein (κ= 0.88); however, the percentage agreement was lower for TFA (76.3 %, κ= 0.42), and slightly lower for vitamin E (88.1 %, κ= 0.60) and vitamin B6 (89.7 %, κ= 0.63). The full list of nutrients and the categorization of participants into fifths based on the previous and the updated version is shown in **Tables 5 and 6**.

**Table 4.**
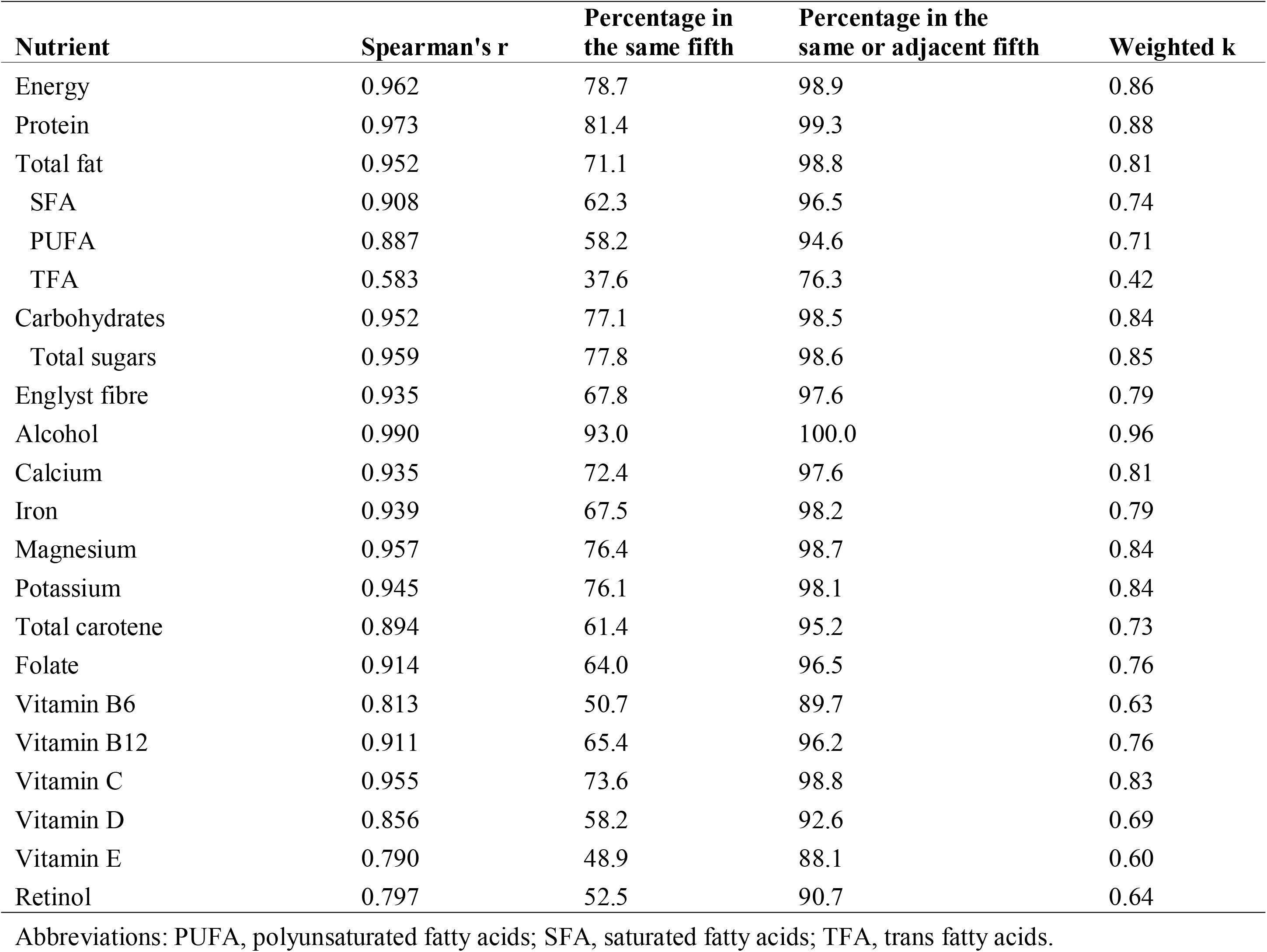
Comparison of total energy and nutrient intake between previous (McCance & Widdowson) and updated (Nutrient databank + other updates) in 207,144 participants from UK Biobank.

**Table 5.** Dietary intakes of energy, macronutrients and fibre by fifths, shaded cells depict participants categorised into the same (dark shading) or adjacent (light shading) quintile using the previous (McCance and Widdowson) and the updated (Nutrient databank + other updates).

| McCance & Widdowson |  | Nutrient databank |  |  |  |  |
| --- | --- | --- | --- | --- | --- | --- |
|  |  | Q1 | Q2 | Q3 | Q4 | Q5 |
| Energy | Q1 | 36733 | 4665 | 31 | 0 | 0 |
|  | Q2 | 3993 | 30595 | 6767 | 74 | 0 |
|  | Q3 | 310 | 5368 | 28870 | 6857 | 24 |
|  | Q4 | 216 | 502 | 5198 | 30468 | 5045 |
|  | Q5 | 177 | 299 | 563 | 4030 | 36359 |
| Protein | Q1 | 37372 | 3993 | 60 | 8 | 1 |
|  | Q2 | 3644 | 31928 | 5707 | 147 | 9 |
|  | Q3 | 260 | 5015 | 30318 | 5738 | 87 |
|  | Q4 | 115 | 373 | 5032 | 31775 | 4134 |
|  | Q5 | 38 | 120 | 312 | 3761 | 37197 |
| Total fat | Q1 | 34662 | 6470 | 285 | 12 | 1 |
|  | Q2 | 6334 | 26253 | 8291 | 542 | 12 |
|  | Q3 | 352 | 8115 | 24532 | 8139 | 288 |
|  | Q4 | 72 | 545 | 8039 | 26702 | 6072 |
|  | Q5 | 9 | 46 | 282 | 6034 | 35055 |
| SFA | Q1 | 31964 | 8772 | 695 | 24 | 1 |
|  | Q2 | 7780 | 21682 | 10825 | 1145 | 23 |
|  | Q3 | 1181 | 8997 | 20100 | 10583 | 543 |
|  | Q4 | 322 | 1613 | 8728 | 22546 | 8196 |
|  | Q5 | 182 | 365 | 1081 | 7131 | 32665 |
| PUFA | Q1 | 32247 | 7891 | 1155 | 162 | 22 |
|  | Q2 | 7920 | 20889 | 10325 | 2121 | 161 |
|  | Q3 | 1032 | 9851 | 18163 | 10885 | 1527 |
|  | Q4 | 230 | 2640 | 9728 | 19162 | 9605 |
|  | Q5 | 0 | 158 | 2058 | 9099 | 30113 |
| Trans fat | Q1 | 21416 | 11056 | 5480 | 2528 | 952 |
|  | Q2 | 9227 | 12235 | 11027 | 6678 | 2259 |
|  | Q3 | 5256 | 8651 | 10975 | 11114 | 5433 |
|  | Q4 | 3371 | 5940 | 8458 | 12098 | 11562 |
|  | Q5 | 2159 | 3547 | 5489 | 9011 | 21222 |
| Carbohydrates | Q1 | 36743 | 4619 | 68 | 1 | 0 |
|  | Q2 | 3877 | 30279 | 7083 | 190 | 2 |
|  | Q3 | 323 | 5400 | 28052 | 7553 | 101 |
|  | Q4 | 171 | 643 | 5414 | 29232 | 5966 |
|  | Q5 | 315 | 488 | 812 | 4453 | 35359 |
| Total sugars | Q1 | 36650 | 4658 | 125 | 0 | 0 |
|  | Q2 | 4105 | 30422 | 6677 | 222 | 0 |
|  | Q3 | 384 | 5360 | 28604 | 7003 | 84 |
|  | Q4 | 181 | 725 | 5206 | 29710 | 5600 |
|  | Q5 | 109 | 264 | 817 | 4494 | 35744 |
| Fibre | Q1 | 34324 | 5742 | 1284 | 124 | 7 |
|  | Q2 | 6974 | 25127 | 7195 | 1974 | 121 |
|  | Q3 | 130 | 10274 | 22479 | 7607 | 941 |
|  | Q4 | 1 | 286 | 10342 | 24472 | 6313 |
|  | Q5 | 0 | 0 | 129 | 7252 | 34046 |
| Alcohol | Q1 | 60178 | 10935 | 0 | 0 | 0 |
|  | Q2 | 0 | 11793 | 259 | 0 | 0 |
|  | Q3 | 0 | 139 | 40345 | 638 | 0 |
|  | Q4 | 0 | 0 | 638 | 39902 | 1086 |
|  | Q5 | 0 | 0 | 0 | 894 | 40337 |

**Table 6.**
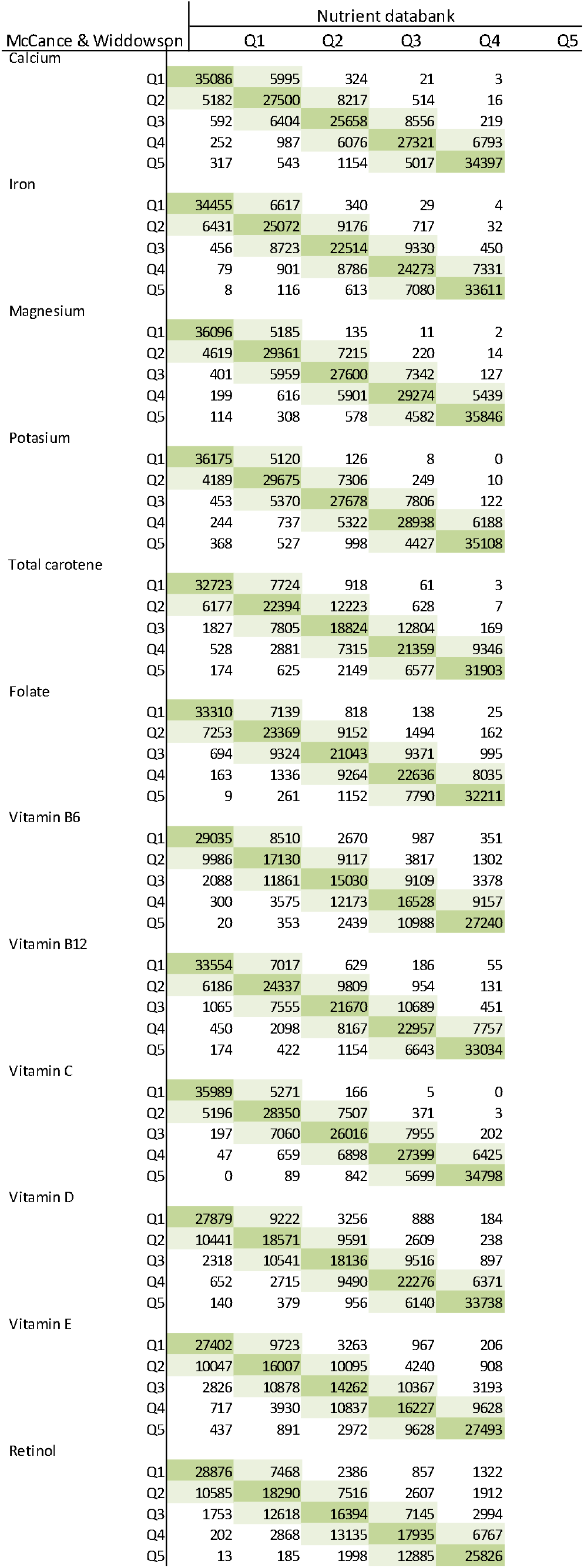
Dietary intakes of micronutrients by fifths, shaded cells depict participants categorised into the same (dark shading) or adjacent (light shading) quintile using the previous (McCance and Widdowson) and the updated (Nutrient databank + other updates).

| McCance & Widdowson |  | Nutrient databank |  |  |  |  |
| --- | --- | --- | --- | --- | --- | --- |
|  |  | Q1 | Q2 | Q3 | Q4 | Q5 |
| Calcium | Q1 | 35086 | 5995 | 324 | 21 | 3 |
|  | Q2 | 5182 | 27500 | 8217 | 514 | 16 |
|  | Q3 | 592 | 6404 | 25658 | 8556 | 219 |
|  | Q4 | 252 | 987 | 6076 | 27321 | 6793 |
|  | Q5 | 317 | 543 | 1154 | 5017 | 34397 |
| Iron | Q1 | 34455 | 6617 | 340 | 29 | 4 |
|  | Q2 | 6431 | 25072 | 9176 | 717 | 32 |
|  | Q3 | 456 | 8723 | 22514 | 9330 | 450 |
|  | Q4 | 79 | 901 | 8786 | 24273 | 7331 |
|  | Q5 | 8 | 116 | 613 | 7080 | 33611 |
| Magnesium | Q1 | 36096 | 5185 | 135 | 11 | 2 |
|  | Q2 | 4619 | 29361 | 7215 | 220 | 14 |
|  | Q3 | 401 | 5959 | 27600 | 7342 | 127 |
|  | Q4 | 199 | 616 | 5901 | 29274 | 5439 |
|  | Q5 | 114 | 308 | 578 | 4582 | 35846 |
| Potassium | Q1 | 36175 | 5120 | 126 | 8 | 0 |
|  | Q2 | 4189 | 29675 | 7306 | 249 | 10 |
|  | Q3 | 453 | 5370 | 27678 | 7806 | 122 |
|  | Q4 | 244 | 737 | 5322 | 28938 | 6188 |
|  | Q5 | 368 | 527 | 998 | 4427 | 35108 |
| Total carotene | Q1 | 32723 | 7724 | 918 | 61 | 3 |
|  | Q2 | 6177 | 22394 | 12223 | 628 | 7 |
|  | Q3 | 1827 | 7805 | 18824 | 12804 | 169 |
|  | Q4 | 528 | 2881 | 7315 | 21359 | 9346 |
|  | Q5 | 174 | 625 | 2149 | 6577 | 31903 |
| Folate | Q1 | 33310 | 7139 | 818 | 138 | 25 |
|  | Q2 | 7253 | 23369 | 9152 | 1494 | 162 |
|  | Q3 | 694 | 9324 | 21043 | 9371 | 995 |
|  | Q4 | 163 | 1336 | 9264 | 22636 | 8035 |
|  | Q5 | 9 | 261 | 1152 | 7790 | 32211 |
| Vitamin B6 | Q1 | 29035 | 8510 | 2670 | 987 | 351 |
|  | Q2 | 9986 | 17130 | 9117 | 3817 | 1302 |
|  | Q3 | 2088 | 11861 | 15030 | 9109 | 3378 |
|  | Q4 | 300 | 3575 | 12173 | 16528 | 9157 |
|  | Q5 | 20 | 353 | 2439 | 10988 | 27240 |
| Vitamin B12 | Q1 | 33554 | 7017 | 629 | 186 | 55 |
|  | Q2 | 6186 | 24337 | 9809 | 954 | 131 |
|  | Q3 | 1065 | 7555 | 21670 | 10689 | 451 |
|  | Q4 | 450 | 2098 | 8167 | 22957 | 7757 |
|  | Q5 | 174 | 422 | 1154 | 6643 | 33034 |
| Vitamin C | Q1 | 35989 | 5271 | 166 | 5 | 0 |
|  | Q2 | 5196 | 28350 | 7507 | 371 | 3 |
|  | Q3 | 197 | 7060 | 26016 | 7955 | 202 |
|  | Q4 | 47 | 659 | 6898 | 27399 | 6425 |
|  | Q5 | 0 | 89 | 842 | 5699 | 34798 |
| Vitamin D | Q1 | 27879 | 9222 | 3256 | 888 | 184 |
|  | Q2 | 10441 | 18571 | 9591 | 2609 | 238 |
|  | Q3 | 2318 | 10541 | 18136 | 9516 | 897 |
|  | Q4 | 652 | 2715 | 9490 | 22276 | 6371 |
|  | Q5 | 140 | 379 | 956 | 6140 | 33738 |
| Vitamin E | Q1 | 27402 | 9723 | 3263 | 967 | 206 |
|  | Q2 | 10047 | 16007 | 10095 | 4240 | 908 |
|  | Q3 | 2826 | 10878 | 14262 | 10367 | 3193 |
|  | Q4 | 717 | 3930 | 10837 | 16227 | 9628 |
|  | Q5 | 437 | 891 | 2972 | 9628 | 27493 |
| Retinol | Q1 | 28876 | 7468 | 2386 | 857 | 1322 |
|  | Q2 | 10585 | 18290 | 7516 | 2607 | 1912 |
|  | Q3 | 1753 | 12618 | 16394 | 7145 | 2994 |
|  | Q4 | 202 | 2868 | 13135 | 17935 | 6767 |
|  | Q5 | 13 | 185 | 1998 | 12885 | 25826 |

## Discussion

We have described the updated version of the Oxford WebQ 24-h dietary assessment and compared it with the previous version of this questionnaire among participants in UK Biobank. In general, small absolute mean differences in nutrient intakes between the two versions were observed, and the ranking of individuals was minimally affected for most nutrients. The only substantial differences were observed for TFA and vitamin C, for which intakes in the updated version were lower and for retinol, vitamin D and E, for which intakes were higher. We have incorporated new dietary variables, which will allow researchers to assess whether they are related to non-communicable diseases. Also, with this update, we have made it easier for future users to continue this updating process using future releases of the UKNDB.

After categorising the nutrient intakes, there was very high agreement between the two versions for total energy intake and macronutrients. The closest agreement was observed for alcohol intake, for which 100% of the participants were in the same or adjacent fifth, followed by total protein. As expected, intakes of TFA were lower in the updated version of the nutrient calculation and there was moderate agreement with the previous version. Most TFA in the diet are produced when converting vegetable oils into semi-solid fats during the process of partial hydrogenation. TFA are well established risk factors for cardiovascular disease (23), and the food industry has voluntarily reduced or eliminated some artificial TFA in processed foods in the UK in the last fifteen years (24). The previous version used FCT in which nutrient content was published from foods chemically analysed up to 2002 (including analytic data pre-dating the publication date), and therefore, the ‘true’ TFA intake in 2009-2012, when the participants completed the Oxford WebQ, was likely lower (25). This previous version also had substantial missing data for TFA, and for this reason this nutrient was not released in UK Biobank. The lower mean TFA intake in the updated version is likely an underestimated difference due to previous missing data on TFA, and also due to food reformulation over time and/or the different imputations of TFA between the two FCT versions of the nutrient calculation. The main sources of TFA in the previous version were likely to be fat spreads and desserts and biscuits, while in the updated version they are likely to be mainly naturally occurring TFA in food produced from ruminant animals. Intakes of TFA are below the dietary reference value of <2% of total energy, and values are consistent with those reported by the UK NDNS (26).

Intakes of SFAs were also lower in the updated version of the nutrient calculation, but with high agreement in ranking between the two versions. One of the major contributors to SFA in this cohort is dairy fat spread, and therefore it is possible that the decrease in SFA may be due to the decrease of 20 to 60% in the portion sizes allocated for some spreads in the revised version (e.g. spreads on crispbreads, slices of bread, bread rolls, and oatcakes, see supplement for more details).

There were also differences in vitamin intakes between the two versions. Vitamin C intake was on average 17% lower in the updated version compared to the previous version. When vitamin C intake was divided into fifths, the majority of the participants remained classified in the same or an adjacent category. The decrease in vitamin C may be due to fruit juice, which is the largest source of vitamin C in this cohort and in which the previous version of the questionnaire had a concentrated fruit juice code not sufficiently diluted with water. On the other hand, we observed an increase in the intake estimates of retinol and vitamins D and E, although there was substantial agreement between the two versions when these nutrients were categorised. This may be due to the incorporation of fats used when cooking in this updated version, which were mainly vegetable oils; increases in vitamin D may also have occurred due to increases in food fortification, although no fortified foods were preferred when allocating food codes to the WebQ items. Moreover, differences in micronutrient content between the different FCTs are to be expected even if these FCT were created from similar sources; this may be due to for example food reformulation, re-analysis of foods resulting in differences due to storage conditions, fortification or season when the food was sampled. Lastly, imputation of missing values in the UKNDB may have contributed to changes in the nutrient intakes observed (27).

Among the new dietary variables that have been incorporated in this updated version, are MUFAs, n-3 and n-6 PUFAs. The UKNDB does not have information on total essential PUFAs, but n-3 and n-6 fatty acids account for the vast majority of PUFAs in the diet; therefore, researchers using this resource could sum these two fatty acids as a proxy of total PUFA. Other dietary variables that have been incorporated are animal and plant fat and protein, and free sugars. The mean intake of free sugars in this population is slightly above the recommended value of <10% of total energy intake by the World Health Organization (28).

This study has some strengths and limitations. The updated FCT has over three times more food codes than the previous one, which allowed for a better matching between reported food intakes and nutrient composition. This updated version of the nutrient calculation was developed to improve accuracy -and in very few cases also validity (where the original food code did not accurately match the food description in the WebQ) of the dietary intakes of the participants when they completed the questionnaire, and so it is expected to decrease measurement error. Non-differential misclassification of dietary intakes may attenuate the relationship in diet-disease associations in prospective studies (29). However, it should be emphasized that, as in all questionnaire-based assessments of dietary intake, there will be some measurement error, especially systematic bias due to underreporting (20).

In conclusion, we have described an updated version of the nutrient calculation of the Oxford WebQ 24-h dietary assessment and compared it with the previous version. Small absolute group differences in nutrient intakes between the two versions were observed and the ranking of individuals was minimally affected for most nutrients. The greatest differences were observed for TFA and vitamin C, for which intakes in the updated version were lower; and for retinol, vitamin D and E, for which the reported intakes were higher. This updated version of the nutrient calculation was developed to improve accuracy and personalisation of the dietary intakes of the participants and therefore, some reduction in non-differential misclassification in diet-disease associations is expected. This new version of the nutrient calculation and new dietary variables will be returned to UK Biobank.

## Data Availability

UK Biobank is an open access resource. Bona fide researchers can apply to use the UK Biobank data set by registering and applying at http://www.ukbio bank.ac.uk/register-apply.

## Declarations

### Ethics approval

The UK Biobank study was conducted according to the guidelines laid down in the Declaration of Helsinki and approved by the North West Multi-Centre Research Ethics Committee (reference number 06/MRE08/65).

### Consent to participate

All participants gave informed consent to participate and be followed-up through data-linkage at recruitment.

### Availability of data and material

UK Biobank is an open access resource. Bona fide researchers can apply to use the UK Biobank data set by registering and applying at http://www.ukbiobank.ac.uk/register-apply.

## Acknowledgements

The food composition data used in this paper were taken from the National Diet and Nutrition Survey (NDNS) and accessed with kind permission of the UK Data Service. We wish to express our gratitude to the participants and those involved in building the UK Biobank resource. This work has been conducted using the UK Biobank Resource under Application Number 24494. We would also like to thank researchers involved in the previous versions of the Oxford WebQ questionnaire.

