## Supplementary material for "Description of the updated nutrition calculation of the Oxford WebQ questionnaire and comparison with the previous version among 207,144 participants in UK Biobank": Sup table 1

**Supplementary Table 1.** Variable description, portion sizes, and differences between each food item in the WebQ between version 1 (McCance and Widdowson) and version 2 (Nutrient databank + other changes).

| | McCance and Widdowson's | | Nutrient databank + other changes | | | |
|---|---|---|---|---|---|---|
| **Variable description** | **Food item** | **Portion size** | **Food item** | **Portion size** | **Portion diff%** | **Description of differences with previous version** |
| Salt added to food | add_salt | 4 | add_salt | 1 | -75% | More realistic portion size for daily amount of 'salt at the table'; qualitative measure, this is the same quantity for everyone responding 'yes'. |
| Alcoholic drinks: beer, lager or cider | alcohol_beercider | 574 | alcohol_beercider | 574 | 0% | |
| Alcoholic drinks: other drinks (e.g. cream liqueurs, medium or high strength liqueurs) | alcohol_other | 25 | alcohol_other | 25 | 0% | Mapped to liqueurs |
| Alcoholic drinks: spirits | alcohol_spirits | 23 | alcohol_spirits | 23 | 0% | |
| Alcoholic drinks: sherry, port, fortified wine | alcohol_wine_fort | 50 | alcohol_wine_fort | 50 | 0% | |
| Alcoholic drinks: red wine | alcohol_wine_red_large | 250 | alcohol_wine_red_large | 250 | 0% | |
| Alcoholic drinks: red wine | alcohol_wine_red_med | 175 | alcohol_wine_red_med | 175 | 0% | |
| Alcoholic drinks: red wine | alcohol_wine_red_small | 125 | alcohol_wine_red_small | 125 | 0% | |
| Alcoholic drinks: rose wine, sparkling rose wine | alcohol_wine_rose_large | 250 | alcohol_wine_rose_large | 250 | 0% | |
| Alcoholic drinks: rose wine, sparkling rose wine | alcohol_wine_rose_med | 175 | alcohol_wine_rose_med | 175 | 0% | |
| Alcoholic drinks: rose wine, sparkling rose wine | alcohol_wine_rose_small | 125 | alcohol_wine_rose_small | 125 | 0% | |
| Alcoholic drinks: white wine, sparkling white wine | alcohol_wine_white_large | 250 | alcohol_wine_white_large | 250 | 0% | |
| Alcoholic drinks: white wine, sparkling white wine | alcohol_wine_white_med | 175 | alcohol_wine_white_med | 175 | 0% | |
| Alcoholic drinks: white wine, sparkling white wine | alcohol_wine_white_small | 125 | alcohol_wine_white_small | 125 | 0% | |
| Biscuits or cookies with chocolate or half coated in chocolate | biscuit_choc | 17 | biscuit_choc | 17 | 0% | |
| Fully coated chocolate biscuits, can include cream or wafer filling | biscuit_choccov | 24 | biscuit_choccov | 24 | 0% | |
| Fully coated chocolate biscuits, can include cream or wafer filling | | | biscuit_choccov_gf | 24 | - | Gluten free version added to the updated version (however no gluten free code available at the time so the non-gluten free code was used) |
| Biscuits or cookies with chocolate or half coated in chocolate | | | biscuit_choc_gf | 17 | - | Gluten free version added to the updated version (however no gluten free code available at the time so the non-gluten free code was used) |
| Biscuits sweet e.g digestive, gingernut, shortbread | biscuit_sweet | 17 | biscuit_sweet | 17 | 0% | |
| Gluten free biscuits sweet e.g digestive, gingernut, shortbread | | | biscuit_sweet_gf | 17 | - | Gluten free version added to the updated version |
| Gluten free baguette, ciabatta, panini, sub: brown or with added fibre | | | bread_baguette_gf_nonwhite | 95 | - | Gluten free version added to the updated version |
| Gluten free baguette, ciabatta, panini, sub: not specified type so includes white, brown or with added fibre | | | bread_baguette_gf_unanswered | 95 | - | Gluten free version added to the updated version |
| Gluten free baguette, ciabatta, panini, sub: white | | | bread_baguette_gf_white | 95 | - | Gluten free version added to the updated version |
| Baguette, ciabatta, panini, sub: granary, brown, high fibre white, 50/50 white and wholemeal | bread_baguette_mixed | 95 | bread_baguette_mixed | 95 | 0% | |
| Baguette, ciabatta, panini, sub: wheatgerm, rye, white sliced | bread_baguette_other | 95 | bread_baguette_other | 95 | 0% | |
| Seeds for seeded bread: e.g sesame, sunflower, poppy | bread_baguette_seeded | 3 | bread_baguette_seeded | 3 | 0% | |
| Fat that is spread (medium amount) onto baguette: ticked butter but not specified amount of % fat | bread_baguette_spread_butter_dunno_med | 12 | bread_baguette_spread_butter_dunno_med | 15 | 25% | 125% of the weight on a bread roll; food composition as butter and spreadable butter |
| Fat that is spread (thick amount) onto baguette: ticked butter but not specified amount of % fat | bread_baguette_spread_butter_dunno_thick | 15 | bread_baguette_spread_butter_dunno_thick | 18.75 | 25% | 125% of the weight on a bread roll; food composition as butter and spreadable butter |
| Fat that is spread (thin amount) onto baguette: ticked butter but not specified amount of % fat | bread_baguette_spread_butter_dunno_thin | 10 | bread_baguette_spread_butter_dunno_thin | 12.5 | 25% | 125% of the weight on a bread roll; food composition as butter and spreadable butter |
| Fat that is spread (medium amount) onto baguette: ticked butter, normal amount of % fat | bread_baguette_spread_butter_fat_med | 12 | bread_baguette_spread_butter_fat_med | 15 | 25% | 125% of the weight on a bread roll |
| Fat that is spread (thick amount) onto baguette: ticked butter, normal amount of % fat | bread_baguette_spread_butter_fat_thick | 15 | bread_baguette_spread_butter_fat_thick | 18.75 | 25% | 125% of the weight on a bread roll |
| Fat that is spread (thin amount) onto baguette: ticked butter, normal amount of % fat | bread_baguette_spread_butter_fat_thin | 10 | bread_baguette_spread_butter_fat_thin | 12.5 | 25% | 125% of the weight on a bread roll |
| Fat that is spread (medium amount) onto baguette: ticked butter, low fat | bread_baguette_spread_butter_lowfat_med | 12 | bread_baguette_spread_butter_lowfat_med | 15 | 25% | 125% of the weight on a bread roll |
| Fat that is spread (thick amount) onto baguette: ticked butter, low fat | bread_baguette_spread_butter_lowfat_thick | 15 | bread_baguette_spread_butter_lowfat_thick | 18.75 | 25% | 125% of the weight on a bread roll |
| Fat that is spread (thin amount) onto baguette: ticked butter, low fat | bread_baguette_spread_butter_lowfat_thin | 10 | bread_baguette_spread_butter_lowfat_thin | 12.5 | 25% | 125% of the weight on a bread roll |
| Fat that is spread (medium amount) onto baguette: ticked spreadable butter with normal amount of % fat | bread_baguette_spread_butter_spread_fat_med | 12 | bread_baguette_spread_butter_spread_fat_med | 15 | 25% | 125% of the weight on a bread roll |
| Fat that is spread (thick amount) onto baguette: ticked spreadable butter with normal amount of % fat | bread_baguette_spread_butter_spread_fat_thick | 15 | bread_baguette_spread_butter_spread_fat_thick | 18.75 | 25% | 125% of the weight on a bread roll |
| Fat that is spread (thin amount) onto baguette: ticked spreadable butter with normal amount of % fat | bread_baguette_spread_butter_spread_fat_thin | 10 | bread_baguette_spread_butter_spread_fat_thin | 12.5 | 25% | 125% of the weight on a bread roll |
| Fat that is spread (medium amount) onto baguette: ticked spreadable butter, low fat | bread_baguette_spread_butter_spread_lowfat_me | 12 | bread_baguette_spread_butter_spread_lowfat_me | 15 | 25% | 125% of the weight on a bread roll |
| Fat that is spread (thick amount) onto baguette: ticked spreadable butter, low fat | bread_baguette_spread_butter_spread_lowfat_thi | 15 | bread_baguette_spread_butter_spread_lowfat_thi | 18.75 | 25% | 125% of the weight on a bread roll |
| Fat that is spread (thin amount) onto baguette: ticked spreadable butter, low fat | bread_baguette_spread_butter_spread_lowfat_thi | 10 | bread_baguette_spread_butter_spread_lowfat_thi | 12.5 | 25% | 125% of the weight on a bread roll |
| Fat that is spread (medium amount) onto baguette: ticked dairy spread which is also cholesterol lowering e.g Benecol Buttery | bread_baguette_spread_dairy_chol_med | 10 | bread_baguette_spread_dairy_chol_med | 12.5 | 25% | 125% of the weight on a bread roll |
| Fat that is spread (thick amount) onto baguette: ticked dairy spread which is also cholesterol lowering e.g Benecol Buttery | bread_baguette_spread_dairy_chol_thick | 12 | bread_baguette_spread_dairy_chol_thick | 15 | 25% | 125% of the weight on a bread roll |
| Fat that is spread (thin amount) onto baguette: ticked dairy spread which is also cholesterol lowering e.g Benecol Buttery | bread_baguette_spread_dairy_chol_thin | 7 | bread_baguette_spread_dairy_chol_thin | 8.75 | 25% | 125% of the weight on a bread roll |
| Fat that is spread (medium amount) onto baguette: ticked dairy spread but not specified amount of % fat | bread_baguette_spread_dairy_dunno_med | 10 | bread_baguette_spread_dairy_dunno_med | 12.5 | 25% | 125% of the weight on a bread roll; food composition as low fat and reduced fat |
| Fat that is spread (thick amount) onto baguette: ticked dairy spread but not specified amount of % fat | bread_baguette_spread_dairy_dunno_thick | 12 | bread_baguette_spread_dairy_dunno_thick | 15 | 25% | 125% of the weight on a bread roll; food composition as low fat and reduced fat |
| Fat that is spread (thin amount) onto baguette: ticked dairy spread but not specified amount of % fat | bread_baguette_spread_dairy_dunno_thin | 7 | bread_baguette_spread_dairy_dunno_thin | 8.75 | 25% | 125% of the weight on a bread roll; food composition as low fat and reduced fat |
| Fat that is spread (medium amount) onto baguette: ticked dairy spread with normal amount of % fat | bread_baguette_spread_dairy_fat_med | 10 | bread_baguette_spread_dairy_fat_med | 12.5 | 25% | 125% of the weight on a bread roll; normal fat is taken as reduced fat (up to 62% fat) |
| Fat that is spread (thick amount) onto baguette: ticked dairy spread with normal amount of % fat | bread_baguette_spread_dairy_fat_thick | 12 | bread_baguette_spread_dairy_fat_thick | 15 | 25% | 125% of the weight on a bread roll; normal fat is taken as reduced fat (up to 62% fat) |
| Fat that is spread (thin amount) onto baguette: ticked dairy spread with normal amount of % fat | bread_baguette_spread_dairy_fat_thin | 7 | bread_baguette_spread_dairy_fat_thin | 8.75 | 25% | 125% of the weight on a bread roll; normal fat is taken as reduced fat (up to 62% fat) |
| Fat that is spread (medium amount) onto baguette: ticked dairy spread, low fat | bread_baguette_spread_dairy_lowfat_med | 10 | bread_baguette_spread_dairy_lowfat_med | 12.5 | 25% | 125% of the weight on a bread roll |
| Fat that is spread (thick amount) onto baguette: ticked dairy spread, low fat | bread_baguette_spread_dairy_lowfat_thick | 12 | bread_baguette_spread_dairy_lowfat_thick | 15 | 25% | 125% of the weight on a bread roll |
| Fat that is spread (thin amount) onto baguette: ticked dairy spread, low fat | bread_baguette_spread_dairy_lowfat_thin | 7 | bread_baguette_spread_dairy_lowfat_thin | 8.75 | 25% | 125% of the weight on a bread roll |
| Fat that is spread (medium amount) onto baguette: ticked dairy spread, very low fat | bread_baguette_spread_dairy_vlowfat_med | 10 | bread_baguette_spread_dairy_vlowfat_med | 12.5 | 25% | 125% of the weight on a bread roll; food composition as low fat spread |
| Fat that is spread (thick amount) onto baguette: ticked dairy spread, very low fat | bread_baguette_spread_dairy_vlowfat_thick | 12 | bread_baguette_spread_dairy_vlowfat_thick | 15 | 25% | 125% of the weight on a bread roll; food composition as low fat spread |
| Fat that is spread (thin amount) onto baguette: ticked dairy spread, very low fat | bread_baguette_spread_dairy_vlowfat_thin | 7 | bread_baguette_spread_dairy_vlowfat_thin | 8.75 | 25% | 125% of the weight on a bread roll; food composition as low fat spread |
| Fat that is spread (medium amount) onto baguette: not specified type of spread but ticked cholesterol lowering e.g Benecol, Flora pro active | bread_baguette_spread_dunno_chol_med | 10 | bread_baguette_spread_dunno_chol_med | 12.5 | 25% | 125% of the weight on a bread roll |
| Fat that is spread (thick amount) onto baguette: not specified type of spread but ticked cholesterol lowering e.g Benecol, Flora pro active | bread_baguette_spread_dunno_chol_thick | 12 | bread_baguette_spread_dunno_chol_thick | 15 | 25% | 125% of the weight on a bread roll |
| Fat that is spread (thin amount) onto baguette: not specified type of spread but ticked cholesterol lowering e.g Benecol, Flora pro active | bread_baguette_spread_dunno_chol_thin | 7 | bread_baguette_spread_dunno_chol_thin | 8.75 | 25% | 125% of the weight on a bread roll |
| Fat that is spread (medium amount) onto baguette: not specified type of spread or amount of % fat | bread_baguette_spread_dunno_dunno_med | 10 | bread_baguette_spread_dunno_dunno_med | 12.5 | 25% | 125% of the weight on a bread roll; food composition as low fat and reduced fat |
| Fat that is spread (thick amount) onto baguette: not specified type of spread or amount of % fat | bread_baguette_spread_dunno_dunno_thick | 12 | bread_baguette_spread_dunno_dunno_thick | 15 | 25% | 125% of the weight on a bread roll; food composition as low fat and reduced fat |
| Fat that is spread (thin amount) onto baguette: not specified type of spread or amount of % fat | bread_baguette_spread_dunno_dunno_thin | 7 | bread_baguette_spread_dunno_dunno_thin | 8.75 | 25% | 125% of the weight on a bread roll; food composition as low fat and reduced fat |
| Fat that is spread (medium amount) onto baguette: not specified type of spread but ticked normal amount % fat | bread_baguette_spread_dunno_fat_med | 10 | bread_baguette_spread_dunno_fat_med | 12.5 | 25% | 125% of the weight on a bread roll; normal fat is taken as reduced fat (up to 62% fat) |
| Fat that is spread (thick amount) onto baguette: not specified type of spread but ticked normal amount % fat | bread_baguette_spread_dunno_fat_thick | 12 | bread_baguette_spread_dunno_fat_thick | 15 | 25% | 125% of the weight on a bread roll; normal fat is taken as reduced fat (up to 62% fat) |
| Fat that is spread (thin amount) onto baguette: not specified type of spread but ticked normal amount % fat | bread_baguette_spread_dunno_fat_thin | 7 | bread_baguette_spread_dunno_fat_thin | 8.75 | 25% | 125% of the weight on a bread roll; normal fat is taken as reduced fat (up to 62% fat) |
| Fat that is spread (medium amount) onto baguette: not specified type of spread but ticked low fat | bread_baguette_spread_dunno_lowfat_med | 10 | bread_baguette_spread_dunno_lowfat_med | 12.5 | 25% | 125% of the weight on a bread roll |
| Fat that is spread (thick amount) onto baguette: not specified type of spread but ticked low fat | bread_baguette_spread_dunno_lowfat_thick | 12 | bread_baguette_spread_dunno_lowfat_thick | 15 | 25% | 125% of the weight on a bread roll |
| Fat that is spread (thin amount) onto baguette: not specified type of spread but ticked low fat | bread_baguette_spread_dunno_lowfat_thin | 7 | bread_baguette_spread_dunno_lowfat_thin | 8.75 | 25% | 125% of the weight on a bread roll |
| Fat that is spread (medium amount) onto baguette: not specified type of spread but ticked very low fat | bread_baguette_spread_dunno_vlowfat_med | 10 | bread_baguette_spread_dunno_vlowfat_med | 12.5 | 25% | 125% of the weight on a bread roll; food composition as low fat spread |
| Fat that is spread (thick amount) onto baguette: not specified type of spread but ticked very low fat | bread_baguette_spread_dunno_vlowfat_thick | 12 | bread_baguette_spread_dunno_vlowfat_thick | 15 | 25% | 125% of the weight on a bread roll; food composition as low fat spread |
| Fat that is spread (thin amount) onto baguette: not specified type of spread but ticked very low fat | bread_baguette_spread_dunno_vlowfat_thin | 7 | bread_baguette_spread_dunno_vlowfat_thin | 8.75 | 25% | 125% of the weight on a bread roll; food composition as low fat spread |
| Fat that is spread (medium amount) onto baguette: ticked hard margarine (hard block margarine in wrapper) | bread_baguette_spread_hardmarg_med | 12 | bread_baguette_spread_hardmarg_med | 15 | 25% | 125% of the weight on a bread roll, spread weight as butter |
| Fat that is spread (thick amount) onto baguette: ticked hard margarine (hard block margarine in wrapper) | bread_baguette_spread_hardmarg_thick | 15 | bread_baguette_spread_hardmarg_thick | 18.75 | 25% | 125% of the weight on a bread roll, spread weight as butter |
| Fat that is spread (thin amount) onto baguette: ticked hard margarine (hard block margarine in wrapper) | bread_baguette_spread_hardmarg_thin | 10 | bread_baguette_spread_hardmarg_thin | 12.5 | 25% | 125% of the weight on a bread roll, spread weight as butter |
| Fat that is spread (medium amount) onto baguette: ticked cholesterol lowering olive spread e.g Benecol/Flora pro active olive spread | bread_baguette_spread_olive_chol_med | 10 | bread_baguette_spread_olive_chol_med | 12.5 | 25% | 125% of the weight on a bread roll |
| Fat that is spread (thick amount) onto baguette: ticked cholesterol lowering olive spread e.g Benecol/Flora pro active olive spread | bread_baguette_spread_olive_chol_thick | 12 | bread_baguette_spread_olive_chol_thick | 15 | 25% | 125% of the weight on a bread roll |
| Fat that is spread (thin amount) onto baguette: ticked cholesterol lowering olive spread e.g Benecol/Flora pro active olive spread | bread_baguette_spread_olive_chol_thin | 7 | bread_baguette_spread_olive_chol_thin | 8.75 | 25% | 125% of the weight on a bread roll |
| Fat that is spread (medium amount) onto baguette: ticked olive spread but not specified amount of fat | bread_baguette_spread_olive_dunno_med | 10 | bread_baguette_spread_olive_dunno_med | 12.5 | 25% | 125% of the weight on a bread roll; food composition as low fat and reduced fat |
| Fat that is spread (thick amount) onto baguette: ticked olive spread but not specified amount of fat | bread_baguette_spread_olive_dunno_thick | 12 | bread_baguette_spread_olive_dunno_thick | 15 | 25% | 125% of the weight on a bread roll; food composition as low fat and reduced fat |
| Fat that is spread (thin amount) onto baguette: ticked olive spread but not specified amount of fat | bread_baguette_spread_olive_dunno_thin | 7 | bread_baguette_spread_olive_dunno_thin | 8.75 | 25% | 125% of the weight on a bread roll; food composition as low fat and reduced fat |
| Fat that is spread (medium amount) onto baguette: ticked olive spread with normal amount of % fat | bread_baguette_spread_olive_fat_med | 10 | bread_baguette_spread_olive_fat_med | 12.5 | 25% | 125% of the weight on a bread roll; normal fat is taken as reduced fat (up to 62% fat) |
| Fat that is spread (thick amount) onto baguette: ticked olive spread with normal amount of % fat | bread_baguette_spread_olive_fat_thick | 12 | bread_baguette_spread_olive_fat_thick | 15 | 25% | 125% of the weight on a bread roll; normal fat is taken as reduced fat (up to 62% fat) |
| Fat that is spread (thin amount) onto baguette: ticked olive spread with normal amount of % fat | bread_baguette_spread_olive_fat_thin | 7 | bread_baguette_spread_olive_fat_thin | 8.75 | 25% | 125% of the weight on a bread roll; normal fat is taken as reduced fat (up to 62% fat) |
| Fat that is spread (medium amount) onto baguette: ticked olive spread, low fat | bread_baguette_spread_olive_lowfat_med | 10 | bread_baguette_spread_olive_lowfat_med | 12.5 | 25% | 125% of the weight on a bread roll |
| Fat that is spread (thick amount) onto baguette: ticked olive spread, low fat | bread_baguette_spread_olive_lowfat_thick | 12 | bread_baguette_spread_olive_lowfat_thick | 15 | 25% | 125% of the weight on a bread roll |
| Fat that is spread (thin amount) onto baguette: ticked olive spread, low fat | bread_baguette_spread_olive_lowfat_thin | 7 | bread_baguette_spread_olive_lowfat_thin | 8.75 | 25% | 125% of the weight on a bread roll |
| Fat that is spread (medium amount) onto baguette: ticked olive spread, very low fat | bread_baguette_spread_olive_vlowfat_med | 10 | bread_baguette_spread_olive_vlowfat_med | 12.5 | 25% | 125% of the weight on a bread roll; food composition as low fat spread |
| Fat that is spread (thick amount) onto baguette: ticked olive spread, very low fat | bread_baguette_spread_olive_vlowfat_thick | 12 | bread_baguette_spread_olive_vlowfat_thick | 15 | 25% | 125% of the weight on a bread roll; food composition as low fat spread |
| Fat that is spread (thin amount) onto baguette: ticked olive spread, very low fat | bread_baguette_spread_olive_vlowfat_thin | 7 | bread_baguette_spread_olive_vlowfat_thin | 8.75 | 25% | 125% of the weight on a bread roll; food composition as low fat spread |
| Fat that is spread (medium amount) onto baguette: ticked other spread e.g ghee | bread_baguette_spread_other_med | 10 | bread_baguette_spread_other_med | 12.5 | 25% | 125% of the weight on a bread roll |
| Fat that is spread (thick amount) onto baguette: ticked other spread e.g ghee | bread_baguette_spread_other_thick | 12 | bread_baguette_spread_other_thick | 15 | 25% | 125% of the weight on a bread roll |
| Fat that is spread (thin amount) onto baguette: ticked other spread e.g ghee | bread_baguette_spread_other_thin | 7 | bread_baguette_spread_other_thin | 8.75 | 25% | 125% of the weight on a bread roll |
| Fat that is spread (medium amount) onto baguette: ticked polyunsaturated margarine (e.g Flora) and also cholesterol lowering | bread_baguette_spread_polymarg_chol_med | 10 | bread_baguette_spread_polymarg_chol_med | 12.5 | 25% | 125% of the weight on a bread roll |
| Fat that is spread (thick amount) onto baguette: ticked polyunsaturated margarine (e.g Flora) and also cholesterol lowering | bread_baguette_spread_polymarg_chol_thick | 12 | bread_baguette_spread_polymarg_chol_thick | 15 | 25% | 125% of the weight on a bread roll |
| Fat that is spread (thin amount) onto baguette: ticked polyunsaturated margarine (e.g Flora) and also cholesterol lowering | bread_baguette_spread_polymarg_chol_thin | 7 | bread_baguette_spread_polymarg_chol_thin | 8.75 | 25% | 125% of the weight on a bread roll |
| Fat that is spread (medium amount) onto baguette: ticked polyunsaturated margarine (e.g Flora) but not specified amount of fat | bread_baguette_spread_polymarg_dunno_med | 10 | bread_baguette_spread_polymarg_dunno_med | 12.5 | 25% | 125% of the weight on a bread roll; food composition as low fat and reduced fat |
| Fat that is spread (thick amount) onto baguette: ticked polyunsaturated margarine (e.g Flora) but not specified amount of fat | bread_baguette_spread_polymarg_dunno_thick | 12 | bread_baguette_spread_polymarg_dunno_thick | 15 | 25% | 125% of the weight on a bread roll; food composition as low fat and reduced fat |
| Fat that is spread (thin amount) onto baguette: ticked polyunsaturated margarine (e.g Flora) but not specified amount of fat | bread_baguette_spread_polymarg_dunno_thin | 7 | bread_baguette_spread_polymarg_dunno_thin | 8.75 | 25% | 125% of the weight on a bread roll; food composition as low fat and reduced fat |
| Fat that is spread (medium amount) onto baguette: ticked polyunsaturated margarine (e.g Flora), normal amount of % fat | bread_baguette_spread_polymarg_fat_med | 10 | bread_baguette_spread_polymarg_fat_med | 12.5 | 25% | 125% of the weight on a bread roll; normal fat is taken as reduced fat (up to 62% fat) |
| Fat that is spread (thick amount) onto baguette: ticked polyunsaturated margarine (e.g Flora), normal amount of % fat | bread_baguette_spread_polymarg_fat_thick | 12 | bread_baguette_spread_polymarg_fat_thick | 15 | 25% | 125% of the weight on a bread roll; normal fat is taken as reduced fat (up to 62% fat) |
| Fat that is spread (thin amount) onto baguette: ticked polyunsaturated margarine (e.g Flora), normal amount of % fat | bread_baguette_spread_polymarg_fat_thin | 7 | bread_baguette_spread_polymarg_fat_thin | 8.75 | 25% | 125% of the weight on a bread roll; normal fat is taken as reduced fat (up to 62% fat) |
| Fat that is spread (medium amount) onto baguette: ticked polyunsaturated margarine (e.g Flora), low fat | bread_baguette_spread_polymarg_lowfat_med | 10 | bread_baguette_spread_polymarg_lowfat_med | 12.5 | 25% | 125% of the weight on a bread roll |
| Fat that is spread (thick amount) onto baguette: ticked polyunsaturated margarine (e.g Flora), low fat | bread_baguette_spread_polymarg_lowfat_thick | 12 | bread_baguette_spread_polymarg_lowfat_thick | 15 | 25% | 125% of the weight on a bread roll |
| Fat that is spread (thin amount) onto baguette: ticked polyunsaturated margarine (e.g Flora), low fat | bread_baguette_spread_polymarg_lowfat_thin | 7 | bread_baguette_spread_polymarg_lowfat_thin | 8.75 | 25% | 125% of the weight on a bread roll |
| Fat that is spread (medium amount) onto baguette: ticked polyunsaturated margarine (e.g Flora), very low fat | bread_baguette_spread_polymarg_vlowfat_med | 10 | bread_baguette_spread_polymarg_vlowfat_med | 12.5 | 25% | 125% of the weight on a bread roll; food composition as low fat spread |
| Fat that is spread (thick amount) onto baguette: ticked polyunsaturated margarine (e.g Flora), very low fat | bread_baguette_spread_polymarg_vlowfat_thick | 12 | bread_baguette_spread_polymarg_vlowfat_thick | 15 | 25% | 125% of the weight on a bread roll; food composition as low fat spread |
| Fat that is spread (thin amount) onto baguette: ticked polyunsaturated margarine (e.g Flora), very low fat | bread_baguette_spread_polymarg_vlowfat_thin | 7 | bread_baguette_spread_polymarg_vlowfat_thin | 8.75 | 25% | 125% of the weight on a bread roll; food composition as low fat spread |
| Fat that is spread (medium amount) onto baguette: ticked soya/vegan/dairy free margarine e.g Pure, and also cholesterol lowering | bread_baguette_spread_soya_chol_med | 10 | bread_baguette_spread_soya_chol_med | 12.5 | 25% | 125% of the weight on a bread roll |

| | McCance and Widdowson's | | Nutrient databank + other changes | | | |
|---|---|---|---|---|---|---|
| Variable description | Food item | Portion size | Food item | Portion size | Portion diff% | Description of differences with previous version |
| Fat that is spread (thick amount) onto baguette: ticked soya/vegan/dairy free margarine e.g Pure, and also cholesterol lowering | bread_baguette_spread_soya_chol_thick | 12 | bread_baguette_spread_soya_chol_thick | 15 | 25% | 125% of the weight on a bread roll |
| Fat that is spread (thin amount) onto baguette: ticked soya/vegan/dairy free margarine e.g Pure, and also cholesterol lowering | bread_baguette_spread_soya_chol_thin | 7 | bread_baguette_spread_soya_chol_thin | 8.75 | 25% | 125% of the weight on a bread roll |
| Fat that is spread (medium amount) onto baguette: ticked soya/vegan/dairy free margarine e.g Pure, and not specified amount of fat | bread_baguette_spread_soya_dunno_med | 10 | bread_baguette_spread_soya_dunno_med | 12.5 | 25% | 125% of the weight on a bread roll; food composition as low fat and reduced fat |
| Fat that is spread (thick amount) onto baguette: ticked soya/vegan/dairy free margarine e.g Pure, and not specified amount of fat | bread_baguette_spread_soya_dunno_thick | 12 | bread_baguette_spread_soya_dunno_thick | 15 | 25% | 125% of the weight on a bread roll; food composition as low fat and reduced fat |
| Fat that is spread (thin amount) onto baguette: ticked soya/vegan/dairy free margarine e.g Pure, and not specified amount of fat | bread_baguette_spread_soya_dunno_thin | 7 | bread_baguette_spread_soya_dunno_thin | 8.75 | 25% | 125% of the weight on a bread roll; food composition as low fat and reduced fat |
| Fat that is spread (medium amount) onto baguette: ticked soya/vegan/dairy free margarine (e.g Pure), normal amount % fat | bread_baguette_spread_soya_fat_med | 10 | bread_baguette_spread_soya_fat_med | 12.5 | 25% | 125% of the weight on a bread roll; normal fat is taken as reduced fat (up to 62% fat) |
| Fat that is spread (thick amount) onto baguette: ticked soya/vegan/dairy free margarine (e.g Pure), normal amount % fat | bread_baguette_spread_soya_fat_thick | 12 | bread_baguette_spread_soya_fat_thick | 15 | 25% | 125% of the weight on a bread roll; normal fat is taken as reduced fat (up to 62% fat) |
| Fat that is spread (thin amount) onto baguette: ticked soya/vegan/dairy free margarine (e.g Pure), normal amount % fat | bread_baguette_spread_soya_fat_thin | 7 | bread_baguette_spread_soya_fat_thin | 8.75 | 25% | 125% of the weight on a bread roll; normal fat is taken as reduced fat (up to 62% fat) |
| Fat that is spread (medium amount) onto baguette: ticked soya/vegan/dairy free margarine (e.g Pure), low fat | bread_baguette_spread_soya_lowfat_med | 10 | bread_baguette_spread_soya_lowfat_med | 12.5 | 25% | 125% of the weight on a bread roll |
| Fat that is spread (thick amount) onto baguette: ticked soya/vegan/dairy free margarine (e.g Pure), low fat | bread_baguette_spread_soya_lowfat_thick | 12 | bread_baguette_spread_soya_lowfat_thick | 15 | 25% | 125% of the weight on a bread roll |
| Fat that is spread (thin amount) onto baguette: ticked soya/vegan/dairy free margarine (e.g Pure), low fat | bread_baguette_spread_soya_lowfat_thin | 7 | bread_baguette_spread_soya_lowfat_thin | 8.75 | 25% | 125% of the weight on a bread roll |
| Fat that is spread (medium amount) onto baguette: ticked soya/vegan/dairy free margarine (e.g Pure), very low fat | bread_baguette_spread_soya_vlowfat_med | 10 | bread_baguette_spread_soya_vlowfat_med | 12.5 | 25% | 125% of the weight on a bread roll; food composition as low fat spread |
| Fat that is spread (thick amount) onto baguette: ticked soya/vegan/dairy free margarine (e.g Pure), very low fat | bread_baguette_spread_soya_vlowfat_thick | 12 | bread_baguette_spread_soya_vlowfat_thick | 15 | 25% | 125% of the weight on a bread roll; food composition as low fat spread |
| Fat that is spread (thin amount) onto baguette: ticked soya/vegan/dairy free margarine (e.g Pure), very low fat | bread_baguette_spread_soya_vlowfat_thin | 7 | bread_baguette_spread_soya_vlowfat_thin | 8.75 | 25% | 125% of the weight on a bread roll; food composition as low fat spread |
| Baguette, ciabatta, panini, sub: not specified type so includes wholemeal, brown or white sliced bread | bread_baguette_unanswered | 95 | bread_baguette_unanswered | 95 | 0% | |
| Baguette, ciabatta, panini, sub: french stick, plain ciabatta | bread_baguette_white | 95 | bread_baguette_white | 95 | 0% | |
| Baguette, ciabatta, panini, sub: wholemeal | bread_baguette_wholemeal | 95 | bread_baguette_wholemeal | 95 | 0% | |
| Crisp bread e.g ryvita, crackers, wholemeal crackers, rice cakes | bread_crisp | 10 | bread_crisp | 10 | 0% | |
| Crisp bread e.g ryvita, crackers, wholemeal crackers, rice cakes | | | bread_crisp_gf | 10 | - | Gluten free version added to the updated version (however no gluten free code available at the time so the non-gluten free code was used) |
| Fat that is spread (medium amount) onto crisp bread: ticked butter but not specified amount of % fat | bread_crisp_spread_butter_dunno_med | 7 | bread_crisp_spread_butter_dunno_med | 3.9 | -44% | 40% crispbread+rice cakes (@60% of slice) + 60% cream crackers (@25% of slice); food composition as butter and spreadable butter |
| Fat that is spread (thick amount) onto crisp bread: ticked butter but not specified amount of % fat | bread_crisp_spread_butter_dunno_thick | 10 | bread_crisp_spread_butter_dunno_thick | 4.7 | -53% | 40% crispbread+rice cakes (@60% of slice) + 60% cream crackers (@25% of slice); food composition as butter and spreadable butter |
| Fat that is spread (thin amount) onto crisp bread: ticked butter but not specified amount of % fat | bread_crisp_spread_butter_dunno_thin | 5 | bread_crisp_spread_butter_dunno_thin | 2.7 | -46% | 40% crispbread+rice cakes (@60% of slice) + 60% cream crackers (@25% of slice); food composition as butter and spreadable butter |
| Fat that is spread (medium amount) onto crisp bread: ticked butter, normal amount of % fat | bread_crisp_spread_butter_fat_med | 7 | bread_crisp_spread_butter_fat_med | 3.9 | -44% | 40% crispbread+rice cakes (@62% of slice) + 62% cream crackers (@25% of slice) |
| Fat that is spread (thick amount) onto crisp bread: ticked butter, normal amount of % fat | bread_crisp_spread_butter_fat_thick | 10 | bread_crisp_spread_butter_fat_thick | 4.7 | -53% | 40% crispbread+rice cakes (@62% of slice) + 62% cream crackers (@25% of slice) |
| Fat that is spread (thin amount) onto crisp bread: ticked butter, normal amount of % fat | bread_crisp_spread_butter_fat_thin | 5 | bread_crisp_spread_butter_fat_thin | 2.7 | -46% | 40% crispbread+rice cakes (@62% of slice) + 62% cream crackers (@25% of slice) |
| Fat that is spread (medium amount) onto crisp bread: ticked butter, low fat | bread_crisp_spread_butter_lowfat_med | 7 | bread_crisp_spread_butter_lowfat_med | 3.9 | -44% | 40% crispbread+rice cakes (@60% of slice) + 60% cream crackers (@25% of slice) |
| Fat that is spread (thick amount) onto crisp bread: ticked butter, low fat | bread_crisp_spread_butter_lowfat_thick | 10 | bread_crisp_spread_butter_lowfat_thick | 4.7 | -53% | 40% crispbread+rice cakes (@60% of slice) + 60% cream crackers (@25% of slice) |
| Fat that is spread (thin amount) onto crisp bread: ticked butter, low fat | bread_crisp_spread_butter_lowfat_thin | 5 | bread_crisp_spread_butter_lowfat_thin | 2.7 | -46% | 40% crispbread+rice cakes (@60% of slice) + 60% cream crackers (@25% of slice) |
| Fat that is spread (medium amount) onto crisp bread: ticked spreadable butter with normal amount of % fat | bread_crisp_spread_butter_spread_fat_med | 7 | bread_crisp_spread_butter_spread_fat_med | 3.9 | -44% | 40% crispbread+rice cakes (@62% of slice) + 62% cream crackers (@25% of slice) |
| Fat that is spread (thick amount) onto crisp bread: ticked spreadable butter with normal amount of % fat | bread_crisp_spread_butter_spread_fat_thick | 10 | bread_crisp_spread_butter_spread_fat_thick | 4.7 | -53% | 40% crispbread+rice cakes (@62% of slice) + 62% cream crackers (@25% of slice) |
| Fat that is spread (thin amount) onto crisp bread: ticked spreadable butter with normal amount of % fat | bread_crisp_spread_butter_spread_fat_thin | 5 | bread_crisp_spread_butter_spread_fat_thin | 2.7 | -46% | 40% crispbread+rice cakes (@62% of slice) + 62% cream crackers (@25% of slice) |
| Fat that is spread (medium amount) onto crisp bread: ticked spreadable butter, low fat | bread_crisp_spread_butter_spread_lowfat_med | 7 | bread_crisp_spread_butter_spread_lowfat_med | 3.9 | -44% | 40% crispbread+rice cakes (@60% of slice) + 60% cream crackers (@25% of slice) |
| Fat that is spread (thick amount) onto crisp bread: ticked spreadable butter, low fat | bread_crisp_spread_butter_spread_lowfat_thick | 10 | bread_crisp_spread_butter_spread_lowfat_thick | 4.7 | -53% | 40% crispbread+rice cakes (@60% of slice) + 60% cream crackers (@25% of slice) |
| Fat that is spread (thin amount) onto crisp bread: ticked spreadable butter, low fat | bread_crisp_spread_butter_spread_lowfat_thin | 5 | bread_crisp_spread_butter_spread_lowfat_thin | 2.7 | -46% | 40% crispbread+rice cakes (@60% of slice) + 60% cream crackers (@25% of slice) |
| Fat that is spread (medium amount) onto crisp bread: ticked dairy spread which is also cholesterol lowering e.g Benecol Buttery | bread_crisp_spread_dairy_chol_med | 5 | bread_crisp_spread_dairy_chol_med | 2.7 | -46% | 40% crispbread+rice cakes (@60% of slice) + 60% cream crackers (@25% of slice) |
| Fat that is spread (thick amount) onto crisp bread: ticked dairy spread which is also cholesterol lowering e.g Benecol Buttery | bread_crisp_spread_dairy_chol_thick | 7 | bread_crisp_spread_dairy_chol_thick | 3.9 | -44% | 40% crispbread+rice cakes (@60% of slice) + 60% cream crackers (@25% of slice) |
| Fat that is spread (thin amount) onto crisp bread: ticked dairy spread which is also cholesterol lowering e.g Benecol Buttery | bread_crisp_spread_dairy_chol_thin | 3 | bread_crisp_spread_dairy_chol_thin | 2 | -33% | 40% crispbread+rice cakes (@60% of slice) + 60% cream crackers (@25% of slice) |
| Fat that is spread (medium amount) onto crisp bread: ticked dairy spread but not specified amount of % fat | bread_crisp_spread_dairy_dunno_med | 5 | bread_crisp_spread_dairy_dunno_med | 2.7 | -46% | 40% crispbread+rice cakes (@60% of slice) + 60% cream crackers (@25% of slice); food composition as low fat and reduced fat |
| Fat that is spread (thick amount) onto crisp bread: ticked dairy spread but not specified amount of % fat | bread_crisp_spread_dairy_dunno_thick | 7 | bread_crisp_spread_dairy_dunno_thick | 3.9 | -44% | 40% crispbread+rice cakes (@60% of slice) + 60% cream crackers (@25% of slice); food composition as low fat and reduced fat |
| Fat that is spread (thin amount) onto crisp bread: ticked dairy spread but not specified amount of % fat | bread_crisp_spread_dairy_dunno_thin | 3 | bread_crisp_spread_dairy_dunno_thin | 2 | -33% | 40% crispbread+rice cakes (@60% of slice) + 60% cream crackers (@25% of slice); food composition as low fat and reduced fat |
| Fat that is spread (medium amount) onto crisp bread: ticked dairy spread with normal amount of % fat | bread_crisp_spread_dairy_fat_med | 5 | bread_crisp_spread_dairy_fat_med | 2.7 | -46% | 40% crispbread+rice cakes (@62% of slice) + 62% cream crackers (@25% of slice); normal fat is taken as reduced fat (up to 62% fat) |
| Fat that is spread (thick amount) onto crisp bread: ticked dairy spread with normal amount of % fat | bread_crisp_spread_dairy_fat_thick | 7 | bread_crisp_spread_dairy_fat_thick | 3.9 | -44% | 40% crispbread+rice cakes (@62% of slice) + 62% cream crackers (@25% of slice); normal fat is taken as reduced fat (up to 62% fat) |
| Fat that is spread (thin amount) onto crisp bread: ticked dairy spread with normal amount of % fat | bread_crisp_spread_dairy_fat_thin | 3 | bread_crisp_spread_dairy_fat_thin | 2 | -33% | 40% crispbread+rice cakes (@62% of slice) + 62% cream crackers (@25% of slice); normal fat is taken as reduced fat (up to 62% fat) |
| Fat that is spread (medium amount) onto crisp bread: ticked dairy spread, low fat | bread_crisp_spread_dairy_lowfat_med | 5 | bread_crisp_spread_dairy_lowfat_med | 2.7 | -46% | 40% crispbread+rice cakes (@60% of slice) + 60% cream crackers (@25% of slice) |
| Fat that is spread (thick amount) onto crisp bread: ticked dairy spread, low fat | bread_crisp_spread_dairy_lowfat_thick | 7 | bread_crisp_spread_dairy_lowfat_thick | 3.9 | -44% | 40% crispbread+rice cakes (@60% of slice) + 60% cream crackers (@25% of slice) |
| Fat that is spread (thin amount) onto crisp bread: ticked dairy spread, low fat | bread_crisp_spread_dairy_lowfat_thin | 3 | bread_crisp_spread_dairy_lowfat_thin | 2 | -33% | 40% crispbread+rice cakes (@60% of slice) + 60% cream crackers (@25% of slice) |
| Fat that is spread (medium amount) onto crisp bread: ticked dairy spread, very low fat | bread_crisp_spread_dairy_vlowfat_med | 5 | bread_crisp_spread_dairy_vlowfat_med | 2.7 | -46% | 40% crispbread+rice cakes (@60% of slice) + 60% cream crackers (@25% of slice); food composition as low fat spread |
| Fat that is spread (thick amount) onto crisp bread: ticked dairy spread, very low fat | bread_crisp_spread_dairy_vlowfat_thick | 7 | bread_crisp_spread_dairy_vlowfat_thick | 3.9 | -44% | 40% crispbread+rice cakes (@60% of slice) + 60% cream crackers (@25% of slice); food composition as low fat spread |
| Fat that is spread (thin amount) onto crisp bread: ticked dairy spread, very low fat | bread_crisp_spread_dairy_vlowfat_thin | 3 | bread_crisp_spread_dairy_vlowfat_thin | 2 | -33% | 40% crispbread+rice cakes (@60% of slice) + 60% cream crackers (@25% of slice); food composition as low fat spread |
| Fat that is spread (medium amount) onto crisp bread: not specified type of spread but ticked cholesterol lowering e.g Benecol, Flora pro active | bread_crisp_spread_dunno_chol_med | 5 | bread_crisp_spread_dunno_chol_med | 2.7 | -46% | 40% crispbread+rice cakes (@60% of slice) + 60% cream crackers (@25% of slice) |
| Fat that is spread (thick amount) onto crisp bread: not specified type of spread but ticked cholesterol lowering e.g Benecol, Flora pro active | bread_crisp_spread_dunno_chol_thick | 7 | bread_crisp_spread_dunno_chol_thick | 3.9 | -44% | 40% crispbread+rice cakes (@60% of slice) + 60% cream crackers (@25% of slice) |
| Fat that is spread (thin amount) onto crisp bread: not specified type of spread but ticked cholesterol lowering e.g Benecol, Flora pro active | bread_crisp_spread_dunno_chol_thin | 3 | bread_crisp_spread_dunno_chol_thin | 2 | -33% | 40% crispbread+rice cakes (@60% of slice) + 60% cream crackers (@25% of slice) |
| Fat that is spread (medium amount) onto crisp bread: not specified type of spread or amount of % fat | bread_crisp_spread_dunno_dunno_med | 5 | bread_crisp_spread_dunno_dunno_med | 2.7 | -46% | 40% crispbread+rice cakes (@60% of slice) + 60% cream crackers (@25% of slice) |
| Fat that is spread (thick amount) onto crisp bread: not specified type of spread or amount of % fat | bread_crisp_spread_dunno_dunno_thick | 7 | bread_crisp_spread_dunno_dunno_thick | 3.9 | -44% | 40% crispbread+rice cakes (@60% of slice) + 60% cream crackers (@25% of slice) |
| Fat that is spread (thin amount) onto crisp bread: not specified type of spread or amount of % fat | bread_crisp_spread_dunno_dunno_thin | 3 | bread_crisp_spread_dunno_dunno_thin | 2 | -33% | 40% crispbread+rice cakes (@60% of slice) + 60% cream crackers (@25% of slice) |
| Fat that is spread (medium amount) onto crisp bread: not specified type of spread but ticked normal amount % fat | bread_crisp_spread_dunno_fat_med | 5 | bread_crisp_spread_dunno_fat_med | 2.7 | -46% | 40% crispbread+rice cakes (@62% of slice) + 62% cream crackers (@25% of slice); normal fat is taken as reduced fat (up to 62% fat) |
| Fat that is spread (thick amount) onto crisp bread: not specified type of spread but ticked normal amount % fat | bread_crisp_spread_dunno_fat_thick | 7 | bread_crisp_spread_dunno_fat_thick | 3.9 | -44% | 40% crispbread+rice cakes (@62% of slice) + 62% cream crackers (@25% of slice); normal fat is taken as reduced fat (up to 62% fat) |
| Fat that is spread (thin amount) onto crisp bread: not specified type of spread but ticked normal amount % fat | bread_crisp_spread_dunno_fat_thin | 3 | bread_crisp_spread_dunno_fat_thin | 2 | -33% | 40% crispbread+rice cakes (@62% of slice) + 62% cream crackers (@25% of slice); normal fat is taken as reduced fat (up to 62% fat) |
| Fat that is spread (medium amount) onto crisp bread: not specified type of spread but ticked low fat | bread_crisp_spread_dunno_lowfat_med | 5 | bread_crisp_spread_dunno_lowfat_med | 2.7 | -46% | 40% crispbread+rice cakes (@60% of slice) + 60% cream crackers (@25% of slice) |
| Fat that is spread (thick amount) onto crisp bread: not specified type of spread but ticked low fat | bread_crisp_spread_dunno_lowfat_thick | 7 | bread_crisp_spread_dunno_lowfat_thick | 3.9 | -44% | 40% crispbread+rice cakes (@60% of slice) + 60% cream crackers (@25% of slice) |
| Fat that is spread (thin amount) onto crisp bread: not specified type of spread but ticked low fat | bread_crisp_spread_dunno_lowfat_thin | 3 | bread_crisp_spread_dunno_lowfat_thin | 2 | -33% | 40% crispbread+rice cakes (@60% of slice) + 60% cream crackers (@25% of slice) |
| Fat that is spread (medium amount) onto crisp bread: not specified type of spread but ticked very low fat | bread_crisp_spread_dunno_vlowfat_med | 5 | bread_crisp_spread_dunno_vlowfat_med | 2.7 | -46% | 40% crispbread+rice cakes (@60% of slice) + 60% cream crackers (@25% of slice); food composition as low fat spread |
| Fat that is spread (thick amount) onto crisp bread: not specified type of spread but ticked very low fat | bread_crisp_spread_dunno_vlowfat_thick | 7 | bread_crisp_spread_dunno_vlowfat_thick | 3.9 | -44% | 40% crispbread+rice cakes (@60% of slice) + 60% cream crackers (@25% of slice); food composition as low fat spread |
| Fat that is spread (thin amount) onto crisp bread: not specified type of spread but ticked very low fat | bread_crisp_spread_dunno_vlowfat_thin | 3 | bread_crisp_spread_dunno_vlowfat_thin | 2 | -33% | 40% crispbread+rice cakes (@60% of slice) + 60% cream crackers (@25% of slice); food composition as low fat spread |
| Fat that is spread (medium amount) onto crisp bread: ticked hard margarine (hard block margarine in wrapper) | bread_crisp_spread_hardmarg_med | 7 | bread_crisp_spread_hardmarg_med | 3.9 | -44% | 40% crispbread+rice cakes (@60% of slice) + 60% cream crackers (@25% of slice); spread weight as butter |
| Fat that is spread (thick amount) onto crisp bread: ticked hard margarine (hard block margarine in wrapper) | bread_crisp_spread_hardmarg_thick | 10 | bread_crisp_spread_hardmarg_thick | 4.7 | -53% | 40% crispbread+rice cakes (@60% of slice) + 60% cream crackers (@25% of slice); spread weight as butter |
| Fat that is spread (thin amount) onto crisp bread: ticked hard margarine (hard block margarine in wrapper) | bread_crisp_spread_hardmarg_thin | 5 | bread_crisp_spread_hardmarg_thin | 2.7 | -46% | 40% crispbread+rice cakes (@60% of slice) + 60% cream crackers (@25% of slice); spread weight as butter |
| Fat that is spread (medium amount) onto crisp bread: ticked cholesterol lowering olive spread e.g Benecol/Flora pro active olive spread | bread_crisp_spread_olive_chol_med | 5 | bread_crisp_spread_olive_chol_med | 2.7 | -46% | 40% crispbread+rice cakes (@60% of slice) + 60% cream crackers (@25% of slice) |
| Fat that is spread (thick amount) onto crisp bread: ticked cholesterol lowering olive spread e.g Benecol/Flora pro active olive spread | bread_crisp_spread_olive_chol_thick | 7 | bread_crisp_spread_olive_chol_thick | 3.9 | -44% | 40% crispbread+rice cakes (@60% of slice) + 60% cream crackers (@25% of slice) |
| Fat that is spread (thin amount) onto crisp bread: ticked cholesterol lowering olive spread e.g Benecol/Flora pro active olive spread | bread_crisp_spread_olive_chol_thin | 3 | bread_crisp_spread_olive_chol_thin | 2 | -33% | 40% crispbread+rice cakes (@60% of slice) + 60% cream crackers (@25% of slice) |
| Fat that is spread (medium amount) onto crisp bread: ticked olive spread but not specified amount of fat | bread_crisp_spread_olive_dunno_med | 5 | bread_crisp_spread_olive_dunno_med | 2.7 | -46% | 40% crispbread+rice cakes (@60% of slice) + 60% cream crackers (@25% of slice) |
| Fat that is spread (thick amount) onto crisp bread: ticked olive spread but not specified amount of fat | bread_crisp_spread_olive_dunno_thick | 7 | bread_crisp_spread_olive_dunno_thick | 3.9 | -44% | 40% crispbread+rice cakes (@60% of slice) + 60% cream crackers (@25% of slice) |
| Fat that is spread (thin amount) onto crisp bread: ticked olive spread but not specified amount of fat | bread_crisp_spread_olive_dunno_thin | 3 | bread_crisp_spread_olive_dunno_thin | 2 | -33% | 40% crispbread+rice cakes (@60% of slice) + 60% cream crackers (@25% of slice) |
| Fat that is spread (medium amount) onto crisp bread: ticked olive spread with normal amount of % fat | bread_crisp_spread_olive_fat_med | 5 | bread_crisp_spread_olive_fat_med | 2.7 | -46% | 40% crispbread+rice cakes (@62% of slice) + 62% cream crackers (@25% of slice); normal fat is taken as reduced fat (up to 62% fat) |
| Fat that is spread (thick amount) onto crisp bread: ticked olive spread with normal amount of % fat | bread_crisp_spread_olive_fat_thick | 7 | bread_crisp_spread_olive_fat_thick | 3.9 | -44% | 40% crispbread+rice cakes (@62% of slice) + 62% cream crackers (@25% of slice); normal fat is taken as reduced fat (up to 62% fat) |
| Fat that is spread (thin amount) onto crisp bread: ticked olive spread with normal amount of % fat | bread_crisp_spread_olive_fat_thin | 3 | bread_crisp_spread_olive_fat_thin | 2 | -33% | 40% crispbread+rice cakes (@62% of slice) + 62% cream crackers (@25% of slice); normal fat is taken as reduced fat (up to 62% fat) |
| Fat that is spread (medium amount) onto crisp bread: ticked olive spread, low fat | bread_crisp_spread_olive_lowfat_med | 5 | bread_crisp_spread_olive_lowfat_med | 2.7 | -46% | 40% crispbread+rice cakes (@60% of slice) + 60% cream crackers (@25% of slice) |
| Fat that is spread (thick amount) onto crisp bread: ticked olive spread, low fat | bread_crisp_spread_olive_lowfat_thick | 7 | bread_crisp_spread_olive_lowfat_thick | 3.9 | -44% | 40% crispbread+rice cakes (@60% of slice) + 60% cream crackers (@25% of slice) |
| Fat that is spread (thin amount) onto crisp bread: ticked olive spread, low fat | bread_crisp_spread_olive_lowfat_thin | 3 | bread_crisp_spread_olive_lowfat_thin | 2 | -33% | 40% crispbread+rice cakes (@60% of slice) + 60% cream crackers (@25% of slice) |
| Fat that is spread (medium amount) onto crisp bread: ticked olive spread, very low fat | bread_crisp_spread_olive_vlowfat_med | 5 | bread_crisp_spread_olive_vlowfat_med | 2.7 | -46% | 40% crispbread+rice cakes (@60% of slice) + 60% cream crackers (@25% of slice); food composition as low fat spread |
| Fat that is spread (thick amount) onto crisp bread: ticked olive spread, very low fat | bread_crisp_spread_olive_vlowfat_thick | 7 | bread_crisp_spread_olive_vlowfat_thick | 3.9 | -44% | 40% crispbread+rice cakes (@60% of slice) + 60% cream crackers (@25% of slice); food composition as low fat spread |
| Fat that is spread (thin amount) onto crisp bread: ticked olive spread, very low fat | bread_crisp_spread_olive_vlowfat_thin | 3 | bread_crisp_spread_olive_vlowfat_thin | 2 | -33% | 40% crispbread+rice cakes (@60% of slice) + 60% cream crackers (@25% of slice); food composition as low fat spread |
| Fat that is spread (medium amount) onto crisp bread: ticked other spread e.g ghee | bread_crisp_spread_other_med | 5 | bread_crisp_spread_other_med | 2.7 | -46% | 40% crispbread+rice cakes (@60% of slice) + 60% cream crackers (@25% of slice) |
| Fat that is spread (thick amount) onto crisp bread: ticked other spread e.g ghee | bread_crisp_spread_other_thick | 7 | bread_crisp_spread_other_thick | 3.9 | -44% | 40% crispbread+rice cakes (@60% of slice) + 60% cream crackers (@25% of slice) |
| Fat that is spread (thin amount) onto crisp bread: ticked other spread e.g ghee | bread_crisp_spread_other_thin | 3 | bread_crisp_spread_other_thin | 2 | -33% | 40% crispbread+rice cakes (@60% of slice) + 60% cream crackers (@25% of slice) |
| Fat that is spread (medium amount) onto crisp bread: ticked polyunsaturated margarine (e.g Flora) and also cholesterol lowering | bread_crisp_spread_polymarg_chol_med | 5 | bread_crisp_spread_polymarg_chol_med | 2.7 | -46% | 40% crispbread+rice cakes (@60% of slice) + 60% cream crackers (@25% of slice) |
| Fat that is spread (thick amount) onto crisp bread: ticked polyunsaturated margarine (e.g Flora) and also cholesterol lowering | bread_crisp_spread_polymarg_chol_thick | 7 | bread_crisp_spread_polymarg_chol_thick | 3.9 | -44% | 40% crispbread+rice cakes (@60% of slice) + 60% cream crackers (@25% of slice) |
| Fat that is spread (thin amount) onto crisp bread: ticked polyunsaturated margarine (e.g Flora) and also cholesterol lowering | bread_crisp_spread_polymarg_chol_thin | 3 | bread_crisp_spread_polymarg_chol_thin | 2 | -33% | 40% crispbread+rice cakes (@60% of slice) + 60% cream crackers (@25% of slice) |
| Fat that is spread (medium amount) onto crisp bread: ticked polyunsaturated margarine (e.g Flora) but not specified amount of fat | bread_crisp_spread_polymarg_dunno_med | 5 | bread_crisp_spread_polymarg_dunno_med | 2.7 | -46% | 40% crispbread+rice cakes (@60% of slice) + 60% cream crackers (@25% of slice) |
| Fat that is spread (thick amount) onto crisp bread: ticked polyunsaturated margarine (e.g Flora) but not specified amount of fat | bread_crisp_spread_polymarg_dunno_thick | 7 | bread_crisp_spread_polymarg_dunno_thick | 3.9 | -44% | 40% crispbread+rice cakes (@60% of slice) + 60% cream crackers (@25% of slice) |
| Fat that is spread (thin amount) onto crisp bread: ticked polyunsaturated margarine (e.g Flora) but not specified amount of fat | bread_crisp_spread_polymarg_dunno_thin | 3 | bread_crisp_spread_polymarg_dunno_thin | 2 | -33% | 40% crispbread+rice cakes (@60% of slice) + 60% cream crackers (@25% of slice) |
| Fat that is spread (medium amount) onto crisp bread: ticked polyunsaturated margarine (e.g Flora), normal amount of % fat | bread_crisp_spread_polymarg_fat_med | 5 | bread_crisp_spread_polymarg_fat_med | 2.7 | -46% | 40% crispbread+rice cakes (@62% of slice) + 62% cream crackers (@25% of slice); normal fat is taken as reduced fat (up to 62% fat) |
| Fat that is spread (thick amount) onto crisp bread: ticked polyunsaturated margarine (e.g Flora), normal amount of % fat | bread_crisp_spread_polymarg_fat_thick | 7 | bread_crisp_spread_polymarg_fat_thick | 3.9 | -44% | 40% crispbread+rice cakes (@62% of slice) + 62% cream crackers (@25% of slice); normal fat is taken as reduced fat (up to 62% fat) |
| Fat that is spread (thin amount) onto crisp bread: ticked polyunsaturated margarine (e.g Flora), normal amount of % fat | bread_crisp_spread_polymarg_fat_thin | 3 | bread_crisp_spread_polymarg_fat_thin | 2 | -33% | 40% crispbread+rice cakes (@62% of slice) + 62% cream crackers (@25% of slice); normal fat is taken as reduced fat (up to 62% fat) |
| Fat that is spread (medium amount) onto crisp bread: ticked polyunsaturated margarine (e.g Flora), low fat | bread_crisp_spread_polymarg_lowfat_med | 5 | bread_crisp_spread_polymarg_lowfat_med | 2.7 | -46% | 40% crispbread+rice cakes (@60% of slice) + 60% cream crackers (@25% of slice) |
| Fat that is spread (thick amount) onto crisp bread: ticked polyunsaturated margarine (e.g Flora), low fat | bread_crisp_spread_polymarg_lowfat_thick | 7 | bread_crisp_spread_polymarg_lowfat_thick | 3.9 | -44% | 40% crispbread+rice cakes (@60% of slice) + 60% cream crackers (@25% of slice) |
| Fat that is spread (thin amount) onto crisp bread: ticked polyunsaturated margarine (e.g Flora), low fat | bread_crisp_spread_polymarg_lowfat_thin | 3 | bread_crisp_spread_polymarg_lowfat_thin | 2 | -33% | 40% crispbread+rice cakes (@60% of slice) + 60% cream crackers (@25% of slice) |
| Fat that is spread (medium amount) onto crisp bread: ticked polyunsaturated margarine (e.g Flora), very low fat | bread_crisp_spread_polymarg_vlowfat_med | 5 | bread_crisp_spread_polymarg_vlowfat_med | 2.7 | -46% | 40% crispbread+rice cakes (@60% of slice) + 60% cream crackers (@25% of slice); food composition as low fat spread |
| Fat that is spread (thick amount) onto crisp bread: ticked polyunsaturated margarine (e.g Flora), very low fat | bread_crisp_spread_polymarg_vlowfat_thick | 7 | bread_crisp_spread_polymarg_vlowfat_thick | 3.9 | -44% | 40% crispbread+rice cakes (@60% of slice) + 60% cream crackers (@25% of slice); food composition as low fat spread |
| Fat that is spread (thin amount) onto crisp bread: ticked polyunsaturated margarine (e.g Flora), very low fat | bread_crisp_spread_polymarg_vlowfat_thin | 3 | bread_crisp_spread_polymarg_vlowfat_thin | 2 | -33% | 40% crispbread+rice cakes (@60% of slice) + 60% cream crackers (@25% of slice); food composition as low fat spread |
| Fat that is spread (medium amount) onto crisp bread: ticked soya/vegan/dairy free margarine e.g Pure, and also cholesterol lowering | bread_crisp_spread_soya_chol_med | 5 | bread_crisp_spread_soya_chol_med | 2.7 | -46% | 40% crispbread+rice cakes (@60% of slice) + 60% cream crackers (@25% of slice) |
| Fat that is spread (thick amount) onto crisp bread: ticked soya/vegan/dairy free margarine e.g Pure, and also cholesterol lowering | bread_crisp_spread_soya_chol_thick | 7 | bread_crisp_spread_soya_chol_thick | 3.9 | -44% | 40% crispbread+rice cakes (@60% of slice) + 60% cream crackers (@25% of slice) |
| Fat that is spread (thin amount) onto crisp bread: ticked soya/vegan/dairy free margarine e.g Pure, and also cholesterol lowering | bread_crisp_spread_soya_chol_thin | 3 | bread_crisp_spread_soya_chol_thin | 2 | -33% | 40% crispbread+rice cakes (@60% of slice) + 60% cream crackers (@25% of slice) |
| Fat that is spread (medium amount) onto crisp bread: ticked soya/vegan/dairy free margarine e.g Pure, and not specified amount of fat | bread_crisp_spread_soya_dunno_med | 5 | bread_crisp_spread_soya_dunno_med | 2.7 | -46% | 40% crispbread+rice cakes (@60% of slice) + 60% cream crackers (@25% of slice) |
| Fat that is spread (thick amount) onto crisp bread: ticked soya/vegan/dairy free margarine e.g Pure, and not specified amount of fat | bread_crisp_spread_soya_dunno_thick | 7 | bread_crisp_spread_soya_dunno_thick | 3.9 | -44% | 40% crispbread+rice cakes (@60% of slice) + 60% cream crackers (@25% of slice) |
| Fat that is spread (thin amount) onto crisp bread: ticked soya/vegan/dairy free margarine e.g Pure, and not specified amount of fat | bread_crisp_spread_soya_dunno_thin | 3 | bread_crisp_spread_soya_dunno_thin | 2 | -33% | 40% crispbread+rice cakes (@60% of slice) + 60% cream crackers (@25% of slice) |
| Fat that is spread (medium amount) onto crisp bread: ticked soya/vegan/dairy free margarine (e.g Pure), normal amount % fat | bread_crisp_spread_soya_fat_med | 5 | bread_crisp_spread_soya_fat_med | 2.7 | -46% | 40% crispbread+rice cakes (@62% of slice) + 62% cream crackers (@25% of slice); normal fat is taken as reduced fat (up to 62% fat) |
| Fat that is spread (thick amount) onto crisp bread: ticked soya/vegan/dairy free margarine (e.g Pure), normal amount % fat | bread_crisp_spread_soya_fat_thick | 7 | bread_crisp_spread_soya_fat_thick | 3.9 | -44% | 40% crispbread+rice cakes (@62% of slice) + 62% cream crackers (@25% of slice); normal fat is taken as reduced fat (up to 62% fat) |
| Fat that is spread (thin amount) onto crisp bread: ticked soya/vegan/dairy free margarine (e.g Pure), normal amount % fat | bread_crisp_spread_soya_fat_thin | 3 | bread_crisp_spread_soya_fat_thin | 2 | -33% | 40% crispbread+rice cakes (@62% of slice) + 62% cream crackers (@25% of slice); normal fat is taken as reduced fat (up to 62% fat) |

| | McCance and Widdowson's | | Nutrient databank + other changes | | | |
|---|---|---|---|---|---|---|
| Variable description | Food item | Portion size | Food item | Portion size | Portion diff% | Description of differences with previous version |
| Fat that is spread (medium amount) onto crisp bread: ticked soya/vegan/dairy free margarine (e.g Pure), low fat | bread_crisp_spread_soya_lowfat_med | 5 | bread_crisp_spread_soya_lowfat_med | 2.7 | -46% | 40% crispbread+rice cakes (@60% of slice) + 60% cream crackers (@25% of slice) |
| Fat that is spread (thick amount) onto crisp bread: ticked soya/vegan/dairy free margarine (e.g Pure), low fat | bread_crisp_spread_soya_lowfat_thick | 7 | bread_crisp_spread_soya_lowfat_thick | 3.9 | -44% | 40% crispbread+rice cakes (@60% of slice) + 60% cream crackers (@25% of slice) |
| Fat that is spread (thin amount) onto crisp bread: ticked soya/vegan/dairy free margarine (e.g Pure), low fat | bread_crisp_spread_soya_lowfat_thin | 3 | bread_crisp_spread_soya_lowfat_thin | 2 | -33% | 40% crispbread+rice cakes (@60% of slice) + 60% cream crackers (@25% of slice) |
| Fat that is spread (medium amount) onto crisp bread: ticked soya/vegan/dairy free margarine (e.g Pure), very low fat | bread_crisp_spread_soya_vlowfat_med | 5 | bread_crisp_spread_soya_vlowfat_med | 2.7 | -46% | 40% crispbread+rice cakes (@60% of slice) + 60% cream crackers (@25% of slice); food composition as low fat spread |
| Fat that is spread (thick amount) onto crisp bread: ticked soya/vegan/dairy free margarine (e.g Pure), very low fat | bread_crisp_spread_soya_vlowfat_thick | 7 | bread_crisp_spread_soya_vlowfat_thick | 3.9 | -44% | 40% crispbread+rice cakes (@60% of slice) + 60% cream crackers (@25% of slice); food composition as low fat spread |
| Fat that is spread (thin amount) onto crisp bread: ticked soya/vegan/dairy free margarine (e.g Pure), very low fat | bread_crisp_spread_soya_vlowfat_thin | 3 | bread_crisp_spread_soya_vlowfat_thin | 2 | -33% | 40% crispbread+rice cakes (@60% of slice) + 60% cream crackers (@25% of slice); food composition as low fat spread |
| Garlic bread | bread_garlic | 20 | bread_garlic | 20 | 0% | |
| Gluten free large bap, stotty, pitta bread: brown or with added fibre | | | bread_large_bap_gf_nonwhite | 90 | - | Gluten free version added to the updated version |
| Gluten free large bap, stotty, pitta bread: not specified type so includes white, brown or with added fibre | | | bread_large_bap_gf_unanswered | 90 | - | Gluten free version added to the updated version |
| Gluten free large bap, stotty, pitta bread: white | | | bread_large_bap_gf_white | 90 | - | Gluten free version added to the updated version |
| Large bap, stotty, pitta bread: brown, granary, wheatgerm, 50/50, soft or crusty | bread_large_bap_mixed | 90 | bread_large_bap_mixed | 90 | 0% | |
| Large bap, stotty, pitta bread: wheatgerm, rye, white | bread_large_bap_other | 90 | bread_large_bap_other | 90 | 0% | |
| Seeds for seeded bread: e.g sesame, sunflower, poppy | bread_large_bap_seeded | 3 | bread_large_bap_seeded | 3 | 0% | |
| Fat that is spread (medium amount) onto large bap: ticked butter but not specified amount of % fat | bread_large_bap_spread_butter_dunno_med | 12 | bread_large_bap_spread_butter_dunno_med | 15 | 25% | 125% of the weight on a bread roll; food composition as butter and spreadable butter |
| Fat that is spread (thick amount) onto large bap: ticked butter but not specified amount of % fat | bread_large_bap_spread_butter_dunno_thick | 15 | bread_large_bap_spread_butter_dunno_thick | 18.75 | 25% | 125% of the weight on a bread roll; food composition as butter and spreadable butter |
| Fat that is spread (thin amount) onto large bap: ticked butter but not specified amount of % fat | bread_large_bap_spread_butter_dunno_thin | 10 | bread_large_bap_spread_butter_dunno_thin | 12.5 | 25% | 125% of the weight on a bread roll; food composition as butter and spreadable butter |
| Fat that is spread (medium amount) onto large bap: ticked butter, normal amount of % fat | bread_large_bap_spread_butter_fat_med | 12 | bread_large_bap_spread_butter_fat_med | 15 | 25% | 125% of the weight on a bread roll |
| Fat that is spread (thick amount) onto large bap: ticked butter, normal amount of % fat | bread_large_bap_spread_butter_fat_thick | 15 | bread_large_bap_spread_butter_fat_thick | 18.75 | 25% | 125% of the weight on a bread roll |
| Fat that is spread (thin amount) onto large bap: ticked butter, normal amount of % fat | bread_large_bap_spread_butter_fat_thin | 10 | bread_large_bap_spread_butter_fat_thin | 12.5 | 25% | 125% of the weight on a bread roll |
| Fat that is spread (medium amount) onto large bap: ticked butter, low fat | bread_large_bap_spread_butter_lowfat_med | 12 | bread_large_bap_spread_butter_lowfat_med | 15 | 25% | 125% of the weight on a bread roll |
| Fat that is spread (thick amount) onto large bap: ticked butter, low fat | bread_large_bap_spread_butter_lowfat_thick | 15 | bread_large_bap_spread_butter_lowfat_thick | 18.75 | 25% | 125% of the weight on a bread roll |
| Fat that is spread (thin amount) onto large bap: ticked butter, low fat | bread_large_bap_spread_butter_lowfat_thin | 10 | bread_large_bap_spread_butter_lowfat_thin | 12.5 | 25% | 125% of the weight on a bread roll |
| Fat that is spread (medium amount) onto large bap: ticked spreadable butter with normal amount of % fat | bread_large_bap_spread_butter_spread_fat_med | 12 | bread_large_bap_spread_butter_spread_fat_med | 15 | 25% | 125% of the weight on a bread roll |
| Fat that is spread (thick amount) onto large bap: ticked spreadable butter with normal amount of % fat | bread_large_bap_spread_butter_spread_fat_thick | 15 | bread_large_bap_spread_butter_spread_fat_thick | 18.75 | 25% | 125% of the weight on a bread roll |
| Fat that is spread (thin amount) onto large bap: ticked spreadable butter with normal amount of % fat | bread_large_bap_spread_butter_spread_fat_thin | 10 | bread_large_bap_spread_butter_spread_fat_thin | 12.5 | 25% | 125% of the weight on a bread roll |
| Fat that is spread (medium amount) onto large bap: ticked spreadable butter, low fat | bread_large_bap_spread_butter_spread_lowfat_m | 12 | bread_large_bap_spread_butter_spread_lowfat_m | 15 | 25% | 125% of the weight on a bread roll |
| Fat that is spread (thick amount) onto large bap: ticked spreadable butter, low fat | bread_large_bap_spread_butter_spread_lowfat_th | 15 | bread_large_bap_spread_butter_spread_lowfat_th | 18.75 | 25% | 125% of the weight on a bread roll |
| Fat that is spread (thin amount) onto large bap: ticked spreadable butter, low fat | bread_large_bap_spread_butter_spread_lowfat_th | 10 | bread_large_bap_spread_butter_spread_lowfat_th | 12.5 | 25% | 125% of the weight on a bread roll |
| Fat that is spread (medium amount) onto large bap: ticked dairy spread which is also cholesterol lowering e.g Benecol Buttery | bread_large_bap_spread_dairy_chol_med | 10 | bread_large_bap_spread_dairy_chol_med | 12.5 | 25% | 125% of the weight on a bread roll |
| Fat that is spread (thick amount) onto large bap: ticked dairy spread which is also cholesterol lowering e.g Benecol Buttery | bread_large_bap_spread_dairy_chol_thick | 12 | bread_large_bap_spread_dairy_chol_thick | 15 | 25% | 125% of the weight on a bread roll |
| Fat that is spread (thin amount) onto large bap: ticked dairy spread which is also cholesterol lowering e.g Benecol Buttery | bread_large_bap_spread_dairy_chol_thin | 7 | bread_large_bap_spread_dairy_chol_thin | 8.75 | 25% | 125% of the weight on a bread roll |
| Fat that is spread (medium amount) onto large bap: ticked dairy spread but not specified amount of % fat | bread_large_bap_spread_dairy_dunno_med | 10 | bread_large_bap_spread_dairy_dunno_med | 12.5 | 25% | 125% of the weight on a bread roll; food composition as low fat and reduced fat |
| Fat that is spread (thick amount) onto large bap: ticked dairy spread but not specified amount of % fat | bread_large_bap_spread_dairy_dunno_thick | 12 | bread_large_bap_spread_dairy_dunno_thick | 15 | 25% | 125% of the weight on a bread roll; food composition as low fat and reduced fat |
| Fat that is spread (thin amount) onto large bap: ticked dairy spread but not specified amount of % fat | bread_large_bap_spread_dairy_dunno_thin | 7 | bread_large_bap_spread_dairy_dunno_thin | 8.75 | 25% | 125% of the weight on a bread roll; food composition as low fat and reduced fat |
| Fat that is spread (medium amount) onto large bap: ticked dairy spread with normal amount of % fat | bread_large_bap_spread_dairy_fat_med | 10 | bread_large_bap_spread_dairy_fat_med | 12.5 | 25% | 125% of the weight on a bread roll; normal fat is taken as reduced fat (up to 62% fat) |
| Fat that is spread (thick amount) onto large bap: ticked dairy spread with normal amount of % fat | bread_large_bap_spread_dairy_fat_thick | 12 | bread_large_bap_spread_dairy_fat_thick | 15 | 25% | 125% of the weight on a bread roll; normal fat is taken as reduced fat (up to 62% fat) |
| Fat that is spread (thin amount) onto large bap: ticked dairy spread with normal amount of % fat | bread_large_bap_spread_dairy_fat_thin | 7 | bread_large_bap_spread_dairy_fat_thin | 8.75 | 25% | 125% of the weight on a bread roll; normal fat is taken as reduced fat (up to 62% fat) |
| Fat that is spread (medium amount) onto large bap: ticked dairy spread, low fat | bread_large_bap_spread_dairy_lowfat_med | 10 | bread_large_bap_spread_dairy_lowfat_med | 12.5 | 25% | 125% of the weight on a bread roll |
| Fat that is spread (thick amount) onto large bap: ticked dairy spread, low fat | bread_large_bap_spread_dairy_lowfat_thick | 12 | bread_large_bap_spread_dairy_lowfat_thick | 15 | 25% | 125% of the weight on a bread roll |
| Fat that is spread (thin amount) onto large bap: ticked dairy spread, low fat | bread_large_bap_spread_dairy_lowfat_thin | 7 | bread_large_bap_spread_dairy_lowfat_thin | 8.75 | 25% | 125% of the weight on a bread roll |
| Fat that is spread (medium amount) onto large bap: ticked dairy spread, very low fat | bread_large_bap_spread_dairy_vlowfat_med | 10 | bread_large_bap_spread_dairy_vlowfat_med | 12.5 | 25% | 125% of the weight on a bread roll; food composition as low fat spread |
| Fat that is spread (thick amount) onto large bap: ticked dairy spread, very low fat | bread_large_bap_spread_dairy_vlowfat_thick | 12 | bread_large_bap_spread_dairy_vlowfat_thick | 15 | 25% | 125% of the weight on a bread roll; food composition as low fat spread |
| Fat that is spread (thin amount) onto large bap: ticked dairy spread, very low fat | bread_large_bap_spread_dairy_vlowfat_thin | 7 | bread_large_bap_spread_dairy_vlowfat_thin | 8.75 | 25% | 125% of the weight on a bread roll; food composition as low fat spread |
| Fat that is spread (medium amount) onto large bap: not specified type of spread but ticked cholesterol lowering e.g Benecol, Flora pro active | bread_large_bap_spread_dunno_chol_med | 10 | bread_large_bap_spread_dunno_chol_med | 12.5 | 25% | 125% of the weight on a bread roll |
| Fat that is spread (thick amount) onto large bap: not specified type of spread but ticked cholesterol lowering e.g Benecol, Flora pro active | bread_large_bap_spread_dunno_chol_thick | 12 | bread_large_bap_spread_dunno_chol_thick | 15 | 25% | 125% of the weight on a bread roll |
| Fat that is spread (thin amount) onto large bap: not specified type of spread but ticked cholesterol lowering e.g Benecol, Flora pro active | bread_large_bap_spread_dunno_chol_thin | 7 | bread_large_bap_spread_dunno_chol_thin | 8.75 | 25% | 125% of the weight on a bread roll |
| Fat that is spread (medium amount) onto large bap: not specified type of spread or amount of % fat | bread_large_bap_spread_dunno_dunno_med | 10 | bread_large_bap_spread_dunno_dunno_med | 12.5 | 25% | 125% of the weight on a bread roll |
| Fat that is spread (thick amount) onto large bap: not specified type of spread or amount of % fat | bread_large_bap_spread_dunno_dunno_thick | 12 | bread_large_bap_spread_dunno_dunno_thick | 15 | 25% | 125% of the weight on a bread roll |
| Fat that is spread (thin amount) onto large bap: not specified type of spread or amount of % fat | bread_large_bap_spread_dunno_dunno_thin | 7 | bread_large_bap_spread_dunno_dunno_thin | 8.75 | 25% | 125% of the weight on a bread roll |
| Fat that is spread (medium amount) onto large bap: not specified type of spread but ticked normal amount % fat | bread_large_bap_spread_dunno_fat_med | 10 | bread_large_bap_spread_dunno_fat_med | 12.5 | 25% | 125% of the weight on a bread roll; normal fat is taken as reduced fat (up to 62% fat) |
| Fat that is spread (thick amount) onto large bap: not specified type of spread but ticked normal amount % fat | bread_large_bap_spread_dunno_fat_thick | 12 | bread_large_bap_spread_dunno_fat_thick | 15 | 25% | 125% of the weight on a bread roll; normal fat is taken as reduced fat (up to 62% fat) |
| Fat that is spread (thin amount) onto large bap: not specified type of spread but ticked normal amount % fat | bread_large_bap_spread_dunno_fat_thin | 7 | bread_large_bap_spread_dunno_fat_thin | 8.75 | 25% | 125% of the weight on a bread roll; normal fat is taken as reduced fat (up to 62% fat) |
| Fat that is spread (medium amount) onto large bap: not specified type of spread but ticked low fat | bread_large_bap_spread_dunno_lowfat_med | 10 | bread_large_bap_spread_dunno_lowfat_med | 12.5 | 25% | 125% of the weight on a bread roll |
| Fat that is spread (thick amount) onto large bap: not specified type of spread but ticked low fat | bread_large_bap_spread_dunno_lowfat_thick | 12 | bread_large_bap_spread_dunno_lowfat_thick | 15 | 25% | 125% of the weight on a bread roll |
| Fat that is spread (thin amount) onto large bap: not specified type of spread but ticked low fat | bread_large_bap_spread_dunno_lowfat_thin | 7 | bread_large_bap_spread_dunno_lowfat_thin | 8.75 | 25% | 125% of the weight on a bread roll |
| Fat that is spread (medium amount) onto large bap: not specified type of spread but ticked very low fat | bread_large_bap_spread_dunno_vlowfat_med | 10 | bread_large_bap_spread_dunno_vlowfat_med | 12.5 | 25% | 125% of the weight on a bread roll; food composition as low fat spread |
| Fat that is spread (thick amount) onto large bap: not specified type of spread but ticked very low fat | bread_large_bap_spread_dunno_vlowfat_thick | 12 | bread_large_bap_spread_dunno_vlowfat_thick | 15 | 25% | 125% of the weight on a bread roll; food composition as low fat spread |
| Fat that is spread (thin amount) onto large bap: not specified type of spread but ticked very low fat | bread_large_bap_spread_dunno_vlowfat_thin | 7 | bread_large_bap_spread_dunno_vlowfat_thin | 8.75 | 25% | 125% of the weight on a bread roll; food composition as low fat spread |
| Fat that is spread (medium amount) onto large bap: ticked hard margarine (hard block margarine in wrapper) | bread_large_bap_spread_hardmarg_med | 12 | bread_large_bap_spread_hardmarg_med | 15 | 25% | 125% of the weight on a bread roll, spread weight as butter |
| Fat that is spread (thick amount) onto large bap: ticked hard margarine (hard block margarine in wrapper) | bread_large_bap_spread_hardmarg_thick | 15 | bread_large_bap_spread_hardmarg_thick | 18.75 | 25% | 125% of the weight on a bread roll, spread weight as butter |
| Fat that is spread (thin amount) onto large bap: ticked hard margarine (hard block margarine in wrapper) | bread_large_bap_spread_hardmarg_thin | 10 | bread_large_bap_spread_hardmarg_thin | 12.5 | 25% | 125% of the weight on a bread roll, spread weight as butter |
| Fat that is spread (medium amount) onto large bap: ticked cholesterol lowering olive spread e.g Benecol/Flora pro active olive spread | bread_large_bap_spread_olive_chol_med | 10 | bread_large_bap_spread_olive_chol_med | 12.5 | 25% | 125% of the weight on a bread roll |
| Fat that is spread (thick amount) onto large bap: ticked cholesterol lowering olive spread e.g Benecol/Flora pro active olive spread | bread_large_bap_spread_olive_chol_thick | 12 | bread_large_bap_spread_olive_chol_thick | 15 | 25% | 125% of the weight on a bread roll |
| Fat that is spread (thin amount) onto large bap: ticked cholesterol lowering olive spread e.g Benecol/Flora pro active olive spread | bread_large_bap_spread_olive_chol_thin | 7 | bread_large_bap_spread_olive_chol_thin | 8.75 | 25% | 125% of the weight on a bread roll |
| Fat that is spread (medium amount) onto large bap: ticked olive spread but not specified amount of fat | bread_large_bap_spread_olive_dunno_med | 10 | bread_large_bap_spread_olive_dunno_med | 12.5 | 25% | 125% of the weight on a bread roll |
| Fat that is spread (thick amount) onto large bap: ticked olive spread but not specified amount of fat | bread_large_bap_spread_olive_dunno_thick | 12 | bread_large_bap_spread_olive_dunno_thick | 15 | 25% | 125% of the weight on a bread roll |
| Fat that is spread (thin amount) onto large bap: ticked olive spread but not specified amount of fat | bread_large_bap_spread_olive_dunno_thin | 7 | bread_large_bap_spread_olive_dunno_thin | 8.75 | 25% | 125% of the weight on a bread roll |
| Fat that is spread (medium amount) onto large bap: ticked olive spread with normal amount of % fat | bread_large_bap_spread_olive_fat_med | 10 | bread_large_bap_spread_olive_fat_med | 12.5 | 25% | 125% of the weight on a bread roll; normal fat is taken as reduced fat (up to 62% fat) |
| Fat that is spread (thick amount) onto large bap: ticked olive spread with normal amount of % fat | bread_large_bap_spread_olive_fat_thick | 12 | bread_large_bap_spread_olive_fat_thick | 15 | 25% | 125% of the weight on a bread roll; normal fat is taken as reduced fat (up to 62% fat) |
| Fat that is spread (thin amount) onto large bap: ticked olive spread with normal amount of % fat | bread_large_bap_spread_olive_fat_thin | 7 | bread_large_bap_spread_olive_fat_thin | 8.75 | 25% | 125% of the weight on a bread roll; normal fat is taken as reduced fat (up to 62% fat) |
| Fat that is spread (medium amount) onto large bap: ticked olive spread, low fat | bread_large_bap_spread_olive_lowfat_med | 10 | bread_large_bap_spread_olive_lowfat_med | 12.5 | 25% | 125% of the weight on a bread roll |
| Fat that is spread (thick amount) onto large bap: ticked olive spread, low fat | bread_large_bap_spread_olive_lowfat_thick | 12 | bread_large_bap_spread_olive_lowfat_thick | 15 | 25% | 125% of the weight on a bread roll |
| Fat that is spread (thin amount) onto large bap: ticked olive spread, low fat | bread_large_bap_spread_olive_lowfat_thin | 7 | bread_large_bap_spread_olive_lowfat_thin | 8.75 | 25% | 125% of the weight on a bread roll |
| Fat that is spread (medium amount) onto large bap: ticked olive spread, very low fat | bread_large_bap_spread_olive_vlowfat_med | 10 | bread_large_bap_spread_olive_vlowfat_med | 12.5 | 25% | 125% of the weight on a bread roll; food composition as low fat spread |
| Fat that is spread (thick amount) onto large bap: ticked olive spread, very low fat | bread_large_bap_spread_olive_vlowfat_thick | 12 | bread_large_bap_spread_olive_vlowfat_thick | 15 | 25% | 125% of the weight on a bread roll; food composition as low fat spread |
| Fat that is spread (thin amount) onto large bap: ticked olive spread, very low fat | bread_large_bap_spread_olive_vlowfat_thin | 7 | bread_large_bap_spread_olive_vlowfat_thin | 8.75 | 25% | 125% of the weight on a bread roll; food composition as low fat spread |
| Fat that is spread (medium amount) onto large bap: ticked other spread e.g ghee | bread_large_bap_spread_other_med | 10 | bread_large_bap_spread_other_med | 12.5 | 25% | 125% of the weight on a bread roll |
| Fat that is spread (thick amount) onto large bap: ticked other spread e.g ghee | bread_large_bap_spread_other_thick | 12 | bread_large_bap_spread_other_thick | 15 | 25% | 125% of the weight on a bread roll |
| Fat that is spread (thin amount) onto large bap: ticked other spread e.g ghee | bread_large_bap_spread_other_thin | 7 | bread_large_bap_spread_other_thin | 8.75 | 25% | 125% of the weight on a bread roll |
| Fat that is spread (medium amount) onto large bap: ticked polyunsaturated margarine (e.g Flora) and also cholesterol lowering | bread_large_bap_spread_polymarg_chol_med | 10 | bread_large_bap_spread_polymarg_chol_med | 12.5 | 25% | 125% of the weight on a bread roll |
| Fat that is spread (thick amount) onto large bap: ticked polyunsaturated margarine (e.g Flora) and also cholesterol lowering | bread_large_bap_spread_polymarg_chol_thick | 12 | bread_large_bap_spread_polymarg_chol_thick | 15 | 25% | 125% of the weight on a bread roll |
| Fat that is spread (thin amount) onto large bap: ticked polyunsaturated margarine (e.g Flora) and also cholesterol lowering | bread_large_bap_spread_polymarg_chol_thin | 7 | bread_large_bap_spread_polymarg_chol_thin | 8.75 | 25% | 125% of the weight on a bread roll |
| Fat that is spread (medium amount) onto large bap: ticked polyunsaturated margarine (e.g Flora) but not specified amount of fat | bread_large_bap_spread_polymarg_dunno_med | 10 | bread_large_bap_spread_polymarg_dunno_med | 12.5 | 25% | 125% of the weight on a bread roll |
| Fat that is spread (thick amount) onto large bap: ticked polyunsaturated margarine (e.g Flora) but not specified amount of fat | bread_large_bap_spread_polymarg_dunno_thick | 12 | bread_large_bap_spread_polymarg_dunno_thick | 15 | 25% | 125% of the weight on a bread roll |
| Fat that is spread (thin amount) onto large bap: ticked polyunsaturated margarine (e.g Flora) but not specified amount of fat | bread_large_bap_spread_polymarg_dunno_thin | 7 | bread_large_bap_spread_polymarg_dunno_thin | 8.75 | 25% | 125% of the weight on a bread roll |
| Fat that is spread (medium amount) onto large bap: ticked polyunsaturated margarine (e.g Flora), normal amount of % fat | bread_large_bap_spread_polymarg_fat_med | 10 | bread_large_bap_spread_polymarg_fat_med | 12.5 | 25% | 125% of the weight on a bread roll; normal fat is taken as reduced fat (up to 62% fat) |
| Fat that is spread (thick amount) onto large bap: ticked polyunsaturated margarine (e.g Flora), normal amount of % fat | bread_large_bap_spread_polymarg_fat_thick | 12 | bread_large_bap_spread_polymarg_fat_thick | 15 | 25% | 125% of the weight on a bread roll; normal fat is taken as reduced fat (up to 62% fat) |
| Fat that is spread (thin amount) onto large bap: ticked polyunsaturated margarine (e.g Flora), normal amount of % fat | bread_large_bap_spread_polymarg_fat_thin | 7 | bread_large_bap_spread_polymarg_fat_thin | 8.75 | 25% | 125% of the weight on a bread roll; normal fat is taken as reduced fat (up to 62% fat) |
| Fat that is spread (medium amount) onto large bap: ticked polyunsaturated margarine (e.g Flora), low fat | bread_large_bap_spread_polymarg_lowfat_med | 10 | bread_large_bap_spread_polymarg_lowfat_med | 12.5 | 25% | 125% of the weight on a bread roll |
| Fat that is spread (thick amount) onto large bap: ticked polyunsaturated margarine (e.g Flora), low fat | bread_large_bap_spread_polymarg_lowfat_thick | 12 | bread_large_bap_spread_polymarg_lowfat_thick | 15 | 25% | 125% of the weight on a bread roll |
| Fat that is spread (thin amount) onto large bap: ticked polyunsaturated margarine (e.g Flora), low fat | bread_large_bap_spread_polymarg_lowfat_thin | 7 | bread_large_bap_spread_polymarg_lowfat_thin | 8.75 | 25% | 125% of the weight on a bread roll |
| Fat that is spread (medium amount) onto large bap: ticked polyunsaturated margarine (e.g Flora), very low fat | bread_large_bap_spread_polymarg_vlowfat_med | 10 | bread_large_bap_spread_polymarg_vlowfat_med | 12.5 | 25% | 125% of the weight on a bread roll; food composition as low fat spread |
| Fat that is spread (thick amount) onto large bap: ticked polyunsaturated margarine (e.g Flora), very low fat | bread_large_bap_spread_polymarg_vlowfat_thick | 12 | bread_large_bap_spread_polymarg_vlowfat_thick | 15 | 25% | 125% of the weight on a bread roll; food composition as low fat spread |
| Fat that is spread (thin amount) onto large bap: ticked polyunsaturated margarine (e.g Flora), very low fat | bread_large_bap_spread_polymarg_vlowfat_thin | 7 | bread_large_bap_spread_polymarg_vlowfat_thin | 8.75 | 25% | 125% of the weight on a bread roll; food composition as low fat spread |
| Fat that is spread (medium amount) onto large bap: ticked soya/vegan/dairy free margarine e.g Pure, and also cholesterol lowering | bread_large_bap_spread_soya_chol_med | 10 | bread_large_bap_spread_soya_chol_med | 12.5 | 25% | 125% of the weight on a bread roll |
| Fat that is spread (thick amount) onto large bap: ticked soya/vegan/dairy free margarine e.g Pure, and also cholesterol lowering | bread_large_bap_spread_soya_chol_thick | 12 | bread_large_bap_spread_soya_chol_thick | 15 | 25% | 125% of the weight on a bread roll |
| Fat that is spread (thin amount) onto large bap: ticked soya/vegan/dairy free margarine e.g Pure, and also cholesterol lowering | bread_large_bap_spread_soya_chol_thin | 7 | bread_large_bap_spread_soya_chol_thin | 8.75 | 25% | 125% of the weight on a bread roll |
| Fat that is spread (medium amount) onto large bap: ticked soya/vegan/dairy free margarine e.g Pure, and not specified amount of fat | bread_large_bap_spread_soya_dunno_med | 10 | bread_large_bap_spread_soya_dunno_med | 12.5 | 25% | 125% of the weight on a bread roll |
| Fat that is spread (thick amount) onto large bap: ticked soya/vegan/dairy free margarine e.g Pure, and not specified amount of fat | bread_large_bap_spread_soya_dunno_thick | 12 | bread_large_bap_spread_soya_dunno_thick | 15 | 25% | 125% of the weight on a bread roll |
| Fat that is spread (thin amount) onto large bap: ticked soya/vegan/dairy free margarine e.g Pure, and not specified amount of fat | bread_large_bap_spread_soya_dunno_thin | 7 | bread_large_bap_spread_soya_dunno_thin | 8.75 | 25% | 125% of the weight on a bread roll |
| Fat that is spread (medium amount) onto large bap: ticked soya/vegan/dairy free margarine (e.g Pure), normal amount % fat | bread_large_bap_spread_soya_fat_med | 10 | bread_large_bap_spread_soya_fat_med | 12.5 | 25% | 125% of the weight on a bread roll; normal fat is taken as reduced fat (up to 62% fat) |
| Fat that is spread (thick amount) onto large bap: ticked soya/vegan/dairy free margarine (e.g Pure), normal amount % fat | bread_large_bap_spread_soya_fat_thick | 12 | bread_large_bap_spread_soya_fat_thick | 15 | 25% | 125% of the weight on a bread roll; normal fat is taken as reduced fat (up to 62% fat) |
| Fat that is spread (thin amount) onto large bap: ticked soya/vegan/dairy free margarine (e.g Pure), normal amount % fat | bread_large_bap_spread_soya_fat_thin | 7 | bread_large_bap_spread_soya_fat_thin | 8.75 | 25% | 125% of the weight on a bread roll; normal fat is taken as reduced fat (up to 62% fat) |
| Fat that is spread (medium amount) onto large bap: ticked soya/vegan/dairy free margarine (e.g Pure), low fat | bread_large_bap_spread_soya_lowfat_med | 10 | bread_large_bap_spread_soya_lowfat_med | 12.5 | 25% | 125% of the weight on a bread roll |
| Fat that is spread (thick amount) onto large bap: ticked soya/vegan/dairy free margarine (e.g Pure), low fat | bread_large_bap_spread_soya_lowfat_thick | 12 | bread_large_bap_spread_soya_lowfat_thick | 15 | 25% | 125% of the weight on a bread roll |
| Fat that is spread (thin amount) onto large bap: ticked soya/vegan/dairy free margarine (e.g Pure), low fat | bread_large_bap_spread_soya_lowfat_thin | 7 | bread_large_bap_spread_soya_lowfat_thin | 8.75 | 25% | 125% of the weight on a bread roll |
| Fat that is spread (medium amount) onto large bap: ticked soya/vegan/dairy free margarine (e.g Pure), very low fat | bread_large_bap_spread_soya_vlowfat_med | 10 | bread_large_bap_spread_soya_vlowfat_med | 12.5 | 25% | 125% of the weight on a bread roll; food composition as low fat spread |
| Fat that is spread (thick amount) onto large bap: ticked soya/vegan/dairy free margarine (e.g Pure), very low fat | bread_large_bap_spread_soya_vlowfat_thick | 12 | bread_large_bap_spread_soya_vlowfat_thick | 15 | 25% | 125% of the weight on a bread roll; food composition as low fat spread |
| Fat that is spread (thin amount) onto large bap: ticked soya/vegan/dairy free margarine (e.g Pure), very low fat | bread_large_bap_spread_soya_vlowfat_thin | 7 | bread_large_bap_spread_soya_vlowfat_thin | 8.75 | 25% | 125% of the weight on a bread roll; food composition as low fat spread |

| | McCance and Widdowson's | | Nutrient databank + other changes | | | |
|---|---|---|---|---|---|---|
| Variable description | Food item | Portion size | Food item | Portion size | Portion diff% | Description of differences with previous version |
| Large bap, stotty, pitta bread: not specified type so includes white, brown, granary, wheatgerm, wholemeal | bread_large_bap_unanswered | 90 | bread_large_bap_unanswered | 90 | 0% | |
| Large bap, stotty, pitta bread: white | bread_large_bap_white | 90 | bread_large_bap_white | 90 | 0% | |
| Large bap, stotty, pitta bread: wholemeal | bread_large_bap_wholemeal | 90 | bread_large_bap_wholemeal | 90 | 0% | |
| Naan bread plain | bread_naan | 160 | bread_naan | 160 | 0% | |
| Other bread e.g crumpets, tortilla wraps, breadsticks | bread_other | 45 | bread_other | 45 | 0% | |
| Other bread e.g crumpets, tortilla wraps, breadsticks | | | bread_other_gf | 45 | - | Gluten free version added to the updated version (however no gluten free code available at the time so the non-gluten free code was used) |
| Fat that is spread (medium amount) onto bread other: ticked butter but not specified amount of % fat | bread_other_spread_butter_dunno_med | 7 | bread_other_spread_butter_dunno_med | 12 | 71% | as weight on a bread roll; food composition as butter and spreadable butter |
| Fat that is spread (thick amount) onto bread other: ticked butter but not specified amount of % fat | bread_other_spread_butter_dunno_thick | 10 | bread_other_spread_butter_dunno_thick | 15 | 50% | as weight on a bread roll; food composition as butter and spreadable butter |
| Fat that is spread (thin amount) onto bread other: ticked butter but not specified amount of % fat | bread_other_spread_butter_dunno_thin | 5 | bread_other_spread_butter_dunno_thin | 10 | 100% | as weight on a bread roll; food composition as butter and spreadable butter |
| Fat that is spread (medium amount) onto bread other: ticked butter, normal amount of % fat | bread_other_spread_butter_fat_med | 7 | bread_other_spread_butter_fat_med | 12 | 71% | as weight on a bread roll |
| Fat that is spread (thick amount) onto bread other: ticked butter, normal amount of % fat | bread_other_spread_butter_fat_thick | 10 | bread_other_spread_butter_fat_thick | 15 | 50% | as weight on a bread roll |
| Fat that is spread (thin amount) onto bread other: ticked butter, normal amount of % fat | bread_other_spread_butter_fat_thin | 5 | bread_other_spread_butter_fat_thin | 10 | 100% | as weight on a bread roll |
| Fat that is spread (medium amount) onto bread other: ticked butter, low fat | bread_other_spread_butter_lowfat_med | 7 | bread_other_spread_butter_lowfat_med | 12 | 71% | as weight on a bread roll |
| Fat that is spread (thick amount) onto bread other: ticked butter, low fat | bread_other_spread_butter_lowfat_thick | 10 | bread_other_spread_butter_lowfat_thick | 15 | 50% | as weight on a bread roll |
| Fat that is spread (thin amount) onto bread other: ticked butter, low fat | bread_other_spread_butter_lowfat_thin | 5 | bread_other_spread_butter_lowfat_thin | 10 | 100% | as weight on a bread roll |
| Fat that is spread (medium amount) onto bread other: ticked spreadable butter with normal amount of % fat | bread_other_spread_butter_spread_fat_med | 7 | bread_other_spread_butter_spread_fat_med | 12 | 71% | as weight on a bread roll |
| Fat that is spread (thick amount) onto bread other: ticked spreadable butter with normal amount of % fat | bread_other_spread_butter_spread_fat_thick | 10 | bread_other_spread_butter_spread_fat_thick | 15 | 50% | as weight on a bread roll |
| Fat that is spread (thin amount) onto bread other: ticked spreadable butter with normal amount of % fat | bread_other_spread_butter_spread_fat_thin | 5 | bread_other_spread_butter_spread_fat_thin | 10 | 100% | as weight on a bread roll |
| Fat that is spread (medium amount) onto bread other: ticked spreadable butter, low fat | bread_other_spread_butter_spread_lowfat_med | 7 | bread_other_spread_butter_spread_lowfat_med | 12 | 71% | as weight on a bread roll |
| Fat that is spread (thick amount) onto bread other: ticked spreadable butter, low fat | bread_other_spread_butter_spread_lowfat_thick | 10 | bread_other_spread_butter_spread_lowfat_thick | 15 | 50% | as weight on a bread roll |
| Fat that is spread (thin amount) onto bread other: ticked spreadable butter, low fat | bread_other_spread_butter_spread_lowfat_thin | 5 | bread_other_spread_butter_spread_lowfat_thin | 10 | 100% | as weight on a bread roll |
| Fat that is spread (medium amount) onto bread other: ticked dairy spread which is also cholesterol lowering e.g Benecol Buttery | bread_other_spread_dairy_chol_med | 5 | bread_other_spread_dairy_chol_med | 10 | 100% | as weight on a bread roll |
| Fat that is spread (thick amount) onto bread other: ticked dairy spread which is also cholesterol lowering e.g Benecol Buttery | bread_other_spread_dairy_chol_thick | 7 | bread_other_spread_dairy_chol_thick | 12 | 71% | as weight on a bread roll |
| Fat that is spread (thin amount) onto bread other: ticked dairy spread which is also cholesterol lowering e.g Benecol Buttery | bread_other_spread_dairy_chol_thin | 3 | bread_other_spread_dairy_chol_thin | 7 | 133% | as weight on a bread roll |
| Fat that is spread (medium amount) onto bread other: ticked dairy spread but not specified amount of % fat | bread_other_spread_dairy_dunno_med | 5 | bread_other_spread_dairy_dunno_med | 10 | 100% | as weight on a bread roll; food composition as low fat and reduced fat |
| Fat that is spread (thick amount) onto bread other: ticked dairy spread but not specified amount of % fat | bread_other_spread_dairy_dunno_thick | 7 | bread_other_spread_dairy_dunno_thick | 12 | 71% | as weight on a bread roll; food composition as low fat and reduced fat |
| Fat that is spread (thin amount) onto bread other: ticked dairy spread but not specified amount of % fat | bread_other_spread_dairy_dunno_thin | 3 | bread_other_spread_dairy_dunno_thin | 7 | 133% | as weight on a bread roll; food composition as low fat and reduced fat |
| Fat that is spread (medium amount) onto bread other: ticked dairy spread with normal amount of % fat | bread_other_spread_dairy_fat_med | 5 | bread_other_spread_dairy_fat_med | 10 | 100% | as weight on a bread roll; normal fat is taken as reduced fat (up to 62% fat) |
| Fat that is spread (thick amount) onto bread other: ticked dairy spread with normal amount of % fat | bread_other_spread_dairy_fat_thick | 7 | bread_other_spread_dairy_fat_thick | 12 | 71% | as weight on a bread roll; normal fat is taken as reduced fat (up to 62% fat) |
| Fat that is spread (thin amount) onto bread other: ticked dairy spread with normal amount of % fat | bread_other_spread_dairy_fat_thin | 3 | bread_other_spread_dairy_fat_thin | 7 | 133% | as weight on a bread roll; normal fat is taken as reduced fat (up to 62% fat) |
| Fat that is spread (medium amount) onto bread other: ticked dairy spread, low fat | bread_other_spread_dairy_lowfat_med | 5 | bread_other_spread_dairy_lowfat_med | 10 | 100% | as weight on a bread roll |
| Fat that is spread (thick amount) onto bread other: ticked dairy spread, low fat | bread_other_spread_dairy_lowfat_thick | 7 | bread_other_spread_dairy_lowfat_thick | 12 | 71% | as weight on a bread roll |
| Fat that is spread (thin amount) onto bread other: ticked dairy spread, low fat | bread_other_spread_dairy_lowfat_thin | 3 | bread_other_spread_dairy_lowfat_thin | 7 | 133% | as weight on a bread roll |
| Fat that is spread (medium amount) onto bread other: ticked dairy spread, very low fat | bread_other_spread_dairy_vlowfat_med | 5 | bread_other_spread_dairy_vlowfat_med | 10 | 100% | as weight on a bread roll; food composition as low fat spread |
| Fat that is spread (thick amount) onto bread other: ticked dairy spread, very low fat | bread_other_spread_dairy_vlowfat_thick | 7 | bread_other_spread_dairy_vlowfat_thick | 12 | 71% | as weight on a bread roll; food composition as low fat spread |
| Fat that is spread (thin amount) onto bread other: ticked dairy spread, very low fat | bread_other_spread_dairy_vlowfat_thin | 3 | bread_other_spread_dairy_vlowfat_thin | 7 | 133% | as weight on a bread roll; food composition as low fat spread |
| Fat that is spread (medium amount) onto bread other: not specified type of spread but ticked cholesterol lowering e.g Benecol, Flora pro active | bread_other_spread_dunno_chol_med | 5 | bread_other_spread_dunno_chol_med | 10 | 100% | as weight on a bread roll |
| Fat that is spread (thick amount) onto bread other: not specified type of spread but ticked cholesterol lowering e.g Benecol, Flora pro active | bread_other_spread_dunno_chol_thick | 7 | bread_other_spread_dunno_chol_thick | 12 | 71% | as weight on a bread roll |
| Fat that is spread (thin amount) onto bread other: not specified type of spread but ticked cholesterol lowering e.g Benecol, Flora pro active | bread_other_spread_dunno_chol_thin | 3 | bread_other_spread_dunno_chol_thin | 7 | 133% | as weight on a bread roll |
| Fat that is spread (medium amount) onto bread other: not specified type of spread or amount of % fat | bread_other_spread_dunno_dunno_med | 5 | bread_other_spread_dunno_dunno_med | 10 | 100% | as weight on a bread roll |
| Fat that is spread (thick amount) onto bread other: not specified type of spread or amount of % fat | bread_other_spread_dunno_dunno_thick | 7 | bread_other_spread_dunno_dunno_thick | 12 | 71% | as weight on a bread roll |
| Fat that is spread (thin amount) onto bread other: not specified type of spread or amount of % fat | bread_other_spread_dunno_dunno_thin | 3 | bread_other_spread_dunno_dunno_thin | 7 | 133% | as weight on a bread roll |
| Fat that is spread (medium amount) onto bread other: not specified type of spread but ticked normal amount % fat | bread_other_spread_dunno_fat_med | 5 | bread_other_spread_dunno_fat_med | 10 | 100% | as weight on a bread roll; normal fat is taken as reduced fat (up to 62% fat) |
| Fat that is spread (thick amount) onto bread other: not specified type of spread but ticked normal amount % fat | bread_other_spread_dunno_fat_thick | 7 | bread_other_spread_dunno_fat_thick | 12 | 71% | as weight on a bread roll; normal fat is taken as reduced fat (up to 62% fat) |
| Fat that is spread (thin amount) onto bread other: not specified type of spread but ticked normal amount % fat | bread_other_spread_dunno_fat_thin | 3 | bread_other_spread_dunno_fat_thin | 7 | 133% | as weight on a bread roll; normal fat is taken as reduced fat (up to 62% fat) |
| Fat that is spread (medium amount) onto bread other: not specified type of spread but ticked low fat | bread_other_spread_dunno_lowfat_med | 5 | bread_other_spread_dunno_lowfat_med | 10 | 100% | as weight on a bread roll |
| Fat that is spread (thick amount) onto bread other: not specified type of spread but ticked low fat | bread_other_spread_dunno_lowfat_thick | 7 | bread_other_spread_dunno_lowfat_thick | 12 | 71% | as weight on a bread roll |
| Fat that is spread (thin amount) onto bread other: not specified type of spread but ticked low fat | bread_other_spread_dunno_lowfat_thin | 3 | bread_other_spread_dunno_lowfat_thin | 7 | 133% | as weight on a bread roll |
| Fat that is spread (medium amount) onto bread other: not specified type of spread but ticked very low fat | bread_other_spread_dunno_vlowfat_med | 5 | bread_other_spread_dunno_vlowfat_med | 10 | 100% | as weight on a bread roll; food composition as low fat spread |
| Fat that is spread (thick amount) onto bread other: not specified type of spread but ticked very low fat | bread_other_spread_dunno_vlowfat_thick | 7 | bread_other_spread_dunno_vlowfat_thick | 12 | 71% | as weight on a bread roll; food composition as low fat spread |
| Fat that is spread (thin amount) onto bread other: not specified type of spread but ticked very low fat | bread_other_spread_dunno_vlowfat_thin | 3 | bread_other_spread_dunno_vlowfat_thin | 7 | 133% | as weight on a bread roll; food composition as low fat spread |
| Fat that is spread (medium amount) onto bread other: ticked hard margarine (hard block margarine in wrapper) | bread_other_spread_hardmarg_med | 7 | bread_other_spread_hardmarg_med | 12 | 71% | as weight on a bread roll; spread weight as butter |
| Fat that is spread (thick amount) onto bread other: ticked hard margarine (hard block margarine in wrapper) | bread_other_spread_hardmarg_thick | 10 | bread_other_spread_hardmarg_thick | 15 | 50% | as weight on a bread roll; spread weight as butter |
| Fat that is spread (thin amount) onto bread other: ticked hard margarine (hard block margarine in wrapper) | bread_other_spread_hardmarg_thin | 5 | bread_other_spread_hardmarg_thin | 10 | 100% | as weight on a bread roll; spread weight as butter |
| Fat that is spread (medium amount) onto bread other: ticked cholesterol lowering olive spread e.g Benecol/Flora pro active olive spread | bread_other_spread_olive_chol_med | 5 | bread_other_spread_olive_chol_med | 10 | 100% | as weight on a bread roll |
| Fat that is spread (thick amount) onto bread other: ticked cholesterol lowering olive spread e.g Benecol/Flora pro active olive spread | bread_other_spread_olive_chol_thick | 7 | bread_other_spread_olive_chol_thick | 12 | 71% | as weight on a bread roll |
| Fat that is spread (thin amount) onto bread other: ticked cholesterol lowering olive spread e.g Benecol/Flora pro active olive spread | bread_other_spread_olive_chol_thin | 3 | bread_other_spread_olive_chol_thin | 7 | 133% | as weight on a bread roll |
| Fat that is spread (medium amount) onto bread other: ticked olive spread but not specified amount of fat | bread_other_spread_olive_dunno_med | 5 | bread_other_spread_olive_dunno_med | 10 | 100% | as weight on a bread roll |
| Fat that is spread (thick amount) onto bread other: ticked olive spread but not specified amount of fat | bread_other_spread_olive_dunno_thick | 7 | bread_other_spread_olive_dunno_thick | 12 | 71% | as weight on a bread roll |
| Fat that is spread (thin amount) onto bread other: ticked olive spread but not specified amount of fat | bread_other_spread_olive_dunno_thin | 3 | bread_other_spread_olive_dunno_thin | 7 | 133% | as weight on a bread roll |
| Fat that is spread (medium amount) onto bread other: ticked olive spread with normal amount of % fat | bread_other_spread_olive_fat_med | 5 | bread_other_spread_olive_fat_med | 10 | 100% | as weight on a bread roll; normal fat is taken as reduced fat (up to 62% fat) |
| Fat that is spread (thick amount) onto bread other: ticked olive spread with normal amount of % fat | bread_other_spread_olive_fat_thick | 7 | bread_other_spread_olive_fat_thick | 12 | 71% | as weight on a bread roll; normal fat is taken as reduced fat (up to 62% fat) |
| Fat that is spread (thin amount) onto bread other: ticked olive spread with normal amount of % fat | bread_other_spread_olive_fat_thin | 3 | bread_other_spread_olive_fat_thin | 7 | 133% | as weight on a bread roll; normal fat is taken as reduced fat (up to 62% fat) |
| Fat that is spread (medium amount) onto bread other: ticked olive spread, low fat | bread_other_spread_olive_lowfat_med | 5 | bread_other_spread_olive_lowfat_med | 10 | 100% | as weight on a bread roll |
| Fat that is spread (thick amount) onto bread other: ticked olive spread, low fat | bread_other_spread_olive_lowfat_thick | 7 | bread_other_spread_olive_lowfat_thick | 12 | 71% | as weight on a bread roll |
| Fat that is spread (thin amount) onto bread other: ticked olive spread, low fat | bread_other_spread_olive_lowfat_thin | 3 | bread_other_spread_olive_lowfat_thin | 7 | 133% | as weight on a bread roll |
| Fat that is spread (medium amount) onto bread other: ticked olive spread, very low fat | bread_other_spread_olive_vlowfat_med | 5 | bread_other_spread_olive_vlowfat_med | 10 | 100% | as weight on a bread roll; food composition as low fat spread |
| Fat that is spread (thick amount) onto bread other: ticked olive spread, very low fat | bread_other_spread_olive_vlowfat_thick | 7 | bread_other_spread_olive_vlowfat_thick | 12 | 71% | as weight on a bread roll; food composition as low fat spread |
| Fat that is spread (thin amount) onto bread other: ticked olive spread, very low fat | bread_other_spread_olive_vlowfat_thin | 3 | bread_other_spread_olive_vlowfat_thin | 7 | 133% | as weight on a bread roll; food composition as low fat spread |
| Fat that is spread (medium amount) onto bread other: ticked other spread e.g ghee | bread_other_spread_other_med | 5 | bread_other_spread_other_med | 10 | 100% | as weight on a bread roll |
| Fat that is spread (thick amount) onto bread other: ticked other spread e.g ghee | bread_other_spread_other_thick | 7 | bread_other_spread_other_thick | 12 | 71% | as weight on a bread roll |
| Fat that is spread (thin amount) onto bread other: ticked other spread e.g ghee | bread_other_spread_other_thin | 3 | bread_other_spread_other_thin | 7 | 133% | as weight on a bread roll |
| Fat that is spread (medium amount) onto bread other: ticked polyunsaturated margarine (e.g Flora) and also cholesterol lowering | bread_other_spread_polymarg_chol_med | 5 | bread_other_spread_polymarg_chol_med | 10 | 100% | as weight on a bread roll |
| Fat that is spread (thick amount) onto bread other: ticked polyunsaturated margarine (e.g Flora) and also cholesterol lowering | bread_other_spread_polymarg_chol_thick | 7 | bread_other_spread_polymarg_chol_thick | 12 | 71% | as weight on a bread roll |
| Fat that is spread (thin amount) onto bread other: ticked polyunsaturated margarine (e.g Flora) and also cholesterol lowering | bread_other_spread_polymarg_chol_thin | 3 | bread_other_spread_polymarg_chol_thin | 7 | 133% | as weight on a bread roll |
| Fat that is spread (medium amount) onto bread other: ticked polyunsaturated margarine (e.g Flora) but not specified amount of fat | bread_other_spread_polymarg_dunno_med | 5 | bread_other_spread_polymarg_dunno_med | 10 | 100% | as weight on a bread roll |
| Fat that is spread (thick amount) onto bread other: ticked polyunsaturated margarine (e.g Flora) but not specified amount of fat | bread_other_spread_polymarg_dunno_thick | 7 | bread_other_spread_polymarg_dunno_thick | 12 | 71% | as weight on a bread roll |
| Fat that is spread (thin amount) onto bread other: ticked polyunsaturated margarine (e.g Flora) but not specified amount of fat | bread_other_spread_polymarg_dunno_thin | 3 | bread_other_spread_polymarg_dunno_thin | 7 | 133% | as weight on a bread roll |
| Fat that is spread (medium amount) onto bread other: ticked polyunsaturated margarine (e.g Flora), normal amount of % fat | bread_other_spread_polymarg_fat_med | 5 | bread_other_spread_polymarg_fat_med | 10 | 100% | as weight on a bread roll; normal fat is taken as reduced fat (up to 62% fat) |
| Fat that is spread (thick amount) onto bread other: ticked polyunsaturated margarine (e.g Flora), normal amount of % fat | bread_other_spread_polymarg_fat_thick | 7 | bread_other_spread_polymarg_fat_thick | 12 | 71% | as weight on a bread roll; normal fat is taken as reduced fat (up to 62% fat) |
| Fat that is spread (thin amount) onto bread other: ticked polyunsaturated margarine (e.g Flora), normal amount of % fat | bread_other_spread_polymarg_fat_thin | 3 | bread_other_spread_polymarg_fat_thin | 7 | 133% | as weight on a bread roll; normal fat is taken as reduced fat (up to 62% fat) |
| Fat that is spread (medium amount) onto bread other: ticked polyunsaturated margarine (e.g Flora), low fat | bread_other_spread_polymarg_lowfat_med | 5 | bread_other_spread_polymarg_lowfat_med | 10 | 100% | as weight on a bread roll |
| Fat that is spread (thick amount) onto bread other: ticked polyunsaturated margarine (e.g Flora), low fat | bread_other_spread_polymarg_lowfat_thick | 7 | bread_other_spread_polymarg_lowfat_thick | 12 | 71% | as weight on a bread roll |
| Fat that is spread (thin amount) onto bread other: ticked polyunsaturated margarine (e.g Flora), low fat | bread_other_spread_polymarg_lowfat_thin | 3 | bread_other_spread_polymarg_lowfat_thin | 7 | 133% | as weight on a bread roll |
| Fat that is spread (medium amount) onto bread other: ticked polyunsaturated margarine (e.g Flora), very low fat | bread_other_spread_polymarg_vlowfat_med | 5 | bread_other_spread_polymarg_vlowfat_med | 10 | 100% | as weight on a bread roll; food composition as low fat spread |
| Fat that is spread (thick amount) onto bread other: ticked polyunsaturated margarine (e.g Flora), very low fat | bread_other_spread_polymarg_vlowfat_thick | 7 | bread_other_spread_polymarg_vlowfat_thick | 12 | 71% | as weight on a bread roll; food composition as low fat spread |
| Fat that is spread (thin amount) onto bread other: ticked polyunsaturated margarine (e.g Flora), very low fat | bread_other_spread_polymarg_vlowfat_thin | 3 | bread_other_spread_polymarg_vlowfat_thin | 7 | 133% | as weight on a bread roll; food composition as low fat spread |
| Fat that is spread (medium amount) onto bread other: ticked soya/vegan/dairy free margarine e.g Pure, and also cholesterol lowering | bread_other_spread_soya_chol_med | 5 | bread_other_spread_soya_chol_med | 10 | 100% | as weight on a bread roll |
| Fat that is spread (thick amount) onto bread other: ticked soya/vegan/dairy free margarine e.g Pure, and also cholesterol lowering | bread_other_spread_soya_chol_thick | 7 | bread_other_spread_soya_chol_thick | 12 | 71% | as weight on a bread roll |
| Fat that is spread (thin amount) onto bread other: ticked soya/vegan/dairy free margarine e.g Pure, and also cholesterol lowering | bread_other_spread_soya_chol_thin | 3 | bread_other_spread_soya_chol_thin | 7 | 133% | as weight on a bread roll |
| Fat that is spread (medium amount) onto bread other: ticked soya/vegan/dairy free margarine e.g Pure, and not specified amount of fat | bread_other_spread_soya_dunno_med | 5 | bread_other_spread_soya_dunno_med | 10 | 100% | as weight on a bread roll |
| Fat that is spread (thick amount) onto bread other: ticked soya/vegan/dairy free margarine e.g Pure, and not specified amount of fat | bread_other_spread_soya_dunno_thick | 7 | bread_other_spread_soya_dunno_thick | 12 | 71% | as weight on a bread roll |
| Fat that is spread (thin amount) onto bread other: ticked soya/vegan/dairy free margarine e.g Pure, and not specified amount of fat | bread_other_spread_soya_dunno_thin | 3 | bread_other_spread_soya_dunno_thin | 7 | 133% | as weight on a bread roll |
| Fat that is spread (medium amount) onto bread other: ticked soya/vegan/dairy free margarine (e.g Pure), normal amount % fat | bread_other_spread_soya_fat_med | 5 | bread_other_spread_soya_fat_med | 10 | 100% | as weight on a bread roll |
| Fat that is spread (thick amount) onto bread other: ticked soya/vegan/dairy free margarine (e.g Pure), normal amount % fat | bread_other_spread_soya_fat_thick | 7 | bread_other_spread_soya_fat_thick | 12 | 71% | as weight on a bread roll |
| Fat that is spread (thin amount) onto bread other: ticked soya/vegan/dairy free margarine (e.g Pure), normal amount % fat | bread_other_spread_soya_fat_thin | 3 | bread_other_spread_soya_fat_thin | 7 | 133% | as weight on a bread roll |
| Fat that is spread (medium amount) onto bread other: ticked soya/vegan/dairy free margarine (e.g Pure), low fat | bread_other_spread_soya_lowfat_med | 5 | bread_other_spread_soya_lowfat_med | 10 | 100% | as weight on a bread roll |
| Fat that is spread (thick amount) onto bread other: ticked soya/vegan/dairy free margarine (e.g Pure), low fat | bread_other_spread_soya_lowfat_thick | 7 | bread_other_spread_soya_lowfat_thick | 12 | 71% | as weight on a bread roll |
| Fat that is spread (thin amount) onto bread other: ticked soya/vegan/dairy free margarine (e.g Pure), low fat | bread_other_spread_soya_lowfat_thin | 3 | bread_other_spread_soya_lowfat_thin | 7 | 133% | as weight on a bread roll |
| Fat that is spread (medium amount) onto bread other: ticked soya/vegan/dairy free margarine (e.g Pure), very low fat | bread_other_spread_soya_vlowfat_med | 5 | bread_other_spread_soya_vlowfat_med | 10 | 100% | as weight on a bread roll; food composition as low fat spread |
| Fat that is spread (thick amount) onto bread other: ticked soya/vegan/dairy free margarine (e.g Pure), very low fat | bread_other_spread_soya_vlowfat_thick | 7 | bread_other_spread_soya_vlowfat_thick | 12 | 71% | as weight on a bread roll; food composition as low fat spread |
| Fat that is spread (thin amount) onto bread other: ticked soya/vegan/dairy free margarine (e.g Pure), very low fat | bread_other_spread_soya_vlowfat_thin | 3 | bread_other_spread_soya_vlowfat_thin | 7 | 133% | as weight on a bread roll; food composition as low fat spread |
| Gluten free bread roll, bap, bun, bagel: brown or white with added fibre | | | bread_roll_gf_nonwhite | 60 | - | Gluten free version added to the updated version |
| Gluten free bread roll, bap, burger bun, hot dog roll, bagel: not specified type so includes white, brown or with added fibre | | | bread_roll_gf_unanswered | 60 | - | Gluten free version added to the updated version |
| Gluten free bread roll, bap, burger bun, hot dog roll, bagel: white | | | bread_roll_gf_white | 60 | - | Gluten free version added to the updated version |
| Bread roll, bap, bun, bagel: brown, granary, wheatgerm, 50/50 | bread_roll_mixed | 60 | bread_roll_mixed | 60 | 0% | |
| Bread roll, bap, bun, bagel: wheatgerm, rye, white sliced | bread_roll_other | 60 | bread_roll_other | 60 | 0% | |
| Seeds for seeded bread: e.g sesame, sunflower, poppy | bread_roll_seeded | 3 | bread_roll_seeded | 3 | 0% | |
| Fat that is spread (medium amount) onto bread roll: ticked butter but not specified amount of % fat | bread_roll_spread_butter_dunno_med | 15 | bread_roll_spread_butter_dunno_med | 12 | -20% | Food portion sizes / MAFF; food composition as butter and spreadable butter; corrected typo in weight |

| | McCance and Widdowson's | | Nutrient databank + other changes | | | |
|---|---|---|---|---|---|---|
| Variable description | Food item | Portion size | Food item | Portion size | Portion diff% | Description of differences with previous version |
| Fat that is spread (thick amount) onto bread roll: ticked butter but not specified amount of % fat | bread_roll_spread_butter_dunno_thick | 15 | bread_roll_spread_butter_dunno_thick | 15 | 0% | Food portion sizes / MAFF; food composition as butter and spreadable butter |
| Fat that is spread (thin amount) onto bread roll: ticked butter but not specified amount of % fat | bread_roll_spread_butter_dunno_thin | 15 | bread_roll_spread_butter_dunno_thin | 10 | -33% | Food portion sizes / MAFF; food composition as butter and spreadable butter; corrected typo in weight |
| Fat that is spread (medium amount) onto bread roll: ticked butter, normal amount of % fat | bread_roll_spread_butter_fat_med | 15 | bread_roll_spread_butter_fat_med | 12 | -20% | Food portion sizes / MAFF; corrected typo in weight |
| Fat that is spread (thick amount) onto bread roll: ticked butter, normal amount of % fat | bread_roll_spread_butter_fat_thick | 15 | bread_roll_spread_butter_fat_thick | 15 | 0% | Food portion sizes / MAFF |
| Fat that is spread (thin amount) onto bread roll: ticked butter, normal amount of % fat | bread_roll_spread_butter_fat_thin | 15 | bread_roll_spread_butter_fat_thin | 10 | -33% | Food portion sizes / MAFF; corrected typo in weight |
| Fat that is spread (medium amount) onto bread roll: ticked butter, low fat | bread_roll_spread_butter_lowfat_med | 15 | bread_roll_spread_butter_lowfat_med | 12 | -20% | Food portion sizes / MAFF; corrected typo in weight |
| Fat that is spread (thick amount) onto bread roll: ticked butter, low fat | bread_roll_spread_butter_lowfat_thick | 15 | bread_roll_spread_butter_lowfat_thick | 15 | 0% | Food portion sizes / MAFF |
| Fat that is spread (thin amount) onto bread roll: ticked butter, low fat | bread_roll_spread_butter_lowfat_thin | 15 | bread_roll_spread_butter_lowfat_thin | 10 | -33% | Food portion sizes / MAFF; corrected typo in weight |
| Fat that is spread (medium amount) onto bread roll: ticked spreadable butter with normal amount of % fat | bread_roll_spread_butter_spread_fat_med | 15 | bread_roll_spread_butter_spread_fat_med | 12 | -20% | Food portion sizes / MAFF; corrected typo in weight |
| Fat that is spread (thick amount) onto bread roll: ticked spreadable butter with normal amount of % fat | bread_roll_spread_butter_spread_fat_thick | 15 | bread_roll_spread_butter_spread_fat_thick | 15 | 0% | Food portion sizes / MAFF |
| Fat that is spread (thin amount) onto bread roll: ticked spreadable butter with normal amount of % fat | bread_roll_spread_butter_spread_fat_thin | 15 | bread_roll_spread_butter_spread_fat_thin | 10 | -33% | Food portion sizes / MAFF; corrected typo in weight |
| Fat that is spread (medium amount) onto bread roll: ticked spreadable butter, low fat | bread_roll_spread_butter_spread_lowfat_med | 15 | bread_roll_spread_butter_spread_lowfat_med | 12 | -20% | Food portion sizes / MAFF; corrected typo in weight |
| Fat that is spread (thick amount) onto bread roll: ticked spreadable butter, low fat | bread_roll_spread_butter_spread_lowfat_thick | 15 | bread_roll_spread_butter_spread_lowfat_thick | 15 | 0% | Food portion sizes / MAFF |
| Fat that is spread (thin amount) onto bread roll: ticked spreadable butter, low fat | bread_roll_spread_butter_spread_lowfat_thin | 15 | bread_roll_spread_butter_spread_lowfat_thin | 10 | -33% | Food portion sizes / MAFF; corrected typo in weight |
| Fat that is spread (medium amount) onto bread roll: ticked dairy spread which is also cholesterol lowering e.g Benecol Buttery | bread_roll_spread_dairy_chol_med | 10 | bread_roll_spread_dairy_chol_med | 10 | 0% | Food portion sizes / MAFF |
| Fat that is spread (thick amount) onto bread roll: ticked dairy spread which is also cholesterol lowering e.g Benecol Buttery | bread_roll_spread_dairy_chol_thick | 12 | bread_roll_spread_dairy_chol_thick | 12 | 0% | Food portion sizes / MAFF |
| Fat that is spread (thin amount) onto bread roll: ticked dairy spread which is also cholesterol lowering e.g Benecol Buttery | bread_roll_spread_dairy_chol_thin | 7 | bread_roll_spread_dairy_chol_thin | 7 | 0% | Food portion sizes / MAFF |
| Fat that is spread (medium amount) onto bread roll: ticked dairy spread but not specified amount of % fat | bread_roll_spread_dairy_dunno_med | 10 | bread_roll_spread_dairy_dunno_med | 10 | 0% | Food portion sizes / MAFF; food composition as low fat and reduced fat |
| Fat that is spread (thick amount) onto bread roll: ticked dairy spread but not specified amount of % fat | bread_roll_spread_dairy_dunno_thick | 12 | bread_roll_spread_dairy_dunno_thick | 12 | 0% | Food portion sizes / MAFF; food composition as low fat and reduced fat |
| Fat that is spread (thin amount) onto bread roll: ticked dairy spread but not specified amount of % fat | bread_roll_spread_dairy_dunno_thin | 7 | bread_roll_spread_dairy_dunno_thin | 7 | 0% | Food portion sizes / MAFF; food composition as low fat and reduced fat |
| Fat that is spread (medium amount) onto bread roll: ticked dairy spread with normal amount of % fat | bread_roll_spread_dairy_fat_med | 10 | bread_roll_spread_dairy_fat_med | 10 | 0% | Food portion sizes / MAFF; normal fat is taken as reduced fat (up to 62% fat) |
| Fat that is spread (thick amount) onto bread roll: ticked dairy spread with normal amount of % fat | bread_roll_spread_dairy_fat_thick | 12 | bread_roll_spread_dairy_fat_thick | 12 | 0% | Food portion sizes / MAFF; normal fat is taken as reduced fat (up to 62% fat) |
| Fat that is spread (thin amount) onto bread roll: ticked dairy spread with normal amount of % fat | bread_roll_spread_dairy_fat_thin | 7 | bread_roll_spread_dairy_fat_thin | 7 | 0% | Food portion sizes / MAFF; normal fat is taken as reduced fat (up to 62% fat) |
| Fat that is spread (medium amount) onto bread roll: ticked dairy spread, low fat | bread_roll_spread_dairy_lowfat_med | 10 | bread_roll_spread_dairy_lowfat_med | 10 | 0% | Food portion sizes / MAFF |
| Fat that is spread (thick amount) onto bread roll: ticked dairy spread, low fat | bread_roll_spread_dairy_lowfat_thick | 12 | bread_roll_spread_dairy_lowfat_thick | 12 | 0% | Food portion sizes / MAFF |
| Fat that is spread (thin amount) onto bread roll: ticked dairy spread, low fat | bread_roll_spread_dairy_lowfat_thin | 7 | bread_roll_spread_dairy_lowfat_thin | 7 | 0% | Food portion sizes / MAFF |
| Fat that is spread (medium amount) onto bread roll: ticked dairy spread, very low fat | bread_roll_spread_dairy_vlowfat_med | 10 | bread_roll_spread_dairy_vlowfat_med | 10 | 0% | Food portion sizes / MAFF; food composition as low fat spread |
| Fat that is spread (thick amount) onto bread roll: ticked dairy spread, very low fat | bread_roll_spread_dairy_vlowfat_thick | 12 | bread_roll_spread_dairy_vlowfat_thick | 12 | 0% | Food portion sizes / MAFF; food composition as low fat spread |
| Fat that is spread (thin amount) onto bread roll: ticked dairy spread, very low fat | bread_roll_spread_dairy_vlowfat_thin | 7 | bread_roll_spread_dairy_vlowfat_thin | 7 | 0% | Food portion sizes / MAFF; food composition as low fat spread |
| Fat that is spread (medium amount) onto bread roll: not specified type of spread but ticked cholesterol lowering e.g Benecol, Flora pro active | bread_roll_spread_dunno_chol_med | 10 | bread_roll_spread_dunno_chol_med | 10 | 0% | Food portion sizes / MAFF |
| Fat that is spread (thick amount) onto bread roll: not specified type of spread but ticked cholesterol lowering e.g Benecol, Flora pro active | bread_roll_spread_dunno_chol_thick | 12 | bread_roll_spread_dunno_chol_thick | 12 | 0% | Food portion sizes / MAFF |
| Fat that is spread (thin amount) onto bread roll: not specified type of spread but ticked cholesterol lowering e.g Benecol, Flora pro active | bread_roll_spread_dunno_chol_thin | 7 | bread_roll_spread_dunno_chol_thin | 7 | 0% | Food portion sizes / MAFF |
| Fat that is spread (medium amount) onto bread roll: not specified type of spread or amount of % fat | bread_roll_spread_dunno_dunno_med | 10 | bread_roll_spread_dunno_dunno_med | 10 | 0% | Food portion sizes / MAFF |
| Fat that is spread (thick amount) onto bread roll: not specified type of spread or amount of % fat | bread_roll_spread_dunno_dunno_thick | 12 | bread_roll_spread_dunno_dunno_thick | 12 | 0% | Food portion sizes / MAFF |
| Fat that is spread (thin amount) onto bread roll: not specified type of spread or amount of % fat | bread_roll_spread_dunno_dunno_thin | 7 | bread_roll_spread_dunno_dunno_thin | 7 | 0% | Food portion sizes / MAFF |
| Fat that is spread (medium amount) onto bread roll: not specified type of spread but ticked normal amount % fat | bread_roll_spread_dunno_fat_med | 10 | bread_roll_spread_dunno_fat_med | 10 | 0% | Food portion sizes / MAFF; normal fat is taken as reduced fat (up to 62% fat) |
| Fat that is spread (thick amount) onto bread roll: not specified type of spread but ticked normal amount % fat | bread_roll_spread_dunno_fat_thick | 12 | bread_roll_spread_dunno_fat_thick | 12 | 0% | Food portion sizes / MAFF; normal fat is taken as reduced fat (up to 62% fat) |
| Fat that is spread (thin amount) onto bread roll: not specified type of spread but ticked normal amount % fat | bread_roll_spread_dunno_fat_thin | 7 | bread_roll_spread_dunno_fat_thin | 7 | 0% | Food portion sizes / MAFF; normal fat is taken as reduced fat (up to 62% fat) |
| Fat that is spread (medium amount) onto bread roll: not specified type of spread but ticked low fat | bread_roll_spread_dunno_lowfat_med | 10 | bread_roll_spread_dunno_lowfat_med | 10 | 0% | Food portion sizes / MAFF |
| Fat that is spread (thick amount) onto bread roll: not specified type of spread but ticked low fat | bread_roll_spread_dunno_lowfat_thick | 12 | bread_roll_spread_dunno_lowfat_thick | 12 | 0% | Food portion sizes / MAFF |
| Fat that is spread (thin amount) onto bread roll: not specified type of spread but ticked low fat | bread_roll_spread_dunno_lowfat_thin | 7 | bread_roll_spread_dunno_lowfat_thin | 7 | 0% | Food portion sizes / MAFF |
| Fat that is spread (medium amount) onto bread roll: not specified type of spread but ticked very low fat | bread_roll_spread_dunno_vlowfat_med | 10 | bread_roll_spread_dunno_vlowfat_med | 10 | 0% | Food portion sizes / MAFF; food composition as low fat spread |
| Fat that is spread (thick amount) onto bread roll: not specified type of spread but ticked very low fat | bread_roll_spread_dunno_vlowfat_thick | 12 | bread_roll_spread_dunno_vlowfat_thick | 12 | 0% | Food portion sizes / MAFF; food composition as low fat spread |
| Fat that is spread (thin amount) onto bread roll: not specified type of spread but ticked very low fat | bread_roll_spread_dunno_vlowfat_thin | 7 | bread_roll_spread_dunno_vlowfat_thin | 7 | 0% | Food portion sizes / MAFF; food composition as low fat spread |
| Fat that is spread (medium amount) onto bread roll: ticked hard margarine (hard block margarine in wrapper) | bread_roll_spread_hardmarg_med | 12 | bread_roll_spread_hardmarg_med | 12 | 0% | Food portion sizes / MAFF; spread weight as butter |
| Fat that is spread (thick amount) onto bread roll: ticked hard margarine (hard block margarine in wrapper) | bread_roll_spread_hardmarg_thick | 15 | bread_roll_spread_hardmarg_thick | 15 | 0% | Food portion sizes / MAFF; spread weight as butter |
| Fat that is spread (thin amount) onto bread roll: ticked hard margarine (hard block margarine in wrapper) | bread_roll_spread_hardmarg_thin | 10 | bread_roll_spread_hardmarg_thin | 10 | 0% | Food portion sizes / MAFF; spread weight as butter |
| Fat that is spread (medium amount) onto bread roll: ticked cholesterol lowering olive spread e.g Benecol/Flora pro active olive spread | bread_roll_spread_olive_chol_med | 10 | bread_roll_spread_olive_chol_med | 10 | 0% | Food portion sizes / MAFF |
| Fat that is spread (thick amount) onto bread roll: ticked cholesterol lowering olive spread e.g Benecol/Flora pro active olive spread | bread_roll_spread_olive_chol_thick | 12 | bread_roll_spread_olive_chol_thick | 12 | 0% | Food portion sizes / MAFF |
| Fat that is spread (thin amount) onto bread roll: ticked cholesterol lowering olive spread e.g Benecol/Flora pro active olive spread | bread_roll_spread_olive_chol_thin | 7 | bread_roll_spread_olive_chol_thin | 7 | 0% | Food portion sizes / MAFF |
| Fat that is spread (medium amount) onto bread roll: ticked olive spread but not specified amount of fat | bread_roll_spread_olive_dunno_med | 10 | bread_roll_spread_olive_dunno_med | 10 | 0% | Food portion sizes / MAFF |
| Fat that is spread (thick amount) onto bread roll: ticked olive spread but not specified amount of fat | bread_roll_spread_olive_dunno_thick | 12 | bread_roll_spread_olive_dunno_thick | 12 | 0% | Food portion sizes / MAFF |
| Fat that is spread (thin amount) onto bread roll: ticked olive spread but not specified amount of fat | bread_roll_spread_olive_dunno_thin | 7 | bread_roll_spread_olive_dunno_thin | 7 | 0% | Food portion sizes / MAFF |
| Fat that is spread (medium amount) onto bread roll: ticked olive spread with normal amount of % fat | bread_roll_spread_olive_fat_med | 10 | bread_roll_spread_olive_fat_med | 10 | 0% | Food portion sizes / MAFF; normal fat is taken as reduced fat (up to 62% fat) |
| Fat that is spread (thick amount) onto bread roll: ticked olive spread with normal amount of % fat | bread_roll_spread_olive_fat_thick | 12 | bread_roll_spread_olive_fat_thick | 12 | 0% | Food portion sizes / MAFF; normal fat is taken as reduced fat (up to 62% fat) |
| Fat that is spread (thin amount) onto bread roll: ticked olive spread with normal amount of % fat | bread_roll_spread_olive_fat_thin | 7 | bread_roll_spread_olive_fat_thin | 7 | 0% | Food portion sizes / MAFF; normal fat is taken as reduced fat (up to 62% fat) |
| Fat that is spread (medium amount) onto bread roll: ticked olive spread, low fat | bread_roll_spread_olive_lowfat_med | 10 | bread_roll_spread_olive_lowfat_med | 10 | 0% | Food portion sizes / MAFF |
| Fat that is spread (thick amount) onto bread roll: ticked olive spread, low fat | bread_roll_spread_olive_lowfat_thick | 12 | bread_roll_spread_olive_lowfat_thick | 12 | 0% | Food portion sizes / MAFF |
| Fat that is spread (thin amount) onto bread roll: ticked olive spread, low fat | bread_roll_spread_olive_lowfat_thin | 7 | bread_roll_spread_olive_lowfat_thin | 7 | 0% | Food portion sizes / MAFF |
| Fat that is spread (medium amount) onto bread roll: ticked olive spread, very low fat | bread_roll_spread_olive_vlowfat_med | 10 | bread_roll_spread_olive_vlowfat_med | 10 | 0% | Food portion sizes / MAFF; food composition as low fat spread |
| Fat that is spread (thick amount) onto bread roll: ticked olive spread, very low fat | bread_roll_spread_olive_vlowfat_thick | 12 | bread_roll_spread_olive_vlowfat_thick | 12 | 0% | Food portion sizes / MAFF; food composition as low fat spread |
| Fat that is spread (thin amount) onto bread roll: ticked olive spread, very low fat | bread_roll_spread_olive_vlowfat_thin | 7 | bread_roll_spread_olive_vlowfat_thin | 7 | 0% | Food portion sizes / MAFF; food composition as low fat spread |
| Fat that is spread (medium amount) onto bread roll: ticked other spread e.g ghee | bread_roll_spread_other_med | 10 | bread_roll_spread_other_med | 10 | 0% | Food portion sizes / MAFF |
| Fat that is spread (thick amount) onto bread roll: ticked other spread e.g ghee | bread_roll_spread_other_thick | 12 | bread_roll_spread_other_thick | 12 | 0% | Food portion sizes / MAFF |
| Fat that is spread (thin amount) onto bread roll: ticked other spread e.g ghee | bread_roll_spread_other_thin | 7 | bread_roll_spread_other_thin | 7 | 0% | Food portion sizes / MAFF |
| Fat that is spread (medium amount) onto bread roll: ticked polyunsaturated margarine (e.g Flora) and also cholesterol lowering | bread_roll_spread_polymarg_chol_med | 10 | bread_roll_spread_polymarg_chol_med | 10 | 0% | Food portion sizes / MAFF |
| Fat that is spread (thick amount) onto bread roll: ticked polyunsaturated margarine (e.g Flora) and also cholesterol lowering | bread_roll_spread_polymarg_chol_thick | 12 | bread_roll_spread_polymarg_chol_thick | 12 | 0% | Food portion sizes / MAFF |
| Fat that is spread (thin amount) onto bread roll: ticked polyunsaturated margarine (e.g Flora) and also cholesterol lowering | bread_roll_spread_polymarg_chol_thin | 7 | bread_roll_spread_polymarg_chol_thin | 7 | 0% | Food portion sizes / MAFF |
| Fat that is spread (medium amount) onto bread roll: ticked polyunsaturated margarine (e.g Flora) but not specified amount of fat | bread_roll_spread_polymarg_dunno_med | 10 | bread_roll_spread_polymarg_dunno_med | 10 | 0% | Food portion sizes / MAFF |
| Fat that is spread (thick amount) onto bread roll: ticked polyunsaturated margarine (e.g Flora) but not specified amount of fat | bread_roll_spread_polymarg_dunno_thick | 12 | bread_roll_spread_polymarg_dunno_thick | 12 | 0% | Food portion sizes / MAFF |
| Fat that is spread (thin amount) onto bread roll: ticked polyunsaturated margarine (e.g Flora) but not specified amount of fat | bread_roll_spread_polymarg_dunno_thin | 7 | bread_roll_spread_polymarg_dunno_thin | 7 | 0% | Food portion sizes / MAFF |
| Fat that is spread (medium amount) onto bread roll: ticked polyunsaturated margarine (e.g Flora), normal amount of % fat | bread_roll_spread_polymarg_fat_med | 10 | bread_roll_spread_polymarg_fat_med | 10 | 0% | Food portion sizes / MAFF; normal fat is taken as reduced fat (up to 62% fat) |
| Fat that is spread (thick amount) onto bread roll: ticked polyunsaturated margarine (e.g Flora), normal amount of % fat | bread_roll_spread_polymarg_fat_thick | 12 | bread_roll_spread_polymarg_fat_thick | 12 | 0% | Food portion sizes / MAFF; normal fat is taken as reduced fat (up to 62% fat) |
| Fat that is spread (thin amount) onto bread roll: ticked polyunsaturated margarine (e.g Flora), normal amount of % fat | bread_roll_spread_polymarg_fat_thin | 7 | bread_roll_spread_polymarg_fat_thin | 7 | 0% | Food portion sizes / MAFF; normal fat is taken as reduced fat (up to 62% fat) |
| Fat that is spread (medium amount) onto bread roll: ticked polyunsaturated margarine (e.g Flora), low fat | bread_roll_spread_polymarg_lowfat_med | 10 | bread_roll_spread_polymarg_lowfat_med | 10 | 0% | Food portion sizes / MAFF |
| Fat that is spread (thick amount) onto bread roll: ticked polyunsaturated margarine (e.g Flora), low fat | bread_roll_spread_polymarg_lowfat_thick | 12 | bread_roll_spread_polymarg_lowfat_thick | 12 | 0% | Food portion sizes / MAFF |
| Fat that is spread (thin amount) onto bread roll: ticked polyunsaturated margarine (e.g Flora), low fat | bread_roll_spread_polymarg_lowfat_thin | 7 | bread_roll_spread_polymarg_lowfat_thin | 7 | 0% | Food portion sizes / MAFF |
| Fat that is spread (medium amount) onto bread roll: ticked polyunsaturated margarine (e.g Flora), very low fat | bread_roll_spread_polymarg_vlowfat_med | 10 | bread_roll_spread_polymarg_vlowfat_med | 10 | 0% | Food portion sizes / MAFF; food composition as low fat spread |
| Fat that is spread (thick amount) onto bread roll: ticked polyunsaturated margarine (e.g Flora), very low fat | bread_roll_spread_polymarg_vlowfat_thick | 12 | bread_roll_spread_polymarg_vlowfat_thick | 12 | 0% | Food portion sizes / MAFF; food composition as low fat spread |
| Fat that is spread (thin amount) onto bread roll: ticked polyunsaturated margarine (e.g Flora), very low fat | bread_roll_spread_polymarg_vlowfat_thin | 7 | bread_roll_spread_polymarg_vlowfat_thin | 7 | 0% | Food portion sizes / MAFF; food composition as low fat spread |
| Fat that is spread (medium amount) onto bread roll: ticked soya/vegan/dairy free margarine e.g Pure, and also cholesterol lowering | bread_roll_spread_soya_chol_med | 10 | bread_roll_spread_soya_chol_med | 10 | 0% | Food portion sizes / MAFF |
| Fat that is spread (thick amount) onto bread roll: ticked soya/vegan/dairy free margarine e.g Pure, and also cholesterol lowering | bread_roll_spread_soya_chol_thick | 12 | bread_roll_spread_soya_chol_thick | 12 | 0% | Food portion sizes / MAFF |
| Fat that is spread (thin amount) onto bread roll: ticked soya/vegan/dairy free margarine e.g Pure, and also cholesterol lowering | bread_roll_spread_soya_chol_thin | 7 | bread_roll_spread_soya_chol_thin | 7 | 0% | Food portion sizes / MAFF |
| Fat that is spread (medium amount) onto bread roll: ticked soya/vegan/dairy free margarine e.g Pure, and not specified amount of fat | bread_roll_spread_soya_dunno_med | 10 | bread_roll_spread_soya_dunno_med | 10 | 0% | Food portion sizes / MAFF |
| Fat that is spread (thick amount) onto bread roll: ticked soya/vegan/dairy free margarine e.g Pure, and not specified amount of fat | bread_roll_spread_soya_dunno_thick | 12 | bread_roll_spread_soya_dunno_thick | 12 | 0% | Food portion sizes / MAFF |
| Fat that is spread (thin amount) onto bread roll: ticked soya/vegan/dairy free margarine e.g Pure, and not specified amount of fat | bread_roll_spread_soya_dunno_thin | 7 | bread_roll_spread_soya_dunno_thin | 7 | 0% | Food portion sizes / MAFF |
| Fat that is spread (medium amount) onto bread roll: ticked soya/vegan/dairy free margarine (e.g Pure), normal amount % fat | bread_roll_spread_soya_fat_med | 10 | bread_roll_spread_soya_fat_med | 10 | 0% | Food portion sizes / MAFF; normal fat is taken as reduced fat (up to 62% fat) |
| Fat that is spread (thick amount) onto bread roll: ticked soya/vegan/dairy free margarine (e.g Pure), normal amount % fat | bread_roll_spread_soya_fat_thick | 12 | bread_roll_spread_soya_fat_thick | 12 | 0% | Food portion sizes / MAFF; normal fat is taken as reduced fat (up to 62% fat) |
| Fat that is spread (thin amount) onto bread roll: ticked soya/vegan/dairy free margarine (e.g Pure), normal amount % fat | bread_roll_spread_soya_fat_thin | 7 | bread_roll_spread_soya_fat_thin | 7 | 0% | Food portion sizes / MAFF; normal fat is taken as reduced fat (up to 62% fat) |
| Fat that is spread (medium amount) onto bread roll: ticked soya/vegan/dairy free margarine (e.g Pure), low fat | bread_roll_spread_soya_lowfat_med | 10 | bread_roll_spread_soya_lowfat_med | 10 | 0% | Food portion sizes / MAFF |
| Fat that is spread (thick amount) onto bread roll: ticked soya/vegan/dairy free margarine (e.g Pure), low fat | bread_roll_spread_soya_lowfat_thick | 12 | bread_roll_spread_soya_lowfat_thick | 12 | 0% | Food portion sizes / MAFF |
| Fat that is spread (thin amount) onto bread roll: ticked soya/vegan/dairy free margarine (e.g Pure), low fat | bread_roll_spread_soya_lowfat_thin | 7 | bread_roll_spread_soya_lowfat_thin | 7 | 0% | Food portion sizes / MAFF |
| Fat that is spread (medium amount) onto bread roll: ticked soya/vegan/dairy free margarine (e.g Pure), very low fat | bread_roll_spread_soya_vlowfat_med | 10 | bread_roll_spread_soya_vlowfat_med | 10 | 0% | Food portion sizes / MAFF; food composition as low fat spread |
| Fat that is spread (thick amount) onto bread roll: ticked soya/vegan/dairy free margarine (e.g Pure), very low fat | bread_roll_spread_soya_vlowfat_thick | 12 | bread_roll_spread_soya_vlowfat_thick | 12 | 0% | Food portion sizes / MAFF; food composition as low fat spread |
| Fat that is spread (thin amount) onto bread roll: ticked soya/vegan/dairy free margarine (e.g Pure), very low fat | bread_roll_spread_soya_vlowfat_thin | 7 | bread_roll_spread_soya_vlowfat_thin | 7 | 0% | Food portion sizes / MAFF; food composition as low fat spread |
| Bread roll, bap, bun, bagel: not specified type so includes white, brown, granary, wholemeal, wheatgerm | bread_roll_unanswered | 60 | bread_roll_unanswered | 60 | 0% | |
| Bread roll, bap, bun, bagel: white | bread_roll_white | 48 | bread_roll_white | 60 | 25% | Portion size is now consistent with other bread rolls |
| Bread roll, bap, bun, bagel: wholemeal | bread_roll_wholemeal | 60 | bread_roll_wholemeal | 60 | 0% | |
| Gluten free sliced bread: brown, white with added fibre | | | bread_sliced_gf_nonwhite | 36 | - | Gluten free version added to the updated version |
| Gluten free sliced bread: not specified type so includes white, brown, white with added fibre | | | bread_sliced_gf_unanswered | 36 | - | Gluten free version added to the updated version |
| Gluten free sliced bread: white | | | bread_sliced_gf_white | 36 | - | Gluten free version added to the updated version |
| Sliced bread: brown, granary, high fibre white, 50/50 | bread_sliced_mixed | 36 | bread_sliced_mixed | 36 | 0% | |
| Sliced bread: brown, wheatgerm, rye, white | bread_sliced_other | 36 | bread_sliced_other | 36 | 0% | |
| Seeds for seeded bread: e.g sesame, sunflower, poppy | bread_sliced_seeded | 3 | bread_sliced_seeded | 3 | 0% | |
| Fat that is spread (medium amount) onto sliced bread: ticked butter but not specified amount of % fat | bread_sliced_spread_butter_dunno_med | 10 | bread_sliced_spread_butter_dunno_med | 10 | 0% | Food portion sizes / MAFF; food composition as butter and spreadable butter |
| Fat that is spread (thick amount) onto sliced bread: ticked butter but not specified amount of % fat | bread_sliced_spread_butter_dunno_thick | 12 | bread_sliced_spread_butter_dunno_thick | 12 | 0% | Food portion sizes / MAFF; food composition as butter and spreadable butter |
| Fat that is spread (thin amount) onto sliced bread: ticked butter but not specified amount of % fat | bread_sliced_spread_butter_dunno_thin | 7 | bread_sliced_spread_butter_dunno_thin | 7 | 0% | Food portion sizes / MAFF; food composition as butter and spreadable butter |
| Fat that is spread (medium amount) onto sliced bread: ticked butter, normal amount of % fat | bread_sliced_spread_butter_fat_med | 10 | bread_sliced_spread_butter_fat_med | 10 | 0% | Food portion sizes / MAFF |
| Fat that is spread (thick amount) onto sliced bread: ticked butter, normal amount of % fat | bread_sliced_spread_butter_fat_thick | 12 | bread_sliced_spread_butter_fat_thick | 12 | 0% | Food portion sizes / MAFF |

| | McCance and Widdowson’s | | Nutrient databank + other changes | | | |
|---|---|---|---|---|---|---|
| **Variable description** | **Food item** | **Portion size** | **Food item** | **Portion size** | **Portion diff%** | **Description of differences with previous version** |
| Fat that is spread (thin amount) onto sliced bread: ticked butter, normal amount of % fat | bread_sliced_spread_butter_fat_thin | 7 | bread_sliced_spread_butter_fat_thin | 7 | 0% | Food portion sizes / MAFF |
| Fat that is spread (medium amount) onto sliced bread: ticked butter, low fat | bread_sliced_spread_butter_lowfat_med | 10 | bread_sliced_spread_butter_lowfat_med | 10 | 0% | Food portion sizes / MAFF |
| Fat that is spread (thick amount) onto sliced bread: ticked butter, low fat | bread_sliced_spread_butter_lowfat_thick | 12 | bread_sliced_spread_butter_lowfat_thick | 12 | 0% | Food portion sizes / MAFF |
| Fat that is spread (thin amount) onto sliced bread: ticked butter, low fat | bread_sliced_spread_butter_lowfat_thin | 7 | bread_sliced_spread_butter_lowfat_thin | 7 | 0% | Food portion sizes / MAFF |
| Fat that is spread (medium amount) onto sliced bread: ticked spreadable butter with normal amount of % fat | bread_sliced_spread_butter_spread_fat_med | 10 | bread_sliced_spread_butter_spread_fat_med | 10 | 0% | Food portion sizes / MAFF |
| Fat that is spread (thick amount) onto sliced bread: ticked spreadable butter with normal amount of % fat | bread_sliced_spread_butter_spread_fat_thick | 12 | bread_sliced_spread_butter_spread_fat_thick | 12 | 0% | Food portion sizes / MAFF |
| Fat that is spread (thin amount) onto sliced bread: ticked spreadable butter with normal amount of % fat | bread_sliced_spread_butter_spread_fat_thin | 7 | bread_sliced_spread_butter_spread_fat_thin | 7 | 0% | Food portion sizes / MAFF |
| Fat that is spread (medium amount) onto sliced bread: ticked spreadable butter, low fat | bread_sliced_spread_butter_spread_lowfat_med | 10 | bread_sliced_spread_butter_spread_lowfat_med | 10 | 0% | Food portion sizes / MAFF |
| Fat that is spread (thick amount) onto sliced bread: ticked spreadable butter, low fat | bread_sliced_spread_butter_spread_lowfat_thick | 12 | bread_sliced_spread_butter_spread_lowfat_thick | 12 | 0% | Food portion sizes / MAFF |
| Fat that is spread (thin amount) onto sliced bread: ticked spreadable butter, low fat | bread_sliced_spread_butter_spread_lowfat_thin | 7 | bread_sliced_spread_butter_spread_lowfat_thin | 7 | 0% | Food portion sizes / MAFF |
| Fat that is spread (medium amount) onto sliced bread: ticked dairy spread which is also cholesterol lowering e.g Benecol Buttery | bread_sliced_spread_dairy_chol_med | 10 | bread_sliced_spread_dairy_chol_med | 7 | -30% | Food portion sizes / MAFF |
| Fat that is spread (thick amount) onto sliced bread: ticked dairy spread which is also cholesterol lowering e.g Benecol Buttery | bread_sliced_spread_dairy_chol_thick | 12 | bread_sliced_spread_dairy_chol_thick | 10 | -17% | Food portion sizes / MAFF |
| Fat that is spread (thin amount) onto sliced bread: ticked dairy spread which is also cholesterol lowering e.g Benecol Buttery | bread_sliced_spread_dairy_chol_thin | 7 | bread_sliced_spread_dairy_chol_thin | 5 | -29% | Food portion sizes / MAFF |
| Fat that is spread (medium amount) onto sliced bread: ticked dairy spread but not specified amount of % fat | bread_sliced_spread_dairy_dunno_med | 10 | bread_sliced_spread_dairy_dunno_med | 7 | -30% | Food portion sizes / MAFF; food composition as low fat and reduced fat |
| Fat that is spread (thick amount) onto sliced bread: ticked dairy spread but not specified amount of % fat | bread_sliced_spread_dairy_dunno_thick | 12 | bread_sliced_spread_dairy_dunno_thick | 10 | -17% | Food portion sizes / MAFF; food composition as low fat and reduced fat |
| Fat that is spread (thin amount) onto sliced bread: ticked dairy spread but not specified amount of % fat | bread_sliced_spread_dairy_dunno_thin | 7 | bread_sliced_spread_dairy_dunno_thin | 5 | -29% | Food portion sizes / MAFF; food composition as low fat and reduced fat |
| Fat that is spread (medium amount) onto sliced bread: ticked dairy spread with normal amount of % fat | bread_sliced_spread_dairy_fat_med | 10 | bread_sliced_spread_dairy_fat_med | 7 | -30% | Food portion sizes / MAFF; normal fat is taken as reduced fat (up to 62% fat) |
| Fat that is spread (thick amount) onto sliced bread: ticked dairy spread with normal amount of % fat | bread_sliced_spread_dairy_fat_thick | 12 | bread_sliced_spread_dairy_fat_thick | 10 | -17% | Food portion sizes / MAFF; normal fat is taken as reduced fat (up to 62% fat) |
| Fat that is spread (thin amount) onto sliced bread: ticked dairy spread with normal amount of % fat | bread_sliced_spread_dairy_fat_thin | 7 | bread_sliced_spread_dairy_fat_thin | 5 | -29% | Food portion sizes / MAFF; normal fat is taken as reduced fat (up to 62% fat) |
| Fat that is spread (medium amount) onto sliced bread: ticked dairy spread, low fat | bread_sliced_spread_dairy_lowfat_med | 10 | bread_sliced_spread_dairy_lowfat_med | 7 | -30% | Food portion sizes / MAFF |
| Fat that is spread (thick amount) onto sliced bread: ticked dairy spread, low fat | bread_sliced_spread_dairy_lowfat_thick | 12 | bread_sliced_spread_dairy_lowfat_thick | 10 | -17% | Food portion sizes / MAFF |
| Fat that is spread (thin amount) onto sliced bread: ticked dairy spread, low fat | bread_sliced_spread_dairy_lowfat_thin | 7 | bread_sliced_spread_dairy_lowfat_thin | 5 | -29% | Food portion sizes / MAFF |
| Fat that is spread (medium amount) onto sliced bread: ticked dairy spread, very low fat | bread_sliced_spread_dairy_vlowfat_med | 10 | bread_sliced_spread_dairy_vlowfat_med | 7 | -30% | Food portion sizes / MAFF; food composition as low fat spread |
| Fat that is spread (thick amount) onto sliced bread: ticked dairy spread, very low fat | bread_sliced_spread_dairy_vlowfat_thick | 12 | bread_sliced_spread_dairy_vlowfat_thick | 10 | -17% | Food portion sizes / MAFF; food composition as low fat spread |
| Fat that is spread (thin amount) onto sliced bread: ticked dairy spread, very low fat | bread_sliced_spread_dairy_vlowfat_thin | 7 | bread_sliced_spread_dairy_vlowfat_thin | 5 | -29% | Food portion sizes / MAFF; food composition as low fat spread |
| Fat that is spread (medium amount) onto sliced bread: not specified type of spread but ticked cholesterol lowering e.g Benecol, Flora pro active | bread_sliced_spread_dunno_chol_med | 10 | bread_sliced_spread_dunno_chol_med | 7 | -30% | Food portion sizes / MAFF |
| Fat that is spread (thick amount) onto sliced bread: not specified type of spread but ticked cholesterol lowering e.g Benecol, Flora pro active | bread_sliced_spread_dunno_chol_thick | 12 | bread_sliced_spread_dunno_chol_thick | 10 | -17% | Food portion sizes / MAFF |
| Fat that is spread (thin amount) onto sliced bread: not specified type of spread but ticked cholesterol lowering e.g Benecol, Flora pro active | bread_sliced_spread_dunno_chol_thin | 7 | bread_sliced_spread_dunno_chol_thin | 5 | -29% | Food portion sizes / MAFF |
| Fat that is spread (medium amount) onto sliced bread: not specified type of spread or amount of % fat | bread_sliced_spread_dunno_dunno_med | 10 | bread_sliced_spread_dunno_dunno_med | 7 | -30% | Food portion sizes / MAFF |
| Fat that is spread (thick amount) onto sliced bread: not specified type of spread or amount of % fat | bread_sliced_spread_dunno_dunno_thick | 12 | bread_sliced_spread_dunno_dunno_thick | 10 | -17% | Food portion sizes / MAFF |
| Fat that is spread (thin amount) onto sliced bread: not specified type of spread or amount of % fat | bread_sliced_spread_dunno_dunno_thin | 7 | bread_sliced_spread_dunno_dunno_thin | 5 | -29% | Food portion sizes / MAFF |
| Fat that is spread (medium amount) onto sliced bread: not specified type of spread but ticked normal amount % fat | bread_sliced_spread_dunno_fat_med | 10 | bread_sliced_spread_dunno_fat_med | 7 | -30% | Food portion sizes / MAFF; normal fat is taken as reduced fat (up to 62% fat) |
| Fat that is spread (thick amount) onto sliced bread: not specified type of spread but ticked normal amount % fat | bread_sliced_spread_dunno_fat_thick | 12 | bread_sliced_spread_dunno_fat_thick | 10 | -17% | Food portion sizes / MAFF; normal fat is taken as reduced fat (up to 62% fat) |
| Fat that is spread (thin amount) onto sliced bread: not specified type of spread but ticked normal amount % fat | bread_sliced_spread_dunno_fat_thin | 7 | bread_sliced_spread_dunno_fat_thin | 5 | -29% | Food portion sizes / MAFF; normal fat is taken as reduced fat (up to 62% fat) |
| Fat that is spread (medium amount) onto sliced bread: not specified type of spread but ticked low fat | bread_sliced_spread_dunno_lowfat_med | 10 | bread_sliced_spread_dunno_lowfat_med | 7 | -30% | Food portion sizes / MAFF |
| Fat that is spread (thick amount) onto sliced bread: not specified type of spread but ticked low fat | bread_sliced_spread_dunno_lowfat_thick | 12 | bread_sliced_spread_dunno_lowfat_thick | 10 | -17% | Food portion sizes / MAFF |
| Fat that is spread (thin amount) onto sliced bread: not specified type of spread but ticked low fat | bread_sliced_spread_dunno_lowfat_thin | 7 | bread_sliced_spread_dunno_lowfat_thin | 5 | -29% | Food portion sizes / MAFF |
| Fat that is spread (medium amount) onto sliced bread: not specified type of spread but ticked very low fat | bread_sliced_spread_dunno_vlowfat_med | 10 | bread_sliced_spread_dunno_vlowfat_med | 7 | -30% | Food portion sizes / MAFF; food composition as low fat spread |
| Fat that is spread (thick amount) onto sliced bread: not specified type of spread but ticked very low fat | bread_sliced_spread_dunno_vlowfat_thick | 12 | bread_sliced_spread_dunno_vlowfat_thick | 10 | -17% | Food portion sizes / MAFF; food composition as low fat spread |
| Fat that is spread (thin amount) onto sliced bread: not specified type of spread but ticked very low fat | bread_sliced_spread_dunno_vlowfat_thin | 7 | bread_sliced_spread_dunno_vlowfat_thin | 5 | -29% | Food portion sizes / MAFF; food composition as low fat spread |
| Fat that is spread (medium amount) onto sliced bread: ticked hard margarine (hard block margarine in wrapper) | bread_sliced_spread_hardmarg_med | 7 | bread_sliced_spread_hardmarg_med | 10 | 43% | Food portion sizes / MAFF as butter |
| Fat that is spread (thick amount) onto sliced bread: ticked hard margarine (hard block margarine in wrapper) | bread_sliced_spread_hardmarg_thick | 10 | bread_sliced_spread_hardmarg_thick | 12 | 20% | Food portion sizes / MAFF as butter |
| Fat that is spread (thin amount) onto sliced bread: ticked hard margarine (hard block margarine in wrapper) | bread_sliced_spread_hardmarg_thin | 5 | bread_sliced_spread_hardmarg_thin | 7 | 40% | Food portion sizes / MAFF as butter |
| Fat that is spread (medium amount) onto sliced bread: ticked cholesterol lowering olive spread e.g Benecol/Flora pro active olive spread | bread_sliced_spread_olive_chol_med | 10 | bread_sliced_spread_olive_chol_med | 7 | -30% | Food portion sizes / MAFF |
| Fat that is spread (thick amount) onto sliced bread: ticked cholesterol lowering olive spread e.g Benecol/Flora pro active olive spread | bread_sliced_spread_olive_chol_thick | 12 | bread_sliced_spread_olive_chol_thick | 10 | -17% | Food portion sizes / MAFF |
| Fat that is spread (thin amount) onto sliced bread: ticked cholesterol lowering olive spread e.g Benecol/Flora pro active olive spread | bread_sliced_spread_olive_chol_thin | 7 | bread_sliced_spread_olive_chol_thin | 5 | -29% | Food portion sizes / MAFF |
| Fat that is spread (medium amount) onto sliced bread: ticked olive spread but not specified amount of fat | bread_sliced_spread_olive_dunno_med | 10 | bread_sliced_spread_olive_dunno_med | 7 | -30% | Food portion sizes / MAFF |
| Fat that is spread (thick amount) onto sliced bread: ticked olive spread but not specified amount of fat | bread_sliced_spread_olive_dunno_thick | 12 | bread_sliced_spread_olive_dunno_thick | 10 | -17% | Food portion sizes / MAFF |
| Fat that is spread (thin amount) onto sliced bread: ticked olive spread but not specified amount of fat | bread_sliced_spread_olive_dunno_thin | 7 | bread_sliced_spread_olive_dunno_thin | 5 | -29% | Food portion sizes / MAFF |
| Fat that is spread (medium amount) onto sliced bread: ticked olive spread with normal amount of % fat | bread_sliced_spread_olive_fat_med | 10 | bread_sliced_spread_olive_fat_med | 7 | -30% | Food portion sizes / MAFF; normal fat is taken as reduced fat (up to 62% fat) |
| Fat that is spread (thick amount) onto sliced bread: ticked olive spread with normal amount of % fat | bread_sliced_spread_olive_fat_thick | 12 | bread_sliced_spread_olive_fat_thick | 10 | -17% | Food portion sizes / MAFF; normal fat is taken as reduced fat (up to 62% fat) |
| Fat that is spread (thin amount) onto sliced bread: ticked olive spread with normal amount of % fat | bread_sliced_spread_olive_fat_thin | 7 | bread_sliced_spread_olive_fat_thin | 5 | -29% | Food portion sizes / MAFF; normal fat is taken as reduced fat (up to 62% fat) |
| Fat that is spread (medium amount) onto sliced bread: ticked olive spread, low fat | bread_sliced_spread_olive_lowfat_med | 10 | bread_sliced_spread_olive_lowfat_med | 7 | -30% | Food portion sizes / MAFF |
| Fat that is spread (thick amount) onto sliced bread: ticked olive spread, low fat | bread_sliced_spread_olive_lowfat_thick | 12 | bread_sliced_spread_olive_lowfat_thick | 10 | -17% | Food portion sizes / MAFF |
| Fat that is spread (thin amount) onto sliced bread: ticked olive spread, low fat | bread_sliced_spread_olive_lowfat_thin | 7 | bread_sliced_spread_olive_lowfat_thin | 5 | -29% | Food portion sizes / MAFF |
| Fat that is spread (medium amount) onto sliced bread: ticked olive spread, very low fat | bread_sliced_spread_olive_vlowfat_med | 10 | bread_sliced_spread_olive_vlowfat_med | 7 | -30% | Food portion sizes / MAFF; food composition as low fat spread |
| Fat that is spread (thick amount) onto sliced bread: ticked olive spread, very low fat | bread_sliced_spread_olive_vlowfat_thick | 12 | bread_sliced_spread_olive_vlowfat_thick | 10 | -17% | Food portion sizes / MAFF; food composition as low fat spread |
| Fat that is spread (thin amount) onto sliced bread: ticked olive spread, very low fat | bread_sliced_spread_olive_vlowfat_thin | 7 | bread_sliced_spread_olive_vlowfat_thin | 5 | -29% | Food portion sizes / MAFF; food composition as low fat spread |
| Fat that is spread (medium amount) onto sliced bread: ticked other spread e.g ghee | bread_sliced_spread_other_med | 7 | bread_sliced_spread_other_med | 7 | 0% | Food portion sizes / MAFF |
| Fat that is spread (thick amount) onto sliced bread: ticked other spread e.g ghee | bread_sliced_spread_other_thick | 10 | bread_sliced_spread_other_thick | 10 | 0% | Food portion sizes / MAFF |
| Fat that is spread (thin amount) onto sliced bread: ticked other spread e.g ghee | bread_sliced_spread_other_thin | 5 | bread_sliced_spread_other_thin | 5 | 0% | Food portion sizes / MAFF |
| Fat that is spread (medium amount) onto sliced bread: ticked polyunsaturated margarine (e.g Flora) and also cholesterol lowering | bread_sliced_spread_polymarg_chol_med | 10 | bread_sliced_spread_polymarg_chol_med | 7 | -30% | Food portion sizes / MAFF |
| Fat that is spread (thick amount) onto sliced bread: ticked polyunsaturated margarine (e.g Flora) and also cholesterol lowering | bread_sliced_spread_polymarg_chol_thick | 12 | bread_sliced_spread_polymarg_chol_thick | 10 | -17% | Food portion sizes / MAFF |
| Fat that is spread (thin amount) onto sliced bread: ticked polyunsaturated margarine (e.g Flora) and also cholesterol lowering | bread_sliced_spread_polymarg_chol_thin | 7 | bread_sliced_spread_polymarg_chol_thin | 5 | -29% | Food portion sizes / MAFF |
| Fat that is spread (medium amount) onto sliced bread: ticked polyunsaturated margarine (e.g Flora) but not specified amount of fat | bread_sliced_spread_polymarg_dunno_med | 10 | bread_sliced_spread_polymarg_dunno_med | 7 | -30% | Food portion sizes / MAFF |
| Fat that is spread (thick amount) onto sliced bread: ticked polyunsaturated margarine (e.g Flora) but not specified amount of fat | bread_sliced_spread_polymarg_dunno_thick | 12 | bread_sliced_spread_polymarg_dunno_thick | 10 | -17% | Food portion sizes / MAFF |
| Fat that is spread (thin amount) onto sliced bread: ticked polyunsaturated margarine (e.g Flora) but not specified amount of fat | bread_sliced_spread_polymarg_dunno_thin | 7 | bread_sliced_spread_polymarg_dunno_thin | 5 | -29% | Food portion sizes / MAFF |
| Fat that is spread (medium amount) onto sliced bread: ticked polyunsaturated margarine (e.g Flora), normal amount of % fat | bread_sliced_spread_polymarg_fat_med | 10 | bread_sliced_spread_polymarg_fat_med | 7 | -30% | Food portion sizes / MAFF; normal fat is taken as reduced fat (up to 62% fat) |
| Fat that is spread (thick amount) onto sliced bread: ticked polyunsaturated margarine (e.g Flora), normal amount of % fat | bread_sliced_spread_polymarg_fat_thick | 12 | bread_sliced_spread_polymarg_fat_thick | 10 | -17% | Food portion sizes / MAFF; normal fat is taken as reduced fat (up to 62% fat) |
| Fat that is spread (thin amount) onto sliced bread: ticked polyunsaturated margarine (e.g Flora), normal amount of % fat | bread_sliced_spread_polymarg_fat_thin | 7 | bread_sliced_spread_polymarg_fat_thin | 5 | -29% | Food portion sizes / MAFF; normal fat is taken as reduced fat (up to 62% fat) |
| Fat that is spread (medium amount) onto sliced bread: ticked polyunsaturated margarine (e.g Flora), low fat | bread_sliced_spread_polymarg_lowfat_med | 10 | bread_sliced_spread_polymarg_lowfat_med | 7 | -30% | Food portion sizes / MAFF |
| Fat that is spread (thick amount) onto sliced bread: ticked polyunsaturated margarine (e.g Flora), low fat | bread_sliced_spread_polymarg_lowfat_thick | 12 | bread_sliced_spread_polymarg_lowfat_thick | 10 | -17% | Food portion sizes / MAFF |
| Fat that is spread (thin amount) onto sliced bread: ticked polyunsaturated margarine (e.g Flora), low fat | bread_sliced_spread_polymarg_lowfat_thin | 7 | bread_sliced_spread_polymarg_lowfat_thin | 5 | -29% | Food portion sizes / MAFF |
| Fat that is spread (medium amount) onto sliced bread: ticked polyunsaturated margarine (e.g Flora), very low fat | bread_sliced_spread_polymarg_vlowfat_med | 10 | bread_sliced_spread_polymarg_vlowfat_med | 7 | -30% | Food portion sizes / MAFF; food composition as low fat spread |
| Fat that is spread (thick amount) onto sliced bread: ticked polyunsaturated margarine (e.g Flora), very low fat | bread_sliced_spread_polymarg_vlowfat_thick | 12 | bread_sliced_spread_polymarg_vlowfat_thick | 10 | -17% | Food portion sizes / MAFF; food composition as low fat spread |
| Fat that is spread (thin amount) onto sliced bread: ticked polyunsaturated margarine (e.g Flora), very low fat | bread_sliced_spread_polymarg_vlowfat_thin | 7 | bread_sliced_spread_polymarg_vlowfat_thin | 5 | -29% | Food portion sizes / MAFF; food composition as low fat spread |
| Fat that is spread (medium amount) onto sliced bread: ticked soya/vegan/dairy free margarine e.g Pure, and also cholesterol lowering | bread_sliced_spread_soya_chol_med | 10 | bread_sliced_spread_soya_chol_med | 7 | -30% | Food portion sizes / MAFF |
| Fat that is spread (thick amount) onto sliced bread: ticked soya/vegan/dairy free margarine e.g Pure, and also cholesterol lowering | bread_sliced_spread_soya_chol_thick | 12 | bread_sliced_spread_soya_chol_thick | 10 | -17% | Food portion sizes / MAFF |
| Fat that is spread (thin amount) onto sliced bread: ticked soya/vegan/dairy free margarine e.g Pure, and also cholesterol lowering | bread_sliced_spread_soya_chol_thin | 7 | bread_sliced_spread_soya_chol_thin | 5 | -29% | Food portion sizes / MAFF |
| Fat that is spread (medium amount) onto sliced bread: ticked soya/vegan/dairy free margarine e.g Pure, and not specified amount of fat | bread_sliced_spread_soya_dunno_med | 10 | bread_sliced_spread_soya_dunno_med | 7 | -30% | Food portion sizes / MAFF |
| Fat that is spread (thick amount) onto sliced bread: ticked soya/vegan/dairy free margarine e.g Pure, and not specified amount of fat | bread_sliced_spread_soya_dunno_thick | 12 | bread_sliced_spread_soya_dunno_thick | 10 | -17% | Food portion sizes / MAFF |
| Fat that is spread (thin amount) onto sliced bread: ticked soya/vegan/dairy free margarine e.g Pure, and not specified amount of fat | bread_sliced_spread_soya_dunno_thin | 7 | bread_sliced_spread_soya_dunno_thin | 5 | -29% | Food portion sizes / MAFF |
| Fat that is spread (medium amount) onto sliced bread: ticked soya/vegan/dairy free margarine (e.g Pure), normal amount % fat | bread_sliced_spread_soya_fat_med | 10 | bread_sliced_spread_soya_fat_med | 7 | -30% | Food portion sizes / MAFF; normal fat is taken as reduced fat (up to 62% fat) |
| Fat that is spread (thick amount) onto sliced bread: ticked soya/vegan/dairy free margarine (e.g Pure), normal amount % fat | bread_sliced_spread_soya_fat_thick | 12 | bread_sliced_spread_soya_fat_thick | 10 | -17% | Food portion sizes / MAFF; normal fat is taken as reduced fat (up to 62% fat) |
| Fat that is spread (thin amount) onto sliced bread: ticked soya/vegan/dairy free margarine (e.g Pure), normal amount % fat | bread_sliced_spread_soya_fat_thin | 7 | bread_sliced_spread_soya_fat_thin | 5 | -29% | Food portion sizes / MAFF; normal fat is taken as reduced fat (up to 62% fat) |
| Fat that is spread (medium amount) onto sliced bread: ticked soya/vegan/dairy free margarine (e.g Pure), low fat | bread_sliced_spread_soya_lowfat_med | 10 | bread_sliced_spread_soya_lowfat_med | 7 | -30% | Food portion sizes / MAFF |
| Fat that is spread (thick amount) onto sliced bread: ticked soya/vegan/dairy free margarine (e.g Pure), low fat | bread_sliced_spread_soya_lowfat_thick | 12 | bread_sliced_spread_soya_lowfat_thick | 10 | -17% | Food portion sizes / MAFF |
| Fat that is spread (thin amount) onto sliced bread: ticked soya/vegan/dairy free margarine (e.g Pure), low fat | bread_sliced_spread_soya_lowfat_thin | 7 | bread_sliced_spread_soya_lowfat_thin | 5 | -29% | Food portion sizes / MAFF |
| Fat that is spread (medium amount) onto sliced bread: ticked soya/vegan/dairy free margarine (e.g Pure), very low fat | bread_sliced_spread_soya_vlowfat_med | 10 | bread_sliced_spread_soya_vlowfat_med | 7 | -30% | Food portion sizes / MAFF; food composition as low fat spread |
| Fat that is spread (thick amount) onto sliced bread: ticked soya/vegan/dairy free margarine (e.g Pure), very low fat | bread_sliced_spread_soya_vlowfat_thick | 12 | bread_sliced_spread_soya_vlowfat_thick | 10 | -17% | Food portion sizes / MAFF; food composition as low fat spread |
| Fat that is spread (thin amount) onto sliced bread: ticked soya/vegan/dairy free margarine (e.g Pure), very low fat | bread_sliced_spread_soya_vlowfat_thin | 7 | bread_sliced_spread_soya_vlowfat_thin | 5 | -29% | Food portion sizes / MAFF; food composition as low fat spread |
| Sliced bread: not specified type so includes white, wholemeal, brown | bread_sliced_unanswered | 36 | bread_sliced_unanswered | 36 | 0% | |
| Sliced bread: white | bread_sliced_white | 36 | bread_sliced_white | 36 | 0% | |
| Sliced bread: wholemeal | bread_sliced_wholemeal | 36 | bread_sliced_wholemeal | 36 | 0% | |
| Cake: also includes muffin, flapjack, brownie, pecan pie | cake | 60 | cake | 60 | 0% | |
| Cake: also includes muffin, flapjack, brownie, pecan pie | | | cake_gf | 60 | - | Gluten free version added to the updated version (however no gluten free code available at the time so the non-gluten free code was used) |
| Cereal bar: includes with fruit and nuts, coated/uncoated | cerealbar | 28 | cerealbar | 28 | 0% | |
| Cereal bar: includes with fruit and nuts, coated/uncoated | | | cerealbar_gf | 28 | - | Gluten free version added to the updated version (however no gluten free code available at the time so the non-gluten free code was used) |
| Bran type breakfast cereal: e.g Bran Flakes, All Bran | cereal_bran | 50 | cereal_bran | 50 | 0% | |
| Bran type breakfast cereal with dried fruit: e.g Fruit 'n' Fibre | cereal_bran_driedfruit | 52 | cereal_bran_driedfruit | 52 | 0% | |
| Muesli e.g Alpen | cereal_muesli | 100 | cereal_muesli | 80 | -20% | PHAT photo series 6, image 4/5 |
| Muesli with extra fruit and nuts | cereal_muesli_driedfruit | 100 | cereal_muesli_driedfruit | 80 | -20% | PHAT photo series 6, image 4/5 |
| Sweetened oat crunch type cereal e.g. Jordan's Country Crisp, Nut clusters | cereal_oatcrunch | 100 | cereal_oatcrunch | 80 | -20% | PHAT photo series 6, image 4/5 |
| Sweetened oat crunch type cereal with extra dried fruit | cereal_oatcrunch_driedfruit | 100 | cereal_oatcrunch_driedfruit | 80 | -20% | PHAT photo series 6, image 4/5 |
| Cereal not specified: includes Cheerio's, Oatibix | cereal_other | 44 | cereal_other | 44 | 0% | Updated mapping based on free text entered by study participants, e.g. oatiflakes |
| Cereal not specified but with dried fruit: includes Cheerio's, Oatiflakes with fruit | cereal_other_driedfruit | 55 | cereal_other_driedfruit | 55 | 0% | Updated mapping based on free text entered by study participants, e.g. oatiflakes |
| Cereals plain: e.g Cornflakes, Rice Krispies | cereal_plain | 30 | cereal_plain | 30 | 0% | |
| Cereals plain with dried fruit: e.g Cornflakes, Rice Krispies, Special K with extra dried fruit | cereal_plain_driedfruit | 44 | cereal_plain_driedfruit | 44 | 0% | |
| Porridge (including instant) made with cholesterol lowering milk | cereal_porridge_milk | 203.5 | cereal_porridge_milk_chol | 203.5 | 0% | Milk type taken into account in the updated version |

| | McCance and Widdowson’s | | Nutrient databank + other changes | | | |
|---|---|---|---|---|---|---|
| **Variable description** | **Food item** | **Portion size** | **Food item** | **Portion size** | **Portion diff%** | **Description of differences with previous version** |
| Porridge (including instant) made with cholesterol lowering milk and added dried fruit | cereal_porridge_milk_driedfruit | 203.5 | cereal_porridge_milk_chol_driedfruit | 233.5 | 15% | Milk type taken into account in the updated version + portion size increased to account for dried fruit added after cooking |
| Porridge (including instant) made with milk (cow's) unspecified (e.g semi skimmed, skimmed, 1% milk) | cereal_porridge_milk | 203.5 | cereal_porridge_milk_dontknow | 203.5 | 0% | Milk type taken into account in the updated version |
| Porridge (including instant) made with milk (cow's) unspecified (e.g semi skimmed, skimmed, 1% milk) and added dried fruit | cereal_porridge_milk_driedfruit | 203.5 | cereal_porridge_milk_dontknow_driedfruit | 233.5 | 15% | Milk type taken into account in the updated version + portion size increased to account for dried fruit added after cooking |
| Porridge (including instant) made with goats or sheep milk | cereal_porridge_milk | 203.5 | cereal_porridge_milk_goatsheep | 203.5 | 0% | Milk type taken into account in the updated version |
| Porridge (including instant) made with goats or sheep milk and added dried fruit | cereal_porridge_milk_driedfruit | 203.5 | cereal_porridge_milk_goatsheep_driedfruit | 233.5 | 15% | Milk type taken into account in the updated version + portion size increased to account for dried fruit added after cooking |
| Porridge (including instant) made with other milk (e.g 1% milk, lactose free milk, almond milk) | cereal_porridge_milk | 203.5 | cereal_porridge_milk_other | 203.5 | 0% | Milk type taken into account in the updated version |
| Porridge (including instant) made with other milk (e.g 1% milk, lactose free milk, almond milk) and added dried fruit | cereal_porridge_milk_driedfruit | 203.5 | cereal_porridge_milk_other_driedfruit | 233.5 | 15% | Milk type taken into account in the updated version + portion size increased to account for dried fruit added after cooking |
| Porridge (including instant) made with powdered milk made up | cereal_porridge_milk | 203.5 | cereal_porridge_milk_powdered | 203.5 | 0% | Milk type taken into account in the updated version |
| Porridge (including instant) made with powdered milk made up and added dried fruit | cereal_porridge_milk_driedfruit | 203.5 | cereal_porridge_milk_powdered_driedfruit | 233.5 | 15% | Milk type taken into account in the updated version + portion size increased to account for dried fruit added after cooking |
| Porridge (including instant) made with rice, oat, almond or coconut milk | cereal_porridge_milk | 203.5 | cereal_porridge_milk_riceoatveg | 203.5 | 0% | Milk type taken into account in the updated version |
| Porridge (including instant) made with rice, oat, almond or coconut milk and added dried fruit | cereal_porridge_milk_driedfruit | 203.5 | cereal_porridge_milk_riceoatveg_driedfruit | 233.5 | 15% | Milk type taken into account in the updated version + portion size increased to account for dried fruit added after cooking |
| Porridge (including instant) made with semi skimmed milk | cereal_porridge_milk | 203.5 | cereal_porridge_milk_semi | 203.5 | 0% | Milk type taken into account in the updated version |
| Porridge (including instant) made with semi skimmed milk and added dried fruit | cereal_porridge_milk_driedfruit | 203.5 | cereal_porridge_milk_semi_driedfruit | 233.5 | 15% | Milk type taken into account in the updated version + portion size increased to account for dried fruit added after cooking |
| Porridge (including instant) made with skimmed milk | cereal_porridge_milk | 203.5 | cereal_porridge_milk_skimmed | 203.5 | 0% | Milk type taken into account in the updated version |
| Porridge (including instant) made with skimmed milk and added dried fruit | cereal_porridge_milk_driedfruit | 203.5 | cereal_porridge_milk_skimmed_driedfruit | 233.5 | 15% | Milk type taken into account in the updated version + portion size increased to account for dried fruit added after cooking |
| Porridge (including instant) made with soya milk with added calcium | cereal_porridge_milk | 203.5 | cereal_porridge_milk_soya_ca | 203.5 | 0% | Milk type taken into account in the updated version |
| Porridge (including instant) made with soya milk with added calcium and added dried fruit | cereal_porridge_milk_driedfruit | 203.5 | cereal_porridge_milk_soya_ca_driedfruit | 233.5 | 15% | Milk type taken into account in the updated version + portion size increased to account for dried fruit added after cooking |
| Porridge (including instant) made with soya milk with no added calcium | cereal_porridge_milk | 203.5 | cereal_porridge_milk_soya_noca | 203.5 | 0% | Milk type taken into account in the updated version |
| Porridge (including instant) made with soya milk with no added calcium and added dried fruit | cereal_porridge_milk_driedfruit | 203.5 | cereal_porridge_milk_soya_noca_driedfruit | 233.5 | 15% | Milk type taken into account in the updated version + portion size increased to account for dried fruit added after cooking |
| Porridge (including instant) made with whole milk | cereal_porridge_milk | 203.5 | cereal_porridge_milk_whole | 203.5 | 0% | Milk type taken into account in the updated version |
| Porridge (including instant) made with whole milk and added dried fruit | cereal_porridge_milk_driedfruit | 203.5 | cereal_porridge_milk_whole_driedfruit | 233.5 | 15% | Milk type taken into account in the updated version + portion size increased to account for dried fruit added after cooking |
| Porridge (including instant) made with water | cereal_porridge_water | 203.5 | cereal_porridge_water | 203.5 | 0% | |
| Porridge (including instant) made with water and added dried fruit | cereal_porridge_water_driedfruit | 203.5 | cereal_porridge_water_driedfruit | 233.5 | 15% | Portion size increased to account for dried food |
| Sugar added onto cereal | cereal_sugar | 6 | cereal_sugar | 6 | 0% | |
| Sweetened cereals e.g Coco Pops, Honey Nut Cornflakes, Ricicles | cereal_sweet | 38 | cereal_sweet | 38 | 0% | |
| Sweetened cereals e.g Coco Pops, Honey Nut Cornflakes, Ricicles with added dried fruit | cereal_sweet_driedfruit | 52 | cereal_sweet_driedfruit | 52 | 0% | Dried fruit added to cereals |
| Wholewheat cereals e.g Weetabix, Shredded Wheat, Shreddies | cereal_wwheat | 44 | cereal_wwheat | 44 | 0% | |
| Wholewheat cereals e.g Weetabix, Shredded Wheat, Shreddies with added dried fruit | cereal_wwheat_driedfruit | 44 | cereal_wwheat_driedfruit | 44 | 0% | Cereals containing dried fruit |
| Blue cheese: Stilton, Danish Blue, Roquefort | cheese_blue | 35 | cheese_blue | 35 | 0% | |
| Cheesecake | cheesecake | 110 | cheesecake | 110 | 0% | |
| Cheesecake | | | cheesecake_gf | 110 | - | Gluten free version added to the updated version (however no gluten free code available at the time so the non-gluten free code was used) |
| Cottage cheese | cheese_cottage | 60 | cheese_cottage | 60 | 0% | |
| Feta cheese | cheese_feta | 40 | cheese_feta | 40 | 0% | |
| Goat's cheese | cheese_goat | 40 | cheese_goat | 40 | 0% | |
| Hard cheese e.g Cheddar | cheese_hard | 40 | cheese_hard | 40 | 0% | |
| Low fat hard cheese e.g Cheddar low fat | cheese_hard_lof | 40 | cheese_hard_lof | 40 | 0% | |
| Mozzarella cheese | cheese_mozzarella | 40 | cheese_mozzarella | 40 | 0% | |
| Other cheese e.g Wensleydale, Halloumi | cheese_other | 40 | cheese_other | 40 | 0% | Updated mapping based on free text entered by study participants, e.g. haloumi |
| Soft cheese e.g Brie | cheese_soft | 40 | cheese_soft | 40 | 0% | |
| Spreadable cheese e.g cream cheese, cheese triangles | cheese_spread | 15 | cheese_spread | 15 | 0% | |
| Low fat spreadable cheese e.g cream cheese, cheese triangles | cheese_spread_lof | 15 | cheese_spread_lof | 15 | 0% | |
| Chocolate bar e.g Crunchie, Snickers | choc_bar | 50 | choc_bar | 50 | 0% | |
| Dark chocolate | choc_dark | 50 | choc_dark | 50 | 0% | |
| Milk chocolate | choc_milk | 50 | choc_milk | 50 | 0% | |
| Chocolate sweets e.g Roses, Milk Tray | choc_sweets | 36 | choc_sweets | 36 | 0% | |
| White chocolate | choc_white | 50 | choc_white | 50 | 0% | |
| Milk chocolate or white chocolate coated raisins | chocyog_raisin | 25 | chocyog_raisin | 25 | 0% | |
| Chutney or pickle | chutney | 20 | chutney | 20 | 0% | |
| Shot of strong coffee for cappuccino | | | cof_capp | 30 | - | Shot of strong coffee for cappuccino separated from milk in updated version |
| Shot of strong decaffeinated coffee for cappuccino | | | cof_capp_decaf | 30 | - | Shot of strong coffee for cappuccino separated from milk in updated version - decaffeinated food code added |
| Milk in cappuccino: cholesterol lowering | cof_capp_other | 190 | cof_capp_milk_chol | 160 | 0% | Milk type taken into account in the updated version; portion size only for milk (30ml shot of coffee in separate food item); sum weight no change |
| Milk in cappuccino: milk (cow's) unspecified (e.g semi skimmed, skimmed, 1% milk) | cof_capp_other | 190 | cof_capp_milk_dontknow | 160 | 0% | Milk type taken into account in the updated version; portion size only for milk (30ml shot of coffee in separate food item); sum weight no change |
| Milk in cappuccino: goat's or sheep's milk | cof_capp_other | 190 | cof_capp_milk_goatsheep | 160 | 0% | Milk type taken into account in the updated version; portion size only for milk (30ml shot of coffee in separate food item); sum weight no change |
| Milk in cappuccino: other milk (e.g 1% milk, lactose free milk, almond milk) | cof_capp_other | 190 | cof_capp_milk_other | 160 | 0% | Milk type taken into account in the updated version; portion size only for milk (30ml shot of coffee in separate food item); sum weight no change |
| Milk in cappuccino: powdered milk made up | cof_capp_other | 190 | cof_capp_milk_powdered | 160 | 0% | Milk type taken into account in the updated version; portion size only for milk (30ml shot of coffee in separate food item); sum weight no change |
| Milk in cappuccino: rice, oat, almond or coconut milk | cof_capp_other | 190 | cof_capp_milk_riceoatveg | 160 | 0% | Milk type taken into account in the updated version; portion size only for milk (30ml shot of coffee in separate food item); sum weight no change |
| Milk in cappuccino: semi skimmed milk | cof_capp_semi | 190 | cof_capp_milk_semi | 160 | 0% | Milk type taken into account in the updated version; portion size only for milk (30ml shot of coffee in separate food item); sum weight no change |
| Milk in cappuccino: skimmed milk | cof_capp_skimmed | 190 | cof_capp_milk_skimmed | 160 | 0% | Milk type taken into account in the updated version; portion size only for milk (30ml shot of coffee in separate food item); sum weight no change |
| Milk in cappuccino: soya milk with added calcium | cof_capp_other | 190 | cof_capp_milk_soya_ca | 160 | 0% | Milk type taken into account in the updated version; portion size only for milk (30ml shot of coffee in separate food item); sum weight no change |
| Milk in cappuccino: soya milk with no added calcium | cof_capp_other | 190 | cof_capp_milk_soya_noca | 160 | 0% | Milk type taken into account in the updated version; portion size only for milk (30ml shot of coffee in separate food item); sum weight no change |
| Milk in cappuccino: whole milk | cof_capp_whole | 190 | cof_capp_milk_whole | 160 | 0% | Milk type taken into account in the updated version; portion size only for milk (30ml shot of coffee in separate food item); sum weight no change |
| Shot of strong coffee for espresso | cof_espresso | 30 | cof_espresso | 30 | 0% | |
| Shot of strong decaffeinated coffee for espresso | cof_espresso_decaf | 30 | cof_espresso_decaf | 30 | 0% | Decaffeinated food code added |
| Filter coffee | cof_filter | 190 | cof_filter | 190 | 0% | |
| Filter coffee decaffeinated | cof_filter_decaf | 190 | cof_filter_decaf | 190 | 0% | Decaffeinated food code added |
| Instant coffee | cof_instant | 190 | cof_instant | 190 | 0% | |
| Instant coffee decaffeinated | cof_instant_decaf | 190 | cof_instant_decaf | 190 | 0% | Decaffeinated food code added |
| Shot of strong coffee for latte | | | cof_latte | 30 | 0% | Shot of strong coffee for cappuccino separated from milk in updated version |
| Shot of strong decaffeinated coffee for latte | | | cof_latte_decaf | 30 | 0% | Shot of strong coffee for cappuccino separated from milk in updated version - decaffeinated food code added |
| Milk in latte: cholesterol lowering | cof_latte_decaf_other | 190 | cof_latte_milk_chol | 160 | 0% | Milk type taken into account in the updated version; portion size only for milk (30ml shot of coffee in separate food item); sum weight no change |
| Milk in latte: milk (cow's) unspecified (e.g semi skimmed, skimmed, 1% milk) | cof_latte_decaf_other | 190 | cof_latte_milk_dontknow | 160 | 0% | Milk type taken into account in the updated version; portion size only for milk (30ml shot of coffee in separate food item); sum weight no change |
| Milk in latte: goat's or sheep's milk | cof_latte_decaf_other | 190 | cof_latte_milk_goatsheep | 160 | 0% | Milk type taken into account in the updated version; portion size only for milk (30ml shot of coffee in separate food item); sum weight no change |
| Milk in latte: other milk (e.g 1% milk, lactose free milk, almond milk) | cof_latte_decaf_other | 190 | cof_latte_milk_other | 160 | 0% | Milk type taken into account in the updated version; portion size only for milk (30ml shot of coffee in separate food item); sum weight no change |
| Milk in latte: powdered milk made up | cof_latte_decaf_other | 190 | cof_latte_milk_powdered | 160 | 0% | Milk type taken into account in the updated version; portion size only for milk (30ml shot of coffee in separate food item); sum weight no change |
| Milk in latte: rice, oat, almond or coconut milk | cof_latte_decaf_other | 190 | cof_latte_milk_riceoatveg | 160 | 0% | Milk type taken into account in the updated version; portion size only for milk (30ml shot of coffee in separate food item); sum weight no change |
| Milk in latte: semi skimmed milk | cof_latte_decaf_semi | 190 | cof_latte_milk_semi | 160 | 0% | Milk type taken into account in the updated version; portion size only for milk (30ml shot of coffee in separate food item); sum weight no change |
| Milk in latte: skimmed milk | cof_latte_decaf_skimmed | 190 | cof_latte_milk_skimmed | 160 | 0% | Milk type taken into account in the updated version; portion size only for milk (30ml shot of coffee in separate food item); sum weight no change |
| Milk in latte: soya milk with added calcium | cof_latte_decaf_other | 190 | cof_latte_milk_soya_ca | 160 | 0% | Milk type taken into account in the updated version; portion size only for milk (30ml shot of coffee in separate food item); sum weight no change |
| Milk in latte: soya milk with no added calcium | cof_latte_decaf_other | 190 | cof_latte_milk_soya_noca | 160 | 0% | Milk type taken into account in the updated version; portion size only for milk (30ml shot of coffee in separate food item); sum weight no change |
| Milk in latte: whole milk | cof_latte_decaf_whole | 190 | cof_latte_milk_whole | 160 | 0% | Milk type taken into account in the updated version; portion size only for milk (30ml shot of coffee in separate food item); sum weight no change |
| Other unspecified coffee | cof_other | 190 | cof_other | 190 | 0% | Updated mapping based on free text entered by study participants, e.g. mixture of instant and filter coffee |
| Other unspecified coffee decaffeinated | cof_other_decaf | 190 | cof_other_decaf | 190 | 0% | Decaffeinated food code added |
| Sugar added to coffee | cof_sugar | 6 | cof_sugar | 6 | 0% | |
| Cream e.g. single, double, sour, crème fraîche | cream | 30 | cream | 30 | 0% | |
| Croissant | croissant | 60 | croissant | 60 | 0% | |
| Crumble topping | crumble | 70 | crumble | 70 | 0% | |
| Danish pastries | danish_pastry | 110 | danish_pastry | 110 | 0% | |
| Milk based desserts other e.g mousse, tiramisu, crème caramel | dessert_milkbased | 60 | dessert_milkbased | 60 | 0% | |
| Milk based desserts e.g custard, rice pudding, blancmange | dessert_milkpuds | 200 | dessert_milkpuds | 200 | 0% | |
| Other desserts e.g apple pie, stewed fruit, trifle | dessert_other | 60 | dessert_other | 60 | 0% | |
| Soya ice cream, soya yogurt, other soya dessert | dessert_soya | 125 | dessert_soya | 125 | 0% | |
| Double crust pie or pasty | double_crust | 60 | double_crust | 60 | 0% | |
| Doughnut | doughnut | 60 | doughnut | 60 | 0% | |
| Low sugar/low fat hot chocolate drink | drink_diethotchoc | 260 | drink_diethotchoc | 260 | 0% | |
| Fizzy/carbonated soft drinks | drink_fizzy | 330 | drink_fizzy | 330 | 0% | |
| Grapefruit juice | drink_grapefruit | 250 | drink_grapefruit | 250 | 0% | Replaced concentrated juice with 'as served' |
| Hot chocolate: chocolate powder and water | | | drink_hotchoc | 143 | - | Chocolate powder and water in updated version (milk separate for non-instant version of hot chocolate) |
| Milk in hot chocolate: cholesterol lowering | drink_hotchoc_other | 260 | drink_hotchoc_milk_chol | 117 | 0% | Milk type taken into account in the updated version + portion size now only for milk (143ml of chocolate in separate food item) |
| Milk in hot chocolate: milk (cow's) unspecified (e.g semi skimmed, skimmed, 1% milk) | drink_hotchoc_other | 260 | drink_hotchoc_milk_dontknow | 117 | 0% | Milk type taken into account in the updated version + portion size now only for milk (143ml of chocolate in separate food item) |
| Milk in hot chocolate: goat's or sheep's milk | drink_hotchoc_other | 260 | drink_hotchoc_milk_goatsheep | 117 | 0% | Milk type taken into account in the updated version + portion size now only for milk (143ml of chocolate in separate food item) |
| Milk in hot chocolate: other milk (e.g 1% milk, lactose free milk, almond milk) | drink_hotchoc_other | 260 | drink_hotchoc_milk_other | 117 | 0% | Milk type taken into account in the updated version + portion size now only for milk (143ml of chocolate in separate food item) |
| Milk in hot chocolate: powdered milk made up | drink_hotchoc_other | 260 | drink_hotchoc_milk_powdered | 117 | 0% | Milk type taken into account in the updated version + portion size now only for milk (143ml of chocolate in separate food item) |
| Milk in hot chocolate: rice, oat, almond or coconut milk | drink_hotchoc_other | 260 | drink_hotchoc_milk_riceoatveg | 117 | 0% | Milk type taken into account in the updated version + portion size now only for milk (143ml of chocolate in separate food item) |
| Milk in hot chocolate: semi skimmed milk | drink_hotchoc_semi | 260 | drink_hotchoc_milk_semi | 117 | 0% | Milk type taken into account in the updated version + portion size now only for milk (143ml of chocolate in separate food item) |
| Milk in hot chocolate: skimmed milk | drink_hotchoc_skimmed | 260 | drink_hotchoc_milk_skimmed | 117 | 0% | Milk type taken into account in the updated version + portion size now only for milk (143ml of chocolate in separate food item) |
| Milk in hot chocolate: soya milk with added calcium | drink_hotchoc_other | 260 | drink_hotchoc_milk_soya_ca | 117 | 0% | Milk type taken into account in the updated version + portion size now only for milk (143ml of chocolate in separate food item) |
| Milk in hot chocolate: soya milk with no added calcium | drink_hotchoc_other | 260 | drink_hotchoc_milk_soya_noca | 117 | 0% | Milk type taken into account in the updated version + portion size now only for milk (143ml of chocolate in separate food item) |
| Milk in hot chocolate: whole milk | drink_hotchoc_whole | 260 | drink_hotchoc_milk_whole | 117 | 0% | Milk type taken into account in the updated version + portion size now only for milk (143ml of chocolate in separate food item) |
| Low calorie/low sugar drink, still or fizzy | drink_lowcal | 330 | drink_lowcal | 330 | 0% | |

| | McCance and Widdowson’s | | Nutrient databank + other changes | | | |
|---|---|---|---|---|---|---|
| Variable description | Food item | Portion size | Food item | Portion size | Portion diff% | Description of differences with previous version |
| Milk based drink e.g Yogurt drinks, flavoured milk or milkshakes | drink_milkbased | 250 | drink_milkbased | 250 | 0% | |
| Orange juice | drink_orange | 250 | drink_orange | 250 | 0% | |
| Drink other e.g Barley cup, other mixed fruit drink | drink_other | 260 | drink_other | 260 | 0% | Updated mapping based on free text entered by study participants, now includes water, milk and barley cup |
| Pure juice other e.g apple, pineapple, tomato, cranberry | drink_purejuice | 250 | drink_purejuice | 250 | 0% | |
| Fruit squash or cordial | drink_squash | 250 | drink_squash | 250 | 0% | |
| Water (still/sparkling) | drink_water | 250 | drink_water | 250 | 0% | Replaced club soda with mineral water |
| Drizzle of oil | drizzle_oil | 10 | drizzle_oil | 10 | 0% | |
| Omelette or scrambled egg | egg_omelet | 120 | egg_omelet | 120 | 0% | Weight based on 2 eggs |
| Other egg dish e.g quiche | egg_other | 75 | egg_other | 75 | 0% | Updated mapping based on free text entered by study participants, e.g. now includes quiche |
| Scotch egg | egg_scotch | 120 | egg_scotch | 120 | 0% | |
| Egg mayonnaise | egg_swich | 120 | egg_swich | 120 | 0% | Weight based on 2 eggs |
| Eggs boiled, poached, fried | egg_whole | 50 | egg_whole | 50 | 0% | |
| White fish in batter | fish_battered | 190 | fish_battered | 150 | -21% | 50% takeaway (180 g) and 50% shop (120 g) |
| White fish in breadcrumbs | fish_breaded | 56 | fish_breaded | 100 | 79% | Now includes breaded fish as well as fish fingers and fish cakes, weight was amended accordingly |
| Lobster or crab | fish_lobcrab | 85 | fish_lobcrab | 85 | 0% | |
| Oily fish e.g salmon, mackerel, herring | fish_oily | 100 | fish_oily | 100 | 0% | |
| Other fish e.g trout, squid, fish pate | fish_other | 100 | fish_other | 100 | 0% | Updated mapping based on free text entered by study participants, e.g. smoked salmon |
| Prawns | fish_prawns | 60 | fish_prawns | 60 | 0% | |
| Shell fish e.g scallops, muscles | fish_shell | 40 | fish_shell | 40 | 0% | |
| Tinned tuna | fish_tinnedtuna | 92 | fish_tinnedtuna | 92 | 0% | |
| White fish e.g cod, haddock | fish_white | 120 | fish_white | 120 | 0% | |
| Apple | fruit_apple | 100 | fruit_apple | 112 | 12% | Weight increased to reflect the inedible part included in the food composition |
| Banana | fruit_banana | 100 | fruit_banana | 100 | 0% | |
| Berries e.g strawberry, raspberry, blueberry, blackberry | fruit_berry | 40 | fruit_berry | 40 | 0% | |
| Fruitcake iced or plain | fruitcake | 70 | fruitcake | 70 | 0% | |
| Fruitcake iced or plain | | | fruitcake_gf | 70 | - | Gluten free version added to the updated version (however no gluten free code available at the time so the non-gluten free code was used) |
| Cherries | fruit_cherry | 24 | fruit_cherry | 34 | 42% | Approximately 8 cherries (MAFF) |
| Dried fruit e.g raisins, dates, dried figs, dried apricots | fruit_dried | 60 | fruit_dried | 60 | 0% | |
| Grapefruit | fruit_grapefruit | 160 | fruit_grapefruit | 148 | -8% | added canned grapefruit, amended portion weight accordingly |
| Grapes: red, green, black | fruit_grapes | 100 | fruit_grapes | 100 | 0% | |
| Mango | fruit_mango | 100 | fruit_mango | 113 | 13% | Based on participant data: mean weight of 1/2 and 1 unit of mango. |
| Melon: honeydew, cantaloupe, watermelon | fruit_melon | 180 | fruit_melon | 180 | 0% | |
| Fresh or tinned fruit salad with or without juice/syrup | fruit_mixed | 105 | fruit_mixed | 105 | 0% | |
| Orange | fruit_orange | 120 | fruit_orange | 120 | 0% | |
| Other fruit e.g kiwi, papaya, pomegranate, apricot, fig | fruit_other | 60 | fruit_other | 60 | 0% | Updated mapping based on free text entered by study participants |
| Peach | fruit_peach | 150 | fruit_peach | 130 | -13% | Weight decreased to reflect the inedible part excluded in the food composition |
| Pear | fruit_pear | 120 | fruit_pear | 160 | 33% | 50% comice 50% conference |
| Pineapple | fruit_pineapple | 80 | fruit_pineapple | 80 | 0% | |
| Plum | fruit_plum | 55 | fruit_plum | 55 | 0% | |
| Prunes | fruit_prunes | 60 | fruit_prunes | 60 | 0% | |
| Tangerine, clementine, mandarin | fruit_satsuma | 70 | fruit_satsuma | 70 | 0% | |
| Stewed fruit | fruit_stewed | 140 | fruit_stewed | 140 | 0% | |
| Cous cous | grains_couscous | 150 | grains_couscous | 150 | 0% | |
| Other grains e.g barley, bulgarwheat, quinoa | grains_other | 157 | grains_other | 157 | 0% | Updated mapping based on free text entered by study participants |
| Guacamole | guacamole | 26 | guacamole | 26 | 0% | |
| Hummus | hummus | 26 | hummus | 26 | 0% | |
| Ice-cream: dairy hard scoop, non dairy soft scoop, non dairy choc ice | icecream | 120 | icecream | 120 | 0% | |
| Indian snack e.g pakora, onion bahji, samosa | indian_snack | 40 | indian_snack | 40 | 0% | |
| Jam, marmalade, honey, golden syrup | jam_honey | 18 | jam_honey | 18 | 0% | |
| Mayonnaise | mayo | 30 | mayo | 30 | 0% | |
| Low fat mayonnaise | mayo_lowfat | 30 | mayo_lowfat | 30 | 0% | |
| Bacon rasher unsmoked lean/fat trimmed | meat_bacon_nofat | 46 | meat_bacon_nofat | 46 | 0% | |
| Bacon rasher untrimmed/with fat | meat_bacon_withfat | 46 | meat_bacon_withfat | 46 | 0% | |
| Beef lean only: roast, steak, burger, minced, curry | meat_beef_nofat | 120 | meat_beef_nofat | 120 | 0% | |
| Beef with fat: roast, steak, burger, minced, curry | meat_beef_withfat | 120 | meat_beef_withfat | 120 | 0% | |
| Ham unspecified | meat_ham_nofat | 23 | meat_ham_nofat | 23 | 0% | |
| Parma ham, salami, pastrami | meat_ham_withfat | 23 | meat_ham_withfat | 23 | 0% | |
| Lamb lean only: chops, roast, stewed, burger | meat_lamb_nofat | 120 | meat_lamb_nofat | 120 | 0% | |
| Lamb with fat: chops, roast, stewed, burger | meat_lamb_withfat | 120 | meat_lamb_withfat | 120 | 0% | |
| Pig's or lambs liver or liver pate | meat_liverpate | 70 | meat_liverpate | 70 | 0% | |
| Duck, venison, goose | meat_other | 100 | meat_other | 100 | 0% | Updated mapping based on free text entered by study participants, e.g. replaced offal with pheasant and venison |
| Pork lean only: roast, chops, steak, diced (e.g sweet and sour) | meat_pork_nofat | 120 | meat_pork_nofat | 120 | 0% | |
| Pork with fat: roast, chops, steak, diced (e.g sweet and sour) | meat_pork_withfat | 120 | meat_pork_withfat | 120 | 0% | |
| Sausages, pork or beef | meat_sausage | 30 | meat_sausage | 30 | 0% | |
| Milk on your cereal: cholesterol lowering milk | milk_chol_cereal | 100 | milk_chol_cereal | 100 | 0% | |
| Milk (splash) in instant or filter coffee or coffee infusion weak/strong: cholesterol lowering milk | milk_chol_coffee | 25 | milk_chol_coffee | 35 | 40% | Portion size increased to account for changes in mapping; coffeemate and cream now excluded |
| Milk in a glass: cholesterol lowering milk | milk_chol_glass | 259 | milk_chol_glass | 259 | 0% | |
| Milk (splash) in tea: cholesterol lowering milk | milk_chol_tea | 35 | milk_chol_tea | 35 | 0% | |
| Milk on your cereal: milk (cow's) unspecified (e.g semi skimmed, skimmed, 1% milk) | milk_dontknow_cereal | 100 | milk_dontknow_cereal | 100 | 0% | |
| Milk (splash) in instant or filter coffee or coffee infusion weak/strong: milk (cow's) unspecified (e.g semi skimmed, skimmed, 1% milk) | milk_dontknow_coffee | 25 | milk_dontknow_coffee | 35 | 40% | Portion size increased to account for changes in mapping; coffeemate and cream now excluded |
| Milk in a glass: milk (cow's) unspecified (e.g semi skimmed, skimmed, 1% milk) | milk_dontknow_glass | 259 | milk_dontknow_glass | 259 | 0% | |
| Milk (splash) in tea: milk (cow's) unspecified (e.g semi skimmed, skimmed, 1% milk) | milk_dontknow_tea | 35 | milk_dontknow_tea | 35 | 0% | |
| Milk on your cereal: goat's or sheep's milk | milk_goatsheep_cereal | 100 | milk_goatsheep_cereal | 100 | 0% | |
| Milk (splash) in instant or filter coffee or coffee infusion weak/strong: goat's or sheep's milk | milk_goatsheep_coffee | 35 | milk_goatsheep_coffee | 35 | 0% | |
| Milk in a glass: goat's or sheep's milk | milk_goatsheep_glass | 259 | milk_goatsheep_glass | 259 | 0% | |
| Milk (splash) in tea: goat's or sheep's milk | milk_goatsheep_tea | 35 | milk_goatsheep_tea | 35 | 0% | |
| Milk on your cereal: other milk (e.g 1% milk, lactose free milk, almond milk) | milk_other_cereal | 100 | milk_other_cereal | 100 | 0% | Updated mapping based on free text entered by study participants: 1% milk, almond milk, lactose free milk |
| Milk (splash) in instant or filter coffee or coffee infusion weak/strong: other milk (e.g 1% milk, lactose free milk, almond milk) | milk_other_coffee | 25 | milk_other_coffee | 35 | 40% | Updated mapping based on free text entered by study participants: 1% milk, almond milk, lactose free milk; coffeemate and cream now excluded |
| Milk in a glass: other milk (e.g 1% milk, lactose free milk, almond milk) | milk_other_glass | 259 | milk_other_glass | 259 | 0% | Updated mapping based on free text entered by study participants: 1% milk, almond milk, lactose free milk |
| Milk (splash) in tea: other milk (e.g 1% milk, lactose free milk, almond milk) | milk_other_tea | 35 | milk_other_tea | 35 | 0% | Updated mapping based on free text entered by study participants: 1% milk, almond milk, lactose free milk |
| Milk on your cereal: powdered milk made up | milk_powdered_cereal | 10 | milk_powdered_cereal | 100 | 900% | Portion size increased to account for changes in mapping; powdered milk, made up with water |
| Milk (splash) in instant or filter coffee or coffee infusion weak/strong: powdered milk made up | milk_powdered_coffee | 3 | milk_powdered_coffee | 4.5 | 50% | Heaped teaspoon of powdered milk * density |
| Milk in a glass: powdered milk made up | milk_powdered_glass | 24 | milk_powdered_glass | 259 | 979% | Portion size increased to account for changes in mapping; powdered milk, made up with water |
| Milk (splash) in tea: powdered milk made up | milk_powdered_tea | 3 | milk_powdered_tea | 3 | 0% | to be updated to 4.5 gram with next update |
| Milk on your cereal: rice, oat, almond or coconut milk | milk_riceoatveg_cereal | 100 | milk_riceoatveg_cereal | 100 | 0% | includes nut-based milks |
| Milk (splash) in instant or filter coffee or coffee infusion weak/strong: rice, oat, almond or coconut milk | milk_riceoatveg_coffee | 35 | milk_riceoatveg_coffee | 35 | 0% | includes nut-based milks |
| Milk in a glass: rice, oat, almond or coconut milk | milk_riceoatveg_glass | 259 | milk_riceoatveg_glass | 259 | 0% | includes nut-based milks |
| Milk (splash) in tea: rice, oat, almond or coconut milk | milk_riceoatveg_tea | 35 | milk_riceoatveg_tea | 35 | 0% | includes nut-based milks |
| Milk on your cereal: semi skimmed milk | milk_semi_cereal | 100 | milk_semi_cereal | 100 | 0% | |
| Milk (splash) in instant or filter coffee or coffee infusion weak/strong: semi skimmed milk | milk_semi_coffee | 25 | milk_semi_coffee | 35 | 40% | Portion size increased to account for changes in mapping; coffeemate and cream now excluded |
| Milk in a glass: semi skimmed milk | milk_semi_glass | 259 | milk_semi_glass | 259 | 0% | |
| Milk (splash) in tea: semi skimmed milk | milk_semi_tea | 35 | milk_semi_tea | 35 | 0% | |
| Milk on your cereal: skimmed milk | milk_skimmed_cereal | 100 | milk_skimmed_cereal | 100 | 0% | |
| Milk (splash) in instant or filter coffee or coffee infusion weak/strong: skimmed milk | milk_skimmed_coffee | 25 | milk_skimmed_coffee | 35 | 40% | Portion size increased to account for changes in mapping; coffeemate and cream now excluded |
| Milk in a glass: skimmed milk | milk_skimmed_glass | 259 | milk_skimmed_glass | 259 | 0% | |
| Milk (splash) in tea: skimmed milk | milk_skimmed_tea | 35 | milk_skimmed_tea | 35 | 0% | |
| Milk on your cereal: soya milk with added calcium | milk_soya_ca_cereal | 100 | milk_soya_ca_cereal | 100 | 0% | |
| Milk (splash) in instant or filter coffee or coffee infusion weak/strong: soya milk with added calcium | milk_soya_ca_coffee | 35 | milk_soya_ca_coffee | 35 | 0% | |
| Milk in a glass: soya milk with added calcium | milk_soya_ca_glass | 259 | milk_soya_ca_glass | 259 | 0% | |
| Milk (splash) in tea: soya milk with added calcium | milk_soya_ca_tea | 35 | milk_soya_ca_tea | 35 | 0% | |
| Milk on your cereal: soya milk with no added calcium | milk_soya_noca_cereal | 100 | milk_soya_noca_cereal | 100 | 0% | |
| Milk (splash) in instant or filter coffee or coffee infusion weak/strong: soya milk with no added calcium | milk_soya_noca_coffee | 35 | milk_soya_noca_coffee | 35 | 0% | |
| Milk in a glass: soya milk with no added calcium | milk_soya_noca_glass | 259 | milk_soya_noca_glass | 259 | 0% | |
| Milk (splash) in tea: soya milk with no added calcium | milk_soya_noca_tea | 35 | milk_soya_noca_tea | 35 | 0% | |
| Milk on your cereal: whole milk | milk_whole_cereal | 100 | milk_whole_cereal | 100 | 0% | |
| Milk (splash) in instant or filter coffee or coffee infusion weak/strong: whole milk | milk_whole_coffee | 25 | milk_whole_coffee | 35 | 40% | Portion size increased to account for changes in mapping; coffeemate and cream now excluded |
| Milk in a glass: whole milk | milk_whole_glass | 259 | milk_whole_glass | 259 | 0% | |
| Milk (splash) in tea: whole milk | milk_whole_tea | 35 | milk_whole_tea | 35 | 0% | |
| Oatcakes | oatcakes | 13 | oatcakes | 13 | 0% | |

| | McCance and Widdowson's | | Nutrient databank + other changes | | | |
|---|---|---|---|---|---|---|
| Variable description | Food item | Portion size | Food item | Portion size | Portion diff% | Description of differences with previous version |
| Fat that is spread (medium amount) onto oatcakes: ticked butter but not specified amount of % fat | oatcakes_spread_butter_dunno_med | 7 | oatcakes_spread_butter_dunno_med | 2.5 | -64% | 25% of weight on a slice; food composition as butter and spreadable butter |
| Fat that is spread (thick amount) onto oatcakes: ticked butter but not specified amount of % fat | oatcakes_spread_butter_dunno_thick | 10 | oatcakes_spread_butter_dunno_thick | 3 | -70% | 25% of weight on a slice; food composition as butter and spreadable butter |
| Fat that is spread (thin amount) onto oatcakes: ticked butter but not specified amount of % fat | oatcakes_spread_butter_dunno_thin | 5 | oatcakes_spread_butter_dunno_thin | 1.75 | -65% | 25% of weight on a slice; food composition as butter and spreadable butter |
| Fat that is spread (medium amount) onto oatcakes: ticked butter, normal amount of % fat | oatcakes_spread_butter_fat_med | 7 | oatcakes_spread_butter_fat_med | 2.5 | -64% | 25% of weight on a slice |
| Fat that is spread (thick amount) onto oatcakes: ticked butter, normal amount of % fat | oatcakes_spread_butter_fat_thick | 10 | oatcakes_spread_butter_fat_thick | 3 | -70% | 25% of weight on a slice |
| Fat that is spread (thin amount) onto oatcakes: ticked butter, normal amount of % fat | oatcakes_spread_butter_fat_thin | 5 | oatcakes_spread_butter_fat_thin | 1.75 | -65% | 25% of weight on a slice |
| Fat that is spread (medium amount) onto oatcakes: ticked butter, low fat | oatcakes_spread_butter_lowfat_med | 7 | oatcakes_spread_butter_lowfat_med | 2.5 | -64% | 25% of weight on a slice |
| Fat that is spread (thick amount) onto oatcakes: ticked butter, low fat | oatcakes_spread_butter_lowfat_thick | 10 | oatcakes_spread_butter_lowfat_thick | 3 | -70% | 25% of weight on a slice |
| Fat that is spread (thin amount) onto oatcakes: ticked butter, low fat | oatcakes_spread_butter_lowfat_thin | 5 | oatcakes_spread_butter_lowfat_thin | 1.75 | -65% | 25% of weight on a slice |
| Fat that is spread (medium amount) onto oatcakes: ticked spreadable butter with normal amount of % fat | oatcakes_spread_butter_spread_fat_med | 7 | oatcakes_spread_butter_spread_fat_med | 2.5 | -64% | 25% of weight on a slice |
| Fat that is spread (thick amount) onto oatcakes: ticked spreadable butter with normal amount of % fat | oatcakes_spread_butter_spread_fat_thick | 10 | oatcakes_spread_butter_spread_fat_thick | 3 | -70% | 25% of weight on a slice |
| Fat that is spread (thin amount) onto oatcakes: ticked spreadable butter with normal amount of % fat | oatcakes_spread_butter_spread_fat_thin | 5 | oatcakes_spread_butter_spread_fat_thin | 1.75 | -65% | 25% of weight on a slice |
| Fat that is spread (medium amount) onto oatcakes: ticked spreadable butter, low fat | oatcakes_spread_butter_spread_lowfat_med | 7 | oatcakes_spread_butter_spread_lowfat_med | 2.5 | -64% | 25% of weight on a slice |
| Fat that is spread (thick amount) onto oatcakes: ticked spreadable butter, low fat | oatcakes_spread_butter_spread_lowfat_thick | 10 | oatcakes_spread_butter_spread_lowfat_thick | 3 | -70% | 25% of weight on a slice |
| Fat that is spread (thin amount) onto oatcakes: ticked spreadable butter, low fat | oatcakes_spread_butter_spread_lowfat_thin | 5 | oatcakes_spread_butter_spread_lowfat_thin | 1.75 | -65% | 25% of weight on a slice |
| Fat that is spread (medium amount) onto oatcakes: ticked dairy spread which is also cholesterol lowering e.g Benecol Buttery | oatcakes_spread_dairy_chol_med | 5 | oatcakes_spread_dairy_chol_med | 1.75 | -65% | 25% of weight on a slice |
| Fat that is spread (thick amount) onto oatcakes: ticked dairy spread which is also cholesterol lowering e.g Benecol Buttery | oatcakes_spread_dairy_chol_thick | 7 | oatcakes_spread_dairy_chol_thick | 2.5 | -64% | 25% of weight on a slice |
| Fat that is spread (thin amount) onto oatcakes: ticked dairy spread which is also cholesterol lowering e.g Benecol Buttery | oatcakes_spread_dairy_chol_thin | 3 | oatcakes_spread_dairy_chol_thin | 1.25 | -58% | 25% of weight on a slice |
| Fat that is spread (medium amount) onto oatcakes: ticked dairy spread but not specified amount of % fat | oatcakes_spread_dairy_dunno_med | 5 | oatcakes_spread_dairy_dunno_med | 1.75 | -65% | 25% of weight on a slice; food composition as low fat and reduced fat |
| Fat that is spread (thick amount) onto oatcakes: ticked dairy spread but not specified amount of % fat | oatcakes_spread_dairy_dunno_thick | 7 | oatcakes_spread_dairy_dunno_thick | 2.5 | -64% | 25% of weight on a slice; food composition as low fat and reduced fat |
| Fat that is spread (thin amount) onto oatcakes: ticked dairy spread but not specified amount of % fat | oatcakes_spread_dairy_dunno_thin | 3 | oatcakes_spread_dairy_dunno_thin | 1.25 | -58% | 25% of weight on a slice; food composition as low fat and reduced fat |
| Fat that is spread (medium amount) onto oatcakes: ticked dairy spread with normal amount of % fat | oatcakes_spread_dairy_fat_med | 5 | oatcakes_spread_dairy_fat_med | 1.75 | -65% | 25% of weight on a slice; normal fat is taken as reduced fat (up to 62% fat) |
| Fat that is spread (thick amount) onto oatcakes: ticked dairy spread with normal amount of % fat | oatcakes_spread_dairy_fat_thick | 7 | oatcakes_spread_dairy_fat_thick | 2.5 | -64% | 25% of weight on a slice; normal fat is taken as reduced fat (up to 62% fat) |
| Fat that is spread (thin amount) onto oatcakes: ticked dairy spread with normal amount of % fat | oatcakes_spread_dairy_fat_thin | 3 | oatcakes_spread_dairy_fat_thin | 1.25 | -58% | 25% of weight on a slice; normal fat is taken as reduced fat (up to 62% fat) |
| Fat that is spread (medium amount) onto oatcakes: ticked dairy spread, low fat | oatcakes_spread_dairy_lowfat_med | 5 | oatcakes_spread_dairy_lowfat_med | 1.75 | -65% | 25% of weight on a slice |
| Fat that is spread (thick amount) onto oatcakes: ticked dairy spread, low fat | oatcakes_spread_dairy_lowfat_thick | 7 | oatcakes_spread_dairy_lowfat_thick | 2.5 | -64% | 25% of weight on a slice |
| Fat that is spread (thin amount) onto oatcakes: ticked dairy spread, low fat | oatcakes_spread_dairy_lowfat_thin | 3 | oatcakes_spread_dairy_lowfat_thin | 1.25 | -58% | 25% of weight on a slice |
| Fat that is spread (medium amount) onto oatcakes: ticked dairy spread, very low fat | oatcakes_spread_dairy_vlowfat_med | 5 | oatcakes_spread_dairy_vlowfat_med | 1.75 | -65% | 25% of weight on a slice; food composition as low fat spread |
| Fat that is spread (thick amount) onto oatcakes: ticked dairy spread, very low fat | oatcakes_spread_dairy_vlowfat_thick | 7 | oatcakes_spread_dairy_vlowfat_thick | 2.5 | -64% | 25% of weight on a slice; food composition as low fat spread |
| Fat that is spread (thin amount) onto oatcakes: ticked dairy spread, very low fat | oatcakes_spread_dairy_vlowfat_thin | 3 | oatcakes_spread_dairy_vlowfat_thin | 1.25 | -58% | 25% of weight on a slice; food composition as low fat spread |
| Fat that is spread (medium amount) onto oatcakes: not specified type of spread but ticked cholesterol lowering e.g Benecol, Flora pro active | oatcakes_spread_dunno_chol_med | 5 | oatcakes_spread_dunno_chol_med | 1.75 | -65% | 25% of weight on a slice |
| Fat that is spread (thick amount) onto oatcakes: not specified type of spread but ticked cholesterol lowering e.g Benecol, Flora pro active | oatcakes_spread_dunno_chol_thick | 7 | oatcakes_spread_dunno_chol_thick | 2.5 | -64% | 25% of weight on a slice |
| Fat that is spread (thin amount) onto oatcakes: not specified type of spread but ticked cholesterol lowering e.g Benecol, Flora pro active | oatcakes_spread_dunno_chol_thin | 3 | oatcakes_spread_dunno_chol_thin | 1.25 | -58% | 25% of weight on a slice |
| Fat that is spread (medium amount) onto oatcakes: not specified type of spread or amount of % fat | oatcakes_spread_dunno_dunno_med | 5 | oatcakes_spread_dunno_dunno_med | 1.75 | -65% | 25% of weight on a slice |
| Fat that is spread (thick amount) onto oatcakes: not specified type of spread or amount of % fat | oatcakes_spread_dunno_dunno_thick | 7 | oatcakes_spread_dunno_dunno_thick | 2.5 | -64% | 25% of weight on a slice |
| Fat that is spread (thin amount) onto oatcakes: not specified type of spread or amount of % fat | oatcakes_spread_dunno_dunno_thin | 3 | oatcakes_spread_dunno_dunno_thin | 1.25 | -58% | 25% of weight on a slice |
| Fat that is spread (medium amount) onto oatcakes: not specified type of spread but ticked normal amount % fat | oatcakes_spread_dunno_fat_med | 5 | oatcakes_spread_dunno_fat_med | 1.75 | -65% | 25% of weight on a slice; normal fat is taken as reduced fat (up to 62% fat) |
| Fat that is spread (thick amount) onto oatcakes: not specified type of spread but ticked normal amount % fat | oatcakes_spread_dunno_fat_thick | 7 | oatcakes_spread_dunno_fat_thick | 2.5 | -64% | 25% of weight on a slice; normal fat is taken as reduced fat (up to 62% fat) |
| Fat that is spread (thin amount) onto oatcakes: not specified type of spread but ticked normal amount % fat | oatcakes_spread_dunno_fat_thin | 3 | oatcakes_spread_dunno_fat_thin | 1.25 | -58% | 25% of weight on a slice; normal fat is taken as reduced fat (up to 62% fat) |
| Fat that is spread (medium amount) onto oatcakes: not specified type of spread but ticked low fat | oatcakes_spread_dunno_lowfat_med | 5 | oatcakes_spread_dunno_lowfat_med | 1.75 | -65% | 25% of weight on a slice |
| Fat that is spread (thick amount) onto oatcakes: not specified type of spread but ticked low fat | oatcakes_spread_dunno_lowfat_thick | 7 | oatcakes_spread_dunno_lowfat_thick | 2.5 | -64% | 25% of weight on a slice |
| Fat that is spread (thin amount) onto oatcakes: not specified type of spread but ticked low fat | oatcakes_spread_dunno_lowfat_thin | 3 | oatcakes_spread_dunno_lowfat_thin | 1.25 | -58% | 25% of weight on a slice |
| Fat that is spread (medium amount) onto oatcakes: not specified type of spread but ticked very low fat | oatcakes_spread_dunno_vlowfat_med | 5 | oatcakes_spread_dunno_vlowfat_med | 1.75 | -65% | 25% of weight on a slice; food composition as low fat spread |
| Fat that is spread (thick amount) onto oatcakes: not specified type of spread but ticked very low fat | oatcakes_spread_dunno_vlowfat_thick | 7 | oatcakes_spread_dunno_vlowfat_thick | 2.5 | -64% | 25% of weight on a slice; food composition as low fat spread |
| Fat that is spread (thin amount) onto oatcakes: not specified type of spread but ticked very low fat | oatcakes_spread_dunno_vlowfat_thin | 3 | oatcakes_spread_dunno_vlowfat_thin | 1.25 | -58% | 25% of weight on a slice; food composition as low fat spread |
| Fat that is spread (medium amount) onto oatcakes: ticked hard margarine (hard block margarine in wrapper) | oatcakes_spread_hardmarg_med | 7 | oatcakes_spread_hardmarg_med | 2.5 | -64% | 25% of weight on a slice; spread weight as butter |
| Fat that is spread (thick amount) onto oatcakes: ticked hard margarine (hard block margarine in wrapper) | oatcakes_spread_hardmarg_thick | 10 | oatcakes_spread_hardmarg_thick | 3 | -70% | 25% of weight on a slice; spread weight as butter |
| Fat that is spread (thin amount) onto oatcakes: ticked hard margarine (hard block margarine in wrapper) | oatcakes_spread_hardmarg_thin | 5 | oatcakes_spread_hardmarg_thin | 1.75 | -65% | 25% of weight on a slice; spread weight as butter |
| Fat that is spread (medium amount) onto oatcakes: ticked cholesterol lowering olive spread e.g Benecol/Flora pro active olive spread | oatcakes_spread_olive_chol_med | 5 | oatcakes_spread_olive_chol_med | 1.75 | -65% | 25% of weight on a slice |
| Fat that is spread (thick amount) onto oatcakes: ticked cholesterol lowering olive spread e.g Benecol/Flora pro active olive spread | oatcakes_spread_olive_chol_thick | 7 | oatcakes_spread_olive_chol_thick | 2.5 | -64% | 25% of weight on a slice |
| Fat that is spread (thin amount) onto oatcakes: ticked cholesterol lowering olive spread e.g Benecol/Flora pro active olive spread | oatcakes_spread_olive_chol_thin | 3 | oatcakes_spread_olive_chol_thin | 1.25 | -58% | 25% of weight on a slice |
| Fat that is spread (medium amount) onto oatcakes: ticked olive spread but not specified amount of fat | oatcakes_spread_olive_dunno_med | 5 | oatcakes_spread_olive_dunno_med | 1.75 | -65% | 25% of weight on a slice |
| Fat that is spread (thick amount) onto oatcakes: ticked olive spread but not specified amount of fat | oatcakes_spread_olive_dunno_thick | 7 | oatcakes_spread_olive_dunno_thick | 2.5 | -64% | 25% of weight on a slice |
| Fat that is spread (thin amount) onto oatcakes: ticked olive spread but not specified amount of fat | oatcakes_spread_olive_dunno_thin | 3 | oatcakes_spread_olive_dunno_thin | 1.25 | -58% | 25% of weight on a slice |
| Fat that is spread (medium amount) onto oatcakes: ticked olive spread with normal amount of % fat | oatcakes_spread_olive_fat_med | 5 | oatcakes_spread_olive_fat_med | 1.75 | -65% | 25% of weight on a slice; normal fat is taken as reduced fat (up to 62% fat) |
| Fat that is spread (thick amount) onto oatcakes: ticked olive spread with normal amount of % fat | oatcakes_spread_olive_fat_thick | 7 | oatcakes_spread_olive_fat_thick | 2.5 | -64% | 25% of weight on a slice; normal fat is taken as reduced fat (up to 62% fat) |
| Fat that is spread (thin amount) onto oatcakes: ticked olive spread with normal amount of % fat | oatcakes_spread_olive_fat_thin | 3 | oatcakes_spread_olive_fat_thin | 1.25 | -58% | 25% of weight on a slice; normal fat is taken as reduced fat (up to 62% fat) |
| Fat that is spread (medium amount) onto oatcakes: ticked olive spread, low fat | oatcakes_spread_olive_lowfat_med | 5 | oatcakes_spread_olive_lowfat_med | 1.75 | -65% | 25% of weight on a slice |
| Fat that is spread (thick amount) onto oatcakes: ticked olive spread, low fat | oatcakes_spread_olive_lowfat_thick | 7 | oatcakes_spread_olive_lowfat_thick | 2.5 | -64% | 25% of weight on a slice |
| Fat that is spread (thin amount) onto oatcakes: ticked olive spread, low fat | oatcakes_spread_olive_lowfat_thin | 3 | oatcakes_spread_olive_lowfat_thin | 1.25 | -58% | 25% of weight on a slice |
| Fat that is spread (medium amount) onto oatcakes: ticked olive spread, very low fat | oatcakes_spread_olive_vlowfat_med | 5 | oatcakes_spread_olive_vlowfat_med | 1.75 | -65% | 25% of weight on a slice; food composition as low fat spread |
| Fat that is spread (thick amount) onto oatcakes: ticked olive spread, very low fat | oatcakes_spread_olive_vlowfat_thick | 7 | oatcakes_spread_olive_vlowfat_thick | 2.5 | -64% | 25% of weight on a slice; food composition as low fat spread |
| Fat that is spread (thin amount) onto oatcakes: ticked olive spread, very low fat | oatcakes_spread_olive_vlowfat_thin | 3 | oatcakes_spread_olive_vlowfat_thin | 1.25 | -58% | 25% of weight on a slice; food composition as low fat spread |
| Fat that is spread (medium amount) onto oatcakes: ticked other spread e.g ghee | oatcakes_spread_other_med | 5 | oatcakes_spread_other_med | 1.75 | -65% | 25% of weight on a slice |
| Fat that is spread (thick amount) onto oatcakes: ticked other spread e.g ghee | oatcakes_spread_other_thick | 7 | oatcakes_spread_other_thick | 2.5 | -64% | 25% of weight on a slice |
| Fat that is spread (thin amount) onto oatcakes: ticked other spread e.g ghee | oatcakes_spread_other_thin | 3 | oatcakes_spread_other_thin | 1.25 | -58% | 25% of weight on a slice |
| Fat that is spread (medium amount) onto oatcakes: ticked polyunsaturated margarine (e.g Flora) and also cholesterol lowering | oatcakes_spread_polymarg_chol_med | 5 | oatcakes_spread_polymarg_chol_med | 1.75 | -65% | 25% of weight on a slice |
| Fat that is spread (thick amount) onto oatcakes: ticked polyunsaturated margarine (e.g Flora) and also cholesterol lowering | oatcakes_spread_polymarg_chol_thick | 7 | oatcakes_spread_polymarg_chol_thick | 2.5 | -64% | 25% of weight on a slice |
| Fat that is spread (thin amount) onto oatcakes: ticked polyunsaturated margarine (e.g Flora) and also cholesterol lowering | oatcakes_spread_polymarg_chol_thin | 3 | oatcakes_spread_polymarg_chol_thin | 1.25 | -58% | 25% of weight on a slice |
| Fat that is spread (medium amount) onto oatcakes: ticked polyunsaturated margarine (e.g Flora) but not specified amount of fat | oatcakes_spread_polymarg_dunno_med | 5 | oatcakes_spread_polymarg_dunno_med | 1.75 | -65% | 25% of weight on a slice |
| Fat that is spread (thick amount) onto oatcakes: ticked polyunsaturated margarine (e.g Flora) but not specified amount of fat | oatcakes_spread_polymarg_dunno_thick | 7 | oatcakes_spread_polymarg_dunno_thick | 2.5 | -64% | 25% of weight on a slice |
| Fat that is spread (thin amount) onto oatcakes: ticked polyunsaturated margarine (e.g Flora) but not specified amount of fat | oatcakes_spread_polymarg_dunno_thin | 3 | oatcakes_spread_polymarg_dunno_thin | 1.25 | -58% | 25% of weight on a slice |
| Fat that is spread (medium amount) onto oatcakes: ticked polyunsaturated margarine (e.g Flora), normal amount of % fat | oatcakes_spread_polymarg_fat_med | 5 | oatcakes_spread_polymarg_fat_med | 1.75 | -65% | 25% of weight on a slice; normal fat is taken as reduced fat (up to 62% fat) |
| Fat that is spread (thick amount) onto oatcakes: ticked polyunsaturated margarine (e.g Flora), normal amount of % fat | oatcakes_spread_polymarg_fat_thick | 7 | oatcakes_spread_polymarg_fat_thick | 2.5 | -64% | 25% of weight on a slice; normal fat is taken as reduced fat (up to 62% fat) |
| Fat that is spread (thin amount) onto oatcakes: ticked polyunsaturated margarine (e.g Flora), normal amount of % fat | oatcakes_spread_polymarg_fat_thin | 3 | oatcakes_spread_polymarg_fat_thin | 1.25 | -58% | 25% of weight on a slice; normal fat is taken as reduced fat (up to 62% fat) |
| Fat that is spread (medium amount) onto oatcakes: ticked polyunsaturated margarine (e.g Flora), low fat | oatcakes_spread_polymarg_lowfat_med | 5 | oatcakes_spread_polymarg_lowfat_med | 1.75 | -65% | 25% of weight on a slice |
| Fat that is spread (thick amount) onto oatcakes: ticked polyunsaturated margarine (e.g Flora), low fat | oatcakes_spread_polymarg_lowfat_thick | 7 | oatcakes_spread_polymarg_lowfat_thick | 2.5 | -64% | 25% of weight on a slice |
| Fat that is spread (thin amount) onto oatcakes: ticked polyunsaturated margarine (e.g Flora), low fat | oatcakes_spread_polymarg_lowfat_thin | 3 | oatcakes_spread_polymarg_lowfat_thin | 1.25 | -58% | 25% of weight on a slice |
| Fat that is spread (medium amount) onto oatcakes: ticked polyunsaturated margarine (e.g Flora), very low fat | oatcakes_spread_polymarg_vlowfat_med | 5 | oatcakes_spread_polymarg_vlowfat_med | 1.75 | -65% | 25% of weight on a slice; food composition as low fat spread |
| Fat that is spread (thick amount) onto oatcakes: ticked polyunsaturated margarine (e.g Flora), very low fat | oatcakes_spread_polymarg_vlowfat_thick | 7 | oatcakes_spread_polymarg_vlowfat_thick | 2.5 | -64% | 25% of weight on a slice; food composition as low fat spread |
| Fat that is spread (thin amount) onto oatcakes: ticked polyunsaturated margarine (e.g Flora), very low fat | oatcakes_spread_polymarg_vlowfat_thin | 3 | oatcakes_spread_polymarg_vlowfat_thin | 1.25 | -58% | 25% of weight on a slice; food composition as low fat spread |
| Fat that is spread (medium amount) onto oatcakes: ticked soya/vegan/dairy free margarine e.g Pure, and also cholesterol lowering | oatcakes_spread_soya_chol_med | 5 | oatcakes_spread_soya_chol_med | 1.75 | -65% | 25% of weight on a slice |
| Fat that is spread (thick amount) onto oatcakes: ticked soya/vegan/dairy free margarine e.g Pure, and also cholesterol lowering | oatcakes_spread_soya_chol_thick | 7 | oatcakes_spread_soya_chol_thick | 2.5 | -64% | 25% of weight on a slice |
| Fat that is spread (thin amount) onto oatcakes: ticked soya/vegan/dairy free margarine e.g Pure, and also cholesterol lowering | oatcakes_spread_soya_chol_thin | 3 | oatcakes_spread_soya_chol_thin | 1.25 | -58% | 25% of weight on a slice |
| Fat that is spread (medium amount) onto oatcakes: ticked soya/vegan/dairy free margarine e.g Pure, and not specified amount of fat | oatcakes_spread_soya_dunno_med | 5 | oatcakes_spread_soya_dunno_med | 1.75 | -65% | 25% of weight on a slice |
| Fat that is spread (thick amount) onto oatcakes: ticked soya/vegan/dairy free margarine e.g Pure, and not specified amount of fat | oatcakes_spread_soya_dunno_thick | 7 | oatcakes_spread_soya_dunno_thick | 2.5 | -64% | 25% of weight on a slice |
| Fat that is spread (thin amount) onto oatcakes: ticked soya/vegan/dairy free margarine e.g Pure, and not specified amount of fat | oatcakes_spread_soya_dunno_thin | 3 | oatcakes_spread_soya_dunno_thin | 1.25 | -58% | 25% of weight on a slice |
| Fat that is spread (medium amount) onto oatcakes: ticked soya/vegan/dairy free margarine (e.g Pure), normal amount % fat | oatcakes_spread_soya_fat_med | 5 | oatcakes_spread_soya_fat_med | 1.75 | -65% | 25% of weight on a slice; normal fat is taken as reduced fat (up to 62% fat) |
| Fat that is spread (thick amount) onto oatcakes: ticked soya/vegan/dairy free margarine (e.g Pure), normal amount % fat | oatcakes_spread_soya_fat_thick | 7 | oatcakes_spread_soya_fat_thick | 2.5 | -64% | 25% of weight on a slice; normal fat is taken as reduced fat (up to 62% fat) |
| Fat that is spread (thin amount) onto oatcakes: ticked soya/vegan/dairy free margarine (e.g Pure), normal amount % fat | oatcakes_spread_soya_fat_thin | 3 | oatcakes_spread_soya_fat_thin | 1.25 | -58% | 25% of weight on a slice; normal fat is taken as reduced fat (up to 62% fat) |
| Fat that is spread (medium amount) onto oatcakes: ticked soya/vegan/dairy free margarine (e.g Pure), low fat | oatcakes_spread_soya_lowfat_med | 5 | oatcakes_spread_soya_lowfat_med | 1.75 | -65% | 25% of weight on a slice |
| Fat that is spread (thick amount) onto oatcakes: ticked soya/vegan/dairy free margarine (e.g Pure), low fat | oatcakes_spread_soya_lowfat_thick | 7 | oatcakes_spread_soya_lowfat_thick | 2.5 | -64% | 25% of weight on a slice |
| Fat that is spread (thin amount) onto oatcakes: ticked soya/vegan/dairy free margarine (e.g Pure), low fat | oatcakes_spread_soya_lowfat_thin | 3 | oatcakes_spread_soya_lowfat_thin | 1.25 | -58% | 25% of weight on a slice |
| Fat that is spread (medium amount) onto oatcakes: ticked soya/vegan/dairy free margarine (e.g Pure), very low fat | oatcakes_spread_soya_vlowfat_med | 5 | oatcakes_spread_soya_vlowfat_med | 1.75 | -65% | 25% of weight on a slice; food composition as low fat spread |
| Fat that is spread (thick amount) onto oatcakes: ticked soya/vegan/dairy free margarine (e.g Pure), very low fat | oatcakes_spread_soya_vlowfat_thick | 7 | oatcakes_spread_soya_vlowfat_thick | 2.5 | -64% | 25% of weight on a slice; food composition as low fat spread |
| Fat that is spread (thin amount) onto oatcakes: ticked soya/vegan/dairy free margarine (e.g Pure), very low fat | oatcakes_spread_soya_vlowfat_thin | 3 | oatcakes_spread_soya_vlowfat_thin | 1.25 | -58% | 25% of weight on a slice; food composition as low fat spread |
| Scotch pancake, blini, American style pancake made with cholesterol lowering milk | pancake_blini | 41 | pancake_blini_chol | 41 | 0% | Milk type taken into account in the updated version |
| Scotch pancake, blini, American style pancake made with milk (cow's) unspecified | pancake_blini | 41 | pancake_blini_dontknow | 41 | 0% | Milk type taken into account in the updated version |
| Scotch pancake, blini, American style pancake made with goat's or sheep's milk | pancake_blini | 41 | pancake_blini_goatsheep | 41 | 0% | Milk type taken into account in the updated version |
| Scotch pancake, blini, American style pancake made with other milk (e.g 1% milk, lactose free milk, almond milk) | pancake_blini | 41 | pancake_blini_other | 41 | 0% | Milk type taken into account in the updated version |
| Scotch pancake, blini, American style pancake made with powdered milk made up | pancake_blini | 41 | pancake_blini_powdered | 41 | 0% | Milk type taken into account in the updated version |
| Scotch pancake, blini, American style pancake made with rice, oat, almond, coconut milk | pancake_blini | 41 | pancake_blini_riceoatveg | 41 | 0% | Milk type taken into account in the updated version |
| Scotch pancake, blini, American style pancake made with semi skimmed milk | pancake_blini | 41 | pancake_blini_semi | 41 | 0% | Milk type taken into account in the updated version |
| Scotch pancake, blini, American style pancake made with skimmed milk | pancake_blini | 41 | pancake_blini_skimmed | 41 | 0% | Milk type taken into account in the updated version |
| Scotch pancake, blini, American style pancake made with soya milk with added calcium | pancake_blini | 41 | pancake_blini_soya_ca | 41 | 0% | Milk type taken into account in the updated version |
| Scotch pancake, blini, American style pancake made with soya milk with no added calcium | pancake_blini | 41 | pancake_blini_soya_noca | 41 | 0% | Milk type taken into account in the updated version |
| Scotch pancake, blini, American style pancake made with whole milk | pancake_blini | 41 | pancake_blini_whole | 41 | 0% | Milk type taken into account in the updated version |
| Pancake, crêpe made with cholesterol lowering milk | pancake_crepe | 110 | pancake_crepe_chol | 110 | 0% | Milk type taken into account in the updated version |
| Pancake, crêpe made with milk (cow's) unspecified | pancake_crepe | 110 | pancake_crepe_dontknow | 110 | 0% | Milk type taken into account in the updated version |

| Variable description | McCance and Widdowson's Food item | Portion size | Nutrient databank + other changes Food item | Portion size | Portion diff% | Description of differences with previous version |
|---|---|---|---|---|---|---|
| Pancake, crêpe made with goat's or sheep's milk | pancake_crepe | 110 | pancake_crepe_goatsheep | 110 | 0% | Milk type taken into account in the updated version |
| Pancake, crêpe made with other milk (e.g 1% milk, lactose free milk, almond milk) | pancake_crepe | 110 | pancake_crepe_other | 110 | 0% | Milk type taken into account in the updated version |
| Pancake, crêpe made with powdered milk made up | pancake_crepe | 110 | pancake_crepe_powdered | 110 | 0% | Milk type taken into account in the updated version |
| Pancake, crêpe made with rice, oat, almond, coconut milk | pancake_crepe | 110 | pancake_crepe_riceoatveg | 110 | 0% | Milk type taken into account in the updated version |
| Pancake, crêpe made with semi skimmed milk | pancake_crepe | 110 | pancake_crepe_semi | 110 | 0% | Milk type taken into account in the updated version |
| Pancake, crêpe made with skimmed milk | pancake_crepe | 110 | pancake_crepe_skimmed | 110 | 0% | Milk type taken into account in the updated version |
| Pancake, crêpe made with soya milk with added calcium | pancake_crepe | 110 | pancake_crepe_soya_ca | 110 | 0% | Milk type taken into account in the updated version |
| Pancake, crêpe made with soya milk with no added calcium | pancake_crepe | 110 | pancake_crepe_soya_noca | 110 | 0% | Milk type taken into account in the updated version |
| Pancake, crêpe made with whole milk | pancake_crepe | 110 | pancake_crepe_whole | 110 | 0% | Milk type taken into account in the updated version |
| Brown pasta | pasta_brown | 230 | pasta_brown | 230 | 0% | |
| Gluten free pasta (rice/millet) | | | pasta_gf | 230 | - | Gluten free version added to the updated version |
| White pasta | pasta_white | 230 | pasta_white | 230 | 0% | |
| Pesto | pesto | 26 | pesto | 26 | 0% | |
| Pizza | pizza | 150 | pizza | 150 | 0% | |
| Pizza | | | pizza_gf | 150 | - | Gluten free version added to the updated version (however no gluten free code available at the time so the non-gluten free code was used) |
| Peanut butter, chocolate/nut spread (e.g. Nutella) | pnutbutter_nutella | 16 | pnutbutter_nutella | 16 | 0% | |
| Potato boiled | potato_boil | 175 | potato_boil | 175 | 0% | |
| Knob of butter or margarine added to boiled potatoes | potato_boil_marg | 25 | potato_boil_marg | 25 | 0% | |
| Potatoes: fried, chips, wedges, roast | potato_fried | 180 | potato_fried | 180 | 0% | |
| Mashed potato with milk and fat: Fat specified as butter but not specified amount of % fat | potato_mashed | 60 | potato_mashed_butter_dunno | 180 | 200% | Fat used taken into account in the updated version + serving increased from 1 to 3 scoops to obtain a portion size similar to other potato items |
| Mashed potato with milk and fat: Fat specified as butter, normal amount of % fat | potato_mashed | 60 | potato_mashed_butter_fat | 180 | 200% | Fat used taken into account in the updated version + serving increased from 1 to 3 scoops to obtain a portion size similar to other potato items |
| Mashed potato with milk and fat: Fat specified as butter, low fat | potato_mashed | 60 | potato_mashed_butter_lowfat | 180 | 200% | Fat used taken into account in the updated version + serving increased from 1 to 3 scoops to obtain a portion size similar to other potato items |
| Mashed potato with milk and fat: Fat specified as spreadable butter with normal amount of % fat | potato_mashed | 60 | potato_mashed_butter_spread_fat | 180 | 200% | Fat used taken into account in the updated version + serving increased from 1 to 3 scoops to obtain a portion size similar to other potato items |
| Mashed potato with milk and fat: Fat specified as spreadable butter, low fat | potato_mashed | 60 | potato_mashed_butter_spread_lowfat | 180 | 200% | Fat used taken into account in the updated version + serving increased from 1 to 3 scoops to obtain a portion size similar to other potato items |
| Mashed potato with milk and fat: Fat specified as not known type of fat or spread | potato_mashed | 60 | potato_mashed_fat_dunno | 180 | 200% | Fat used taken into account in the updated version + serving increased from 1 to 3 scoops to obtain a portion size similar to other potato items |
| Mashed potato with milk and no fat added | potato_mashed | 60 | potato_mashed_fat_none | 180 | 200% | Fat used taken into account in the updated version + serving increased from 1 to 3 scoops to obtain a portion size similar to other potato items |
| Mashed potato with milk and fat: Fat specified as other type of fat or spread e.g ghee, dripping | potato_mashed | 60 | potato_mashed_fat_other | 180 | 200% | Fat used taken into account in the updated version + serving increased from 1 to 3 scoops to obtain a portion size similar to other potato items |
| Mashed potato with milk and fat: Fat specified as lard | potato_mashed | 60 | potato_mashed_lard | 180 | 200% | Fat used taken into account in the updated version + serving increased from 1 to 3 scoops to obtain a portion size similar to other potato items |
| Mashed potato with milk and fat: Fat specified as hard margarine (hard block margarine in wrapper) | potato_mashed | 60 | potato_mashed_marg_hard | 180 | 200% | Fat used taken into account in the updated version + serving increased from 1 to 3 scoops to obtain a portion size similar to other potato items |
| Mashed potato with milk and fat: Fat specified as polyunsaturated margarine (e.g Flora) and also cholesterol lowering | potato_mashed | 60 | potato_mashed_marg_poly_chol | 180 | 200% | Fat used taken into account in the updated version + serving increased from 1 to 3 scoops to obtain a portion size similar to other potato items |
| Mashed potato with milk and fat: Fat specified as polyunsaturated margarine (e.g Flora) but not specified amount of fat | potato_mashed | 60 | potato_mashed_marg_poly_dunno | 180 | 200% | Fat used taken into account in the updated version + serving increased from 1 to 3 scoops to obtain a portion size similar to other potato items |
| Mashed potato with milk and fat: Fat specified as polyunsaturated margarine (e.g Flora), normal amount of % fat | potato_mashed | 60 | potato_mashed_marg_poly_fat | 180 | 200% | Fat used taken into account in the updated version + serving increased from 1 to 3 scoops to obtain a portion size similar to other potato items |
| Mashed potato with milk and fat: Fat specified as polyunsaturated margarine (e.g Flora), low fat | potato_mashed | 60 | potato_mashed_marg_poly_lowfat | 180 | 200% | Fat used taken into account in the updated version + serving increased from 1 to 3 scoops to obtain a portion size similar to other potato items |
| Mashed potato with milk and fat: Fat specified as polyunsaturated margarine (e.g Flora), very low fat | potato_mashed | 60 | potato_mashed_marg_poly_vlowfat | 180 | 200% | Fat used taken into account in the updated version + serving increased from 1 to 3 scoops to obtain a portion size similar to other potato items |
| Mashed potato with milk and fat: Fat specified as soya/vegan/dairy free margarine e.g Pure, and also cholesterol lowering | potato_mashed | 60 | potato_mashed_marg_soya_chol | 180 | 200% | Fat used taken into account in the updated version + serving increased from 1 to 3 scoops to obtain a portion size similar to other potato items |
| Mashed potato with milk and fat: Fat specified as soya/vegan/dairy free margarine e.g Pure, and not specified amount of fat | potato_mashed | 60 | potato_mashed_marg_soya_dunno | 180 | 200% | Fat used taken into account in the updated version + serving increased from 1 to 3 scoops to obtain a portion size similar to other potato items |
| Mashed potato with milk and fat: Fat specified as soya/vegan/dairy free margarine (e.g Pure), normal amount % fat | potato_mashed | 60 | potato_mashed_marg_soya_fat | 180 | 200% | Fat used taken into account in the updated version + serving increased from 1 to 3 scoops to obtain a portion size similar to other potato items |
| Mashed potato with milk and fat: Fat specified as soya/vegan/dairy free margarine (e.g Pure), low fat | potato_mashed | 60 | potato_mashed_marg_soya_lowfat | 180 | 200% | Fat used taken into account in the updated version + serving increased from 1 to 3 scoops to obtain a portion size similar to other potato items |
| Mashed potato with milk and fat: Fat specified as soya/vegan/dairy free margarine (e.g Pure), very low fat | potato_mashed | 60 | potato_mashed_marg_soya_vlowfat | 180 | 200% | Fat used taken into account in the updated version + serving increased from 1 to 3 scoops to obtain a portion size similar to other potato items |
| Mashed potato with milk and fat: Fat specified as olive oil | potato_mashed | 60 | potato_mashed_oil_olive | 180 | 200% | Fat used taken into account in the updated version + serving increased from 1 to 3 scoops to obtain a portion size similar to other potato items |
| Mashed potato with milk and fat: Fat specified as other oil e.g corn, groundnut, rice bran oil | potato_mashed | 60 | potato_mashed_oil_other | 180 | 200% | Fat used taken into account in the updated version + serving increased from 1 to 3 scoops to obtain a portion size similar to other potato items |
| Mashed potato with milk and fat: Fat specified as rapeseed oil | potato_mashed | 60 | potato_mashed_oil_rapeseed | 180 | 200% | Fat used taken into account in the updated version + serving increased from 1 to 3 scoops to obtain a portion size similar to other potato items |
| Mashed potato with milk and fat: Fat specified as sunflower oil | potato_mashed | 60 | potato_mashed_oil_sunflower | 180 | 200% | Fat used taken into account in the updated version + serving increased from 1 to 3 scoops to obtain a portion size similar to other potato items |
| Mashed potato with milk and fat: Fat specified as vegetable oil | potato_mashed | 60 | potato_mashed_oil_veg | 180 | 200% | Fat used taken into account in the updated version + serving increased from 1 to 3 scoops to obtain a portion size similar to other potato items |
| Mashed potato with milk and fat: Fat specified as dairy spread which is also cholesterol lowering e.g Benecol Buttery | potato_mashed | 60 | potato_mashed_spread_dairy_chol | 180 | 200% | Fat used taken into account in the updated version + serving increased from 1 to 3 scoops to obtain a portion size similar to other potato items |
| Mashed potato with milk and fat: Fat specified as dairy spread but not specified amount of % fat | potato_mashed | 60 | potato_mashed_spread_dairy_dunno | 180 | 200% | Fat used taken into account in the updated version + serving increased from 1 to 3 scoops to obtain a portion size similar to other potato items |
| Mashed potato with milk and fat: Fat specified as dairy spread with normal amount of % fat | potato_mashed | 60 | potato_mashed_spread_dairy_fat | 180 | 200% | Fat used taken into account in the updated version + serving increased from 1 to 3 scoops to obtain a portion size similar to other potato items |
| Mashed potato with milk and fat: Fat specified as dairy spread, low fat | potato_mashed | 60 | potato_mashed_spread_dairy_lowfat | 180 | 200% | Fat used taken into account in the updated version + serving increased from 1 to 3 scoops to obtain a portion size similar to other potato items |
| Mashed potato with milk and fat: Fat specified as dairy spread, very low fat | potato_mashed | 60 | potato_mashed_spread_dairy_vlowfat | 180 | 200% | Fat used taken into account in the updated version + serving increased from 1 to 3 scoops to obtain a portion size similar to other potato items |
| Mashed potato with milk and fat: Fat specified as not known type of spread but ticked cholesterol lowering e.g Benecol, Flora pro active | potato_mashed | 60 | potato_mashed_spread_dunno_chol | 180 | 200% | Fat used taken into account in the updated version + serving increased from 1 to 3 scoops to obtain a portion size similar to other potato items |
| Mashed potato with milk and fat: Fat specified as not known type of spread or amount of % fat | potato_mashed | 60 | potato_mashed_spread_dunno_dunno | 180 | 200% | Fat used taken into account in the updated version + serving increased from 1 to 3 scoops to obtain a portion size similar to other potato items |
| Mashed potato with milk and fat: Fat specified as not known type of spread but ticked normal amount % fat | potato_mashed | 60 | potato_mashed_spread_dunno_fat | 180 | 200% | Fat used taken into account in the updated version + serving increased from 1 to 3 scoops to obtain a portion size similar to other potato items |
| Mashed potato with milk and fat: Fat specified as not known type of spread but ticked low fat | potato_mashed | 60 | potato_mashed_spread_dunno_lowfat | 180 | 200% | Fat used taken into account in the updated version + serving increased from 1 to 3 scoops to obtain a portion size similar to other potato items |
| Mashed potato with milk and fat: Fat specified as not known type of spread but ticked very low fat | potato_mashed | 60 | potato_mashed_spread_dunno_vlowfat | 180 | 200% | Fat used taken into account in the updated version + serving increased from 1 to 3 scoops to obtain a portion size similar to other potato items |
| Mashed potato with milk and fat: Fat specified as cholesterol lowering olive spread e.g Benecol/Flora pro active olive spread | potato_mashed | 60 | potato_mashed_spread_olive_chol | 180 | 200% | Fat used taken into account in the updated version + serving increased from 1 to 3 scoops to obtain a portion size similar to other potato items |
| Mashed potato with milk and fat: Fat specified as olive spread but not specified amount of fat | potato_mashed | 60 | potato_mashed_spread_olive_dunno | 180 | 200% | Fat used taken into account in the updated version + serving increased from 1 to 3 scoops to obtain a portion size similar to other potato items |
| Mashed potato with milk and fat: Fat specified as olive spread with normal amount of % fat | potato_mashed | 60 | potato_mashed_spread_olive_fat | 180 | 200% | Fat used taken into account in the updated version + serving increased from 1 to 3 scoops to obtain a portion size similar to other potato items |
| Mashed potato with milk and fat: Fat specified as olive spread, low fat | potato_mashed | 60 | potato_mashed_spread_olive_lowfat | 180 | 200% | Fat used taken into account in the updated version + serving increased from 1 to 3 scoops to obtain a portion size similar to other potato items |
| Mashed potato with milk and fat: Fat specified as olive spread, very low fat | potato_mashed | 60 | potato_mashed_spread_olive_vlowfat | 180 | 200% | Fat used taken into account in the updated version + serving increased from 1 to 3 scoops to obtain a portion size similar to other potato items |
| Chicken or turkey, skin removed, in breadcrumbs or deep fried e.g. nuggets, KFC | poultry_friedbcrumb_noskin | 100 | poultry_friedbcrumb_noskin | 100 | 0% | |
| Chicken or turkey, skin left on, in breadcrumbs or deep fried e.g. nuggets, KFC | poultry_friedbcrumb_withskin | 100 | poultry_friedbcrumb_withskin | 100 | 0% | Removed skin from mapping; unlikely to contain skin due to preparation method |
| Chicken or turkey, skin removed e.g. roast, drumsticks, curry | poultry_noskin | 130 | poultry_noskin | 130 | 0% | |
| Chicken or turkey, skin left on e.g. roast, drumsticks, curry | poultry_withskin | 130 | poultry_withskin | 130 | 0% | |
| Brown rice | rice_brown | 150 | rice_brown | 150 | 0% | |
| White rice | rice_white | 150 | rice_white | 150 | 0% | |
| Salad dressing e.g french dressing | salad_dressing | 15 | salad_dressing | 15 | 0% | |
| Brown sauce/ BBQ sauce | sauce_brown | 15 | sauce_brown | 15 | 0% | |
| Cheese sauce made with cholesterol lowering milk | sauce_cheese | 62 | sauce_cheese_chol | 62 | 0% | Milk type taken into account in the updated version |
| Cheese sauce made with milk (cow's) unspecified | sauce_cheese | 62 | sauce_cheese_dontknow | 62 | 0% | Milk type taken into account in the updated version |
| Cheese sauce made with goat's or sheep's milk | sauce_cheese | 62 | sauce_cheese_goatsheep | 62 | 0% | Milk type taken into account in the updated version |
| Cheese sauce made with other milk (e.g 1% milk, lactose free milk, almond milk) | sauce_cheese | 62 | sauce_cheese_other | 62 | 0% | Milk type taken into account in the updated version |
| Cheese sauce made with powdered milk made up | sauce_cheese | 62 | sauce_cheese_powdered | 62 | 0% | Milk type taken into account in the updated version |
| Cheese sauce made with rice, oat, almond, coconut milk | sauce_cheese | 62 | sauce_cheese_riceoatveg | 62 | 0% | Milk type taken into account in the updated version |
| Cheese sauce made with semi skimmed milk | sauce_cheese | 62 | sauce_cheese_semi | 62 | 0% | Milk type taken into account in the updated version |
| Cheese sauce made with skimmed milk | sauce_cheese | 62 | sauce_cheese_skimmed | 62 | 0% | Milk type taken into account in the updated version |
| Cheese sauce made with soya milk with added calcium | sauce_cheese | 62 | sauce_cheese_soya_ca | 62 | 0% | Milk type taken into account in the updated version |
| Cheese sauce made with soya milk with no added calcium | sauce_cheese | 62 | sauce_cheese_soya_noca | 62 | 0% | Milk type taken into account in the updated version |
| Cheese sauce made with whole milk | sauce_cheese | 62 | sauce_cheese_whole | 62 | 0% | Milk type taken into account in the updated version |
| Gravy | sauce_gravy | 50 | sauce_gravy | 50 | 0% | |
| Tomato ketchup | sauce_ketchup | 30 | sauce_ketchup | 30 | 0% | |
| Tomato based sauce e.g pasta sauce | sauce_tomato | 90 | sauce_tomato | 90 | 0% | |
| White or cream sauce e.g bechamel made with cholesterol lowering milk | sauce_white | 62 | sauce_white_chol | 62 | 0% | Milk type taken into account in the updated version |
| White or cream sauce e.g bechamel made with milk (cow's) unspecified | sauce_white | 62 | sauce_white_dontknow | 62 | 0% | Milk type taken into account in the updated version |
| White or cream sauce e.g bechamel made with goat's or sheep's milk | sauce_white | 62 | sauce_white_goatsheep | 62 | 0% | Milk type taken into account in the updated version |
| White or cream sauce e.g bechamel made with other milk (e.g 1% milk, lactose free milk, almond milk) | sauce_white | 62 | sauce_white_other | 62 | 0% | Milk type taken into account in the updated version |
| White or cream sauce e.g bechamel made with powdered milk made up | sauce_white | 62 | sauce_white_powdered | 62 | 0% | Milk type taken into account in the updated version |
| White or cream sauce e.g bechamel made with rice, oat, almond, coconut milk | sauce_white | 62 | sauce_white_riceoatveg | 62 | 0% | Milk type taken into account in the updated version |
| White or cream sauce e.g bechamel made with semi skimmed milk | sauce_white | 62 | sauce_white_semi | 62 | 0% | Milk type taken into account in the updated version |
| White or cream sauce e.g bechamel made with skimmed milk | sauce_white | 62 | sauce_white_skimmed | 62 | 0% | Milk type taken into account in the updated version |
| White or cream sauce e.g bechamel made with soya milk with added calcium | sauce_white | 62 | sauce_white_soya_ca | 62 | 0% | Milk type taken into account in the updated version |
| White or cream sauce e.g bechamel made with soya milk with no added calcium | sauce_white | 62 | sauce_white_soya_noca | 62 | 0% | Milk type taken into account in the updated version |
| White or cream sauce e.g bechamel made with whole milk | sauce_white | 62 | sauce_white_whole | 62 | 0% | Milk type taken into account in the updated version |
| Scone, plain, fruit, cheese | scone | 48 | scone | 48 | 0% | |
| Gluten free scone | | | scone_gf | 48 | - | Gluten free version added to the updated version (however no gluten free code available at the time so the non-gluten free code was used) |
| Single crust pie/flan e.g. quiche | single_crust | 30 | single_crust | 30 | 0% | |
| Milk/yogurt based smoothie | smoothie_dairy | 250 | smoothie_dairy | 260 | 4% | Slight increase in portion size, to be consistent with portion size of fruit smoothie |
| Fruit based smoothie | smoothie_fruit | 260 | smoothie_fruit | 260 | 0% | |
| Cheesy biscuits e.g Mini Cheddars, Tuc | snack_cheesybis | 40 | snack_cheesybis | 40 | 0% | |
| Crisps e.g. Walkers, Sensations, Doritos, Hula Hoops | snack_crisps | 40 | snack_crisps | 40 | 0% | |
| Olives | snack_olives | 50 | snack_olives | 50 | 0% | |
| Pot noodle style snack | snackpot | 280 | snackpot | 280 | 0% | |
| Salted/roasted nuts e.g. almonds, cashews, pistachios | snack_saltednuts | 40 | snack_saltednuts | 40 | 0% | |
| Peanuts, roasted/salted | snack_saltedpeanuts | 40 | snack_saltedpeanuts | 40 | 0% | |
| Savoury crispbread/corn cake snacks e.g. Snack-a-Jack, flavoured Ryvita snack size | snack_savourybis | 40 | snack_savourybis | 40 | 0% | |
| Seeds e.g. sunflower, pumpkin, linseeds | snack_seeds | 14 | snack_seeds | 14 | 0% | |
| Other savoury snack e.g bombay mix, monster munch, pretzel, popcorn | snack_svyother | 40 | snack_svyother | 40 | 0% | Updated mapping based on free text entered by study participants; nuts removed from mapping |
| Other sweet snack/bar e.g Go Ahead yogurt breaks, sweet popcorn | snack_swtother | 40 | snack_swtother | 40 | 0% | Updated mapping based on free text entered by study participants; croissant removed from mapping |
| Unsalted nuts e.g. almonds, cashews, walnuts | snack_unsaltednuts | 40 | snack_unsaltednuts | 40 | 0% | |
| Peanuts, unsalted e.g monkey nuts | snack_unsaltedpeanuts | 40 | snack_unsaltedpeanuts | 40 | 0% | |

| | McCance and Widdowson's | | Nutrient databank + other changes | | | |
|---|---|---|---|---|---|---|
| **Variable description** | **Food item** | **Portion size** | **Food item** | **Portion size** | **Portion diff%** | **Description of differences with previous version** |
| Carton, pouch, canned soup with fish/ seafood | soup_canned_fish | 220 | soup_canned_fish | 220 | 0% | |
| Carton, pouch, canned soup with meat/ poultry e.g. ham, chicken | soup_canned_meat | 220 | soup_canned_meat | 220 | 0% | |
| Carton, pouch, canned soup, other e.g pea and ham | soup_canned_other | 220 | soup_canned_other | 220 | 0% | |
| Carton, pouch, canned soup with pasta e.g. noodles | soup_canned_pasta | 220 | soup_canned_pasta | 220 | 0% | |
| Carton, pouch, canned soup with peas/ beans/ lentils | soup_canned_pulse | 220 | soup_canned_pulse | 220 | 0% | |
| Carton, pouch, canned soup but not specified the type | soup_canned_unanswered | 220 | soup_canned_unanswered | 220 | 0% | |
| Carton, pouch, canned soup with vegetables e.g. potato, tomato | soup_canned_veg | 220 | soup_canned_veg | 220 | 0% | |
| Homemade soup with fish/ seafood | soup_homemade_fish | 220 | soup_homemade_fish | 220 | 0% | |
| Homemade soup with meat/ poultry e.g. ham, chicken | soup_homemade_meat | 220 | soup_homemade_meat | 220 | 0% | |
| Homemade soup, other e.g pea and ham | soup_homemade_other | 220 | soup_homemade_other | 220 | 0% | |
| Homemade soup with pasta e.g. noodles | soup_homemade_pasta | 220 | soup_homemade_pasta | 220 | 0% | |
| Homemade soup with peas/ beans/ lentils | soup_homemade_pulse | 220 | soup_homemade_pulse | 220 | 0% | |
| Homemade soup but not specified the type | soup_homemade_unanswered | 220 | soup_homemade_unanswered | 220 | 0% | |
| Homemade soup with vegetables e.g. potato, tomato | soup_homemade_veg | 220 | soup_homemade_veg | 220 | 0% | |
| Dried/ powdered soup e.g. Cup-a-Soup | soup_powder | 200 | soup_powder | 200 | 0% | |
| Sponge puddings, plain or with chocolate or fruit sauce | spongepuds | 120 | spongepuds | 120 | 0% | |
| Sponge puddings, plain or with chocolate or fruit sauce | | | spongepuds_gf | 120 | - | Gluten free version added to the updated version (however no gluten free code available at the time so the non-gluten free code was used) |
| Other spread/ sauce/ dip e.g tartar, mint, sweet chilli, Indian curry sauce, mustard | spreadsauce_other | 20 | spreadsauce_other | 20 | 0% | |
| Sushi: sushi rice including seaweed/fish/meat/veg | sushi | 278 | sushi | 278 | 0% | |
| Sweets: hard and soft, e.g. peppermints, toffees, fudge, fruit flavoured sweets | sweets | 36 | sweets | 36 | 0% | |
| Sugar free sweets: hard and soft, e.g. peppermints, toffees, fudge, fruit flavoured sweets | sweets_diet | 18 | sweets_diet | 18 | 0% | |
| Standard tea, black | tea_black | 190 | tea_black | 190 | 0% | |
| Standard tea, black, decaffeinated | tea_black_decaf | 190 | tea_black_decaf | 190 | 0% | |
| Green tea | tea_green | 190 | tea_green | 190 | 0% | |
| Herbal, fruit tea | tea_herbal | 190 | tea_herbal | 190 | 0% | |
| Other tea e.g | tea_other | 190 | tea_other | 190 | 0% | Updated mapping based on free text entered by study participants, which mostly contained flavoured black teas |
| Rooibos/ Redbush tea | tea_rooibos | 190 | tea_rooibos | 190 | 0% | |
| Sugar added to tea | tea_sugar | 6 | tea_sugar | 6 | 0% | |
| Vegieburger or vegie sausage | vegalt_burger | 90 | vegalt_burger | 90 | 0% | |
| Other vegetarian alternative e.g nut roast, falafel | vegalt_other | 90 | vegalt_other | 90 | 0% | Updated mapping based on free text entered by study participants |
| Quorn sausage, burger, pieces | vegalt_quorn | 90 | vegalt_quorn | 90 | 0% | |
| Tofu / tempeh / TVP / soya mince | vegalt_tofu | 90 | vegalt_tofu | 90 | 0% | |
| Avocado | veg_avocado | 136 | veg_avocado | 136 | 0% | |
| Baked beans | veg_bakedbeans | 135 | veg_bakedbeans | 135 | 0% | |
| Beetroot | veg_beetroot | 48 | veg_beetroot | 48 | 0% | |
| Broad beans | veg_broadbeans | 70 | veg_broadbeans | 70 | 0% | |
| Broccoli | veg_broccoli | 80 | veg_broccoli | 80 | 0% | |
| Butternut squash | veg_butternut | 130 | veg_butternut | 130 | 0% | |
| Cabbage, kale | veg_cabbagekale | 90 | veg_cabbagekale | 90 | 0% | |
| Carrots | veg_carrots | 60 | veg_carrots | 60 | 0% | |
| Cauliflower | veg_cauli | 90 | veg_cauli | 90 | 0% | |
| Celery | veg_celery | 30 | veg_celery | 30 | 0% | |
| Courgettes cooked with added fat: Fat specified as butter but not specified amount of % fat | veg_courgette | 90 | veg_courgette_butter_dunno | 90 | 0% | Fat (2.5%) used for cooking taken into account in the updated version - 50% cooked with fat |
| Courgettes cooked with added fat: Fat specified as butter, normal amount of % fat | veg_courgette | 90 | veg_courgette_butter_fat | 90 | 0% | Fat (2.5%) used for cooking taken into account in the updated version - 50% cooked with fat |
| Courgettes cooked with added fat: Fat specified as butter, low fat | veg_courgette | 90 | veg_courgette_butter_lowfat | 90 | 0% | Fat (2.5%) used for cooking taken into account in the updated version - 50% cooked with fat |
| Courgettes cooked with added fat: Fat specified as spreadable butter with normal amount of % fat | veg_courgette | 90 | veg_courgette_butter_spread_fat | 90 | 0% | Fat (2.5%) used for cooking taken into account in the updated version - 50% cooked with fat |
| Courgettes cooked with added fat: Fat specified as spreadable butter, low fat | veg_courgette | 90 | veg_courgette_butter_spread_lowfat | 90 | 0% | Fat (2.5%) used for cooking taken into account in the updated version - 50% cooked with fat |
| Courgettes cooked with added fat: Fat specified as not known type of fat or spread | veg_courgette | 90 | veg_courgette_fat_dunno | 90 | 0% | Fat (2.5%) used for cooking taken into account in the updated version - 50% cooked with fat |
| Courgettes cooked with no added fat | veg_courgette | 90 | veg_courgette_fat_none | 90 | 0% | Fat (2.5%) used for cooking taken into account in the updated version - 50% cooked with fat |
| Courgettes cooked with added fat: Fat specified as other type of fat or spread e.g ghee, dripping | veg_courgette | 90 | veg_courgette_fat_other | 90 | 0% | Fat (2.5%) used for cooking taken into account in the updated version - 50% cooked with fat |
| Courgettes cooked with added fat: Fat specified as lard | veg_courgette | 90 | veg_courgette_lard | 90 | 0% | Fat (2.5%) used for cooking taken into account in the updated version - 50% cooked with fat |
| Courgettes cooked with added fat: Fat specified as hard margarine (hard block margarine in wrapper) | veg_courgette | 90 | veg_courgette_marg_hard | 90 | 0% | Fat (2.5%) used for cooking taken into account in the updated version - 50% cooked with fat |
| Courgettes cooked with added fat: Fat specified as polyunsaturated margarine (e.g Flora) and also cholesterol lowering | veg_courgette | 90 | veg_courgette_marg_poly_chol | 90 | 0% | Fat (2.5%) used for cooking taken into account in the updated version - 50% cooked with fat |
| Courgettes cooked with added fat: Fat specified as polyunsaturated margarine (e.g Flora) but not specified amount of fat | veg_courgette | 90 | veg_courgette_marg_poly_dunno | 90 | 0% | Fat (2.5%) used for cooking taken into account in the updated version - 50% cooked with fat |
| Courgettes cooked with added fat: Fat specified as polyunsaturated margarine (e.g Flora), normal amount of % fat | veg_courgette | 90 | veg_courgette_marg_poly_fat | 90 | 0% | Fat (2.5%) used for cooking taken into account in the updated version - 50% cooked with fat |
| Courgettes cooked with added fat: Fat specified as polyunsaturated margarine (e.g Flora), low fat | veg_courgette | 90 | veg_courgette_marg_poly_lowfat | 90 | 0% | Fat (2.5%) used for cooking taken into account in the updated version - 50% cooked with fat |
| Courgettes cooked with added fat: Fat specified as polyunsaturated margarine (e.g Flora), very low fat | veg_courgette | 90 | veg_courgette_marg_poly_vlowfat | 90 | 0% | Fat (2.5%) used for cooking taken into account in the updated version - 50% cooked with fat |
| Courgettes cooked with added fat: Fat specified as soya/vegan/dairy free margarine e.g Pure, and also cholesterol lowering | veg_courgette | 90 | veg_courgette_marg_soya_chol | 90 | 0% | Fat (2.5%) used for cooking taken into account in the updated version - 50% cooked with fat |
| Courgettes cooked with added fat: Fat specified as soya/vegan/dairy free margarine e.g Pure, and not specified amount of fat | veg_courgette | 90 | veg_courgette_marg_soya_dunno | 90 | 0% | Fat (2.5%) used for cooking taken into account in the updated version - 50% cooked with fat |
| Courgettes cooked with added fat: Fat specified as soya/vegan/dairy free margarine (e.g Pure), normal amount % fat | veg_courgette | 90 | veg_courgette_marg_soya_fat | 90 | 0% | Fat (2.5%) used for cooking taken into account in the updated version - 50% cooked with fat |
| Courgettes cooked with added fat: Fat specified as soya/vegan/dairy free margarine (e.g Pure), low fat | veg_courgette | 90 | veg_courgette_marg_soya_lowfat | 90 | 0% | Fat (2.5%) used for cooking taken into account in the updated version - 50% cooked with fat |
| Courgettes cooked with added fat: Fat specified as soya/vegan/dairy free margarine (e.g Pure), very low fat | veg_courgette | 90 | veg_courgette_marg_soya_vlowfat | 90 | 0% | Fat (2.5%) used for cooking taken into account in the updated version - 50% cooked with fat |
| Courgettes cooked with added fat: Fat specified as olive oil | veg_courgette | 90 | veg_courgette_oil_olive | 90 | 0% | Fat (2.5%) used for cooking taken into account in the updated version - 50% cooked with fat |
| Courgettes cooked with added fat: Fat specified as other oil e.g corn, groundnut, rice bran oil | veg_courgette | 90 | veg_courgette_oil_other | 90 | 0% | Fat (2.5%) used for cooking taken into account in the updated version - 50% cooked with fat |
| Courgettes cooked with added fat: Fat specified as rapeseed oil | veg_courgette | 90 | veg_courgette_oil_rapeseed | 90 | 0% | Fat (2.5%) used for cooking taken into account in the updated version - 50% cooked with fat |
| Courgettes cooked with added fat: Fat specified as sunflower oil | veg_courgette | 90 | veg_courgette_oil_sunflower | 90 | 0% | Fat (2.5%) used for cooking taken into account in the updated version - 50% cooked with fat |
| Courgettes cooked with added fat: Fat specified as vegetable oil | veg_courgette | 90 | veg_courgette_oil_veg | 90 | 0% | Fat (2.5%) used for cooking taken into account in the updated version - 50% cooked with fat |
| Courgettes cooked with added fat: Fat specified as dairy spread which is also cholesterol lowering e.g Benecol Buttery | veg_courgette | 90 | veg_courgette_spread_dairy_chol | 90 | 0% | Fat (2.5%) used for cooking taken into account in the updated version - 50% cooked with fat |
| Courgettes cooked with added fat: Fat specified as dairy spread but not specified amount of % fat | veg_courgette | 90 | veg_courgette_spread_dairy_dunno | 90 | 0% | Fat (2.5%) used for cooking taken into account in the updated version - 50% cooked with fat |
| Courgettes cooked with added fat: Fat specified as dairy spread with normal amount of % fat | veg_courgette | 90 | veg_courgette_spread_dairy_fat | 90 | 0% | Fat (2.5%) used for cooking taken into account in the updated version - 50% cooked with fat |
| Courgettes cooked with added fat: Fat specified as dairy spread, low fat | veg_courgette | 90 | veg_courgette_spread_dairy_lowfat | 90 | 0% | Fat (2.5%) used for cooking taken into account in the updated version - 50% cooked with fat |
| Courgettes cooked with added fat: Fat specified as dairy spread, very low fat | veg_courgette | 90 | veg_courgette_spread_dairy_vlowfat | 90 | 0% | Fat (2.5%) used for cooking taken into account in the updated version - 50% cooked with fat |
| Courgettes cooked with added fat: Fat specified as not known type of spread but ticked cholesterol lowering e.g Benecol, Flora pro active | veg_courgette | 90 | veg_courgette_spread_dunno_chol | 90 | 0% | Fat (2.5%) used for cooking taken into account in the updated version - 50% cooked with fat |
| Courgettes cooked with added fat: Fat specified as not known type of spread or amount of % fat | veg_courgette | 90 | veg_courgette_spread_dunno_dunno | 90 | 0% | Fat (2.5%) used for cooking taken into account in the updated version - 50% cooked with fat |
| Courgettes cooked with added fat: Fat specified as not known type of spread but ticked normal amount % fat | veg_courgette | 90 | veg_courgette_spread_dunno_fat | 90 | 0% | Fat (2.5%) used for cooking taken into account in the updated version - 50% cooked with fat |
| Courgettes cooked with added fat: Fat specified as not known type of spread but ticked low fat | veg_courgette | 90 | veg_courgette_spread_dunno_lowfat | 90 | 0% | Fat (2.5%) used for cooking taken into account in the updated version - 50% cooked with fat |
| Courgettes cooked with added fat: Fat specified as not known type of spread but ticked very low fat | veg_courgette | 90 | veg_courgette_spread_dunno_vlowfat | 90 | 0% | Fat (2.5%) used for cooking taken into account in the updated version - 50% cooked with fat |
| Courgettes cooked with added fat: Fat specified as cholesterol lowering olive spread e.g Benecol/Flora pro active olive spread | veg_courgette | 90 | veg_courgette_spread_olive_chol | 90 | 0% | Fat (2.5%) used for cooking taken into account in the updated version - 50% cooked with fat |
| Courgettes cooked with added fat: Fat specified as olive spread but not specified amount of fat | veg_courgette | 90 | veg_courgette_spread_olive_dunno | 90 | 0% | Fat (2.5%) used for cooking taken into account in the updated version - 50% cooked with fat |
| Courgettes cooked with added fat: Fat specified as olive spread with normal amount of % fat | veg_courgette | 90 | veg_courgette_spread_olive_fat | 90 | 0% | Fat (2.5%) used for cooking taken into account in the updated version - 50% cooked with fat |
| Courgettes cooked with added fat: Fat specified as olive spread, low fat | veg_courgette | 90 | veg_courgette_spread_olive_lowfat | 90 | 0% | Fat (2.5%) used for cooking taken into account in the updated version - 50% cooked with fat |
| Courgettes cooked with added fat: Fat specified as olive spread, very low fat | veg_courgette | 90 | veg_courgette_spread_olive_vlowfat | 90 | 0% | Fat (2.5%) used for cooking taken into account in the updated version - 50% cooked with fat |
| Cucumber | veg_cucumber | 60 | veg_cucumber | 60 | 0% | |
| Garlic | veg_garlic | 5 | veg_garlic | 5 | 0% | |
| Green beans, french beans, runner beans | veg_greenbeans | 70 | veg_greenbeans | 70 | 0% | |
| Leeks cooked with added fat: Fat specified as butter but not specified amount of % fat | veg_leek | 80 | veg_leek_butter_dunno | 80 | 0% | Fat (1.25%) used for cooking taken into account in the updated version - 25% cooked with fat |
| Leeks cooked with added fat: Fat specified as butter, normal amount of % fat | veg_leek | 80 | veg_leek_butter_fat | 80 | 0% | Fat (1.25%) used for cooking taken into account in the updated version - 25% cooked with fat |
| Leeks cooked with added fat: Fat specified as butter, low fat | veg_leek | 80 | veg_leek_butter_lowfat | 80 | 0% | Fat (1.25%) used for cooking taken into account in the updated version - 25% cooked with fat |
| Leeks cooked with added fat: Fat specified as spreadable butter with normal amount of % fat | veg_leek | 80 | veg_leek_butter_spread_fat | 80 | 0% | Fat (1.25%) used for cooking taken into account in the updated version - 25% cooked with fat |
| Leeks cooked with added fat: Fat specified as spreadable butter, low fat | veg_leek | 80 | veg_leek_butter_spread_lowfat | 80 | 0% | Fat (1.25%) used for cooking taken into account in the updated version - 25% cooked with fat |
| Leeks cooked with added fat: Fat specified as not known type of fat or spread | veg_leek | 80 | veg_leek_fat_dunno | 80 | 0% | Fat (1.25%) used for cooking taken into account in the updated version - 25% cooked with fat |
| Leeks cooked with no added fat | veg_leek | 80 | veg_leek_fat_none | 80 | 0% | Fat (1.25%) used for cooking taken into account in the updated version - 25% cooked with fat |
| Leeks cooked with added fat: Fat specified as other type of fat or spread e.g ghee, dripping | veg_leek | 80 | veg_leek_fat_other | 80 | 0% | Fat (1.25%) used for cooking taken into account in the updated version - 25% cooked with fat |
| Leeks cooked with added fat: Fat specified as lard | veg_leek | 80 | veg_leek_lard | 80 | 0% | Fat (1.25%) used for cooking taken into account in the updated version - 25% cooked with fat |
| Leeks cooked with added fat: Fat specified as hard margarine (hard block margarine in wrapper) | veg_leek | 80 | veg_leek_marg_hard | 80 | 0% | Fat (1.25%) used for cooking taken into account in the updated version - 25% cooked with fat |
| Leeks cooked with added fat: Fat specified as polyunsaturated margarine (e.g Flora) and also cholesterol lowering | veg_leek | 80 | veg_leek_marg_poly_chol | 80 | 0% | Fat (1.25%) used for cooking taken into account in the updated version - 25% cooked with fat |
| Leeks cooked with added fat: Fat specified as polyunsaturated margarine (e.g Flora) but not specified amount of fat | veg_leek | 80 | veg_leek_marg_poly_dunno | 80 | 0% | Fat (1.25%) used for cooking taken into account in the updated version - 25% cooked with fat |
| Leeks cooked with added fat: Fat specified as polyunsaturated margarine (e.g Flora), normal amount of % fat | veg_leek | 80 | veg_leek_marg_poly_fat | 80 | 0% | Fat (1.25%) used for cooking taken into account in the updated version - 25% cooked with fat |
| Leeks cooked with added fat: Fat specified as polyunsaturated margarine (e.g Flora), low fat | veg_leek | 80 | veg_leek_marg_poly_lowfat | 80 | 0% | Fat (1.25%) used for cooking taken into account in the updated version - 25% cooked with fat |
| Leeks cooked with added fat: Fat specified as polyunsaturated margarine (e.g Flora), very low fat | veg_leek | 80 | veg_leek_marg_poly_vlowfat | 80 | 0% | Fat (1.25%) used for cooking taken into account in the updated version - 25% cooked with fat |
| Leeks cooked with added fat: Fat specified as soya/vegan/dairy free margarine e.g Pure, and also cholesterol lowering | veg_leek | 80 | veg_leek_marg_soya_chol | 80 | 0% | Fat (1.25%) used for cooking taken into account in the updated version - 25% cooked with fat |
| Leeks cooked with added fat: Fat specified as soya/vegan/dairy free margarine e.g Pure, and not specified amount of fat | veg_leek | 80 | veg_leek_marg_soya_dunno | 80 | 0% | Fat (1.25%) used for cooking taken into account in the updated version - 25% cooked with fat |
| Leeks cooked with added fat: Fat specified as soya/vegan/dairy free margarine (e.g Pure), normal amount % fat | veg_leek | 80 | veg_leek_marg_soya_fat | 80 | 0% | Fat (1.25%) used for cooking taken into account in the updated version - 25% cooked with fat |
| Leeks cooked with added fat: Fat specified as soya/vegan/dairy free margarine (e.g Pure), low fat | veg_leek | 80 | veg_leek_marg_soya_lowfat | 80 | 0% | Fat (1.25%) used for cooking taken into account in the updated version - 25% cooked with fat |
| Leeks cooked with added fat: Fat specified as soya/vegan/dairy free margarine (e.g Pure), very low fat | veg_leek | 80 | veg_leek_marg_soya_vlowfat | 80 | 0% | Fat (1.25%) used for cooking taken into account in the updated version - 25% cooked with fat |
| Leeks cooked with added fat: Fat specified as olive oil | veg_leek | 80 | veg_leek_oil_olive | 80 | 0% | Fat (1.25%) used for cooking taken into account in the updated version - 25% cooked with fat |
| Leeks cooked with added fat: Fat specified as other oil e.g corn, groundnut, rice bran oil | veg_leek | 80 | veg_leek_oil_other | 80 | 0% | Fat (1.25%) used for cooking taken into account in the updated version - 25% cooked with fat |
| Leeks cooked with added fat: Fat specified as rapeseed oil | veg_leek | 80 | veg_leek_oil_rapeseed | 80 | 0% | Fat (1.25%) used for cooking taken into account in the updated version - 25% cooked with fat |
| Leeks cooked with added fat: Fat specified as sunflower oil | veg_leek | 80 | veg_leek_oil_sunflower | 80 | 0% | Fat (1.25%) used for cooking taken into account in the updated version - 25% cooked with fat |

| | McCance and Widdowson's | | Nutrient databank + other changes | | | |
|---|---|---|---|---|---|---|
| Variable description | Food item | Portion size | Food item | Portion size | Portion diff% | Description of differences with previous version |
| Leeks cooked with added fat: Fat specified as vegetable oil | veg_leek | 80 | veg_leek_oil_veg | 80 | 0% | Fat (1.25%) used for cooking taken into account in the updated version - 25% cooked with fat |
| Leeks cooked with added fat: Fat specified as dairy spread which is also cholesterol lowering e.g Benecol Buttery | veg_leek | 80 | veg_leek_spread_dairy_chol | 80 | 0% | Fat (1.25%) used for cooking taken into account in the updated version - 25% cooked with fat |
| Leeks cooked with added fat: Fat specified as dairy spread but not specified amount of % fat | veg_leek | 80 | veg_leek_spread_dairy_dunno | 80 | 0% | Fat (1.25%) used for cooking taken into account in the updated version - 25% cooked with fat |
| Leeks cooked with added fat: Fat specified as dairy spread with normal amount of % fat | veg_leek | 80 | veg_leek_spread_dairy_fat | 80 | 0% | Fat (1.25%) used for cooking taken into account in the updated version - 25% cooked with fat |
| Leeks cooked with added fat: Fat specified as dairy spread, low fat | veg_leek | 80 | veg_leek_spread_dairy_lowfat | 80 | 0% | Fat (1.25%) used for cooking taken into account in the updated version - 25% cooked with fat |
| Leeks cooked with added fat: Fat specified as dairy spread, very low fat | veg_leek | 80 | veg_leek_spread_dairy_vlowfat | 80 | 0% | Fat (1.25%) used for cooking taken into account in the updated version - 25% cooked with fat |
| Leeks cooked with added fat: Fat specified as not known type of spread but ticked cholesterol lowering e.g Benecol, Flora pro active | veg_leek | 80 | veg_leek_spread_dunno_chol | 80 | 0% | Fat (1.25%) used for cooking taken into account in the updated version - 25% cooked with fat |
| Leeks cooked with added fat: Fat specified as not known type of spread or amount of % fat | veg_leek | 80 | veg_leek_spread_dunno_dunno | 80 | 0% | Fat (1.25%) used for cooking taken into account in the updated version - 25% cooked with fat |
| Leeks cooked with added fat: Fat specified as not known type of spread but ticked normal amount % fat | veg_leek | 80 | veg_leek_spread_dunno_fat | 80 | 0% | Fat (1.25%) used for cooking taken into account in the updated version - 25% cooked with fat |
| Leeks cooked with added fat: Fat specified as not known type of spread but ticked low fat | veg_leek | 80 | veg_leek_spread_dunno_lowfat | 80 | 0% | Fat (1.25%) used for cooking taken into account in the updated version - 25% cooked with fat |
| Leeks cooked with added fat: Fat specified as not known type of spread but ticked very low fat | veg_leek | 80 | veg_leek_spread_dunno_vlowfat | 80 | 0% | Fat (1.25%) used for cooking taken into account in the updated version - 25% cooked with fat |
| Leeks cooked with added fat: Fat specified as cholesterol lowering olive spread e.g Benecol/Flora pro active olive spread | veg_leek | 80 | veg_leek_spread_olive_chol | 80 | 0% | Fat (1.25%) used for cooking taken into account in the updated version - 25% cooked with fat |
| Leeks cooked with added fat: Fat specified as olive spread but not specified amount of fat | veg_leek | 80 | veg_leek_spread_olive_dunno | 80 | 0% | Fat (1.25%) used for cooking taken into account in the updated version - 25% cooked with fat |
| Leeks cooked with added fat: Fat specified as olive spread with normal amount of % fat | veg_leek | 80 | veg_leek_spread_olive_fat | 80 | 0% | Fat (1.25%) used for cooking taken into account in the updated version - 25% cooked with fat |
| Leeks cooked with added fat: Fat specified as olive spread, low fat | veg_leek | 80 | veg_leek_spread_olive_lowfat | 80 | 0% | Fat (1.25%) used for cooking taken into account in the updated version - 25% cooked with fat |
| Leeks cooked with added fat: Fat specified as olive spread, very low fat | veg_leek | 80 | veg_leek_spread_olive_vlowfat | 80 | 0% | Fat (1.25%) used for cooking taken into account in the updated version - 25% cooked with fat |
| Lettuce | veg_lettuce | 35 | veg_lettuce | 35 | 0% | |
| Mixed vegetables | veg_mixed | 90 | veg_mixed | 90 | 0% | |
| Mixed veg cooked or stir fried with added fat: Fat specified as butter but not specified amount of % fat | veg_mixtures | 90 | veg_mixtures_butter_dunno | 90 | 0% | Fat (5%) used for cooking taken into account in the updated version; mapping changed to stirfry vegetables |
| Mixed veg cooked or stir fried with added fat: Fat specified as butter, normal amount of % fat | veg_mixtures | 90 | veg_mixtures_butter_fat | 90 | 0% | Fat (5%) used for cooking taken into account in the updated version; mapping changed to stirfry vegetables |
| Mixed veg cooked or stir fried with added fat: Fat specified as butter, low fat | veg_mixtures | 90 | veg_mixtures_butter_lowfat | 90 | 0% | Fat (5%) used for cooking taken into account in the updated version; mapping changed to stirfry vegetables |
| Mixed veg cooked or stir fried with added fat: Fat specified as spreadable butter with normal amount of % fat | veg_mixtures | 90 | veg_mixtures_butter_spread_fat | 90 | 0% | Fat (5%) used for cooking taken into account in the updated version; mapping changed to stirfry vegetables |
| Mixed veg cooked or stir fried with added fat: Fat specified as spreadable butter, low fat | veg_mixtures | 90 | veg_mixtures_butter_spread_lowfat | 90 | 0% | Fat (5%) used for cooking taken into account in the updated version; mapping changed to stirfry vegetables |
| Mixed veg cooked or stir fried with added fat: Fat specified as not known type of fat or spread | veg_mixtures | 90 | veg_mixtures_fat_dunno | 90 | 0% | Fat (5%) used for cooking taken into account in the updated version; mapping changed to stirfry vegetables |
| Mixed veg cooked or stir fried with no added fat | veg_mixtures | 90 | veg_mixtures_fat_none | 90 | 0% | Fat (5%) used for cooking taken into account in the updated version; mapping changed to stirfry vegetables |
| Mixed veg cooked or stir fried with added fat: Fat specified as other type of fat or spread e.g ghee, dripping | veg_mixtures | 90 | veg_mixtures_fat_other | 90 | 0% | Fat (5%) used for cooking taken into account in the updated version; mapping changed to stirfry vegetables |
| Mixed veg cooked or stir fried with added fat: Fat specified as lard | veg_mixtures | 90 | veg_mixtures_lard | 90 | 0% | Fat (5%) used for cooking taken into account in the updated version; mapping changed to stirfry vegetables |
| Mixed veg cooked or stir fried with added fat: Fat specified as hard margarine (hard block margarine in wrapper) | veg_mixtures | 90 | veg_mixtures_marg_hard | 90 | 0% | Fat (5%) used for cooking taken into account in the updated version; mapping changed to stirfry vegetables |
| Mixed veg cooked or stir fried with added fat: Fat specified as polyunsaturated margarine (e.g Flora) and also cholesterol lowering | veg_mixtures | 90 | veg_mixtures_marg_poly_chol | 90 | 0% | Fat (5%) used for cooking taken into account in the updated version; mapping changed to stirfry vegetables |
| Mixed veg cooked or stir fried with added fat: Fat specified as polyunsaturated margarine (e.g Flora) but not specified amount of fat | veg_mixtures | 90 | veg_mixtures_marg_poly_dunno | 90 | 0% | Fat (5%) used for cooking taken into account in the updated version; mapping changed to stirfry vegetables |
| Mixed veg cooked or stir fried with added fat: Fat specified as polyunsaturated margarine (e.g Flora), normal amount of % fat | veg_mixtures | 90 | veg_mixtures_marg_poly_fat | 90 | 0% | Fat (5%) used for cooking taken into account in the updated version; mapping changed to stirfry vegetables |
| Mixed veg cooked or stir fried with added fat: Fat specified as polyunsaturated margarine (e.g Flora), low fat | veg_mixtures | 90 | veg_mixtures_marg_poly_lowfat | 90 | 0% | Fat (5%) used for cooking taken into account in the updated version; mapping changed to stirfry vegetables |
| Mixed veg cooked or stir fried with added fat: Fat specified as polyunsaturated margarine (e.g Flora), very low fat | veg_mixtures | 90 | veg_mixtures_marg_poly_vlowfat | 90 | 0% | Fat (5%) used for cooking taken into account in the updated version; mapping changed to stirfry vegetables |
| Mixed veg cooked or stir fried with added fat: Fat specified as soya/vegan/dairy free margarine e.g Pure, and also cholesterol lowering | veg_mixtures | 90 | veg_mixtures_marg_soya_chol | 90 | 0% | Fat (5%) used for cooking taken into account in the updated version; mapping changed to stirfry vegetables |
| Mixed veg cooked or stir fried with added fat: Fat specified as soya/vegan/dairy free margarine e.g Pure, and not specified amount of fat | veg_mixtures | 90 | veg_mixtures_marg_soya_dunno | 90 | 0% | Fat (5%) used for cooking taken into account in the updated version; mapping changed to stirfry vegetables |
| Mixed veg cooked or stir fried with added fat: Fat specified as soya/vegan/dairy free margarine (e.g Pure), normal amount % fat | veg_mixtures | 90 | veg_mixtures_marg_soya_fat | 90 | 0% | Fat (5%) used for cooking taken into account in the updated version; mapping changed to stirfry vegetables |
| Mixed veg cooked or stir fried with added fat: Fat specified as soya/vegan/dairy free margarine (e.g Pure), low fat | veg_mixtures | 90 | veg_mixtures_marg_soya_lowfat | 90 | 0% | Fat (5%) used for cooking taken into account in the updated version; mapping changed to stirfry vegetables |
| Mixed veg cooked or stir fried with added fat: Fat specified as soya/vegan/dairy free margarine (e.g Pure), very low fat | veg_mixtures | 90 | veg_mixtures_marg_soya_vlowfat | 90 | 0% | Fat (5%) used for cooking taken into account in the updated version; mapping changed to stirfry vegetables |
| Mixed veg cooked or stir fried with added fat: Fat specified as olive oil | veg_mixtures | 90 | veg_mixtures_oil_olive | 90 | 0% | Fat (5%) used for cooking taken into account in the updated version; mapping changed to stirfry vegetables |
| Mixed veg cooked or stir fried with added fat: Fat specified as other oil e.g corn, groundnut, rice bran oil | veg_mixtures | 90 | veg_mixtures_oil_other | 90 | 0% | Fat (5%) used for cooking taken into account in the updated version; mapping changed to stirfry vegetables |
| Mixed veg cooked or stir fried with added fat: Fat specified as rapeseed oil | veg_mixtures | 90 | veg_mixtures_oil_rapeseed | 90 | 0% | Fat (5%) used for cooking taken into account in the updated version; mapping changed to stirfry vegetables |
| Mixed veg cooked or stir fried with added fat: Fat specified as sunflower oil | veg_mixtures | 90 | veg_mixtures_oil_sunflower | 90 | 0% | Fat (5%) used for cooking taken into account in the updated version; mapping changed to stirfry vegetables |
| Mixed veg cooked or stir fried with added fat: Fat specified as vegetable oil | veg_mixtures | 90 | veg_mixtures_oil_veg | 90 | 0% | Fat (5%) used for cooking taken into account in the updated version; mapping changed to stirfry vegetables |
| Mixed veg cooked or stir fried with added fat: Fat specified as dairy spread which is also cholesterol lowering e.g Benecol Buttery | veg_mixtures | 90 | veg_mixtures_spread_dairy_chol | 90 | 0% | Fat (5%) used for cooking taken into account in the updated version; mapping changed to stirfry vegetables |
| Mixed veg cooked or stir fried with added fat: Fat specified as dairy spread but not specified amount of % fat | veg_mixtures | 90 | veg_mixtures_spread_dairy_dunno | 90 | 0% | Fat (5%) used for cooking taken into account in the updated version; mapping changed to stirfry vegetables |
| Mixed veg cooked or stir fried with added fat: Fat specified as dairy spread with normal amount of % fat | veg_mixtures | 90 | veg_mixtures_spread_dairy_fat | 90 | 0% | Fat (5%) used for cooking taken into account in the updated version; mapping changed to stirfry vegetables |
| Mixed veg cooked or stir fried with added fat: Fat specified as dairy spread, low fat | veg_mixtures | 90 | veg_mixtures_spread_dairy_lowfat | 90 | 0% | Fat (5%) used for cooking taken into account in the updated version; mapping changed to stirfry vegetables |
| Mixed veg cooked or stir fried with added fat: Fat specified as dairy spread, very low fat | veg_mixtures | 90 | veg_mixtures_spread_dairy_vlowfat | 90 | 0% | Fat (5%) used for cooking taken into account in the updated version; mapping changed to stirfry vegetables |
| Mixed veg cooked or stir fried with added fat: Fat specified as not known type of spread but ticked cholesterol lowering e.g Benecol, Flora pro active | veg_mixtures | 90 | veg_mixtures_spread_dunno_chol | 90 | 0% | Fat (5%) used for cooking taken into account in the updated version; mapping changed to stirfry vegetables |
| Mixed veg cooked or stir fried with added fat: Fat specified as not known type of spread or amount of % fat | veg_mixtures | 90 | veg_mixtures_spread_dunno_dunno | 90 | 0% | Fat (5%) used for cooking taken into account in the updated version; mapping changed to stirfry vegetables |
| Mixed veg cooked or stir fried with added fat: Fat specified as not known type of spread but ticked normal amount % fat | veg_mixtures | 90 | veg_mixtures_spread_dunno_fat | 90 | 0% | Fat (5%) used for cooking taken into account in the updated version; mapping changed to stirfry vegetables |
| Mixed veg cooked or stir fried with added fat: Fat specified as not known type of spread but ticked low fat | veg_mixtures | 90 | veg_mixtures_spread_dunno_lowfat | 90 | 0% | Fat (5%) used for cooking taken into account in the updated version; mapping changed to stirfry vegetables |
| Mixed veg cooked or stir fried with added fat: Fat specified as not known type of spread but ticked very low fat | veg_mixtures | 90 | veg_mixtures_spread_dunno_vlowfat | 90 | 0% | Fat (5%) used for cooking taken into account in the updated version; mapping changed to stirfry vegetables |
| Mixed veg cooked or stir fried with added fat: Fat specified as cholesterol lowering olive spread e.g Benecol/Flora pro active olive spread | veg_mixtures | 90 | veg_mixtures_spread_olive_chol | 90 | 0% | Fat (5%) used for cooking taken into account in the updated version; mapping changed to stirfry vegetables |
| Mixed veg cooked or stir fried with added fat: Fat specified as olive spread but not specified amount of fat | veg_mixtures | 90 | veg_mixtures_spread_olive_dunno | 90 | 0% | Fat (5%) used for cooking taken into account in the updated version; mapping changed to stirfry vegetables |
| Mixed veg cooked or stir fried with added fat: Fat specified as olive spread with normal amount of % fat | veg_mixtures | 90 | veg_mixtures_spread_olive_fat | 90 | 0% | Fat (5%) used for cooking taken into account in the updated version; mapping changed to stirfry vegetables |
| Mixed veg cooked or stir fried with added fat: Fat specified as olive spread, low fat | veg_mixtures | 90 | veg_mixtures_spread_olive_lowfat | 90 | 0% | Fat (5%) used for cooking taken into account in the updated version; mapping changed to stirfry vegetables |
| Mixed veg cooked or stir fried with added fat: Fat specified as olive spread, very low fat | veg_mixtures | 90 | veg_mixtures_spread_olive_vlowfat | 90 | 0% | Fat (5%) used for cooking taken into account in the updated version; mapping changed to stirfry vegetables |
| Mushrooms cooked with added fat: Fat specified as butter but not specified amount of % fat | veg_mushrooms | 44 | veg_mushrooms_butter_dunno | 44 | 0% | Fat (2.5%) used for cooking taken into account in the updated version - 50% cooked with fat |
| Mushrooms cooked with added fat: Fat specified as butter, normal amount of % fat | veg_mushrooms | 44 | veg_mushrooms_butter_fat | 44 | 0% | Fat (2.5%) used for cooking taken into account in the updated version - 50% cooked with fat |
| Mushrooms cooked with added fat: Fat specified as butter, low fat | veg_mushrooms | 44 | veg_mushrooms_butter_lowfat | 44 | 0% | Fat (2.5%) used for cooking taken into account in the updated version - 50% cooked with fat |
| Mushrooms cooked with added fat: Fat specified as spreadable butter with normal amount of % fat | veg_mushrooms | 44 | veg_mushrooms_butter_spread_fat | 44 | 0% | Fat (2.5%) used for cooking taken into account in the updated version - 50% cooked with fat |
| Mushrooms cooked with added fat: Fat specified as spreadable butter, low fat | veg_mushrooms | 44 | veg_mushrooms_butter_spread_lowfat | 44 | 0% | Fat (2.5%) used for cooking taken into account in the updated version - 50% cooked with fat |
| Mushrooms cooked with added fat: Fat specified as not known type of fat or spread | veg_mushrooms | 44 | veg_mushrooms_fat_dunno | 44 | 0% | Fat (2.5%) used for cooking taken into account in the updated version - 50% cooked with fat |
| Mushrooms cooked with no added fat | veg_mushrooms | 44 | veg_mushrooms_fat_none | 44 | 0% | Fat (2.5%) used for cooking taken into account in the updated version - 50% cooked with fat |
| Mushrooms cooked with added fat: Fat specified as other type of fat or spread e.g ghee, dripping | veg_mushrooms | 44 | veg_mushrooms_fat_other | 44 | 0% | Fat (2.5%) used for cooking taken into account in the updated version - 50% cooked with fat |
| Mushrooms cooked with added fat: Fat specified as lard | veg_mushrooms | 44 | veg_mushrooms_lard | 44 | 0% | Fat (2.5%) used for cooking taken into account in the updated version - 50% cooked with fat |
| Mushrooms cooked with added fat: Fat specified as hard margarine (hard block margarine in wrapper) | veg_mushrooms | 44 | veg_mushrooms_marg_hard | 44 | 0% | Fat (2.5%) used for cooking taken into account in the updated version - 50% cooked with fat |
| Mushrooms cooked with added fat: Fat specified as polyunsaturated margarine (e.g Flora) and also cholesterol lowering | veg_mushrooms | 44 | veg_mushrooms_marg_poly_chol | 44 | 0% | Fat (2.5%) used for cooking taken into account in the updated version - 50% cooked with fat |
| Mushrooms cooked with added fat: Fat specified as polyunsaturated margarine (e.g Flora) but not specified amount of fat | veg_mushrooms | 44 | veg_mushrooms_marg_poly_dunno | 44 | 0% | Fat (2.5%) used for cooking taken into account in the updated version - 50% cooked with fat |
| Mushrooms cooked with added fat: Fat specified as polyunsaturated margarine (e.g Flora), normal amount of % fat | veg_mushrooms | 44 | veg_mushrooms_marg_poly_fat | 44 | 0% | Fat (2.5%) used for cooking taken into account in the updated version - 50% cooked with fat |
| Mushrooms cooked with added fat: Fat specified as polyunsaturated margarine (e.g Flora), low fat | veg_mushrooms | 44 | veg_mushrooms_marg_poly_lowfat | 44 | 0% | Fat (2.5%) used for cooking taken into account in the updated version - 50% cooked with fat |
| Mushrooms cooked with added fat: Fat specified as polyunsaturated margarine (e.g Flora), very low fat | veg_mushrooms | 44 | veg_mushrooms_marg_poly_vlowfat | 44 | 0% | Fat (2.5%) used for cooking taken into account in the updated version - 50% cooked with fat |
| Mushrooms cooked with added fat: Fat specified as soya/vegan/dairy free margarine e.g Pure, and also cholesterol lowering | veg_mushrooms | 44 | veg_mushrooms_marg_soya_chol | 44 | 0% | Fat (2.5%) used for cooking taken into account in the updated version - 50% cooked with fat |
| Mushrooms cooked with added fat: Fat specified as soya/vegan/dairy free margarine e.g Pure, and not specified amount of fat | veg_mushrooms | 44 | veg_mushrooms_marg_soya_dunno | 44 | 0% | Fat (2.5%) used for cooking taken into account in the updated version - 50% cooked with fat |
| Mushrooms cooked with added fat: Fat specified as soya/vegan/dairy free margarine (e.g Pure), normal amount % fat | veg_mushrooms | 44 | veg_mushrooms_marg_soya_fat | 44 | 0% | Fat (2.5%) used for cooking taken into account in the updated version - 50% cooked with fat |
| Mushrooms cooked with added fat: Fat specified as soya/vegan/dairy free margarine (e.g Pure), low fat | veg_mushrooms | 44 | veg_mushrooms_marg_soya_lowfat | 44 | 0% | Fat (2.5%) used for cooking taken into account in the updated version - 50% cooked with fat |
| Mushrooms cooked with added fat: Fat specified as soya/vegan/dairy free margarine (e.g Pure), very low fat | veg_mushrooms | 44 | veg_mushrooms_marg_soya_vlowfat | 44 | 0% | Fat (2.5%) used for cooking taken into account in the updated version - 50% cooked with fat |
| Mushrooms cooked with added fat: Fat specified as olive oil | veg_mushrooms | 44 | veg_mushrooms_oil_olive | 44 | 0% | Fat (2.5%) used for cooking taken into account in the updated version - 50% cooked with fat |
| Mushrooms cooked with added fat: Fat specified as other oil e.g corn, groundnut, rice bran oil | veg_mushrooms | 44 | veg_mushrooms_oil_other | 44 | 0% | Fat (2.5%) used for cooking taken into account in the updated version - 50% cooked with fat |
| Mushrooms cooked with added fat: Fat specified as rapeseed oil | veg_mushrooms | 44 | veg_mushrooms_oil_rapeseed | 44 | 0% | Fat (2.5%) used for cooking taken into account in the updated version - 50% cooked with fat |
| Mushrooms cooked with added fat: Fat specified as sunflower oil | veg_mushrooms | 44 | veg_mushrooms_oil_sunflower | 44 | 0% | Fat (2.5%) used for cooking taken into account in the updated version - 50% cooked with fat |
| Mushrooms cooked with added fat: Fat specified as vegetable oil | veg_mushrooms | 44 | veg_mushrooms_oil_veg | 44 | 0% | Fat (2.5%) used for cooking taken into account in the updated version - 50% cooked with fat |
| Mushrooms cooked with added fat: Fat specified as dairy spread which is also cholesterol lowering e.g Benecol Buttery | veg_mushrooms | 44 | veg_mushrooms_spread_dairy_chol | 44 | 0% | Fat (2.5%) used for cooking taken into account in the updated version - 50% cooked with fat |
| Mushrooms cooked with added fat: Fat specified as dairy spread but not specified amount of % fat | veg_mushrooms | 44 | veg_mushrooms_spread_dairy_dunno | 44 | 0% | Fat (2.5%) used for cooking taken into account in the updated version - 50% cooked with fat |
| Mushrooms cooked with added fat: Fat specified as dairy spread with normal amount of % fat | veg_mushrooms | 44 | veg_mushrooms_spread_dairy_fat | 44 | 0% | Fat (2.5%) used for cooking taken into account in the updated version - 50% cooked with fat |
| Mushrooms cooked with added fat: Fat specified as dairy spread, low fat | veg_mushrooms | 44 | veg_mushrooms_spread_dairy_lowfat | 44 | 0% | Fat (2.5%) used for cooking taken into account in the updated version - 50% cooked with fat |
| Mushrooms cooked with added fat: Fat specified as dairy spread, very low fat | veg_mushrooms | 44 | veg_mushrooms_spread_dairy_vlowfat | 44 | 0% | Fat (2.5%) used for cooking taken into account in the updated version - 50% cooked with fat |
| Mushrooms cooked with added fat: Fat specified as not known type of spread but ticked cholesterol lowering e.g Benecol, Flora pro active | veg_mushrooms | 44 | veg_mushrooms_spread_dunno_chol | 44 | 0% | Fat (2.5%) used for cooking taken into account in the updated version - 50% cooked with fat |
| Mushrooms cooked with added fat: Fat specified as not known type of spread or amount of % fat | veg_mushrooms | 44 | veg_mushrooms_spread_dunno_dunno | 44 | 0% | Fat (2.5%) used for cooking taken into account in the updated version - 50% cooked with fat |
| Mushrooms cooked with added fat: Fat specified as not known type of spread but ticked normal amount % fat | veg_mushrooms | 44 | veg_mushrooms_spread_dunno_fat | 44 | 0% | Fat (2.5%) used for cooking taken into account in the updated version - 50% cooked with fat |
| Mushrooms cooked with added fat: Fat specified as not known type of spread but ticked low fat | veg_mushrooms | 44 | veg_mushrooms_spread_dunno_lowfat | 44 | 0% | Fat (2.5%) used for cooking taken into account in the updated version - 50% cooked with fat |
| Mushrooms cooked with added fat: Fat specified as not known type of spread but ticked very low fat | veg_mushrooms | 44 | veg_mushrooms_spread_dunno_vlowfat | 44 | 0% | Fat (2.5%) used for cooking taken into account in the updated version - 50% cooked with fat |
| Mushrooms cooked with added fat: Fat specified as cholesterol lowering olive spread e.g Benecol/Flora pro active olive spread | veg_mushrooms | 44 | veg_mushrooms_spread_olive_chol | 44 | 0% | Fat (2.5%) used for cooking taken into account in the updated version - 50% cooked with fat |
| Mushrooms cooked with added fat: Fat specified as olive spread but not specified amount of fat | veg_mushrooms | 44 | veg_mushrooms_spread_olive_dunno | 44 | 0% | Fat (2.5%) used for cooking taken into account in the updated version - 50% cooked with fat |
| Mushrooms cooked with added fat: Fat specified as olive spread with normal amount of % fat | veg_mushrooms | 44 | veg_mushrooms_spread_olive_fat | 44 | 0% | Fat (2.5%) used for cooking taken into account in the updated version - 50% cooked with fat |
| Mushrooms cooked with added fat: Fat specified as olive spread, low fat | veg_mushrooms | 44 | veg_mushrooms_spread_olive_lowfat | 44 | 0% | Fat (2.5%) used for cooking taken into account in the updated version - 50% cooked with fat |
| Mushrooms cooked with added fat: Fat specified as olive spread, very low fat | veg_mushrooms | 44 | veg_mushrooms_spread_olive_vlowfat | 44 | 0% | Fat (2.5%) used for cooking taken into account in the updated version - 50% cooked with fat |
| Onions, shallots, spring onion cooked with added fat: Fat specified as butter but not specified amount of % fat | veg_onion | 60 | veg_onion_butter_dunno | 60 | 0% | Fat (2.5%) used for cooking taken into account in the updated version - 50% cooked with fat |
| Onions, shallots, spring onion cooked with added fat: Fat specified as butter, normal amount of % fat | veg_onion | 60 | veg_onion_butter_fat | 60 | 0% | Fat (2.5%) used for cooking taken into account in the updated version - 50% cooked with fat |
| Onions, shallots, spring onion cooked with added fat: Fat specified as butter, low fat | veg_onion | 60 | veg_onion_butter_lowfat | 60 | 0% | Fat (2.5%) used for cooking taken into account in the updated version - 50% cooked with fat |
| Onions, shallots, spring onion cooked with added fat: Fat specified as spreadable butter with normal amount of % fat | veg_onion | 60 | veg_onion_butter_spread_fat | 60 | 0% | Fat (2.5%) used for cooking taken into account in the updated version - 50% cooked with fat |
| Onions, shallots, spring onion cooked with added fat: Fat specified as spreadable butter, low fat | veg_onion | 60 | veg_onion_butter_spread_lowfat | 60 | 0% | Fat (2.5%) used for cooking taken into account in the updated version - 50% cooked with fat |
| Onions, shallots, spring onion cooked with added fat: Fat specified as not known type of fat or spread | veg_onion | 60 | veg_onion_fat_dunno | 60 | 0% | Fat (2.5%) used for cooking taken into account in the updated version - 50% cooked with fat |
| Onions, shallots, spring onion cooked with no added fat | veg_onion | 60 | veg_onion_fat_none | 60 | 0% | Fat (2.5%) used for cooking taken into account in the updated version - 50% cooked with fat |
| Onions, shallots, spring onion cooked with added fat: Fat specified as other type of fat or spread e.g ghee, dripping | veg_onion | 60 | veg_onion_fat_other | 60 | 0% | Fat (2.5%) used for cooking taken into account in the updated version - 50% cooked with fat |
| Onions, shallots, spring onion cooked with added fat: Fat specified as lard | veg_onion | 60 | veg_onion_lard | 60 | 0% | Fat (2.5%) used for cooking taken into account in the updated version - 50% cooked with fat |
| Onions, shallots, spring onion cooked with added fat: Fat specified as hard margarine (hard block margarine in wrapper) | veg_onion | 60 | veg_onion_marg_hard | 60 | 0% | Fat (2.5%) used for cooking taken into account in the updated version - 50% cooked with fat |
| Onions, shallots, spring onion cooked with added fat: Fat specified as polyunsaturated margarine (e.g Flora) and also cholesterol lowering | veg_onion | 60 | veg_onion_marg_poly_chol | 60 | 0% | Fat (2.5%) used for cooking taken into account in the updated version - 50% cooked with fat |

| | McCance and Widdowson's | | Nutrient databank + other changes | | | |
|---|---|---|---|---|---|---|
| Variable description | Food item | Portion size | Food item | Portion size | Portion diff% | Description of differences with previous version |
| Onions, shallots, spring onion cooked with added fat: Fat specified as polyunsaturated margarine (e.g Flora) but not specified amount of fat | veg_onion | 60 | veg_onion_marg_poly_dunno | 60 | 0% | Fat (2.5%) used for cooking taken into account in the updated version - 50% cooked with fat |
| Onions, shallots, spring onion cooked with added fat: Fat specified as polyunsaturated margarine (e.g Flora), normal amount of % fat | veg_onion | 60 | veg_onion_marg_poly_fat | 60 | 0% | Fat (2.5%) used for cooking taken into account in the updated version - 50% cooked with fat |
| Onions, shallots, spring onion cooked with added fat: Fat specified as polyunsaturated margarine (e.g Flora), low fat | veg_onion | 60 | veg_onion_marg_poly_lowfat | 60 | 0% | Fat (2.5%) used for cooking taken into account in the updated version - 50% cooked with fat |
| Onions, shallots, spring onion cooked with added fat: Fat specified as polyunsaturated margarine (e.g Flora), very low fat | veg_onion | 60 | veg_onion_marg_poly_vlowfat | 60 | 0% | Fat (2.5%) used for cooking taken into account in the updated version - 50% cooked with fat |
| Onions, shallots, spring onion cooked with added fat: Fat specified as soya/vegan/dairy free margarine e.g Pure, and also cholesterol lowering | veg_onion | 60 | veg_onion_marg_soya_chol | 60 | 0% | Fat (2.5%) used for cooking taken into account in the updated version - 50% cooked with fat |
| Onions, shallots, spring onion cooked with added fat: Fat specified as soya/vegan/dairy free margarine e.g Pure, and not specified amount of fat | veg_onion | 60 | veg_onion_marg_soya_dunno | 60 | 0% | Fat (2.5%) used for cooking taken into account in the updated version - 50% cooked with fat |
| Onions, shallots, spring onion cooked with added fat: Fat specified as soya/vegan/dairy free margarine (e.g Pure), normal amount % fat | veg_onion | 60 | veg_onion_marg_soya_fat | 60 | 0% | Fat (2.5%) used for cooking taken into account in the updated version - 50% cooked with fat |
| Onions, shallots, spring onion cooked with added fat: Fat specified as soya/vegan/dairy free margarine (e.g Pure), low fat | veg_onion | 60 | veg_onion_marg_soya_lowfat | 60 | 0% | Fat (2.5%) used for cooking taken into account in the updated version - 50% cooked with fat |
| Onions, shallots, spring onion cooked with added fat: Fat specified as soya/vegan/dairy free margarine (e.g Pure), very low fat | veg_onion | 60 | veg_onion_marg_soya_vlowfat | 60 | 0% | Fat (2.5%) used for cooking taken into account in the updated version - 50% cooked with fat |
| Onions, shallots, spring onion cooked with added fat: Fat specified as olive oil | veg_onion | 60 | veg_onion_oil_olive | 60 | 0% | Fat (2.5%) used for cooking taken into account in the updated version - 50% cooked with fat |
| Onions, shallots, spring onion cooked with added fat: Fat specified as other oil e.g corn, groundnut, rice bran oil | veg_onion | 60 | veg_onion_oil_other | 60 | 0% | Fat (2.5%) used for cooking taken into account in the updated version - 50% cooked with fat |
| Onions, shallots, spring onion cooked with added fat: Fat specified as rapeseed oil | veg_onion | 60 | veg_onion_oil_rapeseed | 60 | 0% | Fat (2.5%) used for cooking taken into account in the updated version - 50% cooked with fat |
| Onions, shallots, spring onion cooked with added fat: Fat specified as sunflower oil | veg_onion | 60 | veg_onion_oil_sunflower | 60 | 0% | Fat (2.5%) used for cooking taken into account in the updated version - 50% cooked with fat |
| Onions, shallots, spring onion cooked with added fat: Fat specified as vegetable oil | veg_onion | 60 | veg_onion_oil_veg | 60 | 0% | Fat (2.5%) used for cooking taken into account in the updated version - 50% cooked with fat |
| Onions, shallots, spring onion cooked with added fat: Fat specified as dairy spread which is also cholesterol lowering e.g Benecol Buttery | veg_onion | 60 | veg_onion_spread_dairy_chol | 60 | 0% | Fat (2.5%) used for cooking taken into account in the updated version - 50% cooked with fat |
| Onions, shallots, spring onion cooked with added fat: Fat specified as dairy spread but not specified amount of % fat | veg_onion | 60 | veg_onion_spread_dairy_dunno | 60 | 0% | Fat (2.5%) used for cooking taken into account in the updated version - 50% cooked with fat |
| Onions, shallots, spring onion cooked with added fat: Fat specified as dairy spread with normal amount of % fat | veg_onion | 60 | veg_onion_spread_dairy_fat | 60 | 0% | Fat (2.5%) used for cooking taken into account in the updated version - 50% cooked with fat |
| Onions, shallots, spring onion cooked with added fat: Fat specified as dairy spread, low fat | veg_onion | 60 | veg_onion_spread_dairy_lowfat | 60 | 0% | Fat (2.5%) used for cooking taken into account in the updated version - 50% cooked with fat |
| Onions, shallots, spring onion cooked with added fat: Fat specified as dairy spread, very low fat | veg_onion | 60 | veg_onion_spread_dairy_vlowfat | 60 | 0% | Fat (2.5%) used for cooking taken into account in the updated version - 50% cooked with fat |
| Onions, shallots, spring onion cooked with added fat: Fat specified as not known type of spread but ticked cholesterol lowering e.g Benecol, Flora pro active | veg_onion | 60 | veg_onion_spread_dunno_chol | 60 | 0% | Fat (2.5%) used for cooking taken into account in the updated version - 50% cooked with fat |
| Onions, shallots, spring onion cooked with added fat: Fat specified as not known type of spread or amount of % fat | veg_onion | 60 | veg_onion_spread_dunno_dunno | 60 | 0% | Fat (2.5%) used for cooking taken into account in the updated version - 50% cooked with fat |
| Onions, shallots, spring onion cooked with added fat: Fat specified as not known type of spread but ticked normal amount % fat | veg_onion | 60 | veg_onion_spread_dunno_fat | 60 | 0% | Fat (2.5%) used for cooking taken into account in the updated version - 50% cooked with fat |
| Onions, shallots, spring onion cooked with added fat: Fat specified as not known type of spread but ticked low fat | veg_onion | 60 | veg_onion_spread_dunno_lowfat | 60 | 0% | Fat (2.5%) used for cooking taken into account in the updated version - 50% cooked with fat |
| Onions, shallots, spring onion cooked with added fat: Fat specified as not known type of spread but ticked very low fat | veg_onion | 60 | veg_onion_spread_dunno_vlowfat | 60 | 0% | Fat (2.5%) used for cooking taken into account in the updated version - 50% cooked with fat |
| Onions, shallots, spring onion cooked with added fat: Fat specified as cholesterol lowering olive spread e.g Benecol/Flora pro active olive spread | veg_onion | 60 | veg_onion_spread_olive_chol | 60 | 0% | Fat (2.5%) used for cooking taken into account in the updated version - 50% cooked with fat |
| Onions, shallots, spring onion cooked with added fat: Fat specified as olive spread but not specified amount of fat | veg_onion | 60 | veg_onion_spread_olive_dunno | 60 | 0% | Fat (2.5%) used for cooking taken into account in the updated version - 50% cooked with fat |
| Onions, shallots, spring onion cooked with added fat: Fat specified as olive spread with normal amount of % fat | veg_onion | 60 | veg_onion_spread_olive_fat | 60 | 0% | Fat (2.5%) used for cooking taken into account in the updated version - 50% cooked with fat |
| Onions, shallots, spring onion cooked with added fat: Fat specified as olive spread, low fat | veg_onion | 60 | veg_onion_spread_olive_lowfat | 60 | 0% | Fat (2.5%) used for cooking taken into account in the updated version - 50% cooked with fat |
| Onions, shallots, spring onion cooked with added fat: Fat specified as olive spread, very low fat | veg_onion | 60 | veg_onion_spread_olive_vlowfat | 60 | 0% | Fat (2.5%) used for cooking taken into account in the updated version - 50% cooked with fat |
| Other veg e.g asparagus, aubergine, pumpkin cooked with added fat: Fat specified as butter but not specified amount of % fat | veg_other | 60 | veg_other_butter_dunno | 60 | 0% | Fat (3%) used for cooking taken into account in the updated version; updated mapping based on free text entered by study participants |
| Other veg e.g asparagus, aubergine, pumpkin cooked with added fat: Fat specified as butter, normal amount of % fat | veg_other | 60 | veg_other_butter_fat | 60 | 0% | Fat (3%) used for cooking taken into account in the updated version; updated mapping based on free text entered by study participants |
| Other veg e.g asparagus, aubergine, pumpkin cooked with added fat: Fat specified as butter, low fat | veg_other | 60 | veg_other_butter_lowfat | 60 | 0% | Fat (3%) used for cooking taken into account in the updated version; updated mapping based on free text entered by study participants |
| Other veg e.g asparagus, aubergine, pumpkin cooked with added fat: Fat specified as spreadable butter with normal amount of % fat | veg_other | 60 | veg_other_butter_spread_fat | 60 | 0% | Fat (3%) used for cooking taken into account in the updated version; updated mapping based on free text entered by study participants |
| Other veg e.g asparagus, aubergine, pumpkin cooked with added fat: Fat specified as spreadable butter, low fat | veg_other | 60 | veg_other_butter_spread_lowfat | 60 | 0% | Fat (3%) used for cooking taken into account in the updated version; updated mapping based on free text entered by study participants |
| Other veg e.g asparagus, aubergine, pumpkin cooked with added fat: Fat specified as not known type of fat or spread | veg_other | 60 | veg_other_fat_dunno | 60 | 0% | Fat (3%) used for cooking taken into account in the updated version; updated mapping based on free text entered by study participants |
| Other veg e.g asparagus, aubergine, pumpkin cooked with no added fat | veg_other | 60 | veg_other_fat_none | 60 | 0% | Fat (3%) used for cooking taken into account in the updated version; updated mapping based on free text entered by study participants |
| Other veg e.g asparagus, aubergine, pumpkin cooked with added fat: Fat specified as other type of fat or spread e.g ghee, dripping | veg_other | 60 | veg_other_fat_other | 60 | 0% | Fat (3%) used for cooking taken into account in the updated version; updated mapping based on free text entered by study participants |
| Other veg e.g asparagus, aubergine, pumpkin cooked with added fat: Fat specified as lard | veg_other | 60 | veg_other_lard | 60 | 0% | Fat (3%) used for cooking taken into account in the updated version; updated mapping based on free text entered by study participants |
| Other veg e.g asparagus, aubergine, pumpkin cooked with added fat: Fat specified as hard margarine (hard block margarine in wrapper) | veg_other | 60 | veg_other_marg_hard | 60 | 0% | Fat (3%) used for cooking taken into account in the updated version; updated mapping based on free text entered by study participants |
| Other veg e.g asparagus, aubergine, pumpkin cooked with added fat: Fat specified as polyunsaturated margarine (e.g Flora) and also cholesterol lowering | veg_other | 60 | veg_other_marg_poly_chol | 60 | 0% | Fat (3%) used for cooking taken into account in the updated version; updated mapping based on free text entered by study participants |
| Other veg e.g asparagus, aubergine, pumpkin cooked with added fat: Fat specified as polyunsaturated margarine (e.g Flora) but not specified amount of fat | veg_other | 60 | veg_other_marg_poly_dunno | 60 | 0% | Fat (3%) used for cooking taken into account in the updated version; updated mapping based on free text entered by study participants |
| Other veg e.g asparagus, aubergine, pumpkin cooked with added fat: Fat specified as polyunsaturated margarine (e.g Flora), normal amount of % fat | veg_other | 60 | veg_other_marg_poly_fat | 60 | 0% | Fat (3%) used for cooking taken into account in the updated version; updated mapping based on free text entered by study participants |
| Other veg e.g asparagus, aubergine, pumpkin cooked with added fat: Fat specified as polyunsaturated margarine (e.g Flora), low fat | veg_other | 60 | veg_other_marg_poly_lowfat | 60 | 0% | Fat (3%) used for cooking taken into account in the updated version; updated mapping based on free text entered by study participants |
| Other veg e.g asparagus, aubergine, pumpkin cooked with added fat: Fat specified as polyunsaturated margarine (e.g Flora), very low fat | veg_other | 60 | veg_other_marg_poly_vlowfat | 60 | 0% | Fat (3%) used for cooking taken into account in the updated version; updated mapping based on free text entered by study participants |
| Other veg e.g asparagus, aubergine, pumpkin cooked with added fat: Fat specified as soya/vegan/dairy free margarine e.g Pure, and also cholesterol lowering | veg_other | 60 | veg_other_marg_soya_chol | 60 | 0% | Fat (3%) used for cooking taken into account in the updated version; updated mapping based on free text entered by study participants |
| Other veg e.g asparagus, aubergine, pumpkin cooked with added fat: Fat specified as soya/vegan/dairy free margarine e.g Pure, and not specified amount of fat | veg_other | 60 | veg_other_marg_soya_dunno | 60 | 0% | Fat (3%) used for cooking taken into account in the updated version; updated mapping based on free text entered by study participants |
| Other veg e.g asparagus, aubergine, pumpkin cooked with added fat: Fat specified as soya/vegan/dairy free margarine (e.g Pure), normal amount % fat | veg_other | 60 | veg_other_marg_soya_fat | 60 | 0% | Fat (3%) used for cooking taken into account in the updated version; updated mapping based on free text entered by study participants |
| Other veg e.g asparagus, aubergine, pumpkin cooked with added fat: Fat specified as soya/vegan/dairy free margarine (e.g Pure), low fat | veg_other | 60 | veg_other_marg_soya_lowfat | 60 | 0% | Fat (3%) used for cooking taken into account in the updated version; updated mapping based on free text entered by study participants |
| Other veg e.g asparagus, aubergine, pumpkin cooked with added fat: Fat specified as soya/vegan/dairy free margarine (e.g Pure), very low fat | veg_other | 60 | veg_other_marg_soya_vlowfat | 60 | 0% | Fat (3%) used for cooking taken into account in the updated version; updated mapping based on free text entered by study participants |
| Other veg e.g asparagus, aubergine, pumpkin cooked with added fat: Fat specified as olive oil | veg_other | 60 | veg_other_oil_olive | 60 | 0% | Fat (3%) used for cooking taken into account in the updated version; updated mapping based on free text entered by study participants |
| Other veg e.g asparagus, aubergine, pumpkin cooked with added fat: Fat specified as other oil e.g corn, groundnut, rice bran oil | veg_other | 60 | veg_other_oil_other | 60 | 0% | Fat (3%) used for cooking taken into account in the updated version; updated mapping based on free text entered by study participants |
| Other veg e.g asparagus, aubergine, pumpkin cooked with added fat: Fat specified as rapeseed oil | veg_other | 60 | veg_other_oil_rapeseed | 60 | 0% | Fat (3%) used for cooking taken into account in the updated version; updated mapping based on free text entered by study participants |
| Other veg e.g asparagus, aubergine, pumpkin cooked with added fat: Fat specified as sunflower oil | veg_other | 60 | veg_other_oil_sunflower | 60 | 0% | Fat (3%) used for cooking taken into account in the updated version; updated mapping based on free text entered by study participants |
| Other veg e.g asparagus, aubergine, pumpkin cooked with added fat: Fat specified as vegetable oil | veg_other | 60 | veg_other_oil_veg | 60 | 0% | Fat (3%) used for cooking taken into account in the updated version; updated mapping based on free text entered by study participants |
| Other veg e.g asparagus, aubergine, pumpkin cooked with added fat: Fat specified as dairy spread which is also cholesterol lowering e.g Benecol Buttery | veg_other | 60 | veg_other_spread_dairy_chol | 60 | 0% | Fat (3%) used for cooking taken into account in the updated version; updated mapping based on free text entered by study participants |
| Other veg e.g asparagus, aubergine, pumpkin cooked with added fat: Fat specified as dairy spread but not specified amount of % fat | veg_other | 60 | veg_other_spread_dairy_dunno | 60 | 0% | Fat (3%) used for cooking taken into account in the updated version; updated mapping based on free text entered by study participants |
| Other veg e.g asparagus, aubergine, pumpkin cooked with added fat: Fat specified as dairy spread with normal amount of % fat | veg_other | 60 | veg_other_spread_dairy_fat | 60 | 0% | Fat (3%) used for cooking taken into account in the updated version; updated mapping based on free text entered by study participants |
| Other veg e.g asparagus, aubergine, pumpkin cooked with added fat: Fat specified as dairy spread, low fat | veg_other | 60 | veg_other_spread_dairy_lowfat | 60 | 0% | Fat (3%) used for cooking taken into account in the updated version; updated mapping based on free text entered by study participants |
| Other veg e.g asparagus, aubergine, pumpkin cooked with added fat: Fat specified as dairy spread, very low fat | veg_other | 60 | veg_other_spread_dairy_vlowfat | 60 | 0% | Fat (3%) used for cooking taken into account in the updated version; updated mapping based on free text entered by study participants |
| Other veg e.g asparagus, aubergine, pumpkin cooked with added fat: Fat specified as not known type of spread but ticked cholesterol lowering e.g Benecol, Flora pro | veg_other | 60 | veg_other_spread_dunno_chol | 60 | 0% | Fat (3%) used for cooking taken into account in the updated version; updated mapping based on free text entered by study participants |
| Other veg e.g asparagus, aubergine, pumpkin cooked with added fat: Fat specified as not known type of spread or amount of % fat | veg_other | 60 | veg_other_spread_dunno_dunno | 60 | 0% | Fat (3%) used for cooking taken into account in the updated version; updated mapping based on free text entered by study participants |
| Other veg e.g asparagus, aubergine, pumpkin cooked with added fat: Fat specified as not known type of spread but ticked normal amount % fat | veg_other | 60 | veg_other_spread_dunno_fat | 60 | 0% | Fat (3%) used for cooking taken into account in the updated version; updated mapping based on free text entered by study participants |
| Other veg e.g asparagus, aubergine, pumpkin cooked with added fat: Fat specified as not known type of spread but ticked low fat | veg_other | 60 | veg_other_spread_dunno_lowfat | 60 | 0% | Fat (3%) used for cooking taken into account in the updated version; updated mapping based on free text entered by study participants |
| Other veg e.g asparagus, aubergine, pumpkin cooked with added fat: Fat specified as not known type of spread but ticked very low fat | veg_other | 60 | veg_other_spread_dunno_vlowfat | 60 | 0% | Fat (3%) used for cooking taken into account in the updated version; updated mapping based on free text entered by study participants |
| Other veg e.g asparagus, aubergine, pumpkin cooked with added fat: Fat specified as cholesterol lowering olive spread e.g Benecol/Flora pro active olive spread | veg_other | 60 | veg_other_spread_olive_chol | 60 | 0% | Fat (3%) used for cooking taken into account in the updated version; updated mapping based on free text entered by study participants |
| Other veg e.g asparagus, aubergine, pumpkin cooked with added fat: Fat specified as olive spread but not specified amount of fat | veg_other | 60 | veg_other_spread_olive_dunno | 60 | 0% | Fat (3%) used for cooking taken into account in the updated version; updated mapping based on free text entered by study participants |
| Other veg e.g asparagus, aubergine, pumpkin cooked with added fat: Fat specified as olive spread with normal amount of % fat | veg_other | 60 | veg_other_spread_olive_fat | 60 | 0% | Fat (3%) used for cooking taken into account in the updated version; updated mapping based on free text entered by study participants |
| Other veg e.g asparagus, aubergine, pumpkin cooked with added fat: Fat specified as olive spread, low fat | veg_other | 60 | veg_other_spread_olive_lowfat | 60 | 0% | Fat (3%) used for cooking taken into account in the updated version; updated mapping based on free text entered by study participants |
| Other veg e.g asparagus, aubergine, pumpkin cooked with added fat: Fat specified as olive spread, very low fat | veg_other | 60 | veg_other_spread_olive_vlowfat | 60 | 0% | Fat (3%) used for cooking taken into account in the updated version; updated mapping based on free text entered by study participants |
| Parsnips cooked with added fat: Fat specified as butter but not specified amount of % fat | veg_parsnip | 65 | veg_parsnip_butter_dunno | 65 | 0% | Fat (2.5%) used for cooking taken into account in the updated version - 50% cooked with fat |
| Parsnips cooked with added fat: Fat specified as butter, normal amount of % fat | veg_parsnip | 65 | veg_parsnip_butter_fat | 65 | 0% | Fat (2.5%) used for cooking taken into account in the updated version - 50% cooked with fat |
| Parsnips cooked with added fat: Fat specified as butter, low fat | veg_parsnip | 65 | veg_parsnip_butter_lowfat | 65 | 0% | Fat (2.5%) used for cooking taken into account in the updated version - 50% cooked with fat |
| Parsnips cooked with added fat: Fat specified as spreadable butter with normal amount of % fat | veg_parsnip | 65 | veg_parsnip_butter_spread_fat | 65 | 0% | Fat (2.5%) used for cooking taken into account in the updated version - 50% cooked with fat |
| Parsnips cooked with added fat: Fat specified as spreadable butter, low fat | veg_parsnip | 65 | veg_parsnip_butter_spread_lowfat | 65 | 0% | Fat (2.5%) used for cooking taken into account in the updated version - 50% cooked with fat |
| Parsnips cooked with added fat: Fat specified as not known type of fat or spread | veg_parsnip | 65 | veg_parsnip_fat_dunno | 65 | 0% | Fat (2.5%) used for cooking taken into account in the updated version - 50% cooked with fat |
| Parsnips cooked with no added fat | veg_parsnip | 65 | veg_parsnip_fat_none | 65 | 0% | Fat (2.5%) used for cooking taken into account in the updated version - 50% cooked with fat |
| Parsnips cooked with added fat: Fat specified as other type of fat or spread e.g ghee, dripping | veg_parsnip | 65 | veg_parsnip_fat_other | 65 | 0% | Fat (2.5%) used for cooking taken into account in the updated version - 50% cooked with fat |
| Parsnips cooked with added fat: Fat specified as lard | veg_parsnip | 65 | veg_parsnip_lard | 65 | 0% | Fat (2.5%) used for cooking taken into account in the updated version - 50% cooked with fat |
| Parsnips cooked with added fat: Fat specified as hard margarine (hard block margarine in wrapper) | veg_parsnip | 65 | veg_parsnip_marg_hard | 65 | 0% | Fat (2.5%) used for cooking taken into account in the updated version - 50% cooked with fat |
| Parsnips cooked with added fat: Fat specified as polyunsaturated margarine (e.g Flora) and also cholesterol lowering | veg_parsnip | 65 | veg_parsnip_marg_poly_chol | 65 | 0% | Fat (2.5%) used for cooking taken into account in the updated version - 50% cooked with fat |
| Parsnips cooked with added fat: Fat specified as polyunsaturated margarine (e.g Flora) but not specified amount of fat | veg_parsnip | 65 | veg_parsnip_marg_poly_dunno | 65 | 0% | Fat (2.5%) used for cooking taken into account in the updated version - 50% cooked with fat |
| Parsnips cooked with added fat: Fat specified as polyunsaturated margarine (e.g Flora), normal amount of % fat | veg_parsnip | 65 | veg_parsnip_marg_poly_fat | 65 | 0% | Fat (2.5%) used for cooking taken into account in the updated version - 50% cooked with fat |
| Parsnips cooked with added fat: Fat specified as polyunsaturated margarine (e.g Flora), low fat | veg_parsnip | 65 | veg_parsnip_marg_poly_lowfat | 65 | 0% | Fat (2.5%) used for cooking taken into account in the updated version - 50% cooked with fat |
| Parsnips cooked with added fat: Fat specified as polyunsaturated margarine (e.g Flora), very low fat | veg_parsnip | 65 | veg_parsnip_marg_poly_vlowfat | 65 | 0% | Fat (2.5%) used for cooking taken into account in the updated version - 50% cooked with fat |
| Parsnips cooked with added fat: Fat specified as soya/vegan/dairy free margarine e.g Pure, and also cholesterol lowering | veg_parsnip | 65 | veg_parsnip_marg_soya_chol | 65 | 0% | Fat (2.5%) used for cooking taken into account in the updated version - 50% cooked with fat |
| Parsnips cooked with added fat: Fat specified as soya/vegan/dairy free margarine e.g Pure, and not specified amount of fat | veg_parsnip | 65 | veg_parsnip_marg_soya_dunno | 65 | 0% | Fat (2.5%) used for cooking taken into account in the updated version - 50% cooked with fat |
| Parsnips cooked with added fat: Fat specified as soya/vegan/dairy free margarine (e.g Pure), normal amount % fat | veg_parsnip | 65 | veg_parsnip_marg_soya_fat | 65 | 0% | Fat (2.5%) used for cooking taken into account in the updated version - 50% cooked with fat |
| Parsnips cooked with added fat: Fat specified as soya/vegan/dairy free margarine (e.g Pure), low fat | veg_parsnip | 65 | veg_parsnip_marg_soya_lowfat | 65 | 0% | Fat (2.5%) used for cooking taken into account in the updated version - 50% cooked with fat |
| Parsnips cooked with added fat: Fat specified as soya/vegan/dairy free margarine (e.g Pure), very low fat | veg_parsnip | 65 | veg_parsnip_marg_soya_vlowfat | 65 | 0% | Fat (2.5%) used for cooking taken into account in the updated version - 50% cooked with fat |
| Parsnips cooked with added fat: Fat specified as olive oil | veg_parsnip | 65 | veg_parsnip_oil_olive | 65 | 0% | Fat (2.5%) used for cooking taken into account in the updated version - 50% cooked with fat |
| Parsnips cooked with added fat: Fat specified as other oil e.g corn, groundnut, rice bran oil | veg_parsnip | 65 | veg_parsnip_oil_other | 65 | 0% | Fat (2.5%) used for cooking taken into account in the updated version - 50% cooked with fat |
| Parsnips cooked with added fat: Fat specified as rapeseed oil | veg_parsnip | 65 | veg_parsnip_oil_rapeseed | 65 | 0% | Fat (2.5%) used for cooking taken into account in the updated version - 50% cooked with fat |
| Parsnips cooked with added fat: Fat specified as sunflower oil | veg_parsnip | 65 | veg_parsnip_oil_sunflower | 65 | 0% | Fat (2.5%) used for cooking taken into account in the updated version - 50% cooked with fat |
| Parsnips cooked with added fat: Fat specified as vegetable oil | veg_parsnip | 65 | veg_parsnip_oil_veg | 65 | 0% | Fat (2.5%) used for cooking taken into account in the updated version - 50% cooked with fat |
| Parsnips cooked with added fat: Fat specified as dairy spread which is also cholesterol lowering e.g Benecol Buttery | veg_parsnip | 65 | veg_parsnip_spread_dairy_chol | 65 | 0% | Fat (2.5%) used for cooking taken into account in the updated version - 50% cooked with fat |
| Parsnips cooked with added fat: Fat specified as dairy spread but not specified amount of % fat | veg_parsnip | 65 | veg_parsnip_spread_dairy_dunno | 65 | 0% | Fat (2.5%) used for cooking taken into account in the updated version - 50% cooked with fat |
| Parsnips cooked with added fat: Fat specified as dairy spread with normal amount of % fat | veg_parsnip | 65 | veg_parsnip_spread_dairy_fat | 65 | 0% | Fat (2.5%) used for cooking taken into account in the updated version - 50% cooked with fat |
| Parsnips cooked with added fat: Fat specified as dairy spread, low fat | veg_parsnip | 65 | veg_parsnip_spread_dairy_lowfat | 65 | 0% | Fat (2.5%) used for cooking taken into account in the updated version - 50% cooked with fat |
| Parsnips cooked with added fat: Fat specified as dairy spread, very low fat | veg_parsnip | 65 | veg_parsnip_spread_dairy_vlowfat | 65 | 0% | Fat (2.5%) used for cooking taken into account in the updated version - 50% cooked with fat |
| Parsnips cooked with added fat: Fat specified as not known type of spread but ticked cholesterol lowering e.g Benecol, Flora pro active | veg_parsnip | 65 | veg_parsnip_spread_dunno_chol | 65 | 0% | Fat (2.5%) used for cooking taken into account in the updated version - 50% cooked with fat |
| Parsnips cooked with added fat: Fat specified as not known type of spread or amount of % fat | veg_parsnip | 65 | veg_parsnip_spread_dunno_dunno | 65 | 0% | Fat (2.5%) used for cooking taken into account in the updated version - 50% cooked with fat |
| Parsnips cooked with added fat: Fat specified as not known type of spread but ticked normal amount % fat | veg_parsnip | 65 | veg_parsnip_spread_dunno_fat | 65 | 0% | Fat (2.5%) used for cooking taken into account in the updated version - 50% cooked with fat |
| Parsnips cooked with added fat: Fat specified as not known type of spread but ticked low fat | veg_parsnip | 65 | veg_parsnip_spread_dunno_lowfat | 65 | 0% | Fat (2.5%) used for cooking taken into account in the updated version - 50% cooked with fat |
| Parsnips cooked with added fat: Fat specified as not known type of spread but ticked very low fat | veg_parsnip | 65 | veg_parsnip_spread_dunno_vlowfat | 65 | 0% | Fat (2.5%) used for cooking taken into account in the updated version - 50% cooked with fat |
| Parsnips cooked with added fat: Fat specified as cholesterol lowering olive spread e.g Benecol/Flora pro active olive spread | veg_parsnip | 65 | veg_parsnip_spread_olive_chol | 65 | 0% | Fat (2.5%) used for cooking taken into account in the updated version - 50% cooked with fat |
| Parsnips cooked with added fat: Fat specified as olive spread but not specified amount of fat | veg_parsnip | 65 | veg_parsnip_spread_olive_dunno | 65 | 0% | Fat (2.5%) used for cooking taken into account in the updated version - 50% cooked with fat |
| Parsnips cooked with added fat: Fat specified as olive spread with normal amount of % fat | veg_parsnip | 65 | veg_parsnip_spread_olive_fat | 65 | 0% | Fat (2.5%) used for cooking taken into account in the updated version - 50% cooked with fat |
| Parsnips cooked with added fat: Fat specified as olive spread, low fat | veg_parsnip | 65 | veg_parsnip_spread_olive_lowfat | 65 | 0% | Fat (2.5%) used for cooking taken into account in the updated version - 50% cooked with fat |
| Parsnips cooked with added fat: Fat specified as olive spread, very low fat | veg_parsnip | 65 | veg_parsnip_spread_olive_vlowfat | 65 | 0% | Fat (2.5%) used for cooking taken into account in the updated version - 50% cooked with fat |

| | McCance and Widdowson's | | Nutrient databank + other changes | | | |
|---|---|---|---|---|---|---|
| Variable description | Food item | Portion size | Food item | Portion size | Portion diff% | Description of differences with previous version |
| Garden peas, frozen or canned | veg_peas | 65 | veg_peas | 65 | 0% | |
| Peppers (sweet) cooked with added fat: Fat specified as butter but not specified amount of % fat | veg_pepper_bell | 160 | veg_pepper_bell_butter_dunno | 133 | -17% | Fat (2.5%) used for cooking taken into account in the updated version - 50% cooked with fat + portion weight now excludes inedible part |
| Peppers (sweet) cooked with added fat: Fat specified as butter, normal amount of % fat | veg_pepper_bell | 160 | veg_pepper_bell_butter_fat | 133 | -17% | Fat (2.5%) used for cooking taken into account in the updated version - 50% cooked with fat + portion weight now excludes inedible part |
| Peppers (sweet) cooked with added fat: Fat specified as butter, low fat | veg_pepper_bell | 160 | veg_pepper_bell_butter_lowfat | 133 | -17% | Fat (2.5%) used for cooking taken into account in the updated version - 50% cooked with fat + portion weight now excludes inedible part |
| Peppers (sweet) cooked with added fat: Fat specified as spreadable butter with normal amount of % fat | veg_pepper_bell | 160 | veg_pepper_bell_butter_spread_fat | 133 | -17% | Fat (2.5%) used for cooking taken into account in the updated version - 50% cooked with fat + portion weight now excludes inedible part |
| Peppers (sweet) cooked with added fat: Fat specified as spreadable butter, low fat | veg_pepper_bell | 160 | veg_pepper_bell_butter_spread_lowfat | 133 | -17% | Fat (2.5%) used for cooking taken into account in the updated version - 50% cooked with fat + portion weight now excludes inedible part |
| Peppers (sweet) cooked with added fat: Fat specified as not known type of fat or spread | veg_pepper_bell | 160 | veg_pepper_bell_fat_dunno | 133 | -17% | Fat (2.5%) used for cooking taken into account in the updated version - 50% cooked with fat + portion weight now excludes inedible part |
| Peppers (sweet) cooked with no added fat | veg_pepper_bell | 160 | veg_pepper_bell_fat_none | 133 | -17% | Fat (2.5%) used for cooking taken into account in the updated version - 50% cooked with fat + portion weight now excludes inedible part |
| Peppers (sweet) cooked with added fat: Fat specified as other type of fat or spread e.g ghee, dripping | veg_pepper_bell | 160 | veg_pepper_bell_fat_other | 133 | -17% | Fat (2.5%) used for cooking taken into account in the updated version - 50% cooked with fat + portion weight now excludes inedible part |
| Peppers (sweet) cooked with added fat: Fat specified as lard | veg_pepper_bell | 160 | veg_pepper_bell_lard | 133 | -17% | Fat (2.5%) used for cooking taken into account in the updated version - 50% cooked with fat + portion weight now excludes inedible part |
| Peppers (sweet) cooked with added fat: Fat specified as hard margarine (hard block margarine in wrapper) | veg_pepper_bell | 160 | veg_pepper_bell_marg_hard | 133 | -17% | Fat (2.5%) used for cooking taken into account in the updated version - 50% cooked with fat + portion weight now excludes inedible part |
| Peppers (sweet) cooked with added fat: Fat specified as polyunsaturated margarine (e.g Flora) and also cholesterol lowering | veg_pepper_bell | 160 | veg_pepper_bell_marg_poly_chol | 133 | -17% | Fat (2.5%) used for cooking taken into account in the updated version - 50% cooked with fat + portion weight now excludes inedible part |
| Peppers (sweet) cooked with added fat: Fat specified as polyunsaturated margarine (e.g Flora) but not specified amount of fat | veg_pepper_bell | 160 | veg_pepper_bell_marg_poly_dunno | 133 | -17% | Fat (2.5%) used for cooking taken into account in the updated version - 50% cooked with fat + portion weight now excludes inedible part |
| Peppers (sweet) cooked with added fat: Fat specified as polyunsaturated margarine (e.g Flora), normal amount of % fat | veg_pepper_bell | 160 | veg_pepper_bell_marg_poly_fat | 133 | -17% | Fat (2.5%) used for cooking taken into account in the updated version - 50% cooked with fat + portion weight now excludes inedible part |
| Peppers (sweet) cooked with added fat: Fat specified as polyunsaturated margarine (e.g Flora), low fat | veg_pepper_bell | 160 | veg_pepper_bell_marg_poly_lowfat | 133 | -17% | Fat (2.5%) used for cooking taken into account in the updated version - 50% cooked with fat + portion weight now excludes inedible part |
| Peppers (sweet) cooked with added fat: Fat specified as polyunsaturated margarine (e.g Flora), very low fat | veg_pepper_bell | 160 | veg_pepper_bell_marg_poly_vlowfat | 133 | -17% | Fat (2.5%) used for cooking taken into account in the updated version - 50% cooked with fat + portion weight now excludes inedible part |
| Peppers (sweet) cooked with added fat: Fat specified as soya/vegan/dairy free margarine e.g Pure, and also cholesterol lowering | veg_pepper_bell | 160 | veg_pepper_bell_marg_soya_chol | 133 | -17% | Fat (2.5%) used for cooking taken into account in the updated version - 50% cooked with fat + portion weight now excludes inedible part |
| Peppers (sweet) cooked with added fat: Fat specified as soya/vegan/dairy free margarine e.g Pure, and not specified amount of fat | veg_pepper_bell | 160 | veg_pepper_bell_marg_soya_dunno | 133 | -17% | Fat (2.5%) used for cooking taken into account in the updated version - 50% cooked with fat + portion weight now excludes inedible part |
| Peppers (sweet) cooked with added fat: Fat specified as soya/vegan/dairy free margarine (e.g Pure), normal amount % fat | veg_pepper_bell | 160 | veg_pepper_bell_marg_soya_fat | 133 | -17% | Fat (2.5%) used for cooking taken into account in the updated version - 50% cooked with fat + portion weight now excludes inedible part |
| Peppers (sweet) cooked with added fat: Fat specified as soya/vegan/dairy free margarine (e.g Pure), low fat | veg_pepper_bell | 160 | veg_pepper_bell_marg_soya_lowfat | 133 | -17% | Fat (2.5%) used for cooking taken into account in the updated version - 50% cooked with fat + portion weight now excludes inedible part |
| Peppers (sweet) cooked with added fat: Fat specified as soya/vegan/dairy free margarine (e.g Pure), very low fat | veg_pepper_bell | 160 | veg_pepper_bell_marg_soya_vlowfat | 133 | -17% | Fat (2.5%) used for cooking taken into account in the updated version - 50% cooked with fat + portion weight now excludes inedible part |
| Peppers (sweet) cooked with added fat: Fat specified as olive oil | veg_pepper_bell | 160 | veg_pepper_bell_oil_olive | 133 | -17% | Fat (2.5%) used for cooking taken into account in the updated version - 50% cooked with fat + portion weight now excludes inedible part |
| Peppers (sweet) cooked with added fat: Fat specified as other oil e.g corn, groundnut, rice bran oil | veg_pepper_bell | 160 | veg_pepper_bell_oil_other | 133 | -17% | Fat (2.5%) used for cooking taken into account in the updated version - 50% cooked with fat + portion weight now excludes inedible part |
| Peppers (sweet) cooked with added fat: Fat specified as rapeseed oil | veg_pepper_bell | 160 | veg_pepper_bell_oil_rapeseed | 133 | -17% | Fat (2.5%) used for cooking taken into account in the updated version - 50% cooked with fat + portion weight now excludes inedible part |
| Peppers (sweet) cooked with added fat: Fat specified as sunflower oil | veg_pepper_bell | 160 | veg_pepper_bell_oil_sunflower | 133 | -17% | Fat (2.5%) used for cooking taken into account in the updated version - 50% cooked with fat + portion weight now excludes inedible part |
| Peppers (sweet) cooked with added fat: Fat specified as vegetable oil | veg_pepper_bell | 160 | veg_pepper_bell_oil_veg | 133 | -17% | Fat (2.5%) used for cooking taken into account in the updated version - 50% cooked with fat + portion weight now excludes inedible part |
| Peppers (sweet) cooked with added fat: Fat specified as dairy spread which is also cholesterol lowering e.g Benecol Buttery | veg_pepper_bell | 160 | veg_pepper_bell_spread_dairy_chol | 133 | -17% | Fat (2.5%) used for cooking taken into account in the updated version - 50% cooked with fat + portion weight now excludes inedible part |
| Peppers (sweet) cooked with added fat: Fat specified as dairy spread but not specified amount of % fat | veg_pepper_bell | 160 | veg_pepper_bell_spread_dairy_dunno | 133 | -17% | Fat (2.5%) used for cooking taken into account in the updated version - 50% cooked with fat + portion weight now excludes inedible part |
| Peppers (sweet) cooked with added fat: Fat specified as dairy spread with normal amount of % fat | veg_pepper_bell | 160 | veg_pepper_bell_spread_dairy_fat | 133 | -17% | Fat (2.5%) used for cooking taken into account in the updated version - 50% cooked with fat + portion weight now excludes inedible part |
| Peppers (sweet) cooked with added fat: Fat specified as dairy spread, low fat | veg_pepper_bell | 160 | veg_pepper_bell_spread_dairy_lowfat | 133 | -17% | Fat (2.5%) used for cooking taken into account in the updated version - 50% cooked with fat + portion weight now excludes inedible part |
| Peppers (sweet) cooked with added fat: Fat specified as dairy spread, very low fat | veg_pepper_bell | 160 | veg_pepper_bell_spread_dairy_vlowfat | 133 | -17% | Fat (2.5%) used for cooking taken into account in the updated version - 50% cooked with fat + portion weight now excludes inedible part |
| Peppers (sweet) cooked with added fat: Fat specified as not known type of spread but ticked cholesterol lowering e.g Benecol, Flora pro active | veg_pepper_bell | 160 | veg_pepper_bell_spread_dunno_chol | 133 | -17% | Fat (2.5%) used for cooking taken into account in the updated version - 50% cooked with fat + portion weight now excludes inedible part |
| Peppers (sweet) cooked with added fat: Fat specified as not known type of spread or amount of % fat | veg_pepper_bell | 160 | veg_pepper_bell_spread_dunno_dunno | 133 | -17% | Fat (2.5%) used for cooking taken into account in the updated version - 50% cooked with fat + portion weight now excludes inedible part |
| Peppers (sweet) cooked with added fat: Fat specified as not known type of spread but ticked normal amount % fat | veg_pepper_bell | 160 | veg_pepper_bell_spread_dunno_fat | 133 | -17% | Fat (2.5%) used for cooking taken into account in the updated version - 50% cooked with fat + portion weight now excludes inedible part |
| Peppers (sweet) cooked with added fat: Fat specified as not known type of spread but ticked low fat | veg_pepper_bell | 160 | veg_pepper_bell_spread_dunno_lowfat | 133 | -17% | Fat (2.5%) used for cooking taken into account in the updated version - 50% cooked with fat + portion weight now excludes inedible part |
| Peppers (sweet) cooked with added fat: Fat specified as not known type of spread but ticked very low fat | veg_pepper_bell | 160 | veg_pepper_bell_spread_dunno_vlowfat | 133 | -17% | Fat (2.5%) used for cooking taken into account in the updated version - 50% cooked with fat + portion weight now excludes inedible part |
| Peppers (sweet) cooked with added fat: Fat specified as cholesterol lowering olive spread e.g Benecol/Flora pro active olive spread | veg_pepper_bell | 160 | veg_pepper_bell_spread_olive_chol | 133 | -17% | Fat (2.5%) used for cooking taken into account in the updated version - 50% cooked with fat + portion weight now excludes inedible part |
| Peppers (sweet) cooked with added fat: Fat specified as olive spread but not specified amount of fat | veg_pepper_bell | 160 | veg_pepper_bell_spread_olive_dunno | 133 | -17% | Fat (2.5%) used for cooking taken into account in the updated version - 50% cooked with fat + portion weight now excludes inedible part |
| Peppers (sweet) cooked with added fat: Fat specified as olive spread with normal amount of % fat | veg_pepper_bell | 160 | veg_pepper_bell_spread_olive_fat | 133 | -17% | Fat (2.5%) used for cooking taken into account in the updated version - 50% cooked with fat + portion weight now excludes inedible part |
| Peppers (sweet) cooked with added fat: Fat specified as olive spread, low fat | veg_pepper_bell | 160 | veg_pepper_bell_spread_olive_lowfat | 133 | -17% | Fat (2.5%) used for cooking taken into account in the updated version - 50% cooked with fat + portion weight now excludes inedible part |
| Peppers (sweet) cooked with added fat: Fat specified as olive spread, very low fat | veg_pepper_bell | 160 | veg_pepper_bell_spread_olive_vlowfat | 133 | -17% | Fat (2.5%) used for cooking taken into account in the updated version - 50% cooked with fat + portion weight now excludes inedible part |
| Pulses e.g kidney beans, chick peas, butter beans or lentils | veg_pulses | 70 | veg_pulses | 70 | 0% | |
| Salad cream or mayonnaise | veg_saladmayo | 120 | veg_saladmayo | 120 | 0% | |
| Mixed side salad | veg_sidesalad | 66 | veg_sidesalad | 66 | 0% | |
| Spinach | veg_spinach | 90 | veg_spinach | 90 | 0% | |
| Sprouts | veg_sprouts | 90 | veg_sprouts | 90 | 0% | |
| Sweetcorn, corn on the cob | veg_sweetcorn | 43 | veg_sweetcorn | 43 | 0% | |
| Sweet potato | veg_sweetpot | 130 | veg_sweetpot | 130 | 0% | |
| Tomatoes fresh raw | veg_tomato_fresh | 85 | veg_tomato_fresh | 85 | 0% | |
| Tomatoes canned or cooked | veg_tomato_tinned | 135 | veg_tomato_tinned | 135 | 0% | |
| Turnips | veg_turnip | 60 | veg_turnip | 60 | 0% | |
| Watercress | veg_watercress | 20 | veg_watercress | 20 | 0% | |
| Yeast extract e.g Marmite, Vegemite | yeast_extract | 9 | yeast_extract | 4 | -56% | Weight now refects a thick spread (MAFF) |
| Yogurt full fat, plain or with fruit | yogurt_fullfat | 125 | yogurt_fullfat | 125 | 0% | |
| Yogurt low fat, plain or with fruit | yogurt_lowfat | 125 | yogurt_lowfat | 125 | 0% | |
| Yorkshire pudding | yorkshirepud | 80 | yorkshirepud | 25 | -69% | Portion size amended to reflect shop-bought muffin-sized unit |
