## Supplementary material for "Description of the updated nutrition calculation of the Oxford WebQ questionnaire and comparison with the previous version among 207,144 participants in UK Biobank": Sup table 2

**Supplementary table 2**. Nutrient calculation in the updated version (Nutrient databank).

| | Percentage of the total | | | | | Food codes from UK Nutrient Databank and the % used from each food code | | | | | | | | | | | | | | | | | | | | | |
|---|---|---|---|---|---|---|---|---|---|---|---|---|---|---|---|---|---|---|---|---|---|---|---|---|---|---|---|
| Food item | Free sugars | Plant prot | Animal prot | Plant fat | Animal fat | Code1 | % | Code2 | % | Code3 | % | Code4 | % | Code5 | % | Code6 | % | Code7 | % | Code8 | % | code9 | % | code10 | % | code11 | % |
| add_salt | 0% | 100% | 0% | 100% | 0% | 2522 | 100 | | | | | | | | | | | | | | | | | | | | |
| alcohol_beercider | 100% | 100% | 0% | 100% | 0% | 2374 | 35 | 2362 | 35 | 2379 | 6 | 8350 | 6 | 8349 | 6 | 2380 | 6 | 2380 | 6 | | | | | | | | |
| alcohol_other | 97% | 100% | 0% | 1% | 99% | 2397 | 33.4 | 2399 | 33.3 | 2400 | 33.3 | | | | | | | | | | | | | | | | |
| alcohol_spirits | 0% | 100% | 0% | 100% | 0% | 2402 | 100 | | | | | | | | | | | | | | | | | | | | |
| alcohol_wine_fort | 100% | 100% | 0% | 100% | 0% | 2391 | 16.7 | 2392 | 16.7 | 2393 | 16.6 | 2390 | 50 | | | | | | | | | | | | | | |
| alcohol_wine_red_large | 100% | 100% | 0% | 100% | 0% | 8352 | 100 | | | | | | | | | | | | | | | | | | | | |
| alcohol_wine_red_med | 100% | 100% | 0% | 100% | 0% | 8352 | 100 | | | | | | | | | | | | | | | | | | | | |
| alcohol_wine_red_small | 100% | 100% | 0% | 100% | 0% | 8352 | 100 | | | | | | | | | | | | | | | | | | | | |
| alcohol_wine_rose_large | 100% | 100% | 0% | 100% | 0% | 8353 | 100 | | | | | | | | | | | | | | | | | | | | |
| alcohol_wine_rose_med | 100% | 100% | 0% | 100% | 0% | 8353 | 100 | | | | | | | | | | | | | | | | | | | | |
| alcohol_wine_rose_small | 100% | 100% | 0% | 100% | 0% | 8353 | 100 | | | | | | | | | | | | | | | | | | | | |
| alcohol_wine_white_large | 100% | 100% | 0% | 100% | 0% | 8354 | 25 | 8355 | 25 | 8357 | 25 | 8356 | 25 | | | | | | | | | | | | | | |
| alcohol_wine_white_med | 100% | 100% | 0% | 100% | 0% | 8354 | 25 | 8355 | 25 | 8357 | 25 | 8356 | 25 | | | | | | | | | | | | | | |
| alcohol_wine_white_small | 100% | 100% | 0% | 100% | 0% | 8354 | 25 | 8355 | 25 | 8357 | 25 | 8356 | 25 | | | | | | | | | | | | | | |
| biscuit_choc | 96% | 30% | 70% | 50% | 50% | 7662 | 25 | 260 | 50 | 10516 | 25 | | | | | | | | | | | | | | | | |
| biscuit_choc_gf | 96% | 31% | 69% | 50% | 50% | 7662 | 25 | 260 | 50 | 10516 | 25 | | | | | | | | | | | | | | | | |
| biscuit_choccov | 98% | 30% | 70% | 50% | 50% | 8193 | 34 | 253 | 33 | 8194 | 33 | | | | | | | | | | | | | | | | |
| biscuit_choccov_gf | 98% | 31% | 69% | 50% | 50% | 8193 | 34 | 253 | 33 | 8194 | 33 | | | | | | | | | | | | | | | | |
| biscuit_sweet | 92% | 60% | 40% | 60% | 40% | 259 | 33.4 | 263 | 33.3 | 8162 | 33.3 | | | | | | | | | | | | | | | | |
| biscuit_sweet_gf | 100% | 61% | 39% | 60% | 40% | 8872 | 100 | | | | | | | | | | | | | | | | | | | | |
| bread_baguette_gf_nonwhite | 1% | 99% | 1% | 100% | 0% | 10459 | 50 | 8394 | 50 | | | | | | | | | | | | | | | | | | |
| bread_baguette_gf_unanswered | 1% | 90% | 10% | 100% | 0% | 8864 | 33.4 | 10459 | 33.3 | 8395 | 33.3 | | | | | | | | | | | | | | | | |
| bread_baguette_gf_white | 1% | 90% | 10% | 100% | 0% | 8864 | 100 | | | | | | | | | | | | | | | | | | | | |
| bread_baguette_mixed | 0% | 100% | 0% | 100% | 0% | 112 | 33.4 | 102 | 33.3 | 7609 | 16.7 | 10775 | 16.6 | | | | | | | | | | | | | | |
| bread_baguette_other | 0% | 100% | 0% | 100% | 0% | 110 | 20 | 114 | 20 | 120 | 60 | | | | | | | | | | | | | | | | |
| bread_baguette_seeded | 0% | 100% | 0% | 100% | 0% | 2168 | 33.4 | 2167 | 33.3 | 8148 | 33.3 | | | | | | | | | | | | | | | | |
| bread_baguette_spread_butter_dunno_med | 0% | 20% | 80% | 20% | 80% | 9407 | 25 | 10039 | 25 | 851 | 25 | 852 | 25 | | | | | | | | | | | | | | |
| bread_baguette_spread_butter_dunno_thick | 0% | 20% | 80% | 20% | 80% | 9407 | 25 | 10039 | 25 | 851 | 25 | 852 | 25 | | | | | | | | | | | | | | |
| bread_baguette_spread_butter_dunno_thin | 0% | 20% | 80% | 20% | 80% | 9407 | 25 | 10039 | 25 | 851 | 25 | 852 | 25 | | | | | | | | | | | | | | |
| bread_baguette_spread_butter_fat_med | 0% | 0% | 100% | 0% | 100% | 851 | 50 | 852 | 50 | | | | | | | | | | | | | | | | | | |
| bread_baguette_spread_butter_fat_thick | 0% | 0% | 100% | 0% | 100% | 851 | 50 | 852 | 50 | | | | | | | | | | | | | | | | | | |
| bread_baguette_spread_butter_fat_thin | 0% | 0% | 100% | 0% | 100% | 851 | 50 | 852 | 50 | | | | | | | | | | | | | | | | | | |
| bread_baguette_spread_butter_lowfat_med | 0% | 45% | 55% | 45% | 55% | 10140 | 100 | | | | | | | | | | | | | | | | | | | | |
| bread_baguette_spread_butter_lowfat_thick | 0% | 45% | 55% | 45% | 55% | 10140 | 100 | | | | | | | | | | | | | | | | | | | | |
| bread_baguette_spread_butter_lowfat_thin | 0% | 45% | 55% | 45% | 55% | 10140 | 100 | | | | | | | | | | | | | | | | | | | | |
| bread_baguette_spread_butter_spread_fat_med | 0% | 40% | 60% | 40% | 60% | 9407 | 50 | 10039 | 50 | | | | | | | | | | | | | | | | | | |
| bread_baguette_spread_butter_spread_fat_thick | 0% | 40% | 60% | 40% | 60% | 9407 | 50 | 10039 | 50 | | | | | | | | | | | | | | | | | | |
| bread_baguette_spread_butter_spread_fat_thin | 0% | 40% | 60% | 40% | 60% | 9407 | 50 | 10039 | 50 | | | | | | | | | | | | | | | | | | |
| bread_baguette_spread_butter_spread_lowfat_med | 0% | 45% | 55% | 45% | 55% | 3891 | 50 | 10894 | 50 | | | | | | | | | | | | | | | | | | |
| bread_baguette_spread_butter_spread_lowfat_thick | 0% | 45% | 55% | 45% | 55% | 3891 | 50 | 10894 | 50 | | | | | | | | | | | | | | | | | | |
| bread_baguette_spread_butter_spread_lowfat_thin | 0% | 45% | 55% | 45% | 55% | 3891 | 50 | 10894 | 50 | | | | | | | | | | | | | | | | | | |
| bread_baguette_spread_dairy_chol_med | 0% | 97% | 3% | 97% | 3% | 3848 | 100 | | | | | | | | | | | | | | | | | | | | |
| bread_baguette_spread_dairy_chol_thick | 0% | 97% | 3% | 97% | 3% | 3848 | 100 | | | | | | | | | | | | | | | | | | | | |
| bread_baguette_spread_dairy_chol_thin | 0% | 97% | 3% | 97% | 3% | 3848 | 100 | | | | | | | | | | | | | | | | | | | | |
| bread_baguette_spread_dairy_dunno_med | 0% | 97% | 3% | 97% | 3% | 10047 | 50 | 7775 | 50 | | | | | | | | | | | | | | | | | | |
| bread_baguette_spread_dairy_dunno_thick | 0% | 97% | 3% | 97% | 3% | 10047 | 50 | 7775 | 50 | | | | | | | | | | | | | | | | | | |
| bread_baguette_spread_dairy_dunno_thin | 0% | 97% | 3% | 97% | 3% | 10047 | 50 | 7775 | 50 | | | | | | | | | | | | | | | | | | |
| bread_baguette_spread_dairy_fat_med | 0% | 97% | 3% | 97% | 3% | 7775 | 100 | | | | | | | | | | | | | | | | | | | | |
| bread_baguette_spread_dairy_fat_thick | 0% | 97% | 3% | 97% | 3% | 7775 | 100 | | | | | | | | | | | | | | | | | | | | |
| bread_baguette_spread_dairy_fat_thin | 0% | 97% | 3% | 97% | 3% | 7775 | 100 | | | | | | | | | | | | | | | | | | | | |
| bread_baguette_spread_dairy_lowfat_med | 0% | 99% | 1% | 99% | 1% | 10047 | 100 | | | | | | | | | | | | | | | | | | | | |
| bread_baguette_spread_dairy_lowfat_thick | 0% | 99% | 1% | 99% | 1% | 10047 | 100 | | | | | | | | | | | | | | | | | | | | |
| bread_baguette_spread_dairy_lowfat_thin | 0% | 99% | 1% | 99% | 1% | 10047 | 100 | | | | | | | | | | | | | | | | | | | | |
| bread_baguette_spread_dairy_vlowfat_med | 0% | 99% | 1% | 99% | 1% | 10047 | 100 | | | | | | | | | | | | | | | | | | | | |
| bread_baguette_spread_dairy_vlowfat_thick | 0% | 99% | 1% | 99% | 1% | 10047 | 100 | | | | | | | | | | | | | | | | | | | | |
| bread_baguette_spread_dairy_vlowfat_thin | 0% | 99% | 1% | 99% | 1% | 10047 | 100 | | | | | | | | | | | | | | | | | | | | |
| bread_baguette_spread_dunno_chol_med | 0% | 97% | 3% | 97% | 3% | 3848 | 33.4 | 3243 | 33.3 | 2849 | 33.3 | | | | | | | | | | | | | | | | |
| bread_baguette_spread_dunno_chol_thick | 0% | 97% | 3% | 97% | 3% | 3848 | 33.4 | 3243 | 33.3 | 2849 | 33.3 | | | | | | | | | | | | | | | | |
| bread_baguette_spread_dunno_chol_thin | 0% | 97% | 3% | 97% | 3% | 3848 | 33.4 | 3243 | 33.3 | 2849 | 33.3 | | | | | | | | | | | | | | | | |
| bread_baguette_spread_dunno_dunno_med | 0% | 97% | 3% | 97% | 3% | 10047 | 25 | 10049 | 25 | 10043 | 25 | 7775 | 25 | | | | | | | | | | | | | | |
| bread_baguette_spread_dunno_dunno_thick | 0% | 97% | 3% | 97% | 3% | 10047 | 25 | 10049 | 25 | 10043 | 25 | 7775 | 25 | | | | | | | | | | | | | | |
| bread_baguette_spread_dunno_dunno_thin | 0% | 97% | 3% | 97% | 3% | 10047 | 25 | 10049 | 25 | 10043 | 25 | 7775 | 25 | | | | | | | | | | | | | | |
| bread_baguette_spread_dunno_fat_med | 0% | 97% | 3% | 97% | 3% | 10043 | 50 | 7775 | 50 | | | | | | | | | | | | | | | | | | |
| bread_baguette_spread_dunno_fat_thick | 0% | 97% | 3% | 97% | 3% | 10043 | 50 | 7775 | 50 | | | | | | | | | | | | | | | | | | |
| bread_baguette_spread_dunno_fat_thin | 0% | 97% | 3% | 97% | 3% | 10043 | 50 | 7775 | 50 | | | | | | | | | | | | | | | | | | |
| bread_baguette_spread_dunno_lowfat_med | 0% | 97% | 3% | 97% | 3% | 10047 | 50 | 10049 | 50 | | | | | | | | | | | | | | | | | | |
| bread_baguette_spread_dunno_lowfat_thick | 0% | 97% | 3% | 97% | 3% | 10047 | 50 | 10049 | 50 | | | | | | | | | | | | | | | | | | |
| bread_baguette_spread_dunno_lowfat_thin | 0% | 97% | 3% | 97% | 3% | 10047 | 50 | 10049 | 50 | | | | | | | | | | | | | | | | | | |
| bread_baguette_spread_dunno_vlowfat_med | 0% | 97% | 3% | 97% | 3% | 10047 | 50 | 10049 | 50 | | | | | | | | | | | | | | | | | | |
| bread_baguette_spread_dunno_vlowfat_thick | 0% | 97% | 3% | 97% | 3% | 10047 | 50 | 10049 | 50 | | | | | | | | | | | | | | | | | | |
| bread_baguette_spread_dunno_vlowfat_thin | 0% | 97% | 3% | 97% | 3% | 10047 | 50 | 10049 | 50 | | | | | | | | | | | | | | | | | | |
| bread_baguette_spread_hardmarg_med | 0% | 100% | 0% | 100% | 0% | 860 | 100 | | | | | | | | | | | | | | | | | | | | |
| bread_baguette_spread_hardmarg_thick | 0% | 100% | 0% | 100% | 0% | 860 | 100 | | | | | | | | | | | | | | | | | | | | |
| bread_baguette_spread_hardmarg_thin | 0% | 100% | 0% | 100% | 0% | 860 | 100 | | | | | | | | | | | | | | | | | | | | |
| bread_baguette_spread_olive_chol_med | 0% | 99% | 1% | 99% | 1% | 3364 | 50 | 10053 | 50 | | | | | | | | | | | | | | | | | | |
| bread_baguette_spread_olive_chol_thick | 0% | 99% | 1% | 99% | 1% | 3364 | 50 | 10053 | 50 | | | | | | | | | | | | | | | | | | |
| bread_baguette_spread_olive_chol_thin | 0% | 99% | 1% | 99% | 1% | 3364 | 50 | 10053 | 50 | | | | | | | | | | | | | | | | | | |
| bread_baguette_spread_olive_dunno_med | 0% | 99% | 1% | 99% | 1% | 10048 | 50 | 10042 | 25 | 10131 | 25 | | | | | | | | | | | | | | | | |
| bread_baguette_spread_olive_dunno_thick | 0% | 99% | 1% | 99% | 1% | 10048 | 50 | 10042 | 25 | 10131 | 25 | | | | | | | | | | | | | | | | |
| bread_baguette_spread_olive_dunno_thin | 0% | 99% | 1% | 99% | 1% | 10048 | 50 | 10042 | 25 | 10131 | 25 | | | | | | | | | | | | | | | | |
| bread_baguette_spread_olive_fat_med | 0% | 99% | 1% | 99% | 1% | 10042 | 75 | 10131 | 25 | | | | | | | | | | | | | | | | | | |
| bread_baguette_spread_olive_fat_thick | 0% | 99% | 1% | 99% | 1% | 10042 | 75 | 10131 | 25 | | | | | | | | | | | | | | | | | | |
| bread_baguette_spread_olive_fat_thin | 0% | 99% | 1% | 99% | 1% | 10042 | 75 | 10131 | 25 | | | | | | | | | | | | | | | | | | |
| bread_baguette_spread_olive_lowfat_med | 0% | 100% | 1% | 100% | 0% | 10048 | 100 | | | | | | | | | | | | | | | | | | | | |
| bread_baguette_spread_olive_lowfat_thick | 0% | 100% | 1% | 100% | 0% | 10048 | 100 | | | | | | | | | | | | | | | | | | | | |
| bread_baguette_spread_olive_lowfat_thin | 0% | 100% | 1% | 100% | 0% | 10048 | 100 | | | | | | | | | | | | | | | | | | | | |
| bread_baguette_spread_olive_vlowfat_med | 0% | 100% | 1% | 100% | 0% | 10048 | 100 | | | | | | | | | | | | | | | | | | | | |
| bread_baguette_spread_olive_vlowfat_thick | 0% | 100% | 1% | 100% | 0% | 10048 | 100 | | | | | | | | | | | | | | | | | | | | |
| bread_baguette_spread_olive_vlowfat_thin | 0% | 100% | 1% | 100% | 0% | 10048 | 100 | | | | | | | | | | | | | | | | | | | | |
| bread_baguette_spread_other_med | 0% | 60% | 40% | 60% | 40% | 856 | 18 | 851 | 7.5 | 852 | 7.5 | 3848 | 33.5 | 2849 | 33.5 | | | | | | | | | | | | |
| bread_baguette_spread_other_thick | 0% | 60% | 40% | 60% | 40% | 856 | 18 | 851 | 7.5 | 852 | 7.5 | 3848 | 33.5 | 2849 | 33.5 | | | | | | | | | | | | |
| bread_baguette_spread_other_thin | 0% | 60% | 40% | 60% | 40% | 856 | 18 | 851 | 7.5 | 852 | 7.5 | 3848 | 33.5 | 2849 | 33.5 | | | | | | | | | | | | |
| bread_baguette_spread_polymarg_chol_med | 0% | 100% | 0% | 100% | 0% | 2849 | 33.4 | 3848 | 33.3 | 3243 | 33.3 | | | | | | | | | | | | | | | | |
| bread_baguette_spread_polymarg_chol_thick | 0% | 100% | 0% | 100% | 0% | 2849 | 33.4 | 3848 | 33.3 | 3243 | 33.3 | | | | | | | | | | | | | | | | |
| bread_baguette_spread_polymarg_chol_thin | 0% | 100% | 0% | 100% | 0% | 2849 | 33.4 | 3848 | 33.3 | 3243 | 33.3 | | | | | | | | | | | | | | | | |
| bread_baguette_spread_polymarg_dunno_med | 0% | 100% | 0% | 100% | 0% | 10049 | 50 | 10043 | 25 | 10044 | 25 | | | | | | | | | | | | | | | | |
| bread_baguette_spread_polymarg_dunno_thick | 0% | 100% | 0% | 100% | 0% | 10049 | 50 | 10043 | 25 | 10044 | 25 | | | | | | | | | | | | | | | | |
| bread_baguette_spread_polymarg_dunno_thin | 0% | 100% | 0% | 100% | 0% | 10049 | 50 | 10043 | 25 | 10044 | 25 | | | | | | | | | | | | | | | | |
| bread_baguette_spread_polymarg_fat_med | 0% | 100% | 0% | 100% | 0% | 10043 | 50 | 10044 | 50 | | | | | | | | | | | | | | | | | | |
| bread_baguette_spread_polymarg_fat_thick | 0% | 100% | 0% | 100% | 0% | 10043 | 50 | 10044 | 50 | | | | | | | | | | | | | | | | | | |
| bread_baguette_spread_polymarg_fat_thin | 0% | 100% | 0% | 100% | 0% | 10043 | 50 | 10044 | 50 | | | | | | | | | | | | | | | | | | |
| bread_baguette_spread_polymarg_lowfat_med | 0% | 100% | 0% | 100% | 0% | 10049 | 100 | | | | | | | | | | | | | | | | | | | | |
| bread_baguette_spread_polymarg_lowfat_thick | 0% | 100% | 0% | 100% | 0% | 10049 | 100 | | | | | | | | | | | | | | | | | | | | |
| bread_baguette_spread_polymarg_lowfat_thin | 0% | 100% | 0% | 100% | 0% | 10049 | 100 | | | | | | | | | | | | | | | | | | | | |
| bread_baguette_spread_polymarg_vlowfat_med | 0% | 100% | 0% | 100% | 0% | 10049 | 100 | | | | | | | | | | | | | | | | | | | | |
| bread_baguette_spread_polymarg_vlowfat_thick | 0% | 100% | 0% | 100% | 0% | 10049 | 100 | | | | | | | | | | | | | | | | | | | | |
| bread_baguette_spread_polymarg_vlowfat_thin | 0% | 100% | 0% | 100% | 0% | 10049 | 100 | | | | | | | | | | | | | | | | | | | | |
| bread_baguette_spread_soya_chol_med | 0% | 100% | 1% | 100% | 1% | 3848 | 16.6 | 3243 | 16.6 | 2849 | 16.8 | 10786 | 50 | | | | | | | | | | | | | | |
| bread_baguette_spread_soya_chol_thick | 0% | 100% | 1% | 100% | 1% | 3848 | 16.6 | 3243 | 16.6 | 2849 | 16.8 | 10786 | 50 | | | | | | | | | | | | | | |
| bread_baguette_spread_soya_chol_thin | 0% | 100% | 1% | 100% | 1% | 3848 | 16.6 | 3243 | 16.6 | 2849 | 16.8 | 10786 | 50 | | | | | | | | | | | | | | |
| bread_baguette_spread_soya_dunno_med | 0% | 100% | 0% | 100% | 0% | 10980 | 50 | 10786 | 50 | | | | | | | | | | | | | | | | | | |
| bread_baguette_spread_soya_dunno_thick | 0% | 100% | 0% | 100% | 0% | 10980 | 50 | 10786 | 50 | | | | | | | | | | | | | | | | | | |
| bread_baguette_spread_soya_dunno_thin | 0% | 100% | 0% | 100% | 0% | 10980 | 50 | 10786 | 50 | | | | | | | | | | | | | | | | | | |
| bread_baguette_spread_soya_fat_med | 0% | 100% | 0% | 100% | 0% | 10786 | 100 | | | | | | | | | | | | | | | | | | | | |
| bread_baguette_spread_soya_fat_thick | 0% | 100% | 0% | 100% | 0% | 10786 | 100 | | | | | | | | | | | | | | | | | | | | |
| bread_baguette_spread_soya_fat_thin | 0% | 100% | 0% | 100% | 0% | 10786 | 100 | | | | | | | | | | | | | | | | | | | | |
| bread_baguette_spread_soya_lowfat_med | 0% | 100% | 0% | 100% | 0% | 10980 | 100 | | | | | | | | | | | | | | | | | | | | |
| bread_baguette_spread_soya_lowfat_thick | 0% | 100% | 0% | 100% | 0% | 10980 | 100 | | | | | | | | | | | | | | | | | | | | |
| bread_baguette_spread_soya_lowfat_thin | 0% | 100% | 0% | 100% | 0% | 10980 | 100 | | | | | | | | | | | | | | | | | | | | |
| bread_baguette_spread_soya_vlowfat_med | 0% | 100% | 0% | 100% | 0% | 10980 | 100 | | | | | | | | | | | | | | | | | | | | |
| bread_baguette_spread_soya_vlowfat_thick | 0% | 100% | 0% | 100% | 0% | 10980 | 100 | | | | | | | | | | | | | | | | | | | | |
| bread_baguette_spread_soya_vlowfat_thin | 0% | 100% | 0% | 100% | 0% | 10980 | 100 | | | | | | | | | | | | | | | | | | | | |
| bread_baguette_unanswered | 0% | 100% | 0% | 100% | 0% | 120 | 33.4 | 133 | 33.3 | 102 | 33.3 | | | | | | | | | | | | | | | | |
| bread_baguette_white | 0% | 100% | 0% | 100% | 0% | 127 | 50 | 6977 | 50 | | | | | | | | | | | | | | | | | | |
| bread_baguette_wholemeal | 0% | 100% | 0% | 100% | 0% | 133 | 100 | | | | | | | | | | | | | | | | | | | | |
| bread_crisp | 0% | 100% | 0% | 100% | 0% | 256 | 30 | 255 | 30 | 7652 | 30 | 3267 | 10 | | | | | | | | | | | | | | |
| bread_crisp_gf | 0% | 100% | 0% | 100% | 0% | 256 | 30 | 255 | 30 | 7652 | 30 | 3267 | 10 | | | | | | | | | | | | | | |
| bread_crisp_spread_butter_dunno_med | 0% | 20% | 80% | 20% | 80% | 9407 | 25 | 10039 | 25 | 851 | 25 | 852 | 25 | | | | | | | | | | | | | | |
| bread_crisp_spread_butter_dunno_thick | 0% | 20% | 80% | 20% | 80% | 9407 | 25 | 10039 | 25 | 851 | 25 | 852 | 25 | | | | | | | | | | | | | | |
| bread_crisp_spread_butter_dunno_thin | 0% | 20% | 80% | 20% | 80% | 9407 | 25 | 10039 | 25 | 851 | 25 | 852 | 25 | | | | | | | | | | | | | | |
| bread_crisp_spread_butter_fat_med | 0% | 0% | 100% | 0% | 100% | 851 | 50 | 852 | 50 | | | | | | | | | | | | | | | | | | |
| bread_crisp_spread_butter_fat_thick | 0% | 0% | 100% | 0% | 100% | 851 | 50 | 852 | 50 | | | | | | | | | | | | | | | | | | |
| bread_crisp_spread_butter_fat_thin | 0% | 0% | 100% | 0% | 100% | 851 | 50 | 852 | 50 | | | | | | | | | | | | | | | | | | |
| bread_crisp_spread_butter_lowfat_med | 0% | 45% | 55% | 45% | 55% | 10140 | 100 | | | | | | | | | | | | | | | | | | | | |
| bread_crisp_spread_butter_lowfat_thick | 0% | 45% | 55% | 45% | 55% | 10140 | 100 | | | | | | | | | | | | | | | | | | | | |
| bread_crisp_spread_butter_lowfat_thin | 0% | 45% | 55% | 45% | 55% | 10140 | 100 | | | | | | | | | | | | | | | | | | | | |
| bread_crisp_spread_butter_spread_fat_med | 0% | 40% | 60% | 40% | 60% | 9407 | 50 | 10039 | 50 | | | | | | | | | | | | | | | | | | |
| bread_crisp_spread_butter_spread_fat_thick | 0% | 40% | 60% | 40% | 60% | 9407 | 50 | 10039 | 50 | | | | | | | | | | | | | | | | | | |
| bread_crisp_spread_butter_spread_fat_thin | 0% | 40% | 60% | 40% | 60% | 9407 | 50 | 10039 | 50 | | | | | | | | | | | | | | | | | | |
| bread_crisp_spread_butter_spread_lowfat_med | 0% | 45% | 55% | 45% | 55% | 3891 | 50 | 10894 | 50 | | | | | | | | | | | | | | | | | | |
| bread_crisp_spread_butter_spread_lowfat_thick | 0% | 45% | 55% | 45% | 55% | 3891 | 50 | 10894 | 50 | | | | | | | | | | | | | | | | | | |
| bread_crisp_spread_butter_spread_lowfat_thin | 0% | 45% | 55% | 45% | 55% | 3891 | 50 | 10894 | 50 | | | | | | | | | | | | | | | | | | |
| bread_crisp_spread_dairy_chol_med | 0% | 97% | 3% | 97% | 3% | 3848 | 100 | | | | | | | | | | | | | | | | | | | | |
| bread_crisp_spread_dairy_chol_thick | 0% | 97% | 3% | 97% | 3% | 3848 | 100 | | | | | | | | | | | | | | | | | | | | |
| bread_crisp_spread_dairy_chol_thin | 0% | 97% | 3% | 97% | 3% | 3848 | 100 | | | | | | | | | | | | | | | | | | | | |
| bread_crisp_spread_dairy_dunno_med | 0% | 97% | 3% | 97% | 3% | 10047 | 50 | 7775 | 50 | | | | | | | | | | | | | | | | | | |
| bread_crisp_spread_dairy_dunno_thick | 0% | 97% | 3% | 97% | 3% | 10047 | 50 | 7775 | 50 | | | | | | | | | | | | | | | | | | |
| bread_crisp_spread_dairy_dunno_thin | 0% | 97% | 3% | 97% | 3% | 10047 | 50 | 7775 | 50 | | | | | | | | | | | | | | | | | | |
| bread_crisp_spread_dairy_fat_med | 0% | 97% | 3% | 97% | 3% | 7775 | 100 | | | | | | | | | | | | | | | | | | | | |
| bread_crisp_spread_dairy_fat_thick | 0% | 97% | 3% | 97% | 3% | 7775 | 100 | | | | | | | | | | | | | | | | | | | | |
| bread_crisp_spread_dairy_fat_thin | 0% | 97% | 3% | 97% | 3% | 7775 | 100 | | | | | | | | | | | | | | | | | | | | |
| bread_crisp_spread_dairy_lowfat_med | 0% | 99% | 1% | 99% | 1% | 10047 | 100 | | | | | | | | | | | | | | | | | | | | |
| bread_crisp_spread_dairy_lowfat_thick | 0% | 99% | 1% | 99% | 1% | 10047 | 100 | | | | | | | | | | | | | | | | | | | | |
| bread_crisp_spread_dairy_lowfat_thin | 0% | 99% | 1% | 99% | 1% | 10047 | 100 | | | | | | | | | | | | | | | | | | | | |
| bread_crisp_spread_dairy_vlowfat_med | 0% | 99% | 1% | 99% | 1% | 10047 | 100 | | | | | | | | | | | | | | | | | | | | |
| bread_crisp_spread_dairy_vlowfat_thick | 0% | 99% | 1% | 99% | 1% | 10047 | 100 | | | | | | | | | | | | | | | | | | | | |
| bread_crisp_spread_dairy_vlowfat_thin | 0% | 99% | 1% | 99% | 1% | 10047 | 100 | | | | | | | | | | | | | | | | | | | | |
| bread_crisp_spread_dunno_chol_med | 0% | 97% | 3% | 97% | 3% | 3848 | 33.4 | 3243 | 33.3 | 2849 | 33.3 | | | | | | | | | | | | | | | | |
| bread_crisp_spread_dunno_chol_thick | 0% | 97% | 3% | 97% | 3% | 3848 | 33.4 | 3243 | 33.3 | 2849 | 33.3 | | | | | | | | | | | | | | | | |
| bread_crisp_spread_dunno_chol_thin | 0% | 97% | 3% | 97% | 3% | 3848 | 33.4 | 3243 | 33.3 | 2849 | 33.3 | | | | | | | | | | | | | | | | |
| bread_crisp_spread_dunno_dunno_med | 0% | 97% | 3% | 97% | 3% | 10047 | 25 | 10049 | 25 | 10043 | 25 | 7775 | 25 | | | | | | | | | | | | | | |
| bread_crisp_spread_dunno_dunno_thick | 0% | 97% | 3% | 97% | 3% | 10047 | 25 | 10049 | 25 | 10043 | 25 | 7775 | 25 | | | | | | | | | | | | | | |
| bread_crisp_spread_dunno_dunno_thin | 0% | 97% | 3% | 97% | 3% | 10047 | 25 | 10049 | 25 | 10043 | 25 | 7775 | 25 | | | | | | | | | | | | | | |

| | | | | | | | | | | | | | | | |
|---|---|---|---|---|---|---|---|---|---|---|---|---|---|---|---|
| bread_crisp_spread_dunno_fat_med | 0% | 97% | 3% | 97% | 3% | 10043 | 50 | 7775 | 50 | | | | | | |
| bread_crisp_spread_dunno_fat_thick | 0% | 97% | 3% | 97% | 3% | 10043 | 50 | 7775 | 50 | | | | | | |
| bread_crisp_spread_dunno_fat_thin | 0% | 97% | 3% | 97% | 3% | 10043 | 50 | 7775 | 50 | | | | | | |
| bread_crisp_spread_dunno_lowfat_med | 0% | 97% | 3% | 97% | 3% | 10047 | 50 | 10049 | 50 | | | | | | |
| bread_crisp_spread_dunno_lowfat_thick | 0% | 97% | 3% | 97% | 3% | 10047 | 50 | 10049 | 50 | | | | | | |
| bread_crisp_spread_dunno_lowfat_thin | 0% | 97% | 3% | 97% | 3% | 10047 | 50 | 10049 | 50 | | | | | | |
| bread_crisp_spread_dunno_vlowfat_med | 0% | 97% | 3% | 97% | 3% | 10047 | 50 | 10049 | 50 | | | | | | |
| bread_crisp_spread_dunno_vlowfat_thick | 0% | 97% | 3% | 97% | 3% | 10047 | 50 | 10049 | 50 | | | | | | |
| bread_crisp_spread_dunno_vlowfat_thin | 0% | 97% | 3% | 97% | 3% | 10047 | 50 | 10049 | 50 | | | | | | |
| bread_crisp_spread_hardmarg_med | 0% | 100% | 0% | 100% | 0% | 860 | 100 | | | | | | | | |
| bread_crisp_spread_hardmarg_thick | 0% | 100% | 0% | 100% | 0% | 860 | 100 | | | | | | | | |
| bread_crisp_spread_hardmarg_thin | 0% | 100% | 0% | 100% | 0% | 860 | 100 | | | | | | | | |
| bread_crisp_spread_olive_chol_med | 0% | 99% | 1% | 99% | 1% | 3364 | 50 | 10053 | 50 | | | | | | |
| bread_crisp_spread_olive_chol_thick | 0% | 99% | 1% | 99% | 1% | 3364 | 50 | 10053 | 50 | | | | | | |
| bread_crisp_spread_olive_chol_thin | 0% | 99% | 1% | 99% | 1% | 3364 | 50 | 10053 | 50 | | | | | | |
| bread_crisp_spread_olive_dunno_med | 0% | 99% | 1% | 99% | 1% | 10048 | 50 | 10042 | 25 | 10131 | 25 | | | | |
| bread_crisp_spread_olive_dunno_thick | 0% | 99% | 1% | 99% | 1% | 10048 | 50 | 10042 | 25 | 10131 | 25 | | | | |
| bread_crisp_spread_olive_dunno_thin | 0% | 99% | 1% | 99% | 1% | 10048 | 50 | 10042 | 25 | 10131 | 25 | | | | |
| bread_crisp_spread_olive_fat_med | 0% | 99% | 1% | 99% | 1% | 10042 | 75 | 10131 | 25 | | | | | | |
| bread_crisp_spread_olive_fat_thick | 0% | 99% | 1% | 99% | 1% | 10042 | 75 | 10131 | 25 | | | | | | |
| bread_crisp_spread_olive_fat_thin | 0% | 99% | 1% | 99% | 1% | 10042 | 75 | 10131 | 25 | | | | | | |
| bread_crisp_spread_olive_lowfat_med | 0% | 100% | 1% | 100% | 0% | 10048 | 100 | | | | | | | | |
| bread_crisp_spread_olive_lowfat_thick | 0% | 100% | 1% | 100% | 0% | 10048 | 100 | | | | | | | | |
| bread_crisp_spread_olive_lowfat_thin | 0% | 100% | 1% | 100% | 0% | 10048 | 100 | | | | | | | | |
| bread_crisp_spread_olive_vlowfat_med | 0% | 100% | 1% | 100% | 0% | 10048 | 100 | | | | | | | | |
| bread_crisp_spread_olive_vlowfat_thick | 0% | 100% | 1% | 100% | 0% | 10048 | 100 | | | | | | | | |
| bread_crisp_spread_olive_vlowfat_thin | 0% | 100% | 1% | 100% | 0% | 10048 | 100 | | | | | | | | |
| bread_crisp_spread_other_med | 0% | 60% | 40% | 60% | 40% | 856 | 18 | 851 | 7.5 | 852 | 7.5 | 3848 | 33.5 | 2849 | 33.5 |
| bread_crisp_spread_other_thick | 0% | 60% | 40% | 60% | 40% | 856 | 18 | 851 | 7.5 | 852 | 7.5 | 3848 | 33.5 | 2849 | 33.5 |
| bread_crisp_spread_other_thin | 0% | 60% | 40% | 60% | 40% | 856 | 18 | 851 | 7.5 | 852 | 7.5 | 3848 | 33.5 | 2849 | 33.5 |
| bread_crisp_spread_polymarg_chol_med | 0% | 100% | 0% | 100% | 0% | 2849 | 33.4 | 3848 | 33.3 | 3243 | 33.3 | | | | |
| bread_crisp_spread_polymarg_chol_thick | 0% | 100% | 0% | 100% | 0% | 2849 | 33.4 | 3848 | 33.3 | 3243 | 33.3 | | | | |
| bread_crisp_spread_polymarg_chol_thin | 0% | 100% | 0% | 100% | 0% | 2849 | 33.4 | 3848 | 33.3 | 3243 | 33.3 | | | | |
| bread_crisp_spread_polymarg_dunno_med | 0% | 100% | 0% | 100% | 0% | 10049 | 50 | 10043 | 25 | 10044 | 25 | | | | |
| bread_crisp_spread_polymarg_dunno_thick | 0% | 100% | 0% | 100% | 0% | 10049 | 50 | 10043 | 25 | 10044 | 25 | | | | |
| bread_crisp_spread_polymarg_dunno_thin | 0% | 100% | 0% | 100% | 0% | 10049 | 50 | 10043 | 25 | 10044 | 25 | | | | |
| bread_crisp_spread_polymarg_fat_med | 0% | 100% | 0% | 100% | 0% | 10043 | 50 | 10044 | 50 | | | | | | |
| bread_crisp_spread_polymarg_fat_thick | 0% | 100% | 0% | 100% | 0% | 10043 | 50 | 10044 | 50 | | | | | | |
| bread_crisp_spread_polymarg_fat_thin | 0% | 100% | 0% | 100% | 0% | 10043 | 50 | 10044 | 50 | | | | | | |
| bread_crisp_spread_polymarg_lowfat_med | 0% | 100% | 0% | 100% | 0% | 10049 | 100 | | | | | | | | |
| bread_crisp_spread_polymarg_lowfat_thick | 0% | 100% | 0% | 100% | 0% | 10049 | 100 | | | | | | | | |
| bread_crisp_spread_polymarg_lowfat_thin | 0% | 100% | 0% | 100% | 0% | 10049 | 100 | | | | | | | | |
| bread_crisp_spread_polymarg_vlowfat_med | 0% | 100% | 0% | 100% | 0% | 10049 | 100 | | | | | | | | |
| bread_crisp_spread_polymarg_vlowfat_thick | 0% | 100% | 0% | 100% | 0% | 10049 | 100 | | | | | | | | |
| bread_crisp_spread_polymarg_vlowfat_thin | 0% | 100% | 0% | 100% | 0% | 10049 | 100 | | | | | | | | |
| bread_crisp_spread_soya_chol_med | 0% | 100% | 1% | 100% | 1% | 3848 | 16.6 | 3243 | 16.6 | 2849 | 16.8 | 10786 | 50 | | |
| bread_crisp_spread_soya_chol_thick | 0% | 100% | 1% | 100% | 1% | 3848 | 16.6 | 3243 | 16.6 | 2849 | 16.8 | 10786 | 50 | | |
| bread_crisp_spread_soya_chol_thin | 0% | 100% | 1% | 100% | 1% | 3848 | 16.6 | 3243 | 16.6 | 2849 | 16.8 | 10786 | 50 | | |
| bread_crisp_spread_soya_dunno_med | 0% | 100% | 0% | 100% | 0% | 10980 | 50 | 10786 | 50 | | | | | | |
| bread_crisp_spread_soya_dunno_thick | 0% | 100% | 0% | 100% | 0% | 10980 | 50 | 10786 | 50 | | | | | | |
| bread_crisp_spread_soya_dunno_thin | 0% | 100% | 0% | 100% | 0% | 10980 | 50 | 10786 | 50 | | | | | | |
| bread_crisp_spread_soya_fat_med | 0% | 100% | 0% | 100% | 0% | 10786 | 100 | | | | | | | | |
| bread_crisp_spread_soya_fat_thick | 0% | 100% | 0% | 100% | 0% | 10786 | 100 | | | | | | | | |
| bread_crisp_spread_soya_fat_thin | 0% | 100% | 0% | 100% | 0% | 10786 | 100 | | | | | | | | |
| bread_crisp_spread_soya_lowfat_med | 0% | 100% | 0% | 100% | 0% | 10980 | 100 | | | | | | | | |
| bread_crisp_spread_soya_lowfat_thick | 0% | 100% | 0% | 100% | 0% | 10980 | 100 | | | | | | | | |
| bread_crisp_spread_soya_lowfat_thin | 0% | 100% | 0% | 100% | 0% | 10980 | 100 | | | | | | | | |
| bread_crisp_spread_soya_vlowfat_med | 0% | 100% | 0% | 100% | 0% | 10980 | 100 | | | | | | | | |
| bread_crisp_spread_soya_vlowfat_thick | 0% | 100% | 0% | 100% | 0% | 10980 | 100 | | | | | | | | |
| bread_crisp_spread_soya_vlowfat_thin | 0% | 100% | 0% | 100% | 0% | 10980 | 100 | | | | | | | | |
| bread_garlic | 0% | 99% | 1% | 99% | 1% | 7615 | 100 | | | | | | | | |
| bread_large_bap_gf_nonwhite | 1% | 99% | 1% | 100% | 0% | 10459 | 50 | 8394 | 50 | | | | | | |
| bread_large_bap_gf_unanswered | 1% | 90% | 10% | 100% | 0% | 8864 | 33.4 | 10459 | 33.3 | 8395 | 33.3 | | | | |
| bread_large_bap_gf_white | 1% | 90% | 10% | 100% | 0% | 8864 | 100 | | | | | | | | |
| bread_large_bap_mixed | 0% | 100% | 0% | 100% | 0% | 7620 | 33.4 | 7621 | 33.3 | 10779 | 33.3 | | | | |
| bread_large_bap_other | 0% | 100% | 0% | 100% | 0% | 110 | 20 | 114 | 20 | 120 | 60 | | | | |
| bread_large_bap_seeded | 0% | 100% | 0% | 100% | 0% | 2168 | 33.4 | 2167 | 33.3 | 8148 | 33.3 | | | | |
| bread_large_bap_spread_butter_dunno_med | 0% | 20% | 80% | 20% | 80% | 9407 | 25 | 10039 | 25 | 851 | 25 | 852 | 25 | | |
| bread_large_bap_spread_butter_dunno_thick | 0% | 20% | 80% | 20% | 80% | 9407 | 25 | 10039 | 25 | 851 | 25 | 852 | 25 | | |
| bread_large_bap_spread_butter_dunno_thin | 0% | 20% | 80% | 20% | 80% | 9407 | 25 | 10039 | 25 | 851 | 25 | 852 | 25 | | |
| bread_large_bap_spread_butter_fat_med | 0% | 0% | 100% | 0% | 100% | 851 | 50 | 852 | 50 | | | | | | |
| bread_large_bap_spread_butter_fat_thick | 0% | 0% | 100% | 0% | 100% | 851 | 50 | 852 | 50 | | | | | | |
| bread_large_bap_spread_butter_fat_thin | 0% | 0% | 100% | 0% | 100% | 851 | 50 | 852 | 50 | | | | | | |
| bread_large_bap_spread_butter_lowfat_med | 0% | 45% | 55% | 45% | 55% | 10140 | 100 | | | | | | | | |
| bread_large_bap_spread_butter_lowfat_thick | 0% | 45% | 55% | 45% | 55% | 10140 | 100 | | | | | | | | |
| bread_large_bap_spread_butter_lowfat_thin | 0% | 45% | 55% | 45% | 55% | 10140 | 100 | | | | | | | | |
| bread_large_bap_spread_butter_spread_fat_med | 0% | 40% | 60% | 40% | 60% | 9407 | 50 | 10039 | 50 | | | | | | |
| bread_large_bap_spread_butter_spread_fat_thick | 0% | 40% | 60% | 40% | 60% | 9407 | 50 | 10039 | 50 | | | | | | |
| bread_large_bap_spread_butter_spread_fat_thin | 0% | 40% | 60% | 40% | 60% | 9407 | 50 | 10039 | 50 | | | | | | |
| bread_large_bap_spread_butter_spread_lowfat_med | 0% | 45% | 55% | 45% | 55% | 3891 | 50 | 10894 | 50 | | | | | | |
| bread_large_bap_spread_butter_spread_lowfat_thick | 0% | 45% | 55% | 45% | 55% | 3891 | 50 | 10894 | 50 | | | | | | |
| bread_large_bap_spread_butter_spread_lowfat_thin | 0% | 45% | 55% | 45% | 55% | 3891 | 50 | 10894 | 50 | | | | | | |
| bread_large_bap_spread_dairy_chol_med | 0% | 97% | 3% | 97% | 3% | 3848 | 100 | | | | | | | | |
| bread_large_bap_spread_dairy_chol_thick | 0% | 97% | 3% | 97% | 3% | 3848 | 100 | | | | | | | | |
| bread_large_bap_spread_dairy_chol_thin | 0% | 97% | 3% | 97% | 3% | 3848 | 100 | | | | | | | | |
| bread_large_bap_spread_dairy_dunno_med | 0% | 97% | 3% | 97% | 3% | 10047 | 50 | 7775 | 50 | | | | | | |
| bread_large_bap_spread_dairy_dunno_thick | 0% | 97% | 3% | 97% | 3% | 10047 | 50 | 7775 | 50 | | | | | | |
| bread_large_bap_spread_dairy_dunno_thin | 0% | 97% | 3% | 97% | 3% | 10047 | 50 | 7775 | 50 | | | | | | |
| bread_large_bap_spread_dairy_fat_med | 0% | 97% | 3% | 97% | 3% | 7775 | 100 | | | | | | | | |
| bread_large_bap_spread_dairy_fat_thick | 0% | 97% | 3% | 97% | 3% | 7775 | 100 | | | | | | | | |
| bread_large_bap_spread_dairy_fat_thin | 0% | 97% | 3% | 97% | 3% | 7775 | 100 | | | | | | | | |
| bread_large_bap_spread_dairy_lowfat_med | 0% | 99% | 1% | 99% | 1% | 10047 | 100 | | | | | | | | |
| bread_large_bap_spread_dairy_lowfat_thick | 0% | 99% | 1% | 99% | 1% | 10047 | 100 | | | | | | | | |
| bread_large_bap_spread_dairy_lowfat_thin | 0% | 99% | 1% | 99% | 1% | 10047 | 100 | | | | | | | | |
| bread_large_bap_spread_dairy_vlowfat_med | 0% | 99% | 1% | 99% | 1% | 10047 | 100 | | | | | | | | |
| bread_large_bap_spread_dairy_vlowfat_thick | 0% | 99% | 1% | 99% | 1% | 10047 | 100 | | | | | | | | |
| bread_large_bap_spread_dairy_vlowfat_thin | 0% | 99% | 1% | 99% | 1% | 10047 | 100 | | | | | | | | |
| bread_large_bap_spread_dunno_chol_med | 0% | 97% | 3% | 97% | 3% | 3848 | 33.4 | 3243 | 33.3 | 2849 | 33.3 | | | | |
| bread_large_bap_spread_dunno_chol_thick | 0% | 97% | 3% | 97% | 3% | 3848 | 33.4 | 3243 | 33.3 | 2849 | 33.3 | | | | |
| bread_large_bap_spread_dunno_chol_thin | 0% | 97% | 3% | 97% | 3% | 3848 | 33.4 | 3243 | 33.3 | 2849 | 33.3 | | | | |
| bread_large_bap_spread_dunno_dunno_med | 0% | 97% | 3% | 97% | 3% | 10047 | 25 | 10049 | 25 | 10043 | 25 | 7775 | 25 | | |
| bread_large_bap_spread_dunno_dunno_thick | 0% | 97% | 3% | 97% | 3% | 10047 | 25 | 10049 | 25 | 10043 | 25 | 7775 | 25 | | |
| bread_large_bap_spread_dunno_dunno_thin | 0% | 97% | 3% | 97% | 3% | 10047 | 25 | 10049 | 25 | 10043 | 25 | 7775 | 25 | | |
| bread_large_bap_spread_dunno_fat_med | 0% | 97% | 3% | 97% | 3% | 10043 | 50 | 7775 | 50 | | | | | | |
| bread_large_bap_spread_dunno_fat_thick | 0% | 97% | 3% | 97% | 3% | 10043 | 50 | 7775 | 50 | | | | | | |
| bread_large_bap_spread_dunno_fat_thin | 0% | 97% | 3% | 97% | 3% | 10043 | 50 | 7775 | 50 | | | | | | |
| bread_large_bap_spread_dunno_lowfat_med | 0% | 97% | 3% | 97% | 3% | 10047 | 50 | 10049 | 50 | | | | | | |
| bread_large_bap_spread_dunno_lowfat_thick | 0% | 97% | 3% | 97% | 3% | 10047 | 50 | 10049 | 50 | | | | | | |
| bread_large_bap_spread_dunno_lowfat_thin | 0% | 97% | 3% | 97% | 3% | 10047 | 50 | 10049 | 50 | | | | | | |
| bread_large_bap_spread_dunno_vlowfat_med | 0% | 97% | 3% | 97% | 3% | 10047 | 50 | 10049 | 50 | | | | | | |
| bread_large_bap_spread_dunno_vlowfat_thick | 0% | 97% | 3% | 97% | 3% | 10047 | 50 | 10049 | 50 | | | | | | |
| bread_large_bap_spread_dunno_vlowfat_thin | 0% | 97% | 3% | 97% | 3% | 10047 | 50 | 10049 | 50 | | | | | | |
| bread_large_bap_spread_hardmarg_med | 0% | 100% | 0% | 100% | 0% | 860 | 100 | | | | | | | | |
| bread_large_bap_spread_hardmarg_thick | 0% | 100% | 0% | 100% | 0% | 860 | 100 | | | | | | | | |
| bread_large_bap_spread_hardmarg_thin | 0% | 100% | 0% | 100% | 0% | 860 | 100 | | | | | | | | |
| bread_large_bap_spread_olive_chol_med | 0% | 99% | 1% | 99% | 1% | 3364 | 50 | 10053 | 50 | | | | | | |
| bread_large_bap_spread_olive_chol_thick | 0% | 99% | 1% | 99% | 1% | 3364 | 50 | 10053 | 50 | | | | | | |
| bread_large_bap_spread_olive_chol_thin | 0% | 99% | 1% | 99% | 1% | 3364 | 50 | 10053 | 50 | | | | | | |
| bread_large_bap_spread_olive_dunno_med | 0% | 99% | 1% | 99% | 1% | 10048 | 50 | 10042 | 25 | 10131 | 25 | | | | |
| bread_large_bap_spread_olive_dunno_thick | 0% | 99% | 1% | 99% | 1% | 10048 | 50 | 10042 | 25 | 10131 | 25 | | | | |
| bread_large_bap_spread_olive_dunno_thin | 0% | 99% | 1% | 99% | 1% | 10048 | 50 | 10042 | 25 | 10131 | 25 | | | | |
| bread_large_bap_spread_olive_fat_med | 0% | 99% | 1% | 99% | 1% | 10042 | 75 | 10131 | 25 | | | | | | |
| bread_large_bap_spread_olive_fat_thick | 0% | 99% | 1% | 99% | 1% | 10042 | 75 | 10131 | 25 | | | | | | |
| bread_large_bap_spread_olive_fat_thin | 0% | 99% | 1% | 99% | 1% | 10042 | 75 | 10131 | 25 | | | | | | |
| bread_large_bap_spread_olive_lowfat_med | 0% | 100% | 1% | 100% | 0% | 10048 | 100 | | | | | | | | |
| bread_large_bap_spread_olive_lowfat_thick | 0% | 100% | 1% | 100% | 0% | 10048 | 100 | | | | | | | | |
| bread_large_bap_spread_olive_lowfat_thin | 0% | 100% | 1% | 100% | 0% | 10048 | 100 | | | | | | | | |
| bread_large_bap_spread_olive_vlowfat_med | 0% | 100% | 1% | 100% | 0% | 10048 | 100 | | | | | | | | |
| bread_large_bap_spread_olive_vlowfat_thick | 0% | 100% | 1% | 100% | 0% | 10048 | 100 | | | | | | | | |
| bread_large_bap_spread_olive_vlowfat_thin | 0% | 100% | 1% | 100% | 0% | 10048 | 100 | | | | | | | | |
| bread_large_bap_spread_other_med | 0% | 60% | 40% | 60% | 40% | 856 | 18 | 851 | 7.5 | 852 | 7.5 | 3848 | 33.5 | 2849 | 33.5 |
| bread_large_bap_spread_other_thick | 0% | 60% | 40% | 60% | 40% | 856 | 18 | 851 | 7.5 | 852 | 7.5 | 3848 | 33.5 | 2849 | 33.5 |
| bread_large_bap_spread_other_thin | 0% | 60% | 40% | 60% | 40% | 856 | 18 | 851 | 7.5 | 852 | 7.5 | 3848 | 33.5 | 2849 | 33.5 |
| bread_large_bap_spread_polymarg_chol_med | 0% | 100% | 0% | 100% | 0% | 2849 | 33.4 | 3848 | 33.3 | 3243 | 33.3 | | | | |
| bread_large_bap_spread_polymarg_chol_thick | 0% | 100% | 0% | 100% | 0% | 2849 | 33.4 | 3848 | 33.3 | 3243 | 33.3 | | | | |
| bread_large_bap_spread_polymarg_chol_thin | 0% | 100% | 0% | 100% | 0% | 2849 | 33.4 | 3848 | 33.3 | 3243 | 33.3 | | | | |
| bread_large_bap_spread_polymarg_dunno_med | 0% | 100% | 0% | 100% | 0% | 10049 | 50 | 10043 | 25 | 10044 | 25 | | | | |
| bread_large_bap_spread_polymarg_dunno_thick | 0% | 100% | 0% | 100% | 0% | 10049 | 50 | 10043 | 25 | 10044 | 25 | | | | |
| bread_large_bap_spread_polymarg_dunno_thin | 0% | 100% | 0% | 100% | 0% | 10049 | 50 | 10043 | 25 | 10044 | 25 | | | | |
| bread_large_bap_spread_polymarg_fat_med | 0% | 100% | 0% | 100% | 0% | 10043 | 50 | 10044 | 50 | | | | | | |
| bread_large_bap_spread_polymarg_fat_thick | 0% | 100% | 0% | 100% | 0% | 10043 | 50 | 10044 | 50 | | | | | | |
| bread_large_bap_spread_polymarg_fat_thin | 0% | 100% | 0% | 100% | 0% | 10043 | 50 | 10044 | 50 | | | | | | |
| bread_large_bap_spread_polymarg_lowfat_med | 0% | 100% | 0% | 100% | 0% | 10049 | 100 | | | | | | | | |
| bread_large_bap_spread_polymarg_lowfat_thick | 0% | 100% | 0% | 100% | 0% | 10049 | 100 | | | | | | | | |
| bread_large_bap_spread_polymarg_lowfat_thin | 0% | 100% | 0% | 100% | 0% | 10049 | 100 | | | | | | | | |
| bread_large_bap_spread_polymarg_vlowfat_med | 0% | 100% | 0% | 100% | 0% | 10049 | 100 | | | | | | | | |
| bread_large_bap_spread_polymarg_vlowfat_thick | 0% | 100% | 0% | 100% | 0% | 10049 | 100 | | | | | | | | |
| bread_large_bap_spread_polymarg_vlowfat_thin | 0% | 100% | 0% | 100% | 0% | 10049 | 100 | | | | | | | | |
| bread_large_bap_spread_soya_chol_med | 0% | 100% | 1% | 100% | 1% | 3848 | 16.6 | 3243 | 16.6 | 2849 | 16.8 | 10786 | 50 | | |
| bread_large_bap_spread_soya_chol_thick | 0% | 100% | 1% | 100% | 1% | 3848 | 16.6 | 3243 | 16.6 | 2849 | 16.8 | 10786 | 50 | | |
| bread_large_bap_spread_soya_chol_thin | 0% | 100% | 1% | 100% | 1% | 3848 | 16.6 | 3243 | 16.6 | 2849 | 16.8 | 10786 | 50 | | |
| bread_large_bap_spread_soya_dunno_med | 0% | 100% | 0% | 100% | 0% | 10980 | 50 | 10786 | 50 | | | | | | |
| bread_large_bap_spread_soya_dunno_thick | 0% | 100% | 0% | 100% | 0% | 10980 | 50 | 10786 | 50 | | | | | | |
| bread_large_bap_spread_soya_dunno_thin | 0% | 100% | 0% | 100% | 0% | 10980 | 50 | 10786 | 50 | | | | | | |
| bread_large_bap_spread_soya_fat_med | 0% | 100% | 0% | 100% | 0% | 10786 | 100 | | | | | | | | |
| bread_large_bap_spread_soya_fat_thick | 0% | 100% | 0% | 100% | 0% | 10786 | 100 | | | | | | | | |
| bread_large_bap_spread_soya_fat_thin | 0% | 100% | 0% | 100% | 0% | 10786 | 100 | | | | | | | | |
| bread_large_bap_spread_soya_lowfat_med | 0% | 100% | 0% | 100% | 0% | 10980 | 100 | | | | | | | | |
| bread_large_bap_spread_soya_lowfat_thick | 0% | 100% | 0% | 100% | 0% | 10980 | 100 | | | | | | | | |
| bread_large_bap_spread_soya_lowfat_thin | 0% | 100% | 0% | 100% | 0% | 10980 | 100 | | | | | | | | |
| bread_large_bap_spread_soya_vlowfat_med | 0% | 100% | 0% | 100% | 0% | 10980 | 100 | | | | | | | | |
| bread_large_bap_spread_soya_vlowfat_thick | 0% | 100% | 0% | 100% | 0% | 10980 | 100 | | | | | | | | |
| bread_large_bap_spread_soya_vlowfat_thin | 0% | 100% | 0% | 100% | 0% | 10980 | 100 | | | | | | | | |
| bread_large_bap_unanswered | 0% | 100% | 0% | 100% | 0% | 159 | 33.4 | 7621 | 33.3 | 161 | 33.3 | | | | |
| bread_large_bap_white | 0% | 100% | 0% | 100% | 0% | 116 | 50 | 159 | 50 | | | | | | |
| bread_large_bap_wholemeal | 0% | 100% | 0% | 100% | 0% | 161 | 50 | 117 | 50 | | | | | | |
| bread_naan | 0% | 99% | 1% | 99% | 1% | 7622 | 100 | | | | | | | | |
| bread_other | 0% | 100% | 0% | 100% | 0% | 6838 | 40 | 148 | 40 | 7649 | 10 | 384 | 10 | | |
| bread_other_gf | 1% | 90% | 10% | 100% | 0% | 6838 | 40 | 148 | 40 | 7649 | 10 | 384 | 10 | | |

| | | | | | | | | | | | | | | | |
|---|---|---|---|---|---|---|---|---|---|---|---|---|---|---|---|
| bread_other_spread_butter_dunno_med | 0% | 20% | 80% | 20% | 80% | 9407 | 25 | 10039 | 25 | 851 | 25 | 852 | 25 | | |
| bread_other_spread_butter_dunno_thick | 0% | 20% | 80% | 20% | 80% | 9407 | 25 | 10039 | 25 | 851 | 25 | 852 | 25 | | |
| bread_other_spread_butter_dunno_thin | 0% | 20% | 80% | 20% | 80% | 9407 | 25 | 10039 | 25 | 851 | 25 | 852 | 25 | | |
| bread_other_spread_butter_fat_med | 0% | 0% | 100% | 0% | 100% | 851 | 50 | 852 | 50 | | | | | | |
| bread_other_spread_butter_fat_thick | 0% | 0% | 100% | 0% | 100% | 851 | 50 | 852 | 50 | | | | | | |
| bread_other_spread_butter_fat_thin | 0% | 0% | 100% | 0% | 100% | 851 | 50 | 852 | 50 | | | | | | |
| bread_other_spread_butter_lowfat_med | 0% | 45% | 55% | 45% | 55% | 10140 | 100 | | | | | | | | |
| bread_other_spread_butter_lowfat_thick | 0% | 45% | 55% | 45% | 55% | 10140 | 100 | | | | | | | | |
| bread_other_spread_butter_lowfat_thin | 0% | 45% | 55% | 45% | 55% | 10140 | 100 | | | | | | | | |
| bread_other_spread_butter_spread_fat_med | 0% | 40% | 60% | 40% | 60% | 9407 | 50 | 10039 | 50 | | | | | | |
| bread_other_spread_butter_spread_fat_thick | 0% | 40% | 60% | 40% | 60% | 9407 | 50 | 10039 | 50 | | | | | | |
| bread_other_spread_butter_spread_fat_thin | 0% | 40% | 60% | 40% | 60% | 9407 | 50 | 10039 | 50 | | | | | | |
| bread_other_spread_butter_spread_lowfat_med | 0% | 45% | 55% | 45% | 55% | 3891 | 50 | 10894 | 50 | | | | | | |
| bread_other_spread_butter_spread_lowfat_thick | 0% | 45% | 55% | 45% | 55% | 3891 | 50 | 10894 | 50 | | | | | | |
| bread_other_spread_butter_spread_lowfat_thin | 0% | 45% | 55% | 45% | 55% | 3891 | 50 | 10894 | 50 | | | | | | |
| bread_other_spread_dairy_chol_med | 0% | 97% | 3% | 97% | 3% | 3848 | 100 | | | | | | | | |
| bread_other_spread_dairy_chol_thick | 0% | 97% | 3% | 97% | 3% | 3848 | 100 | | | | | | | | |
| bread_other_spread_dairy_chol_thin | 0% | 97% | 3% | 97% | 3% | 3848 | 100 | | | | | | | | |
| bread_other_spread_dairy_dunno_med | 0% | 97% | 3% | 97% | 3% | 10047 | 50 | 7775 | 50 | | | | | | |
| bread_other_spread_dairy_dunno_thick | 0% | 97% | 3% | 97% | 3% | 10047 | 50 | 7775 | 50 | | | | | | |
| bread_other_spread_dairy_dunno_thin | 0% | 97% | 3% | 97% | 3% | 10047 | 50 | 7775 | 50 | | | | | | |
| bread_other_spread_dairy_fat_med | 0% | 97% | 3% | 97% | 3% | 7775 | 100 | | | | | | | | |
| bread_other_spread_dairy_fat_thick | 0% | 97% | 3% | 97% | 3% | 7775 | 100 | | | | | | | | |
| bread_other_spread_dairy_fat_thin | 0% | 97% | 3% | 97% | 3% | 7775 | 100 | | | | | | | | |
| bread_other_spread_dairy_lowfat_med | 0% | 99% | 1% | 99% | 1% | 10047 | 100 | | | | | | | | |
| bread_other_spread_dairy_lowfat_thick | 0% | 99% | 1% | 99% | 1% | 10047 | 100 | | | | | | | | |
| bread_other_spread_dairy_lowfat_thin | 0% | 99% | 1% | 99% | 1% | 10047 | 100 | | | | | | | | |
| bread_other_spread_dairy_vlowfat_med | 0% | 99% | 1% | 99% | 1% | 10047 | 100 | | | | | | | | |
| bread_other_spread_dairy_vlowfat_thick | 0% | 99% | 1% | 99% | 1% | 10047 | 100 | | | | | | | | |
| bread_other_spread_dairy_vlowfat_thin | 0% | 99% | 1% | 99% | 1% | 10047 | 100 | | | | | | | | |
| bread_other_spread_dunno_chol_med | 0% | 97% | 3% | 97% | 3% | 3848 | 33.4 | 3243 | 33.3 | 2849 | 33.3 | | | | |
| bread_other_spread_dunno_chol_thick | 0% | 97% | 3% | 97% | 3% | 3848 | 33.4 | 3243 | 33.3 | 2849 | 33.3 | | | | |
| bread_other_spread_dunno_chol_thin | 0% | 97% | 3% | 97% | 3% | 3848 | 33.4 | 3243 | 33.3 | 2849 | 33.3 | | | | |
| bread_other_spread_dunno_dunno_med | 0% | 97% | 3% | 97% | 3% | 10047 | 25 | 10049 | 25 | 10043 | 25 | 7775 | 25 | | |
| bread_other_spread_dunno_dunno_thick | 0% | 97% | 3% | 97% | 3% | 10047 | 25 | 10049 | 25 | 10043 | 25 | 7775 | 25 | | |
| bread_other_spread_dunno_dunno_thin | 0% | 97% | 3% | 97% | 3% | 10047 | 25 | 10049 | 25 | 10043 | 25 | 7775 | 25 | | |
| bread_other_spread_dunno_fat_med | 0% | 97% | 3% | 97% | 3% | 10043 | 50 | 7775 | 50 | | | | | | |
| bread_other_spread_dunno_fat_thick | 0% | 97% | 3% | 97% | 3% | 10043 | 50 | 7775 | 50 | | | | | | |
| bread_other_spread_dunno_fat_thin | 0% | 97% | 3% | 97% | 3% | 10043 | 50 | 7775 | 50 | | | | | | |
| bread_other_spread_dunno_lowfat_med | 0% | 97% | 3% | 97% | 3% | 10047 | 50 | 10049 | 50 | | | | | | |
| bread_other_spread_dunno_lowfat_thick | 0% | 97% | 3% | 97% | 3% | 10047 | 50 | 10049 | 50 | | | | | | |
| bread_other_spread_dunno_lowfat_thin | 0% | 97% | 3% | 97% | 3% | 10047 | 50 | 10049 | 50 | | | | | | |
| bread_other_spread_dunno_vlowfat_med | 0% | 97% | 3% | 97% | 3% | 10047 | 50 | 10049 | 50 | | | | | | |
| bread_other_spread_dunno_vlowfat_thick | 0% | 97% | 3% | 97% | 3% | 10047 | 50 | 10049 | 50 | | | | | | |
| bread_other_spread_dunno_vlowfat_thin | 0% | 97% | 3% | 97% | 3% | 10047 | 50 | 10049 | 50 | | | | | | |
| bread_other_spread_hardmarg_med | 0% | 100% | 0% | 100% | 0% | 860 | 100 | | | | | | | | |
| bread_other_spread_hardmarg_thick | 0% | 100% | 0% | 100% | 0% | 860 | 100 | | | | | | | | |
| bread_other_spread_hardmarg_thin | 0% | 100% | 0% | 100% | 0% | 860 | 100 | | | | | | | | |
| bread_other_spread_olive_chol_med | 0% | 99% | 1% | 99% | 1% | 3364 | 50 | 10053 | 50 | | | | | | |
| bread_other_spread_olive_chol_thick | 0% | 99% | 1% | 99% | 1% | 3364 | 50 | 10053 | 50 | | | | | | |
| bread_other_spread_olive_chol_thin | 0% | 99% | 1% | 99% | 1% | 3364 | 50 | 10053 | 50 | | | | | | |
| bread_other_spread_olive_dunno_med | 0% | 99% | 1% | 99% | 1% | 10048 | 50 | 10042 | 25 | 10131 | 25 | | | | |
| bread_other_spread_olive_dunno_thick | 0% | 99% | 1% | 99% | 1% | 10048 | 50 | 10042 | 25 | 10131 | 25 | | | | |
| bread_other_spread_olive_dunno_thin | 0% | 99% | 1% | 99% | 1% | 10048 | 50 | 10042 | 25 | 10131 | 25 | | | | |
| bread_other_spread_olive_fat_med | 0% | 99% | 1% | 99% | 1% | 10042 | 75 | 10131 | 25 | | | | | | |
| bread_other_spread_olive_fat_thick | 0% | 99% | 1% | 99% | 1% | 10042 | 75 | 10131 | 25 | | | | | | |
| bread_other_spread_olive_fat_thin | 0% | 99% | 1% | 99% | 1% | 10042 | 75 | 10131 | 25 | | | | | | |
| bread_other_spread_olive_lowfat_med | 0% | 100% | 1% | 100% | 0% | 10048 | 100 | | | | | | | | |
| bread_other_spread_olive_lowfat_thick | 0% | 100% | 1% | 100% | 0% | 10048 | 100 | | | | | | | | |
| bread_other_spread_olive_lowfat_thin | 0% | 100% | 1% | 100% | 0% | 10048 | 100 | | | | | | | | |
| bread_other_spread_olive_vlowfat_med | 0% | 100% | 1% | 100% | 0% | 10048 | 100 | | | | | | | | |
| bread_other_spread_olive_vlowfat_thick | 0% | 100% | 1% | 100% | 0% | 10048 | 100 | | | | | | | | |
| bread_other_spread_olive_vlowfat_thin | 0% | 100% | 1% | 100% | 0% | 10048 | 100 | | | | | | | | |
| bread_other_spread_other_med | 0% | 60% | 40% | 60% | 40% | 856 | 18 | 851 | 7.5 | 852 | 7.5 | 3848 | 33.5 | 2849 | 33.5 |
| bread_other_spread_other_thick | 0% | 60% | 40% | 60% | 40% | 856 | 18 | 851 | 7.5 | 852 | 7.5 | 3848 | 33.5 | 2849 | 33.5 |
| bread_other_spread_other_thin | 0% | 60% | 40% | 60% | 40% | 856 | 18 | 851 | 7.5 | 852 | 7.5 | 3848 | 33.5 | 2849 | 33.5 |
| bread_other_spread_polymarg_chol_med | 0% | 100% | 0% | 100% | 0% | 2849 | 33.4 | 3848 | 33.3 | 3243 | 33.3 | | | | |
| bread_other_spread_polymarg_chol_thick | 0% | 100% | 0% | 100% | 0% | 2849 | 33.4 | 3848 | 33.3 | 3243 | 33.3 | | | | |
| bread_other_spread_polymarg_chol_thin | 0% | 100% | 0% | 100% | 0% | 2849 | 33.4 | 3848 | 33.3 | 3243 | 33.3 | | | | |
| bread_other_spread_polymarg_dunno_med | 0% | 100% | 0% | 100% | 0% | 10049 | 50 | 10043 | 25 | 10044 | 25 | | | | |
| bread_other_spread_polymarg_dunno_thick | 0% | 100% | 0% | 100% | 0% | 10049 | 50 | 10043 | 25 | 10044 | 25 | | | | |
| bread_other_spread_polymarg_dunno_thin | 0% | 100% | 0% | 100% | 0% | 10049 | 50 | 10043 | 25 | 10044 | 25 | | | | |
| bread_other_spread_polymarg_fat_med | 0% | 100% | 0% | 100% | 0% | 10043 | 50 | 10044 | 50 | | | | | | |
| bread_other_spread_polymarg_fat_thick | 0% | 100% | 0% | 100% | 0% | 10043 | 50 | 10044 | 50 | | | | | | |
| bread_other_spread_polymarg_fat_thin | 0% | 100% | 0% | 100% | 0% | 10043 | 50 | 10044 | 50 | | | | | | |
| bread_other_spread_polymarg_lowfat_med | 0% | 100% | 0% | 100% | 0% | 10049 | 100 | | | | | | | | |
| bread_other_spread_polymarg_lowfat_thick | 0% | 100% | 0% | 100% | 0% | 10049 | 100 | | | | | | | | |
| bread_other_spread_polymarg_lowfat_thin | 0% | 100% | 0% | 100% | 0% | 10049 | 100 | | | | | | | | |
| bread_other_spread_polymarg_vlowfat_med | 0% | 100% | 0% | 100% | 0% | 10049 | 100 | | | | | | | | |
| bread_other_spread_polymarg_vlowfat_thick | 0% | 100% | 0% | 100% | 0% | 10049 | 100 | | | | | | | | |
| bread_other_spread_polymarg_vlowfat_thin | 0% | 100% | 0% | 100% | 0% | 10049 | 100 | | | | | | | | |
| bread_other_spread_soya_chol_med | 0% | 100% | 1% | 100% | 1% | 3848 | 16.6 | 3243 | 16.6 | 2849 | 16.8 | 10786 | 50 | | |
| bread_other_spread_soya_chol_thick | 0% | 100% | 1% | 100% | 1% | 3848 | 16.6 | 3243 | 16.6 | 2849 | 16.8 | 10786 | 50 | | |
| bread_other_spread_soya_chol_thin | 0% | 100% | 1% | 100% | 1% | 3848 | 16.6 | 3243 | 16.6 | 2849 | 16.8 | 10786 | 50 | | |
| bread_other_spread_soya_dunno_med | 0% | 100% | 0% | 100% | 0% | 10980 | 50 | 10786 | 50 | | | | | | |
| bread_other_spread_soya_dunno_thick | 0% | 100% | 0% | 100% | 0% | 10980 | 50 | 10786 | 50 | | | | | | |
| bread_other_spread_soya_dunno_thin | 0% | 100% | 0% | 100% | 0% | 10980 | 50 | 10786 | 50 | | | | | | |
| bread_other_spread_soya_fat_med | 0% | 100% | 0% | 100% | 0% | 10786 | 100 | | | | | | | | |
| bread_other_spread_soya_fat_thick | 0% | 100% | 0% | 100% | 0% | 10786 | 100 | | | | | | | | |
| bread_other_spread_soya_fat_thin | 0% | 100% | 0% | 100% | 0% | 10786 | 100 | | | | | | | | |
| bread_other_spread_soya_lowfat_med | 0% | 100% | 0% | 100% | 0% | 10980 | 100 | | | | | | | | |
| bread_other_spread_soya_lowfat_thick | 0% | 100% | 0% | 100% | 0% | 10980 | 100 | | | | | | | | |
| bread_other_spread_soya_lowfat_thin | 0% | 100% | 0% | 100% | 0% | 10980 | 100 | | | | | | | | |
| bread_other_spread_soya_vlowfat_med | 0% | 100% | 0% | 100% | 0% | 10980 | 100 | | | | | | | | |
| bread_other_spread_soya_vlowfat_thick | 0% | 100% | 0% | 100% | 0% | 10980 | 100 | | | | | | | | |
| bread_other_spread_soya_vlowfat_thin | 0% | 100% | 0% | 100% | 0% | 10980 | 100 | | | | | | | | |
| bread_roll_gf_nonwhite | 1% | 99% | 1% | 100% | 0% | 10459 | 50 | 8394 | 50 | | | | | | |
| bread_roll_gf_unanswered | 1% | 90% | 10% | 100% | 0% | 8864 | 33.4 | 10459 | 33.3 | 8395 | 33.3 | | | | |
| bread_roll_gf_white | 1% | 90% | 10% | 100% | 0% | 8864 | 100 | | | | | | | | |
| bread_roll_mixed | 0% | 100% | 0% | 100% | 0% | 7620 | 33.4 | 7621 | 33.3 | 10779 | 33.3 | | | | |
| bread_roll_other | 0% | 100% | 0% | 100% | 0% | 110 | 20 | 114 | 20 | 120 | 60 | | | | |
| bread_roll_seeded | 0% | 100% | 0% | 100% | 0% | 2168 | 33.4 | 2167 | 33.3 | 8148 | 33.3 | | | | |
| bread_roll_spread_butter_dunno_med | 0% | 20% | 80% | 20% | 80% | 9407 | 25 | 10039 | 25 | 851 | 25 | 852 | 25 | | |
| bread_roll_spread_butter_dunno_thick | 0% | 20% | 80% | 20% | 80% | 9407 | 25 | 10039 | 25 | 851 | 25 | 852 | 25 | | |
| bread_roll_spread_butter_dunno_thin | 0% | 20% | 80% | 20% | 80% | 9407 | 25 | 10039 | 25 | 851 | 25 | 852 | 25 | | |
| bread_roll_spread_butter_fat_med | 0% | 0% | 100% | 0% | 100% | 851 | 50 | 852 | 50 | | | | | | |
| bread_roll_spread_butter_fat_thick | 0% | 0% | 100% | 0% | 100% | 851 | 50 | 852 | 50 | | | | | | |
| bread_roll_spread_butter_fat_thin | 0% | 0% | 100% | 0% | 100% | 851 | 50 | 852 | 50 | | | | | | |
| bread_roll_spread_butter_lowfat_med | 0% | 45% | 55% | 45% | 55% | 10140 | 100 | | | | | | | | |
| bread_roll_spread_butter_lowfat_thick | 0% | 45% | 55% | 45% | 55% | 10140 | 100 | | | | | | | | |
| bread_roll_spread_butter_lowfat_thin | 0% | 45% | 55% | 45% | 55% | 10140 | 100 | | | | | | | | |
| bread_roll_spread_butter_spread_fat_med | 0% | 40% | 60% | 40% | 60% | 9407 | 50 | 10039 | 50 | | | | | | |
| bread_roll_spread_butter_spread_fat_thick | 0% | 40% | 60% | 40% | 60% | 9407 | 50 | 10039 | 50 | | | | | | |
| bread_roll_spread_butter_spread_fat_thin | 0% | 40% | 60% | 40% | 60% | 9407 | 50 | 10039 | 50 | | | | | | |
| bread_roll_spread_butter_spread_lowfat_med | 0% | 45% | 55% | 45% | 55% | 3891 | 50 | 10894 | 50 | | | | | | |
| bread_roll_spread_butter_spread_lowfat_thick | 0% | 45% | 55% | 45% | 55% | 3891 | 50 | 10894 | 50 | | | | | | |
| bread_roll_spread_butter_spread_lowfat_thin | 0% | 45% | 55% | 45% | 55% | 3891 | 50 | 10894 | 50 | | | | | | |
| bread_roll_spread_dairy_chol_med | 0% | 97% | 3% | 97% | 3% | 3848 | 100 | | | | | | | | |
| bread_roll_spread_dairy_chol_thick | 0% | 97% | 3% | 97% | 3% | 3848 | 100 | | | | | | | | |
| bread_roll_spread_dairy_chol_thin | 0% | 97% | 3% | 97% | 3% | 3848 | 100 | | | | | | | | |
| bread_roll_spread_dairy_dunno_med | 0% | 97% | 3% | 97% | 3% | 10047 | 50 | 7775 | 50 | | | | | | |
| bread_roll_spread_dairy_dunno_thick | 0% | 97% | 3% | 97% | 3% | 10047 | 50 | 7775 | 50 | | | | | | |
| bread_roll_spread_dairy_dunno_thin | 0% | 97% | 3% | 97% | 3% | 10047 | 50 | 7775 | 50 | | | | | | |
| bread_roll_spread_dairy_fat_med | 0% | 97% | 3% | 97% | 3% | 7775 | 100 | | | | | | | | |
| bread_roll_spread_dairy_fat_thick | 0% | 97% | 3% | 97% | 3% | 7775 | 100 | | | | | | | | |
| bread_roll_spread_dairy_fat_thin | 0% | 97% | 3% | 97% | 3% | 7775 | 100 | | | | | | | | |
| bread_roll_spread_dairy_lowfat_med | 0% | 99% | 1% | 99% | 1% | 10047 | 100 | | | | | | | | |
| bread_roll_spread_dairy_lowfat_thick | 0% | 99% | 1% | 99% | 1% | 10047 | 100 | | | | | | | | |
| bread_roll_spread_dairy_lowfat_thin | 0% | 99% | 1% | 99% | 1% | 10047 | 100 | | | | | | | | |
| bread_roll_spread_dairy_vlowfat_med | 0% | 99% | 1% | 99% | 1% | 10047 | 100 | | | | | | | | |
| bread_roll_spread_dairy_vlowfat_thick | 0% | 99% | 1% | 99% | 1% | 10047 | 100 | | | | | | | | |
| bread_roll_spread_dairy_vlowfat_thin | 0% | 99% | 1% | 99% | 1% | 10047 | 100 | | | | | | | | |
| bread_roll_spread_dunno_chol_med | 0% | 97% | 3% | 97% | 3% | 3848 | 33.4 | 3243 | 33.3 | 2849 | 33.3 | | | | |
| bread_roll_spread_dunno_chol_thick | 0% | 97% | 3% | 97% | 3% | 3848 | 33.4 | 3243 | 33.3 | 2849 | 33.3 | | | | |
| bread_roll_spread_dunno_chol_thin | 0% | 97% | 3% | 97% | 3% | 3848 | 33.4 | 3243 | 33.3 | 2849 | 33.3 | | | | |
| bread_roll_spread_dunno_dunno_med | 0% | 97% | 3% | 97% | 3% | 10047 | 25 | 10049 | 25 | 10043 | 25 | 7775 | 25 | | |
| bread_roll_spread_dunno_dunno_thick | 0% | 97% | 3% | 97% | 3% | 10047 | 25 | 10049 | 25 | 10043 | 25 | 7775 | 25 | | |
| bread_roll_spread_dunno_dunno_thin | 0% | 97% | 3% | 97% | 3% | 10047 | 25 | 10049 | 25 | 10043 | 25 | 7775 | 25 | | |
| bread_roll_spread_dunno_fat_med | 0% | 97% | 3% | 97% | 3% | 10043 | 50 | 7775 | 50 | | | | | | |
| bread_roll_spread_dunno_fat_thick | 0% | 97% | 3% | 97% | 3% | 10043 | 50 | 7775 | 50 | | | | | | |
| bread_roll_spread_dunno_fat_thin | 0% | 97% | 3% | 97% | 3% | 10043 | 50 | 7775 | 50 | | | | | | |
| bread_roll_spread_dunno_lowfat_med | 0% | 97% | 3% | 97% | 3% | 10047 | 50 | 10049 | 50 | | | | | | |
| bread_roll_spread_dunno_lowfat_thick | 0% | 97% | 3% | 97% | 3% | 10047 | 50 | 10049 | 50 | | | | | | |
| bread_roll_spread_dunno_lowfat_thin | 0% | 97% | 3% | 97% | 3% | 10047 | 50 | 10049 | 50 | | | | | | |
| bread_roll_spread_dunno_vlowfat_med | 0% | 97% | 3% | 97% | 3% | 10047 | 50 | 10049 | 50 | | | | | | |
| bread_roll_spread_dunno_vlowfat_thick | 0% | 97% | 3% | 97% | 3% | 10047 | 50 | 10049 | 50 | | | | | | |
| bread_roll_spread_dunno_vlowfat_thin | 0% | 97% | 3% | 97% | 3% | 10047 | 50 | 10049 | 50 | | | | | | |
| bread_roll_spread_hardmarg_med | 0% | 100% | 0% | 100% | 0% | 860 | 100 | | | | | | | | |
| bread_roll_spread_hardmarg_thick | 0% | 100% | 0% | 100% | 0% | 860 | 100 | | | | | | | | |
| bread_roll_spread_hardmarg_thin | 0% | 100% | 0% | 100% | 0% | 860 | 100 | | | | | | | | |
| bread_roll_spread_olive_chol_med | 0% | 99% | 1% | 99% | 1% | 3364 | 50 | 10053 | 50 | | | | | | |
| bread_roll_spread_olive_chol_thick | 0% | 99% | 1% | 99% | 1% | 3364 | 50 | 10053 | 50 | | | | | | |
| bread_roll_spread_olive_chol_thin | 0% | 99% | 1% | 99% | 1% | 3364 | 50 | 10053 | 50 | | | | | | |
| bread_roll_spread_olive_dunno_med | 0% | 99% | 1% | 99% | 1% | 10048 | 50 | 10042 | 25 | 10131 | 25 | | | | |
| bread_roll_spread_olive_dunno_thick | 0% | 99% | 1% | 99% | 1% | 10048 | 50 | 10042 | 25 | 10131 | 25 | | | | |
| bread_roll_spread_olive_dunno_thin | 0% | 99% | 1% | 99% | 1% | 10048 | 50 | 10042 | 25 | 10131 | 25 | | | | |
| bread_roll_spread_olive_fat_med | 0% | 99% | 1% | 99% | 1% | 10042 | 75 | 10131 | 25 | | | | | | |
| bread_roll_spread_olive_fat_thick | 0% | 99% | 1% | 99% | 1% | 10042 | 75 | 10131 | 25 | | | | | | |
| bread_roll_spread_olive_fat_thin | 0% | 99% | 1% | 99% | 1% | 10042 | 75 | 10131 | 25 | | | | | | |
| bread_roll_spread_olive_lowfat_med | 0% | 100% | 1% | 100% | 0% | 10048 | 100 | | | | | | | | |
| bread_roll_spread_olive_lowfat_thick | 0% | 100% | 1% | 100% | 0% | 10048 | 100 | | | | | | | | |
| bread_roll_spread_olive_lowfat_thin | 0% | 100% | 1% | 100% | 0% | 10048 | 100 | | | | | | | | |
| bread_roll_spread_olive_vlowfat_med | 0% | 100% | 1% | 100% | 0% | 10048 | 100 | | | | | | | | |
| bread_roll_spread_olive_vlowfat_thick | 0% | 100% | 1% | 100% | 0% | 10048 | 100 | | | | | | | | |
| bread_roll_spread_olive_vlowfat_thin | 0% | 100% | 1% | 100% | 0% | 10048 | 100 | | | | | | | | |
| bread_roll_spread_other_med | 0% | 60% | 40% | 60% | 40% | 856 | 18 | 851 | 7.5 | 852 | 7.5 | 3848 | 33.5 | 2849 | 33.5 |
| bread_roll_spread_other_thick | 0% | 60% | 40% | 60% | 40% | 856 | 18 | 851 | 7.5 | 852 | 7.5 | 3848 | 33.5 | 2849 | 33.5 |
| bread_roll_spread_other_thin | 0% | 60% | 40% | 60% | 40% | 856 | 18 | 851 | 7.5 | 852 | 7.5 | 3848 | 33.5 | 2849 | 33.5 |
| bread_roll_spread_polymarg_chol_med | 0% | 100% | 0% | 100% | 0% | 2849 | 33.4 | 3848 | 33.3 | 3243 | 33.3 | | | | |

| | | | | | | | | | | | | | | | | | | | | | | | | | |
|---|---|---|---|---|---|---|---|---|---|---|---|---|---|---|---|---|---|---|---|---|---|---|---|---|---|
| bread_roll_spread_polymarg_chol_thick | 0% | 100% | 0% | 100% | 0% | 2849 | 33.4 | 3848 | 33.3 | 3243 | 33.3 | | | | | | | | | | | | | | |
| bread_roll_spread_polymarg_chol_thin | 0% | 100% | 0% | 100% | 0% | 2849 | 33.4 | 3848 | 33.3 | 3243 | 33.3 | | | | | | | | | | | | | | |
| bread_roll_spread_polymarg_dunno_med | 0% | 100% | 0% | 100% | 0% | 10049 | 50 | 10043 | 25 | 10044 | 25 | | | | | | | | | | | | | | |
| bread_roll_spread_polymarg_dunno_thick | 0% | 100% | 0% | 100% | 0% | 10049 | 50 | 10043 | 25 | 10044 | 25 | | | | | | | | | | | | | | |
| bread_roll_spread_polymarg_dunno_thin | 0% | 100% | 0% | 100% | 0% | 10049 | 50 | 10043 | 25 | 10044 | 25 | | | | | | | | | | | | | | |
| bread_roll_spread_polymarg_fat_med | 0% | 100% | 0% | 100% | 0% | 10043 | 50 | 10044 | 50 | | | | | | | | | | | | | | | | |
| bread_roll_spread_polymarg_fat_thick | 0% | 100% | 0% | 100% | 0% | 10043 | 50 | 10044 | 50 | | | | | | | | | | | | | | | | |
| bread_roll_spread_polymarg_fat_thin | 0% | 100% | 0% | 100% | 0% | 10043 | 50 | 10044 | 50 | | | | | | | | | | | | | | | | |
| bread_roll_spread_polymarg_lowfat_med | 0% | 100% | 0% | 100% | 0% | 10049 | 100 | | | | | | | | | | | | | | | | | | |
| bread_roll_spread_polymarg_lowfat_thick | 0% | 100% | 0% | 100% | 0% | 10049 | 100 | | | | | | | | | | | | | | | | | | |
| bread_roll_spread_polymarg_lowfat_thin | 0% | 100% | 0% | 100% | 0% | 10049 | 100 | | | | | | | | | | | | | | | | | | |
| bread_roll_spread_polymarg_vlowfat_med | 0% | 100% | 0% | 100% | 0% | 10049 | 100 | | | | | | | | | | | | | | | | | | |
| bread_roll_spread_polymarg_vlowfat_thick | 0% | 100% | 0% | 100% | 0% | 10049 | 100 | | | | | | | | | | | | | | | | | | |
| bread_roll_spread_polymarg_vlowfat_thin | 0% | 100% | 0% | 100% | 0% | 10049 | 100 | | | | | | | | | | | | | | | | | | |
| bread_roll_spread_soya_chol_med | 0% | 100% | 1% | 100% | 1% | 3848 | 16.6 | 3243 | 16.6 | 2849 | 16.8 | 10786 | 50 | | | | | | | | | | | | |
| bread_roll_spread_soya_chol_thick | 0% | 100% | 1% | 100% | 1% | 3848 | 16.6 | 3243 | 16.6 | 2849 | 16.8 | 10786 | 50 | | | | | | | | | | | | |
| bread_roll_spread_soya_chol_thin | 0% | 100% | 1% | 100% | 1% | 3848 | 16.6 | 3243 | 16.6 | 2849 | 16.8 | 10786 | 50 | | | | | | | | | | | | |
| bread_roll_spread_soya_dunno_med | 0% | 100% | 0% | 100% | 0% | 10980 | 50 | 10786 | 50 | | | | | | | | | | | | | | | | |
| bread_roll_spread_soya_dunno_thick | 0% | 100% | 0% | 100% | 0% | 10980 | 50 | 10786 | 50 | | | | | | | | | | | | | | | | |
| bread_roll_spread_soya_dunno_thin | 0% | 100% | 0% | 100% | 0% | 10980 | 50 | 10786 | 50 | | | | | | | | | | | | | | | | |
| bread_roll_spread_soya_fat_med | 0% | 100% | 0% | 100% | 0% | 10786 | 100 | | | | | | | | | | | | | | | | | | |
| bread_roll_spread_soya_fat_thick | 0% | 100% | 0% | 100% | 0% | 10786 | 100 | | | | | | | | | | | | | | | | | | |
| bread_roll_spread_soya_fat_thin | 0% | 100% | 0% | 100% | 0% | 10786 | 100 | | | | | | | | | | | | | | | | | | |
| bread_roll_spread_soya_lowfat_med | 0% | 100% | 0% | 100% | 0% | 10980 | 100 | | | | | | | | | | | | | | | | | | |
| bread_roll_spread_soya_lowfat_thick | 0% | 100% | 0% | 100% | 0% | 10980 | 100 | | | | | | | | | | | | | | | | | | |
| bread_roll_spread_soya_lowfat_thin | 0% | 100% | 0% | 100% | 0% | 10980 | 100 | | | | | | | | | | | | | | | | | | |
| bread_roll_spread_soya_vlowfat_med | 0% | 100% | 0% | 100% | 0% | 10980 | 100 | | | | | | | | | | | | | | | | | | |
| bread_roll_spread_soya_vlowfat_thick | 0% | 100% | 0% | 100% | 0% | 10980 | 100 | | | | | | | | | | | | | | | | | | |
| bread_roll_spread_soya_vlowfat_thin | 0% | 100% | 0% | 100% | 0% | 10980 | 100 | | | | | | | | | | | | | | | | | | |
| bread_roll_unanswered | 0% | 100% | 0% | 100% | 0% | 159 | 33.4 | 7621 | 33.3 | 161 | 33.3 | | | | | | | | | | | | | | |
| bread_roll_white | 0% | 100% | 0% | 100% | 0% | 158 | 50 | 157 | 50 | | | | | | | | | | | | | | | | |
| bread_roll_wholemeal | 0% | 100% | 0% | 100% | 0% | 161 | 100 | | | | | | | | | | | | | | | | | | |
| bread_sliced_gf_nonwhite | 1% | 99% | 1% | 100% | 0% | 10459 | 50 | 8394 | 50 | | | | | | | | | | | | | | | | |
| bread_sliced_gf_unanswered | 1% | 90% | 10% | 100% | 0% | 8864 | 33.4 | 10459 | 33.3 | 8394 | 33.3 | | | | | | | | | | | | | | |
| bread_sliced_gf_white | 1% | 90% | 10% | 100% | 0% | 8864 | 100 | | | | | | | | | | | | | | | | | | |
| bread_sliced_mixed | 0% | 100% | 0% | 100% | 0% | 112 | 33.4 | 102 | 33.3 | 7609 | 16.7 | 10775 | 16.6 | | | | | | | | | | | | |
| bread_sliced_other | 0% | 100% | 0% | 100% | 0% | 110 | 25 | 114 | 25 | 102 | 25 | 120 | 25 | | | | | | | | | | | | |
| bread_sliced_seeded | 0% | 100% | 0% | 100% | 0% | 2168 | 33.4 | 2167 | 33.3 | 8148 | 33.3 | | | | | | | | | | | | | | |
| bread_sliced_spread_butter_dunno_med | 0% | 20% | 80% | 20% | 80% | 9407 | 25 | 10039 | 25 | 851 | 25 | 852 | 25 | | | | | | | | | | | | |
| bread_sliced_spread_butter_dunno_thick | 0% | 20% | 80% | 20% | 80% | 9407 | 25 | 10039 | 25 | 851 | 25 | 852 | 25 | | | | | | | | | | | | |
| bread_sliced_spread_butter_dunno_thin | 0% | 20% | 80% | 20% | 80% | 9407 | 25 | 10039 | 25 | 851 | 25 | 852 | 25 | | | | | | | | | | | | |
| bread_sliced_spread_butter_fat_med | 0% | 0% | 100% | 0% | 100% | 851 | 50 | 852 | 50 | | | | | | | | | | | | | | | | |
| bread_sliced_spread_butter_fat_thick | 0% | 0% | 100% | 0% | 100% | 851 | 50 | 852 | 50 | | | | | | | | | | | | | | | | |
| bread_sliced_spread_butter_fat_thin | 0% | 0% | 100% | 0% | 100% | 851 | 50 | 852 | 50 | | | | | | | | | | | | | | | | |
| bread_sliced_spread_butter_lowfat_med | 0% | 45% | 55% | 45% | 55% | 10140 | 100 | | | | | | | | | | | | | | | | | | |
| bread_sliced_spread_butter_lowfat_thick | 0% | 45% | 55% | 45% | 55% | 10140 | 100 | | | | | | | | | | | | | | | | | | |
| bread_sliced_spread_butter_lowfat_thin | 0% | 45% | 55% | 45% | 55% | 10140 | 100 | | | | | | | | | | | | | | | | | | |
| bread_sliced_spread_butter_spread_fat_med | 0% | 40% | 60% | 40% | 60% | 9407 | 50 | 10039 | 50 | | | | | | | | | | | | | | | | |
| bread_sliced_spread_butter_spread_fat_thick | 0% | 40% | 60% | 40% | 60% | 9407 | 50 | 10039 | 50 | | | | | | | | | | | | | | | | |
| bread_sliced_spread_butter_spread_fat_thin | 0% | 40% | 60% | 40% | 60% | 9407 | 50 | 10039 | 50 | | | | | | | | | | | | | | | | |
| bread_sliced_spread_butter_spread_lowfat_med | 0% | 45% | 55% | 45% | 55% | 3891 | 50 | 10894 | 50 | | | | | | | | | | | | | | | | |
| bread_sliced_spread_butter_spread_lowfat_thick | 0% | 45% | 55% | 45% | 55% | 3891 | 50 | 10894 | 50 | | | | | | | | | | | | | | | | |
| bread_sliced_spread_butter_spread_lowfat_thin | 0% | 45% | 55% | 45% | 55% | 3891 | 50 | 10894 | 50 | | | | | | | | | | | | | | | | |
| bread_sliced_spread_dairy_chol_med | 0% | 97% | 3% | 97% | 3% | 3848 | 100 | | | | | | | | | | | | | | | | | | |
| bread_sliced_spread_dairy_chol_thick | 0% | 97% | 3% | 97% | 3% | 3848 | 100 | | | | | | | | | | | | | | | | | | |
| bread_sliced_spread_dairy_chol_thin | 0% | 97% | 3% | 97% | 3% | 3848 | 100 | | | | | | | | | | | | | | | | | | |
| bread_sliced_spread_dairy_dunno_med | 0% | 97% | 3% | 97% | 3% | 10047 | 50 | 7775 | 50 | | | | | | | | | | | | | | | | |
| bread_sliced_spread_dairy_dunno_thick | 0% | 97% | 3% | 97% | 3% | 10047 | 50 | 7775 | 50 | | | | | | | | | | | | | | | | |
| bread_sliced_spread_dairy_dunno_thin | 0% | 97% | 3% | 97% | 3% | 10047 | 50 | 7775 | 50 | | | | | | | | | | | | | | | | |
| bread_sliced_spread_dairy_fat_med | 0% | 97% | 3% | 97% | 3% | 7775 | 100 | | | | | | | | | | | | | | | | | | |
| bread_sliced_spread_dairy_fat_thick | 0% | 97% | 3% | 97% | 3% | 7775 | 100 | | | | | | | | | | | | | | | | | | |
| bread_sliced_spread_dairy_fat_thin | 0% | 97% | 3% | 97% | 3% | 7775 | 100 | | | | | | | | | | | | | | | | | | |
| bread_sliced_spread_dairy_lowfat_med | 0% | 99% | 1% | 99% | 1% | 10047 | 100 | | | | | | | | | | | | | | | | | | |
| bread_sliced_spread_dairy_lowfat_thick | 0% | 99% | 1% | 99% | 1% | 10047 | 100 | | | | | | | | | | | | | | | | | | |
| bread_sliced_spread_dairy_lowfat_thin | 0% | 99% | 1% | 99% | 1% | 10047 | 100 | | | | | | | | | | | | | | | | | | |
| bread_sliced_spread_dairy_vlowfat_med | 0% | 99% | 1% | 99% | 1% | 10047 | 100 | | | | | | | | | | | | | | | | | | |
| bread_sliced_spread_dairy_vlowfat_thick | 0% | 99% | 1% | 99% | 1% | 10047 | 100 | | | | | | | | | | | | | | | | | | |
| bread_sliced_spread_dairy_vlowfat_thin | 0% | 99% | 1% | 99% | 1% | 10047 | 100 | | | | | | | | | | | | | | | | | | |
| bread_sliced_spread_dunno_chol_med | 0% | 97% | 3% | 97% | 3% | 3848 | 33.4 | 3243 | 33.3 | 2849 | 33.3 | | | | | | | | | | | | | | |
| bread_sliced_spread_dunno_chol_thick | 0% | 97% | 3% | 97% | 3% | 3848 | 33.4 | 3243 | 33.3 | 2849 | 33.3 | | | | | | | | | | | | | | |
| bread_sliced_spread_dunno_chol_thin | 0% | 97% | 3% | 97% | 3% | 3848 | 33.4 | 3243 | 33.3 | 2849 | 33.3 | | | | | | | | | | | | | | |
| bread_sliced_spread_dunno_dunno_med | 0% | 97% | 3% | 97% | 3% | 10047 | 25 | 10049 | 25 | 10043 | 25 | 7775 | 25 | | | | | | | | | | | | |
| bread_sliced_spread_dunno_dunno_thick | 0% | 97% | 3% | 97% | 3% | 10047 | 25 | 10049 | 25 | 10043 | 25 | 7775 | 25 | | | | | | | | | | | | |
| bread_sliced_spread_dunno_dunno_thin | 0% | 97% | 3% | 97% | 3% | 10047 | 25 | 10049 | 25 | 10043 | 25 | 7775 | 25 | | | | | | | | | | | | |
| bread_sliced_spread_dunno_fat_med | 0% | 97% | 3% | 97% | 3% | 10043 | 50 | 7775 | 50 | | | | | | | | | | | | | | | | |
| bread_sliced_spread_dunno_fat_thick | 0% | 97% | 3% | 97% | 3% | 10043 | 50 | 7775 | 50 | | | | | | | | | | | | | | | | |
| bread_sliced_spread_dunno_fat_thin | 0% | 97% | 3% | 97% | 3% | 10043 | 50 | 7775 | 50 | | | | | | | | | | | | | | | | |
| bread_sliced_spread_dunno_lowfat_med | 0% | 97% | 3% | 97% | 3% | 10047 | 50 | 10049 | 50 | | | | | | | | | | | | | | | | |
| bread_sliced_spread_dunno_lowfat_thick | 0% | 97% | 3% | 97% | 3% | 10047 | 50 | 10049 | 50 | | | | | | | | | | | | | | | | |
| bread_sliced_spread_dunno_lowfat_thin | 0% | 97% | 3% | 97% | 3% | 10047 | 50 | 10049 | 50 | | | | | | | | | | | | | | | | |
| bread_sliced_spread_dunno_vlowfat_med | 0% | 97% | 3% | 97% | 3% | 10047 | 50 | 10049 | 50 | | | | | | | | | | | | | | | | |
| bread_sliced_spread_dunno_vlowfat_thick | 0% | 97% | 3% | 97% | 3% | 10047 | 50 | 10049 | 50 | | | | | | | | | | | | | | | | |
| bread_sliced_spread_dunno_vlowfat_thin | 0% | 97% | 3% | 97% | 3% | 10047 | 50 | 10049 | 50 | | | | | | | | | | | | | | | | |
| bread_sliced_spread_hardmarg_med | 0% | 100% | 0% | 100% | 0% | 860 | 100 | | | | | | | | | | | | | | | | | | |
| bread_sliced_spread_hardmarg_thick | 0% | 100% | 0% | 100% | 0% | 860 | 100 | | | | | | | | | | | | | | | | | | |
| bread_sliced_spread_hardmarg_thin | 0% | 100% | 0% | 100% | 0% | 860 | 100 | | | | | | | | | | | | | | | | | | |
| bread_sliced_spread_olive_chol_med | 0% | 99% | 1% | 99% | 1% | 3364 | 50 | 10053 | 50 | | | | | | | | | | | | | | | | |
| bread_sliced_spread_olive_chol_thick | 0% | 99% | 1% | 99% | 1% | 3364 | 50 | 10053 | 50 | | | | | | | | | | | | | | | | |
| bread_sliced_spread_olive_chol_thin | 0% | 99% | 1% | 99% | 1% | 3364 | 50 | 10053 | 50 | | | | | | | | | | | | | | | | |
| bread_sliced_spread_olive_dunno_med | 0% | 99% | 1% | 99% | 1% | 10048 | 50 | 10042 | 25 | 10131 | 25 | | | | | | | | | | | | | | |
| bread_sliced_spread_olive_dunno_thick | 0% | 99% | 1% | 99% | 1% | 10048 | 50 | 10042 | 25 | 10131 | 25 | | | | | | | | | | | | | | |
| bread_sliced_spread_olive_dunno_thin | 0% | 99% | 1% | 99% | 1% | 10048 | 50 | 10042 | 25 | 10131 | 25 | | | | | | | | | | | | | | |
| bread_sliced_spread_olive_fat_med | 0% | 99% | 1% | 99% | 1% | 10042 | 75 | 10131 | 25 | | | | | | | | | | | | | | | | |
| bread_sliced_spread_olive_fat_thick | 0% | 99% | 1% | 99% | 1% | 10042 | 75 | 10131 | 25 | | | | | | | | | | | | | | | | |
| bread_sliced_spread_olive_fat_thin | 0% | 99% | 1% | 99% | 1% | 10042 | 75 | 10131 | 25 | | | | | | | | | | | | | | | | |
| bread_sliced_spread_olive_lowfat_med | 0% | 100% | 1% | 100% | 0% | 10048 | 100 | | | | | | | | | | | | | | | | | | |
| bread_sliced_spread_olive_lowfat_thick | 0% | 100% | 1% | 100% | 0% | 10048 | 100 | | | | | | | | | | | | | | | | | | |
| bread_sliced_spread_olive_lowfat_thin | 0% | 100% | 1% | 100% | 0% | 10048 | 100 | | | | | | | | | | | | | | | | | | |
| bread_sliced_spread_olive_vlowfat_med | 0% | 100% | 1% | 100% | 0% | 10048 | 100 | | | | | | | | | | | | | | | | | | |
| bread_sliced_spread_olive_vlowfat_thick | 0% | 100% | 1% | 100% | 0% | 10048 | 100 | | | | | | | | | | | | | | | | | | |
| bread_sliced_spread_olive_vlowfat_thin | 0% | 100% | 1% | 100% | 0% | 10048 | 100 | | | | | | | | | | | | | | | | | | |
| bread_sliced_spread_other_med | 0% | 60% | 40% | 60% | 40% | 856 | 18 | 851 | 7.5 | 852 | 7.5 | 3848 | 33.5 | 2849 | 33.5 | | | | | | | | | | |
| bread_sliced_spread_other_thick | 0% | 60% | 40% | 60% | 40% | 856 | 18 | 851 | 7.5 | 852 | 7.5 | 3848 | 33.5 | 2849 | 33.5 | | | | | | | | | | |
| bread_sliced_spread_other_thin | 0% | 60% | 40% | 60% | 40% | 856 | 18 | 851 | 7.5 | 852 | 7.5 | 3848 | 33.5 | 2849 | 33.5 | | | | | | | | | | |
| bread_sliced_spread_polymarg_chol_med | 0% | 100% | 0% | 100% | 0% | 2849 | 33.4 | 3848 | 33.3 | 3243 | 33.3 | | | | | | | | | | | | | | |
| bread_sliced_spread_polymarg_chol_thick | 0% | 100% | 0% | 100% | 0% | 2849 | 33.4 | 3848 | 33.3 | 3243 | 33.3 | | | | | | | | | | | | | | |
| bread_sliced_spread_polymarg_chol_thin | 0% | 100% | 0% | 100% | 0% | 2849 | 33.4 | 3848 | 33.3 | 3243 | 33.3 | | | | | | | | | | | | | | |
| bread_sliced_spread_polymarg_dunno_med | 0% | 100% | 0% | 100% | 0% | 10049 | 50 | 10043 | 25 | 10044 | 25 | | | | | | | | | | | | | | |
| bread_sliced_spread_polymarg_dunno_thick | 0% | 100% | 0% | 100% | 0% | 10049 | 50 | 10043 | 25 | 10044 | 25 | | | | | | | | | | | | | | |
| bread_sliced_spread_polymarg_dunno_thin | 0% | 100% | 0% | 100% | 0% | 10049 | 50 | 10043 | 25 | 10044 | 25 | | | | | | | | | | | | | | |
| bread_sliced_spread_polymarg_fat_med | 0% | 100% | 0% | 100% | 0% | 10043 | 50 | 10044 | 50 | | | | | | | | | | | | | | | | |
| bread_sliced_spread_polymarg_fat_thick | 0% | 100% | 0% | 100% | 0% | 10043 | 50 | 10044 | 50 | | | | | | | | | | | | | | | | |
| bread_sliced_spread_polymarg_fat_thin | 0% | 100% | 0% | 100% | 0% | 10043 | 50 | 10044 | 50 | | | | | | | | | | | | | | | | |
| bread_sliced_spread_polymarg_lowfat_med | 0% | 100% | 0% | 100% | 0% | 10049 | 100 | | | | | | | | | | | | | | | | | | |
| bread_sliced_spread_polymarg_lowfat_thick | 0% | 100% | 0% | 100% | 0% | 10049 | 100 | | | | | | | | | | | | | | | | | | |
| bread_sliced_spread_polymarg_lowfat_thin | 0% | 100% | 0% | 100% | 0% | 10049 | 100 | | | | | | | | | | | | | | | | | | |
| bread_sliced_spread_polymarg_vlowfat_med | 0% | 100% | 0% | 100% | 0% | 10049 | 100 | | | | | | | | | | | | | | | | | | |
| bread_sliced_spread_polymarg_vlowfat_thick | 0% | 100% | 0% | 100% | 0% | 10049 | 100 | | | | | | | | | | | | | | | | | | |
| bread_sliced_spread_polymarg_vlowfat_thin | 0% | 100% | 0% | 100% | 0% | 10049 | 100 | | | | | | | | | | | | | | | | | | |
| bread_sliced_spread_soya_chol_med | 0% | 100% | 1% | 100% | 1% | 3848 | 16.6 | 3243 | 16.6 | 2849 | 16.8 | 10786 | 50 | | | | | | | | | | | | |
| bread_sliced_spread_soya_chol_thick | 0% | 100% | 1% | 100% | 1% | 3848 | 16.6 | 3243 | 16.6 | 2849 | 16.8 | 10786 | 50 | | | | | | | | | | | | |
| bread_sliced_spread_soya_chol_thin | 0% | 100% | 1% | 100% | 1% | 3848 | 16.6 | 3243 | 16.6 | 2849 | 16.8 | 10786 | 50 | | | | | | | | | | | | |
| bread_sliced_spread_soya_dunno_med | 0% | 100% | 0% | 100% | 0% | 10980 | 50 | 10786 | 50 | | | | | | | | | | | | | | | | |
| bread_sliced_spread_soya_dunno_thick | 0% | 100% | 0% | 100% | 0% | 10980 | 50 | 10786 | 50 | | | | | | | | | | | | | | | | |
| bread_sliced_spread_soya_dunno_thin | 0% | 100% | 0% | 100% | 0% | 10980 | 50 | 10786 | 50 | | | | | | | | | | | | | | | | |
| bread_sliced_spread_soya_fat_med | 0% | 100% | 0% | 100% | 0% | 10786 | 100 | | | | | | | | | | | | | | | | | | |
| bread_sliced_spread_soya_fat_thick | 0% | 100% | 0% | 100% | 0% | 10786 | 100 | | | | | | | | | | | | | | | | | | |
| bread_sliced_spread_soya_fat_thin | 0% | 100% | 0% | 100% | 0% | 10786 | 100 | | | | | | | | | | | | | | | | | | |
| bread_sliced_spread_soya_lowfat_med | 0% | 100% | 0% | 100% | 0% | 10980 | 100 | | | | | | | | | | | | | | | | | | |
| bread_sliced_spread_soya_lowfat_thick | 0% | 100% | 0% | 100% | 0% | 10980 | 100 | | | | | | | | | | | | | | | | | | |
| bread_sliced_spread_soya_lowfat_thin | 0% | 100% | 0% | 100% | 0% | 10980 | 100 | | | | | | | | | | | | | | | | | | |
| bread_sliced_spread_soya_vlowfat_med | 0% | 100% | 0% | 100% | 0% | 10980 | 100 | | | | | | | | | | | | | | | | | | |
| bread_sliced_spread_soya_vlowfat_thick | 0% | 100% | 0% | 100% | 0% | 10980 | 100 | | | | | | | | | | | | | | | | | | |
| bread_sliced_spread_soya_vlowfat_thin | 0% | 100% | 0% | 100% | 0% | 10980 | 100 | | | | | | | | | | | | | | | | | | |
| bread_sliced_unanswered | 0% | 100% | 0% | 100% | 0% | 120 | 33.4 | 133 | 33.3 | 102 | 33.3 | | | | | | | | | | | | | | |
| bread_sliced_white | 0% | 100% | 0% | 100% | 0% | 120 | 100 | | | | | | | | | | | | | | | | | | |
| bread_sliced_wholemeal | 0% | 100% | 0% | 100% | 0% | 133 | 100 | | | | | | | | | | | | | | | | | | |
| cake | 99% | 30% | 70% | 20% | 80% | 3897 | 20 | 357 | 20 | 3899 | 20 | 6965 | 20 | 6597 | 20 | | | | | | | | | | |
| cake_gf | 99% | 31% | 69% | 20% | 80% | 3897 | 20 | 357 | 20 | 3899 | 20 | 6965 | 20 | 6597 | 20 | | | | | | | | | | |
| cereal_bran | 88% | 100% | 0% | 100% | 0% | 202 | 25 | 7623 | 25 | 201 | 25 | 8482 | 25 | | | | | | | | | | | | |
| cereal_bran_driedfruit | 66% | 100% | 0% | 100% | 0% | 229 | 25 | 5327 | 25 | 7624 | 25 | 203 | 25 | | | | | | | | | | | | |
| cereal_muesli | 38% | 99% | 1% | 99% | 1% | 212 | 50 | 214 | 50 | | | | | | | | | | | | | | | | |
| cereal_muesli_driedfruit | 50% | 99% | 1% | 99% | 1% | 7629 | 50 | 6836 | 50 | | | | | | | | | | | | | | | | |
| cereal_oatcrunch | 81% | 100% | 0% | 100% | 0% | 5328 | 50 | 8712 | 50 | | | | | | | | | | | | | | | | |
| cereal_oatcrunch_driedfruit | 72% | 100% | 0% | 100% | 0% | 5328 | 45 | 8712 | 45 | 2665 | 10 | | | | | | | | | | | | | | |
| cereal_other | 80% | 100% | 0% | 100% | 0% | 7637 | 20 | 8410 | 40 | 8103 | 40 | | | | | | | | | | | | | | |
| cereal_other_driedfruit | 71% | 100% | 0% | 100% | 0% | 7637 | 15 | 8088 | 32.5 | 8103 | 32.5 | 2665 | 20 | | | | | | | | | | | | |
| cereal_plain | 87% | 100% | 0% | 100% | 0% | 223 | 33.4 | 205 | 16.7 | 206 | 16.6 | 220 | 16.7 | 7630 | 16.6 | | | | | | | | | | |
| cereal_plain_driedfruit | 75% | 100% | 0% | 100% | 0% | 223 | 22.7 | 205 | 11.4 | 206 | 11.3 | 220 | 11.3 | 7630 | 11.3 | 2665 | 32 | | | | | | | | |
| cereal_porridge_milk_chol | 0% | 60% | 40% | 70% | 30% | 10514 | 3.5 | 10515 | 3.5 | 9215 | 7 | 7183 | 86 | | | | | | | | | | | | |
| cereal_porridge_milk_chol_driedfruit | 3% | 61% | 39% | 70% | 30% | 10514 | 2.75 | 10515 | 2.75 | 9215 | 5.5 | 2665 | 13 | 7183 | 76 | | | | | | | | | | |
| cereal_porridge_milk_dontknow | 0% | 60% | 40% | 70% | 30% | 10514 | 3.5 | 10515 | 3.5 | 9215 | 7 | 608 | 21.5 | 8543 | 21.5 | 613 | 8.6 | 8544 | 8.6 | 602 | 8.6 | 603 | 8.6 | 10251 | 8.6 |
| cereal_porridge_milk_dontknow_driedfruit | 3% | 61% | 39% | 70% | 30% | 10514 | 2.75 | 10515 | 2.75 | 9215 | 5.5 | 2665 | 13 | 608 | 19 | 8543 | 19 | 613 | 7.6 | 8544 | 7.6 | 602 | 7.6 | 603 | 7.6 | 10251 | 7.6 |
| cereal_porridge_milk_goatsheep | 0% | 60% | 40% | 70% | 30% | 10514 | 3.5 | 10515 | 3.5 | 9215 | 7 | 10397 | 43 | 625 | 21.5 | 626 | 21.5 | | | | | | | | |
| cereal_porridge_milk_goatsheep_driedfruit | 3% | 61% | 39% | 70% | 30% | 10514 | 2.75 | 10515 | 2.75 | 9215 | 5.5 | 2665 | 13 | 10397 | 38 | 625 | 19 | 626 | 19 | | | | | | |
| cereal_porridge_milk_other | 0% | 60% | 40% | 70% | 30% | 10514 | 3.5 | 10515 | 3.5 | 9215 | 7 | 10251 | 23 | 9493 | 14 | 10932 | 14 | 10898 | 35 | | | | | | |
| cereal_porridge_milk_other_driedfruit | 3% | 61% | 39% | 70% | 30% | 10514 | 2.75 | 10515 | 2.75 | 9215 | 5.5 | 2665 | 13 | 10251 | 20 | 9493 | 13 | 10932 | 13 | 10898 | 30 | | | | |
| cereal_porridge_milk_powdered | 0% | 60% | 40% | 70% | 30% | 10514 | 3.5 | 10515 | 3.5 | 9215 | 7 | 695 | 43 | 696 | 43 | | | | | | | | | | |
| cereal_porridge_milk_powdered_driedfruit | 3% | 61% | 39% | 70% | 30% | 10514 | 2.75 | 10515 | 2.75 | 9215 | 5.5 | 2665 | 13 | 695 | 38 | 696 | 38 | | | | | | | | |
| cereal_porridge_milk_riceoatveg | 80% | 100% | 0% | 100% | 0% | 10514 | 3.5 | 10515 | 3.5 | 9215 | 7 | 10572 | 21.5 | 10898 | 21.5 | 10159 | 21.5 | 10836 | 21.5 | | | | | | |
| cereal_porridge_milk_riceoatveg_driedfruit | 83% | 100% | 0% | 100% | 0% | 10514 | 2.75 | 10515 | 2.75 | 9215 | 5.5 | 2665 | 13 | 10572 | 19 | 10898 | 19 | 10159 | 19 | 10836 | 19 | | | | |
| cereal_porridge_milk_semi | 0% | 60% | 40% | 70% | 30% | 10514 | 3.5 | 10515 | 3.5 | 9215 | 7 | 608 | 43 | 8543 | 43 | | | | | | | | | | |
| cereal_porridge_milk_semi_driedfruit | 3% | 61% | 39% | 70% | 30% | 10514 | 2.75 | 10515 | 2.75 | 9215 | 5.5 | 2665 | 13 | 608 | 38 | 8543 | 38 | | | | | | | | |
| cereal_porridge_milk_skimmed | 0% | 60% | 40% | 70% | 30% | 10514 | 3.5 | 10515 | 3.5 | 9215 | 7 | 613 | 43 | 8544 | 43 | | | | | | | | | | |
| cereal_porridge_milk_skimmed_driedfruit | 3% | 61% | 39% | 70% | 30% | 10514 | 2.75 | 10515 | 2.75 | 9215 | 5.5 | 2665 | 13 | 613 | 38 | 8544 | 38 | | | | | | | | |
| cereal_porridge_milk_soya_ca | 80% | 100% | 0% | 100% | 0% | 10514 | 3.5 | 10515 | 3.5 | 9215 | 7 | 8726 | 28.7 | 10974 | 28.7 | 3769 | 28.6 | | | | | | | | |
| cereal_porridge_milk_soya_ca_driedfruit | 83% | 100% | 0% | 100% | 0% | 10514 | 2.75 | 10515 | 2.75 | 9215 | 5.5 | 2665 | 13 | 8726 | 25.4 | 10974 | 25.3 | 3769 | 25.3 | | | | | | |
| cereal_porridge_milk_soya_noca | 80% | 100% | 0% | 100% | 0% | 10514 | 3.5 | 10515 | 3.5 | 9215 | 7 | 8512 | 43 | 650 | 43 | | | | | | | | | | |
| cereal_porridge_milk_soya_noca_driedfruit | 83% | 100% | 0% | 100% | 0% | 10514 | 2.75 | 10515 | 2.75 | 9215 | 5.5 | 2665 | 13 | 8512 | 38 | 650 | 38 | | | | | | | | |

| | | | | | | | | | | | | | | | | | | | | | |
|---|---|---|---|---|---|---|---|---|---|---|---|---|---|---|---|---|---|---|---|---|---|
| cereal_porridge_milk_whole | 0% | 60% | 40% | 70% | 30% | 10514 | 3.5 | 10515 | 3.5 | 9215 | 7 | 602 | 43 | 603 | 43 | | | | | | |
| cereal_porridge_milk_whole_driedfruit | 3% | 61% | 39% | 70% | 30% | 10514 | 2.75 | 10515 | 2.75 | 9215 | 5.5 | 2665 | 13 | 602 | 38 | 603 | 38 | | | | |
| cereal_porridge_water | 0% | 100% | 0% | 100% | 0% | 10514 | 3.5 | 10515 | 3.5 | 9215 | 7 | 5106 | 86 | | | | | | | | |
| cereal_porridge_water_driedfruit | 3% | 100% | 0% | 100% | 0% | 10514 | 2.75 | 10515 | 2.75 | 9215 | 5.5 | 2665 | 13 | 5106 | 76 | | | | | | |
| cereal_sugar | 100% | 75% | 25% | 100% | 0% | 2205 | 25 | 2217 | 25 | 2214 | 25 | 2206 | 25 | | | | | | | | |
| cereal_sweet | 99% | 100% | 0% | 100% | 0% | 9275 | 16.7 | 227 | 8.4 | 7626 | 8.3 | 10512 | 16.7 | 10513 | 16.6 | 204 | 16.7 | 8483 | 16.6 | | |
| cereal_sweet_driedfruit | 72% | 100% | 0% | 100% | 0% | 9275 | 18.3 | 227 | 9.1 | 7626 | 9.1 | 10512 | 18.25 | 10513 | 18.3 | 2665 | 27 | | | | |
| cereal_wwheat | 49% | 100% | 0% | 100% | 0% | 221 | 33.4 | 10510 | 16.7 | 10511 | 16.6 | 225 | 33.3 | | | | | | | | |
| cereal_wwheat_driedfruit | 71% | 100% | 0% | 100% | 0% | 8190 | 25 | 7051 | 25 | 6132 | 50 | | | | | | | | | | |
| cerealbar | 98% | 90% | 10% | 80% | 20% | 6883 | 10 | 10060 | 10 | 10059 | 10 | 10057 | 10 | 10058 | 10 | 7656 | 25 | 8442 | 25 | | |
| cerealbar_gf | 98% | 91% | 9% | 80% | 20% | 6883 | 10 | 10060 | 10 | 10059 | 10 | 10057 | 10 | 10058 | 10 | 7656 | 25 | 8442 | 25 | | |
| cheese_blue | 0% | 0% | 100% | 0% | 100% | 681 | 33.4 | 664 | 33.3 | 679 | 33.3 | | | | | | | | | | |
| cheese_cottage | 0% | 0% | 100% | 0% | 100% | 687 | 33.4 | 7725 | 33.3 | 686 | 33.3 | | | | | | | | | | |
| cheese_feta | 0% | 0% | 100% | 0% | 100% | 669 | 100 | | | | | | | | | | | | | | |
| cheese_goat | 0% | 0% | 100% | 0% | 100% | 7057 | 100 | | | | | | | | | | | | | | |
| cheese_hard | 0% | 0% | 100% | 0% | 100% | 654 | 20 | 665 | 20 | 667 | 20 | 672 | 20 | 676 | 20 | | | | | | |
| cheese_hard_lof | 0% | 0% | 100% | 0% | 100% | 659 | 50 | 7731 | 50 | | | | | | | | | | | | |
| cheese_mozzarella | 0% | 0% | 100% | 0% | 100% | 675 | 100 | | | | | | | | | | | | | | |
| cheese_other | 0% | 0% | 100% | 0% | 100% | 654 | 20 | 651 | 30 | 683 | 30 | 6981 | 10 | 7057 | 10 | | | | | | |
| cheese_soft | 0% | 0% | 100% | 0% | 100% | 691 | 50 | 652 | 50 | | | | | | | | | | | | |
| cheese_spread | 0% | 0% | 100% | 0% | 100% | 684 | 25 | 2703 | 50 | 10413 | 25 | | | | | | | | | | |
| cheese_spread_lof | 0% | 0% | 100% | 0% | 100% | 10503 | 16.7 | 10504 | 16.7 | 10505 | 16.6 | 6979 | 25 | 7112 | 25 | | | | | | |
| cheesecake | 75% | 10% | 90% | 10% | 90% | 509 | 50 | 510 | 50 | | | | | | | | | | | | |
| cheesecake_gf | 75% | 11% | 89% | 10% | 90% | 509 | 50 | 510 | 50 | | | | | | | | | | | | |
| choc_bar | 86% | 30% | 70% | 50% | 50% | 2265 | 34 | 7964 | 33 | 7963 | 33 | | | | | | | | | | |
| choc_dark | 100% | 100% | 0% | 100% | 0% | 2255 | 100 | | | | | | | | | | | | | | |
| choc_milk | 84% | 20% | 80% | 50% | 50% | 2254 | 100 | | | | | | | | | | | | | | |
| choc_sweets | 94% | 20% | 80% | 50% | 50% | 10427 | 100 | | | | | | | | | | | | | | |
| choc_white | 82% | 20% | 80% | 50% | 50% | 7960 | 100 | | | | | | | | | | | | | | |
| chocyog_raisin | 65% | 30% | 70% | 50% | 50% | 7958 | 50 | 9274 | 50 | | | | | | | | | | | | |
| chutney | 92% | 100% | 0% | 100% | 0% | 2417 | 25 | 2440 | 50 | 2418 | 25 | | | | | | | | | | |
| cof_capp | 0% | 100% | 0% | 100% | 0% | 2307 | 100 | | | | | | | | | | | | | | |
| cof_capp_decaf | 0% | 100% | 0% | 100% | 0% | 8312 | 100 | | | | | | | | | | | | | | |
| cof_capp_milk_chol | 0% | 0% | 100% | 0% | 100% | 7183 | 92.5 | 5103 | 7.5 | | | | | | | | | | | | |
| cof_capp_milk_dontknow | 0% | 0% | 100% | 0% | 100% | 608 | 23.13 | 8543 | 23.1 | 613 | 9.25 | 8544 | 9.25 | 602 | 9.25 | 603 | 9.25 | 10251 | 9.25 | 5103 | 7.5 |
| cof_capp_milk_goatsheep | 0% | 0% | 100% | 0% | 100% | 625 | 23.13 | 626 | 23.1 | 10397 | 46.25 | 5103 | 7.5 | | | | | | | | |
| cof_capp_milk_other | 0% | 0% | 100% | 0% | 100% | 10251 | 46.25 | 10932 | 23.1 | 9493 | 23.125 | 5103 | 7.5 | | | | | | | | |
| cof_capp_milk_powdered | 0% | 0% | 100% | 0% | 100% | 695 | 46.25 | 696 | 46.3 | 5103 | 7.5 | | | | | | | | | | |
| cof_capp_milk_riceoatveg | 100% | 100% | 0% | 100% | 0% | 11150 | 11.1 | 10572 | 11.1 | 10898 | 23.125 | 10159 | 12.03 | 10966 | 12 | 10836 | 23.1 | 5103 | 7.5 | | |
| cof_capp_milk_semi | 0% | 0% | 100% | 0% | 100% | 608 | 46.25 | 8543 | 46.3 | 5103 | 7.5 | | | | | | | | | | |
| cof_capp_milk_skimmed | 0% | 0% | 100% | 0% | 100% | 613 | 46.25 | 8544 | 46.3 | 5103 | 7.5 | | | | | | | | | | |
| cof_capp_milk_soya_ca | 100% | 100% | 0% | 100% | 0% | 8726 | 31.45 | 10974 | 30.5 | 3769 | 30.525 | 5103 | 7.5 | | | | | | | | |
| cof_capp_milk_soya_noca | 100% | 100% | 0% | 100% | 0% | 8512 | 46.25 | 650 | 46.3 | 5103 | 7.5 | | | | | | | | | | |
| cof_capp_milk_whole | 0% | 0% | 100% | 0% | 100% | 602 | 46.25 | 603 | 46.3 | 5103 | 7.5 | | | | | | | | | | |
| cof_espresso | 0% | 100% | 0% | 100% | 0% | 2307 | 100 | | | | | | | | | | | | | | |
| cof_espresso_decaf | 0% | 100% | 0% | 100% | 0% | 8312 | 100 | | | | | | | | | | | | | | |
| cof_filter | 0% | 100% | 0% | 100% | 0% | 2307 | 50 | 2306 | 50 | | | | | | | | | | | | |
| cof_filter_decaf | 0% | 100% | 0% | 100% | 0% | 8312 | 50 | 8313 | 50 | | | | | | | | | | | | |
| cof_instant | 0% | 100% | 0% | 100% | 0% | 2308 | 0.9 | 5103 | 99.1 | | | | | | | | | | | | |
| cof_instant_decaf | 0% | 100% | 0% | 100% | 0% | 8314 | 0.9 | 5103 | 99.1 | | | | | | | | | | | | |
| cof_latte | 0% | 100% | 0% | 100% | 0% | 2307 | 100 | | | | | | | | | | | | | | |
| cof_latte_decaf | 0% | 100% | 0% | 100% | 0% | 8312 | 100 | | | | | | | | | | | | | | |
| cof_latte_milk_chol | 0% | 0% | 100% | 0% | 100% | 7183 | 96.2 | 5103 | 3.8 | | | | | | | | | | | | |
| cof_latte_milk_dontknow | 0% | 0% | 100% | 0% | 100% | 608 | 24.05 | 8543 | 24.1 | 613 | 9.62 | 8544 | 9.62 | 602 | 9.62 | 603 | 9.62 | 10251 | 9.62 | 5103 | 3.8 |
| cof_latte_milk_goatsheep | 0% | 0% | 100% | 0% | 100% | 625 | 24.05 | 626 | 24.1 | 10397 | 48.1 | 5103 | 3.8 | | | | | | | | |
| cof_latte_milk_other | 0% | 0% | 100% | 0% | 100% | 10251 | 48.1 | 10932 | 24.1 | 9493 | 24.05 | 5103 | 3.8 | | | | | | | | |
| cof_latte_milk_powdered | 0% | 0% | 100% | 0% | 100% | 695 | 48.1 | 696 | 48.1 | 5103 | 3.8 | | | | | | | | | | |
| cof_latte_milk_riceoatveg | 100% | 100% | 0% | 100% | 0% | 11150 | 11.54 | 10572 | 11.5 | 10898 | 24.05 | 10159 | 12.51 | 10966 | 12.5 | 10836 | 24.1 | 5103 | 3.8 | | |
| cof_latte_milk_semi | 0% | 0% | 100% | 0% | 100% | 608 | 48.1 | 8543 | 48.1 | 5103 | 3.8 | | | | | | | | | | |
| cof_latte_milk_skimmed | 0% | 0% | 100% | 0% | 100% | 613 | 48.1 | 8544 | 48.1 | 5103 | 3.8 | | | | | | | | | | |
| cof_latte_milk_soya_ca | 100% | 100% | 0% | 100% | 0% | 8726 | 32.71 | 10974 | 31.7 | 3769 | 31.746 | 5103 | 3.8 | | | | | | | | |
| cof_latte_milk_soya_noca | 100% | 100% | 0% | 100% | 0% | 8512 | 48.1 | 650 | 48.1 | 5103 | 3.8 | | | | | | | | | | |
| cof_latte_milk_whole | 0% | 0% | 100% | 0% | 100% | 602 | 48.1 | 603 | 48.1 | 5103 | 3.8 | | | | | | | | | | |
| cof_other | 0% | 100% | 0% | 100% | 0% | 2308 | 0.45 | 5103 | 49.6 | 2307 | 25 | 2306 | 25 | | | | | | | | |
| cof_other_decaf | 0% | 100% | 0% | 100% | 0% | 8314 | 0.45 | 5103 | 49.6 | 8312 | 25 | 8313 | 25 | | | | | | | | |
| cof_sugar | 100% | 100% | 0% | 100% | 0% | 2205 | 66.7 | 2203 | 33.3 | | | | | | | | | | | | |
| cream | 0% | 0% | 100% | 0% | 100% | 638 | 16.7 | 634 | 16.7 | 633 | 16.6 | 3014 | 16.7 | 632 | 16.7 | 644 | 16.6 | | | | |
| croissant | 0% | 20% | 80% | 0% | 100% | 166 | 100 | | | | | | | | | | | | | | |
| crumble | 20% | 70% | 30% | 20% | 80% | 15 | 38 | 24 | 10 | 2205 | 20 | 852 | 29 | 654 | 3 | | | | | | |
| danish_pastry | 76% | 30% | 70% | 20% | 80% | 318 | 100 | | | | | | | | | | | | | | |
| dessert_milkbased | 96% | 20% | 80% | 20% | 80% | 582 | 50 | 7695 | 25 | 9533 | 25 | | | | | | | | | | |
| dessert_milkpuds | 67% | 20% | 80% | 10% | 90% | 546 | 17 | 548 | 17 | 559 | 33 | 9636 | 33 | | | | | | | | |
| dessert_other | 84% | 20% | 80% | 10% | 90% | 6967 | 40 | 1959 | 22.5 | 2150 | 22.5 | 574 | 7.5 | 573 | 7.5 | | | | | | |
| dessert_soya | 79% | 100% | 0% | 100% | 0% | 9115 | 17 | 10575 | 17 | 6541 | 33 | 9053 | 33 | | | | | | | | |
| double_crust | 0% | 60% | 40% | 5% | 95% | 400 | 90 | 405 | 10 | | | | | | | | | | | | |
| doughnut | 99% | 20% | 80% | 40% | 60% | 323 | 50 | 7678 | 25 | 324 | 25 | | | | | | | | | | |
| drink_diethotchoc | 100% | 1% | 99% | 3% | 97% | 2633 | 2.75 | 7888 | 2.75 | 5106 | 94.5 | | | | | | | | | | |
| drink_fizzy | 100% | 100% | 0% | 100% | 0% | 7895 | 33.4 | 8328 | 33.3 | 2322 | 33.3 | | | | | | | | | | |
| drink_grapefruit | 100% | 100% | 0% | 100% | 0% | 2329 | 50 | 2330 | 50 | | | | | | | | | | | | |
| drink_hotchoc | 100% | 1% | 99% | 3% | 97% | 2309 | 6.2 | 2632 | 6 | 2310 | 6 | 5106 | 81.8 | | | | | | | | |
| drink_hotchoc_milk_chol | 0% | 0% | 100% | 0% | 100% | 7183 | 100 | | | | | | | | | | | | | | |
| drink_hotchoc_milk_dontknow | 0% | 0% | 100% | 0% | 100% | 608 | 25 | 8543 | 25 | 613 | 10 | 8544 | 10 | 602 | 10 | 603 | 10 | 10251 | 10 | | |
| drink_hotchoc_milk_goatsheep | 0% | 0% | 100% | 0% | 100% | 10397 | 50 | 625 | 25 | 626 | 25 | | | | | | | | | | |
| drink_hotchoc_milk_other | 0% | 0% | 100% | 0% | 100% | 10251 | 50 | 9493 | 25 | 10932 | 25 | | | | | | | | | | |
| drink_hotchoc_milk_powdered | 0% | 0% | 100% | 0% | 100% | 695 | 50 | 696 | 50 | | | | | | | | | | | | |
| drink_hotchoc_milk_riceoatveg | 100% | 100% | 0% | 100% | 0% | 10572 | 25 | 10898 | 25 | 10159 | 25 | 10836 | 25 | | | | | | | | |
| drink_hotchoc_milk_semi | 0% | 0% | 100% | 0% | 100% | 608 | 50 | 8543 | 50 | | | | | | | | | | | | |
| drink_hotchoc_milk_skimmed | 0% | 0% | 100% | 0% | 100% | 613 | 50 | 8544 | 50 | | | | | | | | | | | | |
| drink_hotchoc_milk_soya_ca | 100% | 100% | 0% | 100% | 0% | 8726 | 34 | 10974 | 33 | 3769 | 33 | | | | | | | | | | |
| drink_hotchoc_milk_soya_noca | 100% | 100% | 0% | 100% | 0% | 8512 | 50 | 650 | 50 | | | | | | | | | | | | |
| drink_hotchoc_milk_whole | 0% | 0% | 100% | 0% | 100% | 602 | 50 | 603 | 50 | | | | | | | | | | | | |
| drink_lowcal | 0% | 100% | 0% | 100% | 0% | 7897 | 50 | 7902 | 25 | 2351 | 5 | 5102 | 20 | | | | | | | | |
| drink_milkbased | 64% | 1% | 99% | 2% | 98% | 711 | 20 | 8212 | 20 | 629 | 20 | 612 | 20 | 11128 | 0.6 | 11127 | 0.6 | 608 | 9.4 | 8543 | 9.4 |
| drink_orange | 100% | 100% | 0% | 100% | 0% | 2337 | 33.4 | 2338 | 33.3 | 2339 | 33.3 | | | | | | | | | | |
| drink_other | 33% | 3% | 97% | 1% | 99% | 8333 | 35 | 608 | 15 | 8543 | 15 | 7053 | 0.09 | 5103 | 9.91 | 2338 | 12.5 | 2358 | 12.5 | | |
| drink_purejuice | 100% | 100% | 0% | 100% | 0% | 2319 | 20 | 2318 | 20 | 2345 | 20 | 2355 | 20 | 3846 | 20 | | | | | | |
| drink_squash | 100% | 100% | 0% | 100% | 0% | 9997 | 5 | 5101 | 60 | 2358 | 25 | 7913 | 5 | 2331 | 5 | | | | | | |
| drink_water | 0% | 100% | 0% | 100% | 0% | 5000 | 80 | 8333 | 20 | | | | | | | | | | | | |
| drizzle_oil | 0% | 100% | 0% | 100% | 0% | 874 | 100 | | | | | | | | | | | | | | |
| egg_omelet | 0% | 0% | 100% | 0% | 100% | 7765 | 25 | 9303 | 25 | 763 | 50 | | | | | | | | | | |
| egg_other | 0% | 10% | 90% | 10% | 90% | 8711 | 50 | 813 | 25 | 9861 | 25 | | | | | | | | | | |
| egg_scotch | 0% | 10% | 90% | 10% | 90% | 814 | 100 | | | | | | | | | | | | | | |
| egg_swich | 0% | 10% | 90% | 10% | 90% | 8066 | 50 | 755 | 45 | 2432 | 5 | | | | | | | | | | |
| egg_whole | 0% | 0% | 100% | 0% | 100% | 755 | 33.4 | 762 | 33.3 | 756 | 33.3 | | | | | | | | | | |
| fish_battered | 0% | 5% | 95% | 90% | 10% | 1415 | 50 | 7808 | 50 | | | | | | | | | | | | |
| fish_breaded | 0% | 5% | 95% | 50% | 50% | 11006 | 16.5 | 1606 | 16.5 | 9524 | 34 | 9574 | 33 | | | | | | | | |
| fish_lobcrab | 0% | 0% | 100% | 0% | 100% | 1561 | 50 | 1565 | 50 | | | | | | | | | | | | |
| fish_oily | 0% | 0% | 100% | 0% | 100% | 1495 | 10 | 1502 | 10 | 1514 | 10 | 9541 | 30 | 1523 | 10 | 9267 | 20 | 9932 | 10 | | |
| fish_other | 0% | 0% | 100% | 15% | 85% | 1522 | 60 | 10357 | 5 | 1639 | 15 | 11004 | 10 | 7819 | 10 | | | | | | |
| fish_prawns | 0% | 0% | 100% | 0% | 100% | 1573 | 100 | | | | | | | | | | | | | | |
| fish_shell | 0% | 0% | 100% | 0% | 100% | 1576 | 30 | 1568 | 70 | | | | | | | | | | | | |
| fish_tinnedtuna | 0% | 0% | 100% | 0% | 100% | 1534 | 50 | 1533 | 50 | | | | | | | | | | | | |
| fish_white | 0% | 0% | 100% | 0% | 100% | 7798 | 50 | 7799 | 50 | | | | | | | | | | | | |
| fruit_apple | 0% | 100% | 0% | 100% | 0% | 2601 | 100 | | | | | | | | | | | | | | |
| fruit_banana | 0% | 100% | 0% | 100% | 0% | 1977 | 100 | | | | | | | | | | | | | | |
| fruit_berry | 0% | 100% | 0% | 100% | 0% | 2143 | 25 | 2153 | 25 | 2001 | 5 | 11115 | 25 | 1985 | 20 | | | | | | |
| fruit_cherry | 0% | 100% | 0% | 100% | 0% | 1991 | 100 | | | | | | | | | | | | | | |
| fruit_dried | 0% | 100% | 0% | 100% | 0% | 8547 | 12.5 | 1960 | 12.5 | 2142 | 50 | 2021 | 12.5 | 2024 | 12.5 | | | | | | |
| fruit_grapefruit | 0% | 100% | 0% | 100% | 0% | 2045 | 70 | 2049 | 30 | | | | | | | | | | | | |
| fruit_grapes | 0% | 100% | 0% | 100% | 0% | 2041 | 50 | 2043 | 50 | | | | | | | | | | | | |
| fruit_mango | 0% | 100% | 0% | 100% | 0% | 2078 | 100 | | | | | | | | | | | | | | |
| fruit_melon | 0% | 100% | 0% | 100% | 0% | 2081 | 33.4 | 2083 | 33.3 | 2085 | 33.3 | | | | | | | | | | |
| fruit_mixed | 0% | 100% | 0% | 100% | 0% | 2035 | 35 | 2030 | 30 | 5954 | 35 | | | | | | | | | | |
| fruit_orange | 0% | 100% | 0% | 100% | 0% | 2092 | 100 | | | | | | | | | | | | | | |
| fruit_other | 0% | 100% | 0% | 100% | 0% | 2061 | 60 | 2099 | 10 | 8115 | 10 | 1963 | 10 | 2023 | 10 | | | | | | |
| fruit_peach | 0% | 100% | 0% | 100% | 0% | 2101 | 50 | 2088 | 50 | | | | | | | | | | | | |
| fruit_pear | 0% | 100% | 0% | 100% | 0% | 2241 | 100 | | | | | | | | | | | | | | |
| fruit_pineapple | 0% | 100% | 0% | 100% | 0% | 2117 | 70 | 2121 | 30 | | | | | | | | | | | | |
| fruit_plum | 0% | 100% | 0% | 100% | 0% | 2123 | 100 | | | | | | | | | | | | | | |
| fruit_prunes | 0% | 100% | 0% | 100% | 0% | 8558 | 100 | | | | | | | | | | | | | | |
| fruit_satsuma | 0% | 100% | 0% | 100% | 0% | 2157 | 100 | | | | | | | | | | | | | | |
| fruit_stewed | 0% | 100% | 0% | 100% | 0% | 1958 | 33.4 | 2150 | 33.3 | 2129 | 33.3 | | | | | | | | | | |
| fruitcake | 71% | 30% | 70% | 20% | 80% | 333 | 50 | 8105 | 50 | | | | | | | | | | | | |
| fruitcake_gf | 71% | 31% | 69% | 20% | 80% | 333 | 50 | 8105 | 50 | | | | | | | | | | | | |
| grains_couscous | 0% | 100% | 0% | 100% | 0% | 7600 | 50 | 8466 | 50 | | | | | | | | | | | | |
| grains_other | 0% | 100% | 0% | 100% | 0% | 5 | 20 | 7028 | 35 | 10424 | 35 | 10017 | 10 | | | | | | | | |
| guacamole | 0% | 90% | 10% | 90% | 10% | 8121 | 75 | 8126 | 25 | | | | | | | | | | | | |
| hummus | 0% | 100% | 0% | 100% | 0% | 1817 | 75 | 2978 | 25 | | | | | | | | | | | | |
| icecream | 76% | 0% | 100% | 60% | 40% | 720 | 33.4 | 6970 | 16.7 | 730 | 33.3 | 726 | 16.6 | | | | | | | | |
| indian_snack | 0% | 70% | 30% | 20% | 80% | 1797 | 50 | 1354 | 25 | 2623 | 25 | | | | | | | | | | |
| jam_honey | 97% | 80% | 20% | 100% | 0% | 2217 | 35 | 2214 | 20 | 2206 | 10 | 8559 | 35 | | | | | | | | |
| mayo | 100% | 20% | 80% | 80% | 20% | 2432 | 50 | 2441 | 50 | | | | | | | | | | | | |
| mayo_lowfat | 100% | 20% | 80% | 80% | 20% | 2433 | 50 | 10805 | 50 | | | | | | | | | | | | |
| meat_bacon_nofat | 0% | 0% | 100% | 0% | 100% | 9464 | 100 | | | | | | | | | | | | | | |
| meat_bacon_withfat | 0% | 0% | 100% | 0% | 100% | 914 | 33.4 | 915 | 33.3 | 916 | 33.3 | | | | | | | | | | |
| meat_beef_nofat | 0% | 0% | 100% | 0% | 100% | 953 | 25 | 970 | 25 | 942 | 25 | 8264 | 25 | | | | | | | | |
| meat_beef_withfat | 0% | 0% | 100% | 1% | 99% | 952 | 25 | 969 | 25 | 3829 | 25 | 8264 | 25 | | | | | | | | |
| meat_ham_nofat | 0% | 0% | 100% | 0% | 100% | 9508 | 100 | | | | | | | | | | | | | | |
| meat_ham_withfat | 0% | 0% | 100% | 0% | 100% | 8089 | 80 | 1274 | 10 | 8043 | 10 | | | | | | | | | | |
| meat_lamb_nofat | 0% | 0% | 100% | 0% | 100% | 980 | 30 | 9434 | 30 | 1003 | 20 | 5217 | 20 | | | | | | | | |
| meat_lamb_withfat | 0% | 0% | 100% | 0% | 100% | 982 | 30 | 9433 | 30 | 1002 | 20 | 5217 | 20 | | | | | | | | |
| meat_liverpate | 0% | 0% | 100% | 0% | 100% | 1191 | 20 | 1204 | 20 | 1258 | 60 | | | | | | | | | | |
| meat_other | 0% | 0% | 100% | 0% | 100% | 1131 | 64 | 1167 | 25 | 1139 | 11 | | | | | | | | | | |
| meat_pork_nofat | 0% | 0% | 100% | 0% | 100% | 1024 | 33.4 | 9451 | 33.3 | 9460 | 33.3 | | | | | | | | | | |
| meat_pork_withfat | 0% | 0% | 100% | 0% | 100% | 1026 | 33.4 | 9450 | 33.3 | 9460 | 33.3 | | | | | | | | | | |
| meat_sausage | 0% | 1% | 99% | 1% | 99% | 7791 | 25 | 1280 | 75 | | | | | | | | | | | | |
| milk_chol_cereal | 0% | 0% | 100% | 0% | 100% | 7183 | 100 | | | | | | | | | | | | | | |
| milk_chol_coffee | 0% | 0% | 100% | 0% | 100% | 7183 | 100 | | | | | | | | | | | | | | |
| milk_chol_glass | 0% | 0% | 100% | 0% | 100% | 7183 | 100 | | | | | | | | | | | | | | |
| milk_chol_tea | 0% | 0% | 100% | 0% | 100% | 7183 | 100 | | | | | | | | | | | | | | |
| milk_dontknow_cereal | 0% | 0% | 100% | 0% | 100% | 608 | 25 | 8543 | 25 | 613 | 10 | 8544 | 10 | 602 | 10 | 603 | 10 | 10251 | 10 | | |
| milk_dontknow_coffee | 0% | 0% | 100% | 0% | 100% | 608 | 25 | 8543 | 25 | 613 | 10 | 8544 | 10 | 602 | 10 | 603 | 10 | 10251 | 10 | | |
| milk_dontknow_glass | 0% | 0% | 100% | 0% | 100% | 608 | 25 | 8543 | 25 | 613 | 10 | 8544 | 10 | 602 | 10 | 603 | 10 | 10251 | 10 | | |
| milk_dontknow_tea | 0% | 0% | 100% | 0% | 100% | 608 | 25 | 8543 | 25 | 613 | 10 | 8544 | 10 | 602 | 10 | 603 | 10 | 10251 | 10 | | |
| milk_goatsheep_cereal | 0% | 0% | 100% | 0% | 100% | 623 | 16.7 | 624 | 16.7 | 10397 | 33.3 | 10416 | 33.3 | | | | | | | | |
| milk_goatsheep_coffee | 0% | 0% | 100% | 0% | 100% | 623 | 12.5 | 624 | 12.5 | 10397 | 25 | 10416 | 25 | 625 | 12.5 | 626 | 12.5 | | | | |
| milk_goatsheep_glass | 0% | 0% | 100% | 0% | 100% | 623 | 12.5 | 624 | 12.5 | 10397 | 25 | 10416 | 25 | 625 | 12.5 | 626 | 12.5 | | | | |
| milk_goatsheep_tea | 0% | 0% | 100% | 0% | 100% | 623 | 12.5 | 624 | 12.5 | 10397 | 25 | 10416 | 25 | 625 | 12.5 | 626 | 12.5 | | | | |
| milk_other_cereal | 0% | 0% | 100% | 0% | 100% | 10251 | 27 | 10898 | 40 | 9493 | 16.5 | 10932 | 16.5 | | | | | | | | |

| | | | | | | | | | | | | | | | | | | | | | |
|---|---|---|---|---|---|---|---|---|---|---|---|---|---|---|---|---|---|---|---|---|---|
| milk_other_coffee | 0% | 0% | 100% | 0% | 100% | 10251 | 27 | 10898 | 40 | 9493 | 16.5 | 10932 | 16.5 | | | | | | | | |
| milk_other_glass | 0% | 0% | 100% | 0% | 100% | 10251 | 27 | 10898 | 40 | 9493 | 16.5 | 10932 | 16.5 | | | | | | | | |
| milk_other_tea | 0% | 0% | 100% | 0% | 100% | 10251 | 27 | 10898 | 40 | 9493 | 16.5 | 10932 | 16.5 | | | | | | | | |
| milk_powdered_cereal | 0% | 0% | 100% | 0% | 100% | 695 | 50 | 696 | 50 | | | | | | | | | | | | |
| milk_powdered_coffee | 0% | 0% | 100% | 0% | 100% | 8149 | 25 | 10498 | 25 | 7717 | 25 | 8213 | 25 | | | | | | | | |
| milk_powdered_glass | 0% | 0% | 100% | 0% | 100% | 695 | 50 | 696 | 50 | | | | | | | | | | | | |
| milk_powdered_tea | 0% | 0% | 100% | 0% | 100% | 8149 | 50 | 10498 | 50 | | | | | | | | | | | | |
| milk_riceoatveg_cereal | 100% | 100% | 0% | 100% | 0% | 11150 | 16.7 | 10898 | 16.7 | 10572 | 16.7 | 10159 | 16.7 | 10966 | 16.6 | 10836 | 16.6 | | | | |
| milk_riceoatveg_coffee | 100% | 100% | 0% | 100% | 0% | 11150 | 16.7 | 10898 | 16.7 | 10572 | 16.7 | 10159 | 16.7 | 10966 | 16.6 | 10836 | 16.6 | | | | |
| milk_riceoatveg_glass | 100% | 100% | 0% | 100% | 0% | 11150 | 16.7 | 10898 | 16.7 | 10572 | 16.7 | 10159 | 16.7 | 10966 | 16.6 | 10836 | 16.6 | | | | |
| milk_riceoatveg_tea | 100% | 100% | 0% | 100% | 0% | 11150 | 16.7 | 10898 | 16.7 | 10572 | 16.7 | 10159 | 16.7 | 10966 | 16.6 | 10836 | 16.6 | | | | |
| milk_semi_cereal | 0% | 0% | 100% | 0% | 100% | 608 | 50 | 8543 | 50 | | | | | | | | | | | | |
| milk_semi_coffee | 0% | 0% | 100% | 0% | 100% | 608 | 50 | 8543 | 50 | | | | | | | | | | | | |
| milk_semi_glass | 0% | 0% | 100% | 0% | 100% | 608 | 50 | 8543 | 50 | | | | | | | | | | | | |
| milk_semi_tea | 0% | 0% | 100% | 0% | 100% | 608 | 50 | 8543 | 50 | | | | | | | | | | | | |
| milk_skimmed_cereal | 0% | 0% | 100% | 0% | 100% | 613 | 50 | 8544 | 50 | | | | | | | | | | | | |
| milk_skimmed_coffee | 0% | 0% | 100% | 0% | 100% | 613 | 50 | 8544 | 50 | | | | | | | | | | | | |
| milk_skimmed_glass | 0% | 0% | 100% | 0% | 100% | 613 | 50 | 8544 | 50 | | | | | | | | | | | | |
| milk_skimmed_tea | 0% | 0% | 100% | 0% | 100% | 613 | 50 | 8544 | 50 | | | | | | | | | | | | |
| milk_soya_ca_cereal | 100% | 100% | 0% | 100% | 0% | 8726 | 34 | 10974 | 33 | 3769 | 33 | | | | | | | | | | |
| milk_soya_ca_coffee | 100% | 100% | 0% | 100% | 0% | 8726 | 34 | 10974 | 33 | 3769 | 33 | | | | | | | | | | |
| milk_soya_ca_glass | 100% | 100% | 0% | 100% | 0% | 8726 | 34 | 10974 | 33 | 3769 | 33 | | | | | | | | | | |
| milk_soya_ca_tea | 100% | 100% | 0% | 100% | 0% | 8726 | 34 | 10974 | 33 | 3769 | 33 | | | | | | | | | | |
| milk_soya_noca_cereal | 100% | 100% | 0% | 100% | 0% | 8512 | 50 | 650 | 50 | | | | | | | | | | | | |
| milk_soya_noca_coffee | 100% | 100% | 0% | 100% | 0% | 8512 | 50 | 650 | 50 | | | | | | | | | | | | |
| milk_soya_noca_glass | 100% | 100% | 0% | 100% | 0% | 8512 | 50 | 650 | 50 | | | | | | | | | | | | |
| milk_soya_noca_tea | 100% | 100% | 0% | 100% | 0% | 8512 | 50 | 650 | 50 | | | | | | | | | | | | |
| milk_whole_cereal | 0% | 0% | 100% | 0% | 100% | 602 | 50 | 603 | 50 | | | | | | | | | | | | |
| milk_whole_coffee | 0% | 0% | 100% | 0% | 100% | 602 | 50 | 603 | 50 | | | | | | | | | | | | |
| milk_whole_glass | 0% | 0% | 100% | 0% | 100% | 602 | 50 | 603 | 50 | | | | | | | | | | | | |
| milk_whole_tea | 0% | 0% | 100% | 0% | 100% | 602 | 50 | 603 | 50 | | | | | | | | | | | | |
| oatcakes | 0% | 100% | 0% | 100% | 0% | 267 | 100 | | | | | | | | | | | | | | |
| oatcakes_spread_butter_dunno_med | 0% | 20% | 80% | 20% | 80% | 9407 | 25 | 10039 | 25 | 851 | 25 | 852 | 25 | | | | | | | | |
| oatcakes_spread_butter_dunno_thick | 0% | 20% | 80% | 20% | 80% | 9407 | 25 | 10039 | 25 | 851 | 25 | 852 | 25 | | | | | | | | |
| oatcakes_spread_butter_dunno_thin | 0% | 20% | 80% | 20% | 80% | 9407 | 25 | 10039 | 25 | 851 | 25 | 852 | 25 | | | | | | | | |
| oatcakes_spread_butter_fat_med | 0% | 0% | 100% | 0% | 100% | 851 | 50 | 852 | 50 | | | | | | | | | | | | |
| oatcakes_spread_butter_fat_thick | 0% | 0% | 100% | 0% | 100% | 851 | 50 | 852 | 50 | | | | | | | | | | | | |
| oatcakes_spread_butter_fat_thin | 0% | 0% | 100% | 0% | 100% | 851 | 50 | 852 | 50 | | | | | | | | | | | | |
| oatcakes_spread_butter_lowfat_med | 0% | 45% | 55% | 45% | 55% | 10140 | 100 | | | | | | | | | | | | | | |
| oatcakes_spread_butter_lowfat_thick | 0% | 45% | 55% | 45% | 55% | 10140 | 100 | | | | | | | | | | | | | | |
| oatcakes_spread_butter_lowfat_thin | 0% | 45% | 55% | 45% | 55% | 10140 | 100 | | | | | | | | | | | | | | |
| oatcakes_spread_butter_spread_fat_med | 0% | 40% | 60% | 40% | 60% | 9407 | 50 | 10039 | 50 | | | | | | | | | | | | |
| oatcakes_spread_butter_spread_fat_thick | 0% | 40% | 60% | 40% | 60% | 9407 | 50 | 10039 | 50 | | | | | | | | | | | | |
| oatcakes_spread_butter_spread_fat_thin | 0% | 40% | 60% | 40% | 60% | 9407 | 50 | 10039 | 50 | | | | | | | | | | | | |
| oatcakes_spread_butter_spread_lowfat_med | 0% | 45% | 55% | 45% | 55% | 3891 | 50 | 10894 | 50 | | | | | | | | | | | | |
| oatcakes_spread_butter_spread_lowfat_thick | 0% | 45% | 55% | 45% | 55% | 3891 | 50 | 10894 | 50 | | | | | | | | | | | | |
| oatcakes_spread_butter_spread_lowfat_thin | 0% | 45% | 55% | 45% | 55% | 3891 | 50 | 10894 | 50 | | | | | | | | | | | | |
| oatcakes_spread_dairy_chol_med | 0% | 97% | 3% | 97% | 3% | 3848 | 100 | | | | | | | | | | | | | | |
| oatcakes_spread_dairy_chol_thick | 0% | 97% | 3% | 97% | 3% | 3848 | 100 | | | | | | | | | | | | | | |
| oatcakes_spread_dairy_chol_thin | 0% | 97% | 3% | 97% | 3% | 3848 | 100 | | | | | | | | | | | | | | |
| oatcakes_spread_dairy_dunno_med | 0% | 97% | 3% | 97% | 3% | 10047 | 50 | 7775 | 50 | | | | | | | | | | | | |
| oatcakes_spread_dairy_dunno_thick | 0% | 97% | 3% | 97% | 3% | 10047 | 50 | 7775 | 50 | | | | | | | | | | | | |
| oatcakes_spread_dairy_dunno_thin | 0% | 97% | 3% | 97% | 3% | 10047 | 50 | 7775 | 50 | | | | | | | | | | | | |
| oatcakes_spread_dairy_fat_med | 0% | 97% | 3% | 97% | 3% | 7775 | 100 | | | | | | | | | | | | | | |
| oatcakes_spread_dairy_fat_thick | 0% | 97% | 3% | 97% | 3% | 7775 | 100 | | | | | | | | | | | | | | |
| oatcakes_spread_dairy_fat_thin | 0% | 97% | 3% | 97% | 3% | 7775 | 100 | | | | | | | | | | | | | | |
| oatcakes_spread_dairy_lowfat_med | 0% | 99% | 1% | 99% | 1% | 10047 | 100 | | | | | | | | | | | | | | |
| oatcakes_spread_dairy_lowfat_thick | 0% | 99% | 1% | 99% | 1% | 10047 | 100 | | | | | | | | | | | | | | |
| oatcakes_spread_dairy_lowfat_thin | 0% | 99% | 1% | 99% | 1% | 10047 | 100 | | | | | | | | | | | | | | |
| oatcakes_spread_dairy_vlowfat_med | 0% | 99% | 1% | 99% | 1% | 10047 | 100 | | | | | | | | | | | | | | |
| oatcakes_spread_dairy_vlowfat_thick | 0% | 99% | 1% | 99% | 1% | 10047 | 100 | | | | | | | | | | | | | | |
| oatcakes_spread_dairy_vlowfat_thin | 0% | 99% | 1% | 99% | 1% | 10047 | 100 | | | | | | | | | | | | | | |
| oatcakes_spread_dunno_chol_med | 0% | 97% | 3% | 97% | 3% | 3848 | 33.4 | 3243 | 33.3 | 2849 | 33.3 | | | | | | | | | | |
| oatcakes_spread_dunno_chol_thick | 0% | 97% | 3% | 97% | 3% | 3848 | 33.4 | 3243 | 33.3 | 2849 | 33.3 | | | | | | | | | | |
| oatcakes_spread_dunno_chol_thin | 0% | 97% | 3% | 97% | 3% | 3848 | 33.4 | 3243 | 33.3 | 2849 | 33.3 | | | | | | | | | | |
| oatcakes_spread_dunno_dunno_med | 0% | 97% | 3% | 97% | 3% | 10047 | 25 | 10049 | 25 | 10043 | 25 | 7775 | 25 | | | | | | | | |
| oatcakes_spread_dunno_dunno_thick | 0% | 97% | 3% | 97% | 3% | 10047 | 25 | 10049 | 25 | 10043 | 25 | 7775 | 25 | | | | | | | | |
| oatcakes_spread_dunno_dunno_thin | 0% | 97% | 3% | 97% | 3% | 10047 | 25 | 10049 | 25 | 10043 | 25 | 7775 | 25 | | | | | | | | |
| oatcakes_spread_dunno_fat_med | 0% | 97% | 3% | 97% | 3% | 10043 | 50 | 7775 | 50 | | | | | | | | | | | | |
| oatcakes_spread_dunno_fat_thick | 0% | 97% | 3% | 97% | 3% | 10043 | 50 | 7775 | 50 | | | | | | | | | | | | |
| oatcakes_spread_dunno_fat_thin | 0% | 97% | 3% | 97% | 3% | 10043 | 50 | 7775 | 50 | | | | | | | | | | | | |
| oatcakes_spread_dunno_lowfat_med | 0% | 97% | 3% | 97% | 3% | 10047 | 50 | 10049 | 50 | | | | | | | | | | | | |
| oatcakes_spread_dunno_lowfat_thick | 0% | 97% | 3% | 97% | 3% | 10047 | 50 | 10049 | 50 | | | | | | | | | | | | |
| oatcakes_spread_dunno_lowfat_thin | 0% | 97% | 3% | 97% | 3% | 10047 | 50 | 10049 | 50 | | | | | | | | | | | | |
| oatcakes_spread_dunno_vlowfat_med | 0% | 97% | 3% | 97% | 3% | 10047 | 50 | 10049 | 50 | | | | | | | | | | | | |
| oatcakes_spread_dunno_vlowfat_thick | 0% | 97% | 3% | 97% | 3% | 10047 | 50 | 10049 | 50 | | | | | | | | | | | | |
| oatcakes_spread_dunno_vlowfat_thin | 0% | 97% | 3% | 97% | 3% | 10047 | 50 | 10049 | 50 | | | | | | | | | | | | |
| oatcakes_spread_hardmarg_med | 0% | 100% | 0% | 100% | 0% | 860 | 100 | | | | | | | | | | | | | | |
| oatcakes_spread_hardmarg_thick | 0% | 100% | 0% | 100% | 0% | 860 | 100 | | | | | | | | | | | | | | |
| oatcakes_spread_hardmarg_thin | 0% | 100% | 0% | 100% | 0% | 860 | 100 | | | | | | | | | | | | | | |
| oatcakes_spread_olive_chol_med | 0% | 99% | 1% | 99% | 1% | 3364 | 50 | 10053 | 50 | | | | | | | | | | | | |
| oatcakes_spread_olive_chol_thick | 0% | 99% | 1% | 99% | 1% | 3364 | 50 | 10053 | 50 | | | | | | | | | | | | |
| oatcakes_spread_olive_chol_thin | 0% | 99% | 1% | 99% | 1% | 3364 | 50 | 10053 | 50 | | | | | | | | | | | | |
| oatcakes_spread_olive_dunno_med | 0% | 99% | 1% | 99% | 1% | 10048 | 50 | 10042 | 25 | 10131 | 25 | | | | | | | | | | |
| oatcakes_spread_olive_dunno_thick | 0% | 99% | 1% | 99% | 1% | 10048 | 50 | 10042 | 25 | 10131 | 25 | | | | | | | | | | |
| oatcakes_spread_olive_dunno_thin | 0% | 99% | 1% | 99% | 1% | 10048 | 50 | 10042 | 25 | 10131 | 25 | | | | | | | | | | |
| oatcakes_spread_olive_fat_med | 0% | 99% | 1% | 99% | 1% | 10042 | 75 | 10131 | 25 | | | | | | | | | | | | |
| oatcakes_spread_olive_fat_thick | 0% | 99% | 1% | 99% | 1% | 10042 | 75 | 10131 | 25 | | | | | | | | | | | | |
| oatcakes_spread_olive_fat_thin | 0% | 99% | 1% | 99% | 1% | 10042 | 75 | 10131 | 25 | | | | | | | | | | | | |
| oatcakes_spread_olive_lowfat_med | 0% | 100% | 1% | 100% | 0% | 10048 | 100 | | | | | | | | | | | | | | |
| oatcakes_spread_olive_lowfat_thick | 0% | 100% | 1% | 100% | 0% | 10048 | 100 | | | | | | | | | | | | | | |
| oatcakes_spread_olive_lowfat_thin | 0% | 100% | 1% | 100% | 0% | 10048 | 100 | | | | | | | | | | | | | | |
| oatcakes_spread_olive_vlowfat_med | 0% | 100% | 1% | 100% | 0% | 10048 | 100 | | | | | | | | | | | | | | |
| oatcakes_spread_olive_vlowfat_thick | 0% | 100% | 1% | 100% | 0% | 10048 | 100 | | | | | | | | | | | | | | |
| oatcakes_spread_olive_vlowfat_thin | 0% | 100% | 1% | 100% | 0% | 10048 | 100 | | | | | | | | | | | | | | |
| oatcakes_spread_other_med | 0% | 60% | 40% | 60% | 40% | 856 | 18 | 851 | 7.5 | 852 | 7.5 | 3848 | 33.5 | 2849 | 33.5 | | | | | | |
| oatcakes_spread_other_thick | 0% | 60% | 40% | 60% | 40% | 856 | 18 | 851 | 7.5 | 852 | 7.5 | 3848 | 33.5 | 2849 | 33.5 | | | | | | |
| oatcakes_spread_other_thin | 0% | 60% | 40% | 60% | 40% | 856 | 18 | 851 | 7.5 | 852 | 7.5 | 3848 | 33.5 | 2849 | 33.5 | | | | | | |
| oatcakes_spread_polymarg_chol_med | 0% | 100% | 0% | 100% | 0% | 2849 | 33.4 | 3848 | 33.3 | 3243 | 33.3 | | | | | | | | | | |
| oatcakes_spread_polymarg_chol_thick | 0% | 100% | 0% | 100% | 0% | 2849 | 33.4 | 3848 | 33.3 | 3243 | 33.3 | | | | | | | | | | |
| oatcakes_spread_polymarg_chol_thin | 0% | 100% | 0% | 100% | 0% | 2849 | 33.4 | 3848 | 33.3 | 3243 | 33.3 | | | | | | | | | | |
| oatcakes_spread_polymarg_dunno_med | 0% | 100% | 0% | 100% | 0% | 10049 | 50 | 10043 | 25 | 10044 | 25 | | | | | | | | | | |
| oatcakes_spread_polymarg_dunno_thick | 0% | 100% | 0% | 100% | 0% | 10049 | 50 | 10043 | 25 | 10044 | 25 | | | | | | | | | | |
| oatcakes_spread_polymarg_dunno_thin | 0% | 100% | 0% | 100% | 0% | 10049 | 50 | 10043 | 25 | 10044 | 25 | | | | | | | | | | |
| oatcakes_spread_polymarg_fat_med | 0% | 100% | 0% | 100% | 0% | 10043 | 50 | 10044 | 50 | | | | | | | | | | | | |
| oatcakes_spread_polymarg_fat_thick | 0% | 100% | 0% | 100% | 0% | 10043 | 50 | 10044 | 50 | | | | | | | | | | | | |
| oatcakes_spread_polymarg_fat_thin | 0% | 100% | 0% | 100% | 0% | 10043 | 50 | 10044 | 50 | | | | | | | | | | | | |
| oatcakes_spread_polymarg_lowfat_med | 0% | 100% | 0% | 100% | 0% | 10049 | 100 | | | | | | | | | | | | | | |
| oatcakes_spread_polymarg_lowfat_thick | 0% | 100% | 0% | 100% | 0% | 10049 | 100 | | | | | | | | | | | | | | |
| oatcakes_spread_polymarg_lowfat_thin | 0% | 100% | 0% | 100% | 0% | 10049 | 100 | | | | | | | | | | | | | | |
| oatcakes_spread_polymarg_vlowfat_med | 0% | 100% | 0% | 100% | 0% | 10049 | 100 | | | | | | | | | | | | | | |
| oatcakes_spread_polymarg_vlowfat_thick | 0% | 100% | 0% | 100% | 0% | 10049 | 100 | | | | | | | | | | | | | | |
| oatcakes_spread_polymarg_vlowfat_thin | 0% | 100% | 0% | 100% | 0% | 10049 | 100 | | | | | | | | | | | | | | |
| oatcakes_spread_soya_chol_med | 0% | 100% | 1% | 100% | 1% | 3848 | 16.6 | 3243 | 16.6 | 2849 | 16.8 | 10786 | 50 | | | | | | | | |
| oatcakes_spread_soya_chol_thick | 0% | 100% | 1% | 100% | 1% | 3848 | 16.6 | 3243 | 16.6 | 2849 | 16.8 | 10786 | 50 | | | | | | | | |
| oatcakes_spread_soya_chol_thin | 0% | 100% | 1% | 100% | 1% | 3848 | 16.6 | 3243 | 16.6 | 2849 | 16.8 | 10786 | 50 | | | | | | | | |
| oatcakes_spread_soya_dunno_med | 0% | 100% | 0% | 100% | 0% | 10980 | 50 | 10786 | 50 | | | | | | | | | | | | |
| oatcakes_spread_soya_dunno_thick | 0% | 100% | 0% | 100% | 0% | 10980 | 50 | 10786 | 50 | | | | | | | | | | | | |
| oatcakes_spread_soya_dunno_thin | 0% | 100% | 0% | 100% | 0% | 10980 | 50 | 10786 | 50 | | | | | | | | | | | | |
| oatcakes_spread_soya_fat_med | 0% | 100% | 0% | 100% | 0% | 10786 | 100 | | | | | | | | | | | | | | |
| oatcakes_spread_soya_fat_thick | 0% | 100% | 0% | 100% | 0% | 10786 | 100 | | | | | | | | | | | | | | |
| oatcakes_spread_soya_fat_thin | 0% | 100% | 0% | 100% | 0% | 10786 | 100 | | | | | | | | | | | | | | |
| oatcakes_spread_soya_lowfat_med | 0% | 100% | 0% | 100% | 0% | 10980 | 100 | | | | | | | | | | | | | | |
| oatcakes_spread_soya_lowfat_thick | 0% | 100% | 0% | 100% | 0% | 10980 | 100 | | | | | | | | | | | | | | |
| oatcakes_spread_soya_lowfat_thin | 0% | 100% | 0% | 100% | 0% | 10980 | 100 | | | | | | | | | | | | | | |
| oatcakes_spread_soya_vlowfat_med | 0% | 100% | 0% | 100% | 0% | 10980 | 100 | | | | | | | | | | | | | | |
| oatcakes_spread_soya_vlowfat_thick | 0% | 100% | 0% | 100% | 0% | 10980 | 100 | | | | | | | | | | | | | | |
| oatcakes_spread_soya_vlowfat_thin | 0% | 100% | 0% | 100% | 0% | 10980 | 100 | | | | | | | | | | | | | | |
| pancake_blini_chol | 0% | 30% | 70% | 10% | 90% | 15 | 26 | 755 | 12 | 871 | 6 | 7183 | 56 | | | | | | | | |
| pancake_blini_dontknow | 0% | 30% | 70% | 10% | 90% | 15 | 26 | 755 | 12 | 871 | 6 | 608 | 14 | 8543 | 14 | 613 | 5.6 | 8544 | 5.6 | 602 | 5.6 | 603 | 5.6 | 10251 | 5.6 |
| pancake_blini_goatsheep | 0% | 30% | 70% | 10% | 90% | 15 | 26 | 755 | 12 | 871 | 6 | 10397 | 28 | 625 | 14 | 626 | 14 | | | | |
| pancake_blini_other | 0% | 30% | 70% | 10% | 90% | 15 | 26 | 755 | 12 | 871 | 6 | 10251 | 28 | 9493 | 14 | 10932 | 14 | | | | |
| pancake_blini_powdered | 0% | 30% | 70% | 10% | 90% | 15 | 26 | 755 | 12 | 871 | 6 | 695 | 28 | 696 | 28 | | | | | | |
| pancake_blini_riceoatveg | 74% | 90% | 10% | 90% | 10% | 15 | 26 | 755 | 12 | 871 | 6 | 10572 | 14 | 10898 | 14 | 10159 | 14 | 10836 | 14 | | |
| pancake_blini_semi | 0% | 30% | 70% | 10% | 90% | 15 | 26 | 755 | 12 | 871 | 6 | 608 | 28 | 8543 | 28 | | | | | | |
| pancake_blini_skimmed | 0% | 30% | 70% | 10% | 90% | 15 | 26 | 755 | 12 | 871 | 6 | 613 | 28 | 8544 | 28 | | | | | | |
| pancake_blini_soya_ca | 74% | 90% | 10% | 90% | 10% | 15 | 26 | 755 | 12 | 871 | 6 | 8726 | 18.7 | 10974 | 18.7 | 3769 | 18.6 | | | | |
| pancake_blini_soya_noca | 74% | 90% | 10% | 90% | 10% | 15 | 26 | 755 | 12 | 871 | 6 | 8512 | 28 | 650 | 28 | | | | | | |
| pancake_blini_whole | 0% | 30% | 70% | 90% | 10% | 15 | 26 | 755 | 12 | 871 | 6 | 602 | 28 | 603 | 28 | | | | | | |
| pancake_crepe_chol | 0% | 30% | 70% | 10% | 90% | 15 | 26 | 755 | 12 | 871 | 6 | 7183 | 56 | | | | | | | | |
| pancake_crepe_dontknow | 0% | 30% | 70% | 10% | 90% | 15 | 26 | 755 | 12 | 871 | 6 | 608 | 14 | 8543 | 14 | 613 | 5.6 | 8544 | 5.6 | 602 | 5.6 | 603 | 5.6 | 10251 | 5.6 |
| pancake_crepe_goatsheep | 0% | 30% | 70% | 10% | 90% | 15 | 26 | 755 | 12 | 871 | 6 | 10397 | 28 | 625 | 14 | 626 | 14 | | | | |
| pancake_crepe_other | 0% | 30% | 70% | 10% | 90% | 15 | 26 | 755 | 12 | 871 | 6 | 10251 | 28 | 9493 | 14 | 10932 | 14 | | | | |
| pancake_crepe_powdered | 0% | 30% | 70% | 10% | 90% | 15 | 26 | 755 | 12 | 871 | 6 | 695 | 28 | 696 | 28 | | | | | | |
| pancake_crepe_riceoatveg | 74% | 90% | 10% | 90% | 10% | 15 | 26 | 755 | 12 | 871 | 6 | 10572 | 14 | 10898 | 14 | 10159 | 14 | 10836 | 14 | | |
| pancake_crepe_semi | 0% | 30% | 70% | 10% | 90% | 15 | 26 | 755 | 12 | 871 | 6 | 608 | 28 | 8543 | 28 | | | | | | |
| pancake_crepe_skimmed | 0% | 30% | 70% | 10% | 90% | 15 | 26 | 755 | 12 | 871 | 6 | 613 | 28 | 8544 | 28 | | | | | | |
| pancake_crepe_soya_ca | 74% | 90% | 10% | 90% | 10% | 15 | 26 | 755 | 12 | 871 | 6 | 8726 | 18.7 | 10974 | 18.7 | 3769 | 18.6 | | | | |
| pancake_crepe_soya_noca | 74% | 90% | 10% | 90% | 10% | 15 | 26 | 755 | 12 | 871 | 6 | 8512 | 28 | 650 | 28 | | | | | | |
| pancake_crepe_whole | 0% | 30% | 70% | 90% | 10% | 15 | 26 | 755 | 12 | 871 | 6 | 602 | 28 | 603 | 28 | | | | | | |
| pasta_brown | 0% | 100% | 0% | 100% | 0% | 36 | 100 | | | | | | | | | | | | | | |
| pasta_gf | 0% | 100% | 0% | 100% | 0% | 7100 | 50 | 8716 | 50 | | | | | | | | | | | | |
| pasta_white | 0% | 50% | 50% | 60% | 40% | 9371 | 25 | 34 | 50 | 32 | 25 | | | | | | | | | | |
| pesto | 0% | 60% | 40% | 70% | 30% | 6036 | 100 | | | | | | | | | | | | | | |
| pizza | 0% | 30% | 70% | 10% | 90% | 10026 | 100 | | | | | | | | | | | | | | |
| pizza_gf | 0% | 30% | 70% | 10% | 90% | 10026 | 100 | | | | | | | | | | | | | | |
| pnutbutter_nutella | 100% | 100% | 1% | 100% | 1% | 2195 | 70 | 2212 | 30 | | | | | | | | | | | | |
| potato_boil | 0% | 100% | 0% | 100% | 0% | 8294 | 25 | 1830 | 25 | 1829 | 25 | 11117 | 25 | | | | | | | | |
| potato_boil_marg | 0% | 50% | 50% | 50% | 50% | 10043 | 25 | 7775 | 25 | 851 | 25 | 852 | 25 | | | | | | | | |
| potato_fried | 0% | 100% | 0% | 100% | 0% | 10561 | 20 | 1841 | 20 | 11123 | 20 | 1949 | 20 | 8684 | 20 | | | | | | |
| potato_mashed_butter_dunno | 0% | 70% | 30% | 10% | 90% | 1829 | 89 | 608 | 3 | 8543 | 3 | 9407 | 1.25 | 10039 | 1.25 | 851 | 1.25 | 852 | 1.25 | | |
| potato_mashed_butter_fat | 0% | 70% | 30% | 1% | 99% | 1829 | 89 | 608 | 3 | 8543 | 3 | 851 | 2.5 | 852 | 2.5 | | | | | | |
| potato_mashed_butter_lowfat | 0% | 70% | 30% | 40% | 60% | 1829 | 89 | 608 | 3 | 8543 | 3 | 10140 | 5 | | | | | | | | |
| potato_mashed_butter_spread_fat | 0% | 70% | 30% | 30% | 70% | 1829 | 89 | 608 | 3 | 8543 | 3 | 9407 | 2.5 | 10039 | 2.5 | | | | | | |
| potato_mashed_butter_spread_lowfat | 0% | 70% | 30% | 40% | 60% | 1829 | 89 | 608 | 3 | 8543 | 3 | 10894 | 2.5 | 3891 | 2.5 | | | | | | |
| potato_mashed_fat_dunno | 0% | 70% | 30% | 90% | 10% | 1829 | 89 | 608 | 3 | 8543 | 3 | 875 | 5 | | | | | | | | |
| potato_mashed_fat_none | 0% | 70% | 30% | 10% | 90% | 1829 | 93 | 608 | 3.5 | 8543 | 3.5 | | | | | | | | | | |
| potato_mashed_fat_other | 0% | 70% | 30% | 1% | 99% | 1829 | 89 | 608 | 3 | 8543 | 3 | 856 | 2.5 | 855 | 2.5 | | | | | | |
| potato_mashed_lard | 0% | 70% | 30% | 1% | 99% | 1829 | 89 | 608 | 3 | 8543 | 3 | 858 | 5 | | | | | | | | |

| | | | | | | | | | | | | | | | | | | | | | |
|---|---|---|---|---|---|---|---|---|---|---|---|---|---|---|---|---|---|---|---|---|---|
| potato_mashed_marg_hard | 0% | 70% | 30% | 90% | 10% | 1829 | 89 | 608 | 3 | 8543 | 3 | 860 | 5 | | | | | | | | |
| potato_mashed_marg_poly_chol | 0% | 70% | 30% | 90% | 10% | 1829 | 89 | 608 | 3 | 8543 | 3 | 2849 | 1.7 | 3848 | 1.7 | 3243 | 1.6 | | | | |
| potato_mashed_marg_poly_dunno | 0% | 70% | 30% | 90% | 10% | 1829 | 89 | 608 | 3 | 8543 | 3 | 10049 | 2.5 | 10043 | 1.25 | 10044 | 1.25 | | | | |
| potato_mashed_marg_poly_fat | 0% | 70% | 30% | 90% | 10% | 1829 | 89 | 608 | 3 | 8543 | 3 | 10043 | 2.5 | 10044 | 2.5 | | | | | | |
| potato_mashed_marg_poly_lowfat | 0% | 70% | 30% | 90% | 10% | 1829 | 89 | 608 | 3 | 8543 | 3 | 10049 | 5 | | | | | | | | |
| potato_mashed_marg_poly_vlowfat | 0% | 70% | 30% | 90% | 10% | 1829 | 89 | 608 | 3 | 8543 | 3 | 10049 | 5 | | | | | | | | |
| potato_mashed_marg_soya_chol | 0% | 70% | 30% | 90% | 10% | 1829 | 89 | 608 | 3 | 8543 | 3 | 3848 | 0.83 | 3243 | 0.83 | 2849 | 0.84 | 10786 | 2.5 | | |
| potato_mashed_marg_soya_dunno | 0% | 70% | 30% | 90% | 10% | 1829 | 89 | 608 | 3 | 8543 | 3 | 10980 | 2.5 | 10786 | 2.5 | | | | | | |
| potato_mashed_marg_soya_fat | 0% | 70% | 30% | 90% | 10% | 1829 | 89 | 608 | 3 | 8543 | 3 | 10786 | 5 | | | | | | | | |
| potato_mashed_marg_soya_lowfat | 0% | 70% | 30% | 90% | 10% | 1829 | 89 | 608 | 3 | 8543 | 3 | 10980 | 5 | | | | | | | | |
| potato_mashed_marg_soya_vlowfat | 0% | 70% | 30% | 90% | 10% | 1829 | 89 | 608 | 3 | 8543 | 3 | 10980 | 5 | | | | | | | | |
| potato_mashed_oil_olive | 0% | 70% | 30% | 90% | 10% | 1829 | 89 | 608 | 3 | 8543 | 3 | 874 | 5 | | | | | | | | |
| potato_mashed_oil_other | 0% | 70% | 30% | 90% | 10% | 1829 | 89 | 608 | 3 | 8543 | 3 | 870 | 1.5 | 870 | 1 | 10963 | 1.25 | 871 | 0.625 | 5000 | 0.63 |
| potato_mashed_oil_rapeseed | 0% | 70% | 30% | 90% | 10% | 1829 | 89 | 608 | 3 | 8543 | 3 | 7990 | 5 | | | | | | | | |
| potato_mashed_oil_sunflower | 0% | 70% | 30% | 90% | 10% | 1829 | 89 | 608 | 3 | 8543 | 3 | 873 | 5 | | | | | | | | |
| potato_mashed_oil_veg | 0% | 70% | 30% | 90% | 10% | 1829 | 89 | 608 | 3 | 8543 | 3 | 872 | 2.5 | 871 | 2.5 | | | | | | |
| potato_mashed_spread_dairy_chol | 0% | 70% | 30% | 80% | 20% | 1829 | 89 | 608 | 3 | 8543 | 3 | 3848 | 5 | | | | | | | | |
| potato_mashed_spread_dairy_dunno | 0% | 70% | 30% | 80% | 20% | 1829 | 89 | 608 | 3 | 8543 | 3 | 10047 | 2.5 | 7775 | 2.5 | | | | | | |
| potato_mashed_spread_dairy_fat | 0% | 70% | 30% | 80% | 20% | 1829 | 89 | 608 | 3 | 8543 | 3 | 7775 | 5 | | | | | | | | |
| potato_mashed_spread_dairy_lowfat | 0% | 70% | 30% | 80% | 20% | 1829 | 89 | 608 | 3 | 8543 | 3 | 10047 | 5 | | | | | | | | |
| potato_mashed_spread_dairy_vlowfat | 0% | 70% | 30% | 80% | 20% | 1829 | 89 | 608 | 3 | 8543 | 3 | 10047 | 5 | | | | | | | | |
| potato_mashed_spread_dunno_chol | 0% | 70% | 30% | 80% | 20% | 1829 | 89 | 608 | 3 | 8543 | 3 | 3848 | 1.7 | 3243 | 1.7 | 2849 | 1.6 | | | | |
| potato_mashed_spread_dunno_dunno | 0% | 70% | 30% | 80% | 20% | 1829 | 89 | 608 | 3 | 8543 | 3 | 10047 | 1.25 | 10049 | 1.25 | 10043 | 1.25 | 7775 | 1.25 | | |
| potato_mashed_spread_dunno_fat | 0% | 70% | 30% | 80% | 20% | 1829 | 89 | 608 | 3 | 8543 | 3 | 10043 | 2.5 | 7775 | 2.5 | | | | | | |
| potato_mashed_spread_dunno_lowfat | 0% | 70% | 30% | 80% | 20% | 1829 | 89 | 608 | 3 | 8543 | 3 | 10047 | 2.5 | 10049 | 2.5 | | | | | | |
| potato_mashed_spread_dunno_vlowfat | 0% | 70% | 30% | 80% | 20% | 1829 | 89 | 608 | 3 | 8543 | 3 | 10047 | 2.5 | 10049 | 2.5 | | | | | | |
| potato_mashed_spread_olive_chol | 0% | 70% | 30% | 80% | 20% | 1829 | 89 | 608 | 3 | 8543 | 3 | 3364 | 2.5 | 10053 | 2.5 | | | | | | |
| potato_mashed_spread_olive_dunno | 0% | 70% | 30% | 80% | 20% | 1829 | 89 | 608 | 3 | 8543 | 3 | 10048 | 2.5 | 10042 | 1.25 | 10131 | 1.25 | | | | |
| potato_mashed_spread_olive_fat | 0% | 70% | 30% | 80% | 20% | 1829 | 89 | 608 | 3 | 8543 | 3 | 10042 | 3.75 | 10131 | 1.25 | | | | | | |
| potato_mashed_spread_olive_lowfat | 0% | 70% | 30% | 80% | 20% | 1829 | 89 | 608 | 3 | 8543 | 3 | 10048 | 5 | | | | | | | | |
| potato_mashed_spread_olive_vlowfat | 0% | 70% | 30% | 80% | 20% | 1829 | 89 | 608 | 3 | 8543 | 3 | 10048 | 5 | | | | | | | | |
| poultry_friedbcrumb_noskin | 0% | 1% | 99% | 10% | 90% | 5263 | 67 | 1087 | 33 | | | | | | | | | | | | |
| poultry_friedbcrumb_withskin | 0% | 1% | 99% | 10% | 90% | 5263 | 67 | 1087 | 33 | | | | | | | | | | | | |
| poultry_noskin | 0% | 0% | 100% | 1% | 99% | 1091 | 30 | 1090 | 30 | 5270 | 15 | 5265 | 15 | 1147 | 10 | | | | | | |
| poultry_withskin | 0% | 0% | 100% | 0% | 100% | 1089 | 60 | 5271 | 15 | 5265 | 15 | 1146 | 10 | | | | | | | | |
| rice_brown | 0% | 100% | 0% | 100% | 0% | 49 | 33.4 | 10009 | 33.3 | 10011 | 33.3 | | | | | | | | | | |
| rice_white | 0% | 100% | 0% | 100% | 0% | 58 | 33.4 | 42 | 33.3 | 55 | 33.3 | | | | | | | | | | |
| salad_dressing | 100% | 100% | 0% | 100% | 0% | 9391 | 100 | | | | | | | | | | | | | | |
| sauce_brown | 72% | 100% | 0% | 100% | 0% | 2412 | 50 | 2409 | 50 | | | | | | | | | | | | |
| sauce_cheese_chol | 0% | 10% | 90% | 10% | 90% | 8947 | 12.5 | 7183 | 87.5 | | | | | | | | | | | | |
| sauce_cheese_dontknow | 0% | 10% | 90% | 10% | 90% | 8947 | 12.5 | 608 | 21.9 | 8543 | 21.875 | 613 | 8.75 | 8544 | 8.75 | 602 | 8.75 | 603 | 8.75 | 10251 | 8.75 |
| sauce_cheese_goatsheep | 0% | 10% | 90% | 10% | 90% | 8947 | 12.5 | 10397 | 43.8 | 625 | 21.875 | 626 | 21.88 | | | | | | | | |
| sauce_cheese_other | 0% | 10% | 90% | 10% | 90% | 8947 | 12.5 | 10251 | 43.8 | 9493 | 21.875 | 10932 | 21.88 | | | | | | | | |
| sauce_cheese_powdered | 0% | 10% | 90% | 10% | 90% | 8947 | 12.5 | 695 | 43.8 | 696 | 43.75 | | | | | | | | | | |
| sauce_cheese_riceoatveg | 88% | 30% | 70% | 30% | 70% | 8947 | 12.5 | 10572 | 21.9 | 10898 | 21.875 | 10159 | 21.88 | 10836 | 21.9 | | | | | | |
| sauce_cheese_semi | 0% | 10% | 90% | 10% | 90% | 8947 | 12.5 | 608 | 43.8 | 8543 | 43.75 | | | | | | | | | | |
| sauce_cheese_skimmed | 0% | 10% | 90% | 10% | 90% | 8947 | 12.5 | 613 | 43.8 | 8544 | 43.75 | | | | | | | | | | |
| sauce_cheese_soya_ca | 88% | 50% | 50% | 50% | 50% | 8947 | 12.5 | 8726 | 29.2 | 10974 | 29.17 | 3769 | 29.16 | | | | | | | | |
| sauce_cheese_soya_noca | 88% | 50% | 50% | 50% | 50% | 8947 | 12.5 | 8512 | 43.8 | 650 | 43.75 | | | | | | | | | | |
| sauce_cheese_whole | 0% | 10% | 90% | 10% | 90% | 8947 | 12.5 | 602 | 43.8 | 603 | 43.75 | | | | | | | | | | |
| sauce_gravy | 0% | 10% | 90% | 10% | 90% | 2426 | 50 | 2427 | 50 | | | | | | | | | | | | |
| sauce_ketchup | 84% | 100% | 0% | 100% | 0% | 2448 | 100 | | | | | | | | | | | | | | |
| sauce_tomato | 90% | 70% | 30% | 30% | 70% | 2450 | 50 | 3997 | 25 | 10311 | 25 | | | | | | | | | | |
| sauce_white_chol | 0% | 10% | 90% | 10% | 90% | 9377 | 12.5 | 7183 | 87.5 | | | | | | | | | | | | |
| sauce_white_dontknow | 0% | 10% | 90% | 10% | 90% | 9377 | 12.5 | 608 | 21.9 | 8543 | 21.875 | 613 | 8.75 | 8544 | 8.75 | 602 | 8.75 | 603 | 8.75 | 10251 | 8.75 |
| sauce_white_goatsheep | 0% | 10% | 90% | 10% | 90% | 9377 | 12.5 | 10397 | 43.8 | 625 | 21.875 | 626 | 21.88 | | | | | | | | |
| sauce_white_other | 0% | 10% | 90% | 10% | 90% | 9377 | 12.5 | 10251 | 43.8 | 9493 | 21.875 | 10932 | 21.88 | | | | | | | | |
| sauce_white_powdered | 0% | 10% | 90% | 10% | 90% | 9377 | 12.5 | 695 | 43.8 | 696 | 43.75 | | | | | | | | | | |
| sauce_white_riceoatveg | 96% | 30% | 70% | 30% | 70% | 9377 | 12.5 | 10572 | 21.9 | 10898 | 21.875 | 10159 | 21.88 | 10836 | 21.9 | | | | | | |
| sauce_white_semi | 0% | 10% | 90% | 10% | 90% | 9377 | 12.5 | 608 | 43.8 | 8543 | 43.75 | | | | | | | | | | |
| sauce_white_skimmed | 0% | 10% | 90% | 10% | 90% | 9377 | 12.5 | 613 | 43.8 | 8544 | 43.75 | | | | | | | | | | |
| sauce_white_soya_ca | 96% | 50% | 50% | 50% | 50% | 9377 | 12.5 | 8726 | 29.2 | 10974 | 29.17 | 3769 | 29.16 | | | | | | | | |
| sauce_white_soya_noca | 96% | 50% | 50% | 50% | 50% | 9377 | 12.5 | 8512 | 43.8 | 650 | 43.75 | | | | | | | | | | |
| sauce_white_whole | 0% | 10% | 90% | 10% | 90% | 9377 | 12.5 | 602 | 43.8 | 603 | 43.75 | | | | | | | | | | |
| scone | 60% | 30% | 70% | 10% | 90% | 369 | 40 | 368 | 40 | 367 | 20 | | | | | | | | | | |
| scone_gf | 76% | 30% | 70% | 11% | 89% | 8870 | 100 | | | | | | | | | | | | | | |
| single_crust | 0% | 60% | 40% | 5% | 95% | 393 | 40 | 400 | 50 | 405 | 10 | | | | | | | | | | |
| smoothie_dairy | 47% | 3% | 97% | 5% | 95% | 10038 | 100 | | | | | | | | | | | | | | |
| smoothie_fruit | 78% | 100% | 0% | 100% | 0% | 8023 | 20 | 8024 | 20 | 10534 | 20 | 10746 | 20 | 10747 | 20 | | | | | | |
| snack_cheesybis | 0% | 50% | 50% | 50% | 50% | 251 | 100 | | | | | | | | | | | | | | |
| snack_crisps | 0% | 100% | 0% | 100% | 0% | 10004 | 16.7 | 7872 | 25 | 10070 | 25 | 10001 | 16.7 | 10000 | 16.6 | | | | | | |
| snack_olives | 0% | 100% | 0% | 100% | 0% | 2090 | 100 | | | | | | | | | | | | | | |
| snack_saltednuts | 0% | 100% | 0% | 100% | 0% | 7304 | 25 | 8548 | 25 | 7884 | 50 | | | | | | | | | | |
| snack_saltedpeanuts | 0% | 100% | 0% | 100% | 0% | 2193 | 100 | | | | | | | | | | | | | | |
| snack_savourybis | 0% | 100% | 0% | 100% | 0% | 3236 | 34 | 3973 | 33 | 8155 | 33 | | | | | | | | | | |
| snack_seeds | 0% | 100% | 0% | 100% | 0% | 2168 | 25 | 2167 | 25 | 2166 | 25 | 9741 | 25 | | | | | | | | |
| snack_svyother | 0% | 100% | 0% | 100% | 0% | 2605 | 10 | 10563 | 30 | 10000 | 30 | 6825 | 20 | 4408 | 10 | | | | | | |
| snack_swtother | 83% | 30% | 70% | 50% | 50% | 8136 | 10 | 8193 | 30 | 7662 | 30 | 2254 | 20 | 2269 | 10 | | | | | | |
| snack_unsaltednuts | 0% | 100% | 0% | 100% | 0% | 2177 | 25 | 2199 | 25 | 2169 | 25 | 2197 | 25 | | | | | | | | |
| snack_unsaltedpeanuts | 0% | 100% | 0% | 100% | 0% | 2191 | 100 | | | | | | | | | | | | | | |
| snackpot | 71% | 60% | 40% | 60% | 40% | 10392 | 100 | | | | | | | | | | | | | | |
| soup_canned_fish | 0% | 30% | 70% | 20% | 80% | 2951 | 100 | | | | | | | | | | | | | | |
| soup_canned_meat | 67% | 30% | 70% | 20% | 80% | 2463 | 33.4 | 7925 | 33.3 | 2473 | 33.3 | | | | | | | | | | |
| soup_canned_other | 50% | 70% | 30% | 60% | 40% | 2483 | 50 | 10960 | 40 | 7926 | 10 | | | | | | | | | | |
| soup_canned_pasta | 0% | 70% | 30% | 20% | 80% | 8890 | 50 | 2467 | 50 | | | | | | | | | | | | |
| soup_canned_pulse | 60% | 80% | 20% | 60% | 40% | 3772 | 50 | 10960 | 50 | | | | | | | | | | | | |
| soup_canned_unanswered | 60% | 80% | 20% | 70% | 30% | 3772 | 33.3 | 2463 | 33.3 | 2483 | 33.4 | | | | | | | | | | |
| soup_canned_veg | 50% | 100% | 0% | 100% | 0% | 7926 | 12.5 | 2478 | 25 | 2483 | 25 | 10731 | 12.5 | 2472 | 12.5 | 9971 | 12.5 | | | | |
| soup_homemade_fish | 0% | 30% | 70% | 20% | 80% | 2951 | 100 | | | | | | | | | | | | | | |
| soup_homemade_meat | 0% | 30% | 70% | 20% | 80% | 5668 | 50 | 6790 | 50 | | | | | | | | | | | | |
| soup_homemade_other | 0% | 70% | 30% | 60% | 40% | 8659 | 10 | 9075 | 40 | 9166 | 50 | | | | | | | | | | |
| soup_homemade_pasta | 0% | 70% | 30% | 20% | 80% | 8890 | 100 | | | | | | | | | | | | | | |
| soup_homemade_pulse | 0% | 80% | 20% | 60% | 40% | 2469 | 50 | 2476 | 50 | | | | | | | | | | | | |
| soup_homemade_unanswered | 0% | 80% | 20% | 70% | 30% | 2469 | 33.3 | 5668 | 33.3 | 9166 | 33.4 | | | | | | | | | | |
| soup_homemade_veg | 0% | 100% | 0% | 100% | 0% | 8659 | 20 | 6880 | 20 | 9166 | 20 | 5997 | 20 | 5996 | 20 | | | | | | |
| soup_powder | 97% | 60% | 40% | 50% | 50% | 2468 | 100 | | | | | | | | | | | | | | |
| spongepuds | 81% | 20% | 80% | 20% | 80% | 567 | 34 | 11010 | 33 | 5749 | 33 | | | | | | | | | | |
| spongepuds_gf | 81% | 21% | 79% | 20% | 80% | 567 | 34 | 11010 | 33 | 5749 | 33 | | | | | | | | | | |
| spreadsauce_other | 100% | 50% | 50% | 50% | 50% | 2454 | 12.5 | 8074 | 12.5 | 9392 | 12.5 | 2159 | 12.5 | 3980 | 12.5 | 3981 | 12.5 | 2435 | 12.5 | 2428 | 12.5 |
| sushi | 16% | 20% | 80% | 20% | 80% | 9153 | 33.4 | 2820 | 33.3 | 2821 | 33.3 | | | | | | | | | | |
| sweets | 100% | 20% | 80% | 20% | 80% | 2251 | 25 | 2272 | 25 | 11013 | 12.5 | 8303 | 25 | 11014 | 12.5 | | | | | | |
| sweets_diet | 0% | 20% | 80% | 20% | 80% | 8646 | 34 | 7968 | 33 | 10707 | 33 | | | | | | | | | | |
| tea_black | 0% | 100% | 0% | 100% | 0% | 2314 | 50 | 2315 | 50 | | | | | | | | | | | | |
| tea_black_decaf | 0% | 100% | 0% | 100% | 0% | 8316 | 50 | 8317 | 50 | | | | | | | | | | | | |
| tea_green | 0% | 100% | 0% | 100% | 0% | 8006 | 100 | | | | | | | | | | | | | | |
| tea_herbal | 0% | 100% | 0% | 100% | 0% | 5341 | 50 | 5340 | 50 | | | | | | | | | | | | |
| tea_other | 0% | 100% | 0% | 100% | 0% | 2316 | 1.3 | 5104 | 48.7 | 5341 | 20 | 2314 | 15 | 2315 | 15 | | | | | | |
| tea_rooibos | 0% | 100% | 0% | 100% | 0% | 5341 | 100 | | | | | | | | | | | | | | |
| tea_sugar | 100% | 100% | 0% | 100% | 0% | 2205 | 100 | | | | | | | | | | | | | | |
| veg_avocado | 0% | 100% | 0% | 100% | 0% | 1976 | 100 | | | | | | | | | | | | | | |
| veg_bakedbeans | 90% | 100% | 0% | 100% | 0% | 1662 | 75 | 2646 | 25 | | | | | | | | | | | | |
| veg_beetroot | 0% | 100% | 0% | 100% | 0% | 2456 | 50 | 1691 | 50 | | | | | | | | | | | | |
| veg_broadbeans | 0% | 100% | 0% | 100% | 0% | 1666 | 50 | 1668 | 50 | | | | | | | | | | | | |
| veg_broccoli | 0% | 100% | 0% | 100% | 0% | 11109 | 75 | 1694 | 25 | | | | | | | | | | | | |
| veg_butternut | 0% | 100% | 0% | 100% | 0% | 3954 | 100 | | | | | | | | | | | | | | |
| veg_cabbagekale | 0% | 100% | 0% | 100% | 0% | 1705 | 33.4 | 7845 | 33.3 | 1708 | 33.3 | | | | | | | | | | |
| veg_carrots | 0% | 100% | 0% | 100% | 0% | 1711 | 40 | 1713 | 30 | 1712 | 10 | 1714 | 20 | | | | | | | | |
| veg_cauli | 0% | 100% | 0% | 100% | 0% | 1719 | 100 | | | | | | | | | | | | | | |
| veg_celery | 0% | 100% | 0% | 100% | 0% | 1725 | 100 | | | | | | | | | | | | | | |
| veg_courgette_butter_dunno | 0% | 99% | 1% | 10% | 90% | 1734 | 97.5 | 9407 | 0.63 | 10039 | 0.625 | 851 | 0.625 | 852 | 0.63 | | | | | | |
| veg_courgette_butter_fat | 0% | 99% | 1% | 1% | 99% | 1734 | 97.5 | 851 | 1.25 | 852 | 1.25 | | | | | | | | | | |
| veg_courgette_butter_lowfat | 0% | 99% | 1% | 40% | 60% | 1734 | 97.5 | 10140 | 2.5 | | | | | | | | | | | | |
| veg_courgette_butter_spread_fat | 0% | 99% | 1% | 30% | 70% | 1734 | 97.5 | 9407 | 1.25 | 10039 | 1.25 | | | | | | | | | | |
| veg_courgette_butter_spread_lowfat | 0% | 99% | 1% | 40% | 60% | 1734 | 97.5 | 10894 | 1.25 | 3891 | 1.25 | | | | | | | | | | |
| veg_courgette_fat_dunno | 0% | 99% | 1% | 100% | 0% | 1734 | 97.5 | 875 | 2.5 | | | | | | | | | | | | |
| veg_courgette_fat_none | 0% | 100% | 0% | 100% | 0% | 1734 | 100 | | | | | | | | | | | | | | |
| veg_courgette_fat_other | 0% | 99% | 1% | 1% | 99% | 1734 | 97.5 | 856 | 1.25 | 855 | 1.25 | | | | | | | | | | |
| veg_courgette_lard | 0% | 99% | 1% | 1% | 99% | 1734 | 97.5 | 858 | 2.5 | | | | | | | | | | | | |
| veg_courgette_marg_hard | 0% | 99% | 1% | 99% | 1% | 1734 | 97.5 | 860 | 2.5 | | | | | | | | | | | | |
| veg_courgette_marg_poly_chol | 0% | 99% | 1% | 90% | 10% | 1734 | 97.5 | 2849 | 0.84 | 3848 | 0.83 | 3243 | 0.83 | | | | | | | | |
| veg_courgette_marg_poly_dunno | 0% | 99% | 1% | 90% | 10% | 1734 | 97.5 | 10049 | 1.25 | 10043 | 0.625 | 10044 | 0.625 | | | | | | | | |
| veg_courgette_marg_poly_fat | 0% | 99% | 1% | 90% | 10% | 1734 | 97.5 | 10043 | 1.25 | 10044 | 1.25 | | | | | | | | | | |
| veg_courgette_marg_poly_lowfat | 0% | 99% | 1% | 90% | 10% | 1734 | 97.5 | 10049 | 2.5 | | | | | | | | | | | | |
| veg_courgette_marg_poly_vlowfat | 0% | 99% | 1% | 90% | 10% | 1734 | 97.5 | 10049 | 2.5 | | | | | | | | | | | | |
| veg_courgette_marg_soya_chol | 0% | 99% | 1% | 99% | 1% | 1734 | 97.5 | 3848 | 0.42 | 3243 | 0.41 | 2849 | 0.42 | 10786 | 1.25 | | | | | | |
| veg_courgette_marg_soya_dunno | 0% | 99% | 1% | 100% | 0% | 1734 | 97.5 | 10980 | 1.25 | 10786 | 1.25 | | | | | | | | | | |
| veg_courgette_marg_soya_fat | 0% | 99% | 1% | 100% | 0% | 1734 | 97.5 | 10786 | 2.5 | | | | | | | | | | | | |
| veg_courgette_marg_soya_lowfat | 0% | 99% | 1% | 100% | 0% | 1734 | 97.5 | 10980 | 2.5 | | | | | | | | | | | | |
| veg_courgette_marg_soya_vlowfat | 0% | 99% | 1% | 100% | 0% | 1734 | 97.5 | 10980 | 2.5 | | | | | | | | | | | | |
| veg_courgette_oil_olive | 0% | 100% | 0% | 100% | 0% | 1734 | 97.5 | 874 | 2.5 | | | | | | | | | | | | |
| veg_courgette_oil_other | 0% | 100% | 0% | 100% | 0% | 1734 | 97.5 | 870 | 0.75 | 870 | 0.5 | 10963 | 0.625 | 871 | 0.31 | 5000 | 0.31 | | | | |
| veg_courgette_oil_rapeseed | 0% | 100% | 0% | 100% | 0% | 1734 | 97.5 | 7990 | 2.5 | | | | | | | | | | | | |
| veg_courgette_oil_sunflower | 0% | 100% | 0% | 100% | 0% | 1734 | 97.5 | 873 | 2.5 | | | | | | | | | | | | |
| veg_courgette_oil_veg | 0% | 100% | 0% | 100% | 0% | 1734 | 97.5 | 872 | 1.25 | 871 | 1.25 | | | | | | | | | | |
| veg_courgette_spread_dairy_chol | 0% | 99% | 1% | 99% | 1% | 1734 | 97.5 | 3848 | 2.5 | | | | | | | | | | | | |
| veg_courgette_spread_dairy_dunno | 0% | 99% | 1% | 99% | 1% | 1734 | 97.5 | 10047 | 1.25 | 7775 | 1.25 | | | | | | | | | | |
| veg_courgette_spread_dairy_fat | 0% | 99% | 1% | 99% | 1% | 1734 | 97.5 | 7775 | 2.5 | | | | | | | | | | | | |
| veg_courgette_spread_dairy_lowfat | 0% | 99% | 1% | 99% | 1% | 1734 | 97.5 | 10047 | 2.5 | | | | | | | | | | | | |
| veg_courgette_spread_dairy_vlowfat | 0% | 99% | 1% | 99% | 1% | 1734 | 97.5 | 10047 | 2.5 | | | | | | | | | | | | |
| veg_courgette_spread_dunno_chol | 0% | 99% | 1% | 99% | 1% | 1734 | 97.5 | 3848 | 0.84 | 3243 | 0.83 | 2849 | 0.83 | | | | | | | | |
| veg_courgette_spread_dunno_dunno | 0% | 99% | 1% | 99% | 1% | 1734 | 97.5 | 10047 | 0.63 | 10049 | 0.625 | 10043 | 0.625 | 7775 | 0.63 | | | | | | |
| veg_courgette_spread_dunno_fat | 0% | 99% | 1% | 99% | 1% | 1734 | 97.5 | 10043 | 1.25 | 7775 | 1.25 | | | | | | | | | | |
| veg_courgette_spread_dunno_lowfat | 0% | 99% | 1% | 99% | 1% | 1734 | 97.5 | 10047 | 1.25 | 10049 | 1.25 | | | | | | | | | | |
| veg_courgette_spread_dunno_vlowfat | 0% | 99% | 1% | 99% | 1% | 1734 | 97.5 | 10047 | 1.25 | 10049 | 1.25 | | | | | | | | | | |
| veg_courgette_spread_olive_chol | 0% | 99% | 1% | 99% | 1% | 1734 | 97.5 | 3364 | 1.25 | 10053 | 1.25 | | | | | | | | | | |
| veg_courgette_spread_olive_dunno | 0% | 99% | 1% | 99% | 1% | 1734 | 97.5 | 10048 | 1.25 | 10042 | 0.625 | 10131 | 0.625 | | | | | | | | |
| veg_courgette_spread_olive_fat | 0% | 99% | 1% | 99% | 1% | 1734 | 97.5 | 10042 | 1.88 | 10131 | 0.625 | | | | | | | | | | |
| veg_courgette_spread_olive_lowfat | 0% | 99% | 1% | 99% | 1% | 1734 | 97.5 | 10048 | 2.5 | | | | | | | | | | | | |
| veg_courgette_spread_olive_vlowfat | 0% | 99% | 1% | 99% | 1% | 1734 | 97.5 | 10048 | 2.5 | | | | | | | | | | | | |
| veg_cucumber | 0% | 100% | 0% | 100% | 0% | 1740 | 100 | | | | | | | | | | | | | | |
| veg_garlic | 0% | 100% | 0% | 100% | 0% | 1743 | 100 | | | | | | | | | | | | | | |
| veg_greenbeans | 0% | 100% | 0% | 100% | 0% | 1671 | 75 | 1681 | 12.5 | 1683 | 12.5 | | | | | | | | | | |
| veg_leek_butter_dunno | 0% | 99% | 1% | 10% | 90% | 1756 | 98.75 | 9407 | 0.31 | 10039 | 0.3125 | 851 | 0.313 | 852 | 0.31 | | | | | | |
| veg_leek_butter_fat | 0% | 99% | 1% | 1% | 99% | 1756 | 98.75 | 851 | 0.63 | 852 | 0.625 | | | | | | | | | | |
| veg_leek_butter_lowfat | 0% | 99% | 1% | 40% | 60% | 1756 | 98.75 | 10140 | 1.25 | | | | | | | | | | | | |
| veg_leek_butter_spread_fat | 0% | 99% | 1% | 30% | 70% | 1756 | 98.75 | 9407 | 0.63 | 10039 | 0.625 | | | | | | | | | | |
| veg_leek_butter_spread_lowfat | 0% | 99% | 1% | 40% | 60% | 1756 | 98.75 | 10894 | 0.63 | 3891 | 0.625 | | | | | | | | | | |
| veg_leek_fat_dunno | 0% | 99% | 1% | 100% | 0% | 1756 | 98.75 | 875 | 1.25 | | | | | | | | | | | | |
| veg_leek_fat_none | 0% | 100% | 0% | 100% | 0% | 1756 | 100 | | | | | | | | | | | | | | |

| | | | | | | | | | | | | | | | | | | | | | | | | | |
|---|---|---|---|---|---|---|---|---|---|---|---|---|---|---|---|---|---|---|---|---|---|---|---|---|---|
| veg_leek_fat_other | 0% | 99% | 1% | 1% | 99% | 1756 | 98.75 | 856 | 0.63 | 855 | 0.625 | | | | | | | | | | | | | | |
| veg_leek_lard | 0% | 99% | 1% | 1% | 99% | 1756 | 98.75 | 858 | 1.25 | | | | | | | | | | | | | | | | |
| veg_leek_marg_hard | 0% | 99% | 1% | 99% | 1% | 1756 | 98.75 | 860 | 1.25 | | | | | | | | | | | | | | | | |
| veg_leek_marg_poly_chol | 0% | 99% | 1% | 90% | 10% | 1756 | 98.75 | 2849 | 0.42 | 3848 | 0.42 | 3243 | 0.41 | | | | | | | | | | | | |
| veg_leek_marg_poly_dunno | 0% | 99% | 1% | 90% | 10% | 1756 | 98.75 | 10049 | 0.63 | 10043 | 0.3125 | 10044 | 0.313 | | | | | | | | | | | | |
| veg_leek_marg_poly_fat | 0% | 99% | 1% | 90% | 10% | 1756 | 98.75 | 10043 | 0.63 | 10044 | 0.625 | | | | | | | | | | | | | | |
| veg_leek_marg_poly_lowfat | 0% | 99% | 1% | 90% | 10% | 1756 | 98.75 | 10049 | 1.25 | | | | | | | | | | | | | | | | |
| veg_leek_marg_poly_vlowfat | 0% | 99% | 1% | 90% | 10% | 1756 | 98.75 | 10049 | 1.25 | | | | | | | | | | | | | | | | |
| veg_leek_marg_soya_chol | 0% | 99% | 1% | 99% | 1% | 1756 | 98.75 | 3848 | 0.21 | 3243 | 0.21 | 2849 | 0.205 | 10786 | 0.63 | | | | | | | | | | |
| veg_leek_marg_soya_dunno | 0% | 99% | 1% | 100% | 0% | 1756 | 98.75 | 10980 | 0.63 | 10786 | 0.625 | | | | | | | | | | | | | | |
| veg_leek_marg_soya_fat | 0% | 99% | 1% | 100% | 0% | 1756 | 98.75 | 10786 | 1.25 | | | | | | | | | | | | | | | | |
| veg_leek_marg_soya_lowfat | 0% | 99% | 1% | 100% | 0% | 1756 | 98.75 | 10980 | 1.25 | | | | | | | | | | | | | | | | |
| veg_leek_marg_soya_vlowfat | 0% | 99% | 1% | 100% | 0% | 1756 | 98.75 | 10980 | 1.25 | | | | | | | | | | | | | | | | |
| veg_leek_oil_olive | 0% | 100% | 0% | 100% | 0% | 1756 | 98.75 | 874 | 1.25 | | | | | | | | | | | | | | | | |
| veg_leek_oil_other | 0% | 100% | 0% | 100% | 0% | 1756 | 98.75 | 870 | 0.38 | 870 | 0.25 | 10963 | 0.31 | 871 | 0.16 | 5000 | 0.15 | | | | | | | | |
| veg_leek_oil_rapeseed | 0% | 100% | 0% | 100% | 0% | 1756 | 98.75 | 7990 | 1.25 | | | | | | | | | | | | | | | | |
| veg_leek_oil_sunflower | 0% | 100% | 0% | 100% | 0% | 1756 | 98.75 | 873 | 1.25 | | | | | | | | | | | | | | | | |
| veg_leek_oil_veg | 0% | 100% | 0% | 100% | 0% | 1756 | 98.75 | 872 | 0.63 | 871 | 0.625 | | | | | | | | | | | | | | |
| veg_leek_spread_dairy_chol | 0% | 99% | 1% | 99% | 1% | 1756 | 98.75 | 3848 | 1.25 | | | | | | | | | | | | | | | | |
| veg_leek_spread_dairy_dunno | 0% | 99% | 1% | 99% | 1% | 1756 | 98.75 | 10047 | 0.63 | 7775 | 0.625 | | | | | | | | | | | | | | |
| veg_leek_spread_dairy_fat | 0% | 99% | 1% | 99% | 1% | 1756 | 98.75 | 7775 | 1.25 | | | | | | | | | | | | | | | | |
| veg_leek_spread_dairy_lowfat | 0% | 99% | 1% | 99% | 1% | 1756 | 98.75 | 10047 | 1.25 | | | | | | | | | | | | | | | | |
| veg_leek_spread_dairy_vlowfat | 0% | 99% | 1% | 99% | 1% | 1756 | 98.75 | 10047 | 1.25 | | | | | | | | | | | | | | | | |
| veg_leek_spread_dunno_chol | 0% | 99% | 1% | 99% | 1% | 1756 | 98.75 | 3848 | 0.42 | 3243 | 0.42 | 2849 | 0.41 | | | | | | | | | | | | |
| veg_leek_spread_dunno_dunno | 0% | 99% | 1% | 99% | 1% | 1756 | 98.75 | 10047 | 0.31 | 10049 | 0.3125 | 10043 | 0.313 | 7775 | 0.31 | | | | | | | | | | |
| veg_leek_spread_dunno_fat | 0% | 99% | 1% | 99% | 1% | 1756 | 98.75 | 10043 | 0.63 | 7775 | 0.625 | | | | | | | | | | | | | | |
| veg_leek_spread_dunno_lowfat | 0% | 99% | 1% | 99% | 1% | 1756 | 98.75 | 10047 | 0.63 | 10049 | 0.625 | | | | | | | | | | | | | | |
| veg_leek_spread_dunno_vlowfat | 0% | 99% | 1% | 99% | 1% | 1756 | 98.75 | 10047 | 0.63 | 10049 | 0.625 | | | | | | | | | | | | | | |
| veg_leek_spread_olive_chol | 0% | 99% | 1% | 99% | 1% | 1756 | 98.75 | 3364 | 0.63 | 10053 | 0.625 | | | | | | | | | | | | | | |
| veg_leek_spread_olive_dunno | 0% | 99% | 1% | 99% | 1% | 1756 | 98.75 | 10048 | 0.63 | 10042 | 0.31 | 10131 | 0.31 | | | | | | | | | | | | |
| veg_leek_spread_olive_fat | 0% | 99% | 1% | 99% | 1% | 1756 | 98.75 | 10042 | 0.94 | 10131 | 0.3125 | | | | | | | | | | | | | | |
| veg_leek_spread_olive_lowfat | 0% | 99% | 1% | 99% | 1% | 1756 | 98.75 | 10048 | 1.25 | | | | | | | | | | | | | | | | |
| veg_leek_spread_olive_vlowfat | 0% | 99% | 1% | 99% | 1% | 1756 | 98.75 | 10048 | 1.25 | | | | | | | | | | | | | | | | |
| veg_lettuce | 0% | 100% | 0% | 100% | 0% | 1762 | 100 | | | | | | | | | | | | | | | | | | |
| veg_mixed | 0% | 100% | 0% | 100% | 0% | 1771 | 100 | | | | | | | | | | | | | | | | | | |
| veg_mixtures_butter_dunno | 0% | 99% | 1% | 10% | 90% | 8377 | 95 | 9407 | 1.25 | 10039 | 1.25 | 851 | 1.25 | 852 | 1.25 | | | | | | | | | | |
| veg_mixtures_butter_fat | 0% | 99% | 1% | 1% | 99% | 8377 | 95 | 851 | 2.5 | 852 | 2.5 | | | | | | | | | | | | | | |
| veg_mixtures_butter_lowfat | 0% | 99% | 1% | 40% | 60% | 8377 | 95 | 10140 | 5 | | | | | | | | | | | | | | | | |
| veg_mixtures_butter_spread_fat | 0% | 99% | 1% | 30% | 70% | 8377 | 95 | 9407 | 2.5 | 10039 | 2.5 | | | | | | | | | | | | | | |
| veg_mixtures_butter_spread_lowfat | 0% | 99% | 1% | 40% | 60% | 8377 | 95 | 10894 | 2.5 | 3891 | 2.5 | | | | | | | | | | | | | | |
| veg_mixtures_fat_dunno | 0% | 99% | 1% | 100% | 0% | 8377 | 95 | 875 | 5 | | | | | | | | | | | | | | | | |
| veg_mixtures_fat_none | 0% | 99% | 1% | 100% | 0% | 8377 | 100 | | | | | | | | | | | | | | | | | | |
| veg_mixtures_fat_other | 0% | 99% | 1% | 1% | 99% | 8377 | 95 | 856 | 2.5 | 855 | 2.5 | | | | | | | | | | | | | | |
| veg_mixtures_lard | 0% | 99% | 1% | 1% | 99% | 8377 | 95 | 858 | 5 | | | | | | | | | | | | | | | | |
| veg_mixtures_marg_hard | 0% | 99% | 1% | 99% | 1% | 8377 | 95 | 860 | 5 | | | | | | | | | | | | | | | | |
| veg_mixtures_marg_poly_chol | 0% | 99% | 1% | 90% | 10% | 8377 | 95 | 2849 | 1.7 | 3848 | 1.7 | 3243 | 1.6 | | | | | | | | | | | | |
| veg_mixtures_marg_poly_dunno | 0% | 99% | 1% | 90% | 10% | 8377 | 95 | 10049 | 2.5 | 10043 | 1.25 | 10044 | 1.25 | | | | | | | | | | | | |
| veg_mixtures_marg_poly_fat | 0% | 99% | 1% | 90% | 10% | 8377 | 95 | 10043 | 2.5 | 10044 | 2.5 | | | | | | | | | | | | | | |
| veg_mixtures_marg_poly_lowfat | 0% | 99% | 1% | 90% | 10% | 8377 | 95 | 10049 | 5 | | | | | | | | | | | | | | | | |
| veg_mixtures_marg_poly_vlowfat | 0% | 99% | 1% | 90% | 10% | 8377 | 95 | 10049 | 5 | | | | | | | | | | | | | | | | |
| veg_mixtures_marg_soya_chol | 0% | 99% | 1% | 99% | 1% | 8377 | 95 | 3848 | 0.83 | 3243 | 0.83 | 2849 | 0.84 | 10786 | 2.5 | | | | | | | | | | |
| veg_mixtures_marg_soya_dunno | 0% | 99% | 1% | 100% | 0% | 8377 | 95 | 10980 | 2.5 | 10786 | 2.5 | | | | | | | | | | | | | | |
| veg_mixtures_marg_soya_fat | 0% | 99% | 1% | 100% | 0% | 8377 | 95 | 10786 | 5 | | | | | | | | | | | | | | | | |
| veg_mixtures_marg_soya_lowfat | 0% | 99% | 1% | 100% | 0% | 8377 | 95 | 10980 | 5 | | | | | | | | | | | | | | | | |
| veg_mixtures_marg_soya_vlowfat | 0% | 99% | 1% | 100% | 0% | 8377 | 95 | 10980 | 5 | | | | | | | | | | | | | | | | |
| veg_mixtures_oil_olive | 0% | 100% | 0% | 100% | 0% | 8377 | 95 | 874 | 5 | | | | | | | | | | | | | | | | |
| veg_mixtures_oil_other | 0% | 100% | 0% | 100% | 0% | 8377 | 95 | 870 | 1.5 | 870 | 1 | 10963 | 1.25 | 871 | 0.63 | 5000 | 0.63 | | | | | | | | |
| veg_mixtures_oil_rapeseed | 0% | 100% | 0% | 100% | 0% | 8377 | 95 | 7990 | 5 | | | | | | | | | | | | | | | | |
| veg_mixtures_oil_sunflower | 0% | 100% | 0% | 100% | 0% | 8377 | 95 | 873 | 5 | | | | | | | | | | | | | | | | |
| veg_mixtures_oil_veg | 0% | 100% | 0% | 100% | 0% | 8377 | 95 | 872 | 2.5 | 871 | 2.5 | | | | | | | | | | | | | | |
| veg_mixtures_spread_dairy_chol | 0% | 99% | 1% | 99% | 1% | 8377 | 95 | 3848 | 5 | | | | | | | | | | | | | | | | |
| veg_mixtures_spread_dairy_dunno | 0% | 99% | 1% | 99% | 1% | 8377 | 95 | 10047 | 2.5 | 7775 | 2.5 | | | | | | | | | | | | | | |
| veg_mixtures_spread_dairy_fat | 0% | 99% | 1% | 99% | 1% | 8377 | 95 | 7775 | 5 | | | | | | | | | | | | | | | | |
| veg_mixtures_spread_dairy_lowfat | 0% | 99% | 1% | 99% | 1% | 8377 | 95 | 10047 | 5 | | | | | | | | | | | | | | | | |
| veg_mixtures_spread_dairy_vlowfat | 0% | 99% | 1% | 99% | 1% | 8377 | 95 | 10047 | 5 | | | | | | | | | | | | | | | | |
| veg_mixtures_spread_dunno_chol | 0% | 99% | 1% | 99% | 1% | 8377 | 95 | 3848 | 1.7 | 3243 | 1.7 | 2849 | 1.6 | | | | | | | | | | | | |
| veg_mixtures_spread_dunno_dunno | 0% | 99% | 1% | 99% | 1% | 8377 | 95 | 10047 | 1.25 | 10049 | 1.25 | 10043 | 1.25 | 7775 | 1.25 | | | | | | | | | | |
| veg_mixtures_spread_dunno_fat | 0% | 99% | 1% | 99% | 1% | 8377 | 95 | 10043 | 2.5 | 7775 | 2.5 | | | | | | | | | | | | | | |
| veg_mixtures_spread_dunno_lowfat | 0% | 99% | 1% | 99% | 1% | 8377 | 95 | 10047 | 2.5 | 10049 | 2.5 | | | | | | | | | | | | | | |
| veg_mixtures_spread_dunno_vlowfat | 0% | 99% | 1% | 99% | 1% | 8377 | 95 | 10047 | 2.5 | 10049 | 2.5 | | | | | | | | | | | | | | |
| veg_mixtures_spread_olive_chol | 0% | 99% | 1% | 99% | 1% | 8377 | 95 | 3364 | 2.5 | 10053 | 2.5 | | | | | | | | | | | | | | |
| veg_mixtures_spread_olive_dunno | 0% | 99% | 1% | 99% | 1% | 8377 | 95 | 10048 | 2.5 | 10042 | 1.25 | 10131 | 1.25 | | | | | | | | | | | | |
| veg_mixtures_spread_olive_fat | 0% | 99% | 1% | 99% | 1% | 8377 | 95 | 10042 | 3.75 | 10131 | 1.25 | | | | | | | | | | | | | | |
| veg_mixtures_spread_olive_lowfat | 0% | 99% | 1% | 99% | 1% | 8377 | 95 | 10048 | 5 | | | | | | | | | | | | | | | | |
| veg_mixtures_spread_olive_vlowfat | 0% | 99% | 1% | 99% | 1% | 8377 | 95 | 10048 | 5 | | | | | | | | | | | | | | | | |
| veg_mushrooms_butter_dunno | 0% | 99% | 1% | 10% | 90% | 1779 | 97.5 | 9407 | 0.63 | 10039 | 0.625 | 851 | 0.625 | 852 | 0.63 | | | | | | | | | | |
| veg_mushrooms_butter_fat | 0% | 99% | 1% | 1% | 99% | 1779 | 97.5 | 851 | 1.25 | 852 | 1.25 | | | | | | | | | | | | | | |
| veg_mushrooms_butter_lowfat | 0% | 99% | 1% | 40% | 60% | 1779 | 97.5 | 10140 | 2.5 | | | | | | | | | | | | | | | | |
| veg_mushrooms_butter_spread_fat | 0% | 99% | 1% | 30% | 70% | 1779 | 97.5 | 9407 | 1.25 | 10039 | 1.25 | | | | | | | | | | | | | | |
| veg_mushrooms_butter_spread_lowfat | 0% | 99% | 1% | 40% | 60% | 1779 | 97.5 | 10894 | 1.25 | 3891 | 1.25 | | | | | | | | | | | | | | |
| veg_mushrooms_fat_dunno | 0% | 99% | 1% | 100% | 0% | 1779 | 97.5 | 875 | 2.5 | | | | | | | | | | | | | | | | |
| veg_mushrooms_fat_none | 0% | 100% | 0% | 100% | 0% | 1779 | 100 | | | | | | | | | | | | | | | | | | |
| veg_mushrooms_fat_other | 0% | 99% | 1% | 1% | 99% | 1779 | 97.5 | 856 | 1.25 | 855 | 1.25 | | | | | | | | | | | | | | |
| veg_mushrooms_lard | 0% | 99% | 1% | 1% | 99% | 1779 | 97.5 | 858 | 2.5 | | | | | | | | | | | | | | | | |
| veg_mushrooms_marg_hard | 0% | 99% | 1% | 99% | 1% | 1779 | 97.5 | 860 | 2.5 | | | | | | | | | | | | | | | | |
| veg_mushrooms_marg_poly_chol | 0% | 99% | 1% | 90% | 10% | 1779 | 97.5 | 2849 | 0.84 | 3848 | 0.83 | 3243 | 0.83 | | | | | | | | | | | | |
| veg_mushrooms_marg_poly_dunno | 0% | 99% | 1% | 90% | 10% | 1779 | 97.5 | 10049 | 1.25 | 10043 | 0.625 | 10044 | 0.625 | | | | | | | | | | | | |
| veg_mushrooms_marg_poly_fat | 0% | 99% | 1% | 90% | 10% | 1779 | 97.5 | 10043 | 1.25 | 10044 | 1.25 | | | | | | | | | | | | | | |
| veg_mushrooms_marg_poly_lowfat | 0% | 99% | 1% | 90% | 10% | 1779 | 97.5 | 10049 | 2.5 | | | | | | | | | | | | | | | | |
| veg_mushrooms_marg_poly_vlowfat | 0% | 99% | 1% | 90% | 10% | 1779 | 97.5 | 10049 | 2.5 | | | | | | | | | | | | | | | | |
| veg_mushrooms_marg_soya_chol | 0% | 99% | 1% | 99% | 1% | 1779 | 97.5 | 3848 | 0.42 | 3243 | 0.41 | 2849 | 0.42 | 10786 | 1.25 | | | | | | | | | | |
| veg_mushrooms_marg_soya_dunno | 0% | 99% | 1% | 100% | 0% | 1779 | 97.5 | 10980 | 1.25 | 10786 | 1.25 | | | | | | | | | | | | | | |
| veg_mushrooms_marg_soya_fat | 0% | 99% | 1% | 100% | 0% | 1779 | 97.5 | 10786 | 2.5 | | | | | | | | | | | | | | | | |
| veg_mushrooms_marg_soya_lowfat | 0% | 99% | 1% | 100% | 0% | 1779 | 97.5 | 10980 | 2.5 | | | | | | | | | | | | | | | | |
| veg_mushrooms_marg_soya_vlowfat | 0% | 99% | 1% | 100% | 0% | 1779 | 97.5 | 10980 | 2.5 | | | | | | | | | | | | | | | | |
| veg_mushrooms_oil_olive | 0% | 100% | 0% | 100% | 0% | 1779 | 97.5 | 874 | 2.5 | | | | | | | | | | | | | | | | |
| veg_mushrooms_oil_other | 0% | 100% | 0% | 100% | 0% | 1779 | 97.5 | 870 | 0.75 | 870 | 0.5 | 10963 | 0.625 | 871 | 0.31 | 5000 | 0.31 | | | | | | | | |
| veg_mushrooms_oil_rapeseed | 0% | 100% | 0% | 100% | 0% | 1779 | 97.5 | 7990 | 2.5 | | | | | | | | | | | | | | | | |
| veg_mushrooms_oil_sunflower | 0% | 100% | 0% | 100% | 0% | 1779 | 97.5 | 873 | 2.5 | | | | | | | | | | | | | | | | |
| veg_mushrooms_oil_veg | 0% | 100% | 0% | 100% | 0% | 1779 | 97.5 | 872 | 1.25 | 871 | 1.25 | | | | | | | | | | | | | | |
| veg_mushrooms_spread_dairy_chol | 0% | 99% | 1% | 99% | 1% | 1779 | 97.5 | 3848 | 2.5 | | | | | | | | | | | | | | | | |
| veg_mushrooms_spread_dairy_dunno | 0% | 99% | 1% | 99% | 1% | 1779 | 97.5 | 10047 | 1.25 | 7775 | 1.25 | | | | | | | | | | | | | | |
| veg_mushrooms_spread_dairy_fat | 0% | 99% | 1% | 99% | 1% | 1779 | 97.5 | 7775 | 2.5 | | | | | | | | | | | | | | | | |
| veg_mushrooms_spread_dairy_lowfat | 0% | 99% | 1% | 99% | 1% | 1779 | 97.5 | 10047 | 2.5 | | | | | | | | | | | | | | | | |
| veg_mushrooms_spread_dairy_vlowfat | 0% | 99% | 1% | 99% | 1% | 1779 | 97.5 | 10047 | 2.5 | | | | | | | | | | | | | | | | |
| veg_mushrooms_spread_dunno_chol | 0% | 99% | 1% | 99% | 1% | 1779 | 97.5 | 3848 | 0.84 | 3243 | 0.83 | 2849 | 0.83 | | | | | | | | | | | | |
| veg_mushrooms_spread_dunno_dunno | 0% | 99% | 1% | 99% | 1% | 1779 | 97.5 | 10047 | 0.63 | 10049 | 0.625 | 10043 | 0.625 | 7775 | 0.63 | | | | | | | | | | |
| veg_mushrooms_spread_dunno_fat | 0% | 99% | 1% | 99% | 1% | 1779 | 97.5 | 10043 | 1.25 | 7775 | 1.25 | | | | | | | | | | | | | | |
| veg_mushrooms_spread_dunno_lowfat | 0% | 99% | 1% | 99% | 1% | 1779 | 97.5 | 10047 | 1.25 | 10049 | 1.25 | | | | | | | | | | | | | | |
| veg_mushrooms_spread_dunno_vlowfat | 0% | 99% | 1% | 99% | 1% | 1779 | 97.5 | 10047 | 1.25 | 10049 | 1.25 | | | | | | | | | | | | | | |
| veg_mushrooms_spread_olive_chol | 0% | 99% | 1% | 99% | 1% | 1779 | 97.5 | 3364 | 1.25 | 10053 | 1.25 | | | | | | | | | | | | | | |
| veg_mushrooms_spread_olive_dunno | 0% | 99% | 1% | 99% | 1% | 1779 | 97.5 | 10048 | 1.25 | 10042 | 0.625 | 10131 | 0.625 | | | | | | | | | | | | |
| veg_mushrooms_spread_olive_fat | 0% | 99% | 1% | 99% | 1% | 1779 | 97.5 | 10042 | 1.88 | 10131 | 0.625 | | | | | | | | | | | | | | |
| veg_mushrooms_spread_olive_lowfat | 0% | 99% | 1% | 99% | 1% | 1779 | 97.5 | 10048 | 2.5 | | | | | | | | | | | | | | | | |
| veg_mushrooms_spread_olive_vlowfat | 0% | 99% | 1% | 99% | 1% | 1779 | 97.5 | 10048 | 2.5 | | | | | | | | | | | | | | | | |
| veg_onion_butter_dunno | 0% | 99% | 1% | 10% | 90% | 1785 | 20 | 9293 | 77.5 | 9407 | 0.625 | 10039 | 0.625 | 851 | 0.63 | 852 | 0.63 | | | | | | | | |
| veg_onion_butter_fat | 0% | 99% | 1% | 1% | 99% | 1785 | 20 | 9293 | 77.5 | 851 | 1.25 | 852 | 1.25 | | | | | | | | | | | | |
| veg_onion_butter_lowfat | 0% | 99% | 1% | 40% | 60% | 1785 | 20 | 9293 | 77.5 | 10140 | 2.5 | | | | | | | | | | | | | | |
| veg_onion_butter_spread_fat | 0% | 99% | 1% | 30% | 70% | 1785 | 20 | 9293 | 77.5 | 9407 | 1.25 | 10039 | 1.25 | | | | | | | | | | | | |
| veg_onion_butter_spread_lowfat | 0% | 99% | 1% | 40% | 60% | 1785 | 20 | 9293 | 77.5 | 10894 | 1.25 | 3891 | 1.25 | | | | | | | | | | | | |
| veg_onion_fat_dunno | 0% | 99% | 1% | 100% | 0% | 1785 | 20 | 9293 | 77.5 | 875 | 2.5 | | | | | | | | | | | | | | |
| veg_onion_fat_none | 0% | 100% | 0% | 100% | 0% | 1785 | 20 | 9293 | 80 | | | | | | | | | | | | | | | | |
| veg_onion_fat_other | 0% | 99% | 1% | 1% | 99% | 1785 | 20 | 9293 | 77.5 | 856 | 1.25 | 855 | 1.25 | | | | | | | | | | | | |
| veg_onion_lard | 0% | 99% | 1% | 1% | 99% | 1785 | 20 | 9293 | 77.5 | 858 | 2.5 | | | | | | | | | | | | | | |
| veg_onion_marg_hard | 0% | 99% | 1% | 99% | 1% | 1785 | 20 | 9293 | 77.5 | 860 | 2.5 | | | | | | | | | | | | | | |
| veg_onion_marg_poly_chol | 0% | 99% | 1% | 90% | 10% | 1785 | 20 | 9293 | 77.5 | 2849 | 0.84 | 3848 | 0.83 | 3243 | 0.83 | | | | | | | | | | |
| veg_onion_marg_poly_dunno | 0% | 99% | 1% | 90% | 10% | 1785 | 20 | 9293 | 77.5 | 10049 | 1.25 | 10043 | 0.625 | 10044 | 0.63 | | | | | | | | | | |
| veg_onion_marg_poly_fat | 0% | 99% | 1% | 90% | 10% | 1785 | 20 | 9293 | 77.5 | 10043 | 1.25 | 10044 | 1.25 | | | | | | | | | | | | |
| veg_onion_marg_poly_lowfat | 0% | 99% | 1% | 90% | 10% | 1785 | 20 | 9293 | 77.5 | 10049 | 2.5 | | | | | | | | | | | | | | |
| veg_onion_marg_poly_vlowfat | 0% | 99% | 1% | 90% | 10% | 1785 | 20 | 9293 | 77.5 | 10049 | 2.5 | | | | | | | | | | | | | | |
| veg_onion_marg_soya_chol | 0% | 99% | 1% | 99% | 1% | 1785 | 20 | 9293 | 77.5 | 3848 | 0.42 | 3243 | 0.41 | 2849 | 0.42 | 10786 | 1.25 | | | | | | | | |
| veg_onion_marg_soya_dunno | 0% | 99% | 1% | 100% | 0% | 1785 | 20 | 9293 | 77.5 | 10980 | 1.25 | 10786 | 1.25 | | | | | | | | | | | | |
| veg_onion_marg_soya_fat | 0% | 99% | 1% | 100% | 0% | 1785 | 20 | 9293 | 77.5 | 10786 | 2.5 | | | | | | | | | | | | | | |
| veg_onion_marg_soya_lowfat | 0% | 99% | 1% | 100% | 0% | 1785 | 20 | 9293 | 77.5 | 10980 | 2.5 | | | | | | | | | | | | | | |
| veg_onion_marg_soya_vlowfat | 0% | 99% | 1% | 100% | 0% | 1785 | 20 | 9293 | 77.5 | 10980 | 2.5 | | | | | | | | | | | | | | |
| veg_onion_oil_olive | 0% | 100% | 0% | 100% | 0% | 1785 | 20 | 9293 | 77.5 | 874 | 2.5 | | | | | | | | | | | | | | |
| veg_onion_oil_other | 0% | 100% | 0% | 100% | 0% | 1785 | 20 | 9293 | 77.5 | 870 | 0.75 | 870 | 0.5 | 10963 | 0.63 | 871 | 0.31 | 5000 | 0.313 | | | | | | |
| veg_onion_oil_rapeseed | 0% | 100% | 0% | 100% | 0% | 1785 | 20 | 9293 | 77.5 | 7990 | 2.5 | | | | | | | | | | | | | | |
| veg_onion_oil_sunflower | 0% | 100% | 0% | 100% | 0% | 1785 | 20 | 9293 | 77.5 | 873 | 2.5 | | | | | | | | | | | | | | |
| veg_onion_oil_veg | 0% | 100% | 0% | 100% | 0% | 1785 | 20 | 9293 | 77.5 | 872 | 1.25 | 871 | 1.25 | | | | | | | | | | | | |
| veg_onion_spread_dairy_chol | 0% | 99% | 1% | 99% | 1% | 1785 | 20 | 9293 | 77.5 | 3848 | 2.5 | | | | | | | | | | | | | | |
| veg_onion_spread_dairy_dunno | 0% | 99% | 1% | 99% | 1% | 1785 | 20 | 9293 | 77.5 | 10047 | 1.25 | 7775 | 1.25 | | | | | | | | | | | | |
| veg_onion_spread_dairy_fat | 0% | 99% | 1% | 99% | 1% | 1785 | 20 | 9293 | 77.5 | 7775 | 2.5 | | | | | | | | | | | | | | |
| veg_onion_spread_dairy_lowfat | 0% | 99% | 1% | 99% | 1% | 1785 | 20 | 9293 | 77.5 | 10047 | 2.5 | | | | | | | | | | | | | | |
| veg_onion_spread_dairy_vlowfat | 0% | 99% | 1% | 99% | 1% | 1785 | 20 | 9293 | 77.5 | 10047 | 2.5 | | | | | | | | | | | | | | |
| veg_onion_spread_dunno_chol | 0% | 99% | 1% | 99% | 1% | 1785 | 20 | 9293 | 77.5 | 3848 | 0.84 | 3243 | 0.83 | 2849 | 0.83 | | | | | | | | | | |
| veg_onion_spread_dunno_dunno | 0% | 99% | 1% | 99% | 1% | 1785 | 20 | 9293 | 77.5 | 10047 | 0.625 | 10049 | 0.625 | 10043 | 0.63 | 7775 | 0.63 | | | | | | | | |
| veg_onion_spread_dunno_fat | 0% | 99% | 1% | 99% | 1% | 1785 | 20 | 9293 | 77.5 | 10043 | 1.25 | 7775 | 1.25 | | | | | | | | | | | | |
| veg_onion_spread_dunno_lowfat | 0% | 99% | 1% | 99% | 1% | 1785 | 20 | 9293 | 77.5 | 10047 | 1.25 | 10049 | 1.25 | | | | | | | | | | | | |
| veg_onion_spread_dunno_vlowfat | 0% | 99% | 1% | 99% | 1% | 1785 | 20 | 9293 | 77.5 | 10047 | 1.25 | 10049 | 1.25 | | | | | | | | | | | | |
| veg_onion_spread_olive_chol | 0% | 99% | 1% | 99% | 1% | 1785 | 20 | 9293 | 77.5 | 3364 | 1.25 | 10053 | 1.25 | | | | | | | | | | | | |
| veg_onion_spread_olive_dunno | 0% | 99% | 1% | 99% | 1% | 1785 | 20 | 9293 | 77.5 | 10048 | 1.25 | 10042 | 0.625 | 10131 | 0.63 | | | | | | | | | | |
| veg_onion_spread_olive_fat | 0% | 99% | 1% | 99% | 1% | 1785 | 20 | 9293 | 77.5 | 10042 | 1.875 | 10131 | 0.625 | | | | | | | | | | | | |
| veg_onion_spread_olive_lowfat | 0% | 99% | 1% | 99% | 1% | 1785 | 20 | 9293 | 77.5 | 10048 | 2.5 | | | | | | | | | | | | | | |
| veg_onion_spread_olive_vlowfat | 0% | 99% | 1% | 99% | 1% | 1785 | 20 | 9293 | 77.5 | 10048 | 2.5 | | | | | | | | | | | | | | |
| veg_other_butter_dunno | 0% | 99% | 1% | 10% | 90% | 8478 | 5 | 1655 | 42.5 | 1908 | 5 | 1724 | 5 | 8471 | 34.5 | 1909 | 5 | 9407 | 0.75 | 10039 | 0.75 | 851 | 0.75 | 852 | 0.75 |
| veg_other_butter_fat | 0% | 99% | 1% | 1% | 99% | 8478 | 5 | 1655 | 42.5 | 1908 | 5 | 1724 | 5 | 8471 | 34.5 | 1909 | 5 | 851 | 1.5 | 852 | 1.5 | | | | |
| veg_other_butter_lowfat | 0% | 99% | 1% | 40% | 60% | 8478 | 5 | 1655 | 42.5 | 1908 | 5 | 1724 | 5 | 8471 | 34.5 | 1909 | 5 | 10140 | 3 | | | | | | |
| veg_other_butter_spread_fat | 0% | 99% | 1% | 30% | 70% | 8478 | 5 | 1655 | 42.5 | 1908 | 5 | 1724 | 5 | 8471 | 34.5 | 1909 | 5 | 3891 | 1.5 | 10894 | 1.5 | | | | |
| veg_other_butter_spread_lowfat | 0% | 99% | 1% | 40% | 60% | 8478 | 5 | 1655 | 42.5 | 1908 | 5 | 1724 | 5 | 8471 | 34.5 | 1909 | 5 | 9407 | 1.5 | 10039 | 1.5 | | | | |
| veg_other_fat_dunno | 0% | 99% | 1% | 100% | 0% | 8478 | 5 | 1655 | 42.5 | 1908 | 5 | 1724 | 5 | 8471 | 34.5 | 1909 | 5 | 875 | 3 | | | | | | |
| veg_other_fat_none | 0% | 100% | 0% | 100% | 0% | 8478 | 5 | 1655 | 44 | 1908 | 5 | 1724 | 5 | 8471 | 36 | 1909 | 5 | | | | | | | | |
| veg_other_fat_other | 0% | 99% | 1% | 1% | 99% | 8478 | 5 | 1655 | 42.5 | 1908 | 5 | 1724 | 5 | 8471 | 34.5 | 1909 | 5 | 856 | 1.5 | 855 | 1.5 | | | | |
| veg_other_lard | 0% | 99% | 1% | 1% | 99% | 8478 | 5 | 1655 | 42.5 | 1908 | 5 | 1724 | 5 | 8471 | 34.5 | 1909 | 5 | 858 | 3 | | | | | | |
| veg_other_marg_hard | 0% | 99% | 1% | 99% | 1% | 8478 | 5 | 1655 | 42.5 | 1908 | 5 | 1724 | 5 | 8471 | 34.5 | 1909 | 5 | 860 | 3 | | | | | | |
| veg_other_marg_poly_chol | 0% | 99% | 1% | 90% | 10% | 8478 | 5 | 1655 | 42.5 | 1908 | 5 | 1724 | 5 | 8471 | 34.5 | 1909 | 5 | 2849 | 1 | 3848 | 1 | 3243 | 1 | | |
| veg_other_marg_poly_dunno | 0% | 99% | 1% | 90% | 10% | 8478 | 5 | 1655 | 42.5 | 1908 | 5 | 1724 | 5 | 8471 | 34.5 | 1909 | 5 | 10049 | 1.5 | 10043 | 0.75 | 10044 | 0.75 | | |
| veg_other_marg_poly_fat | 0% | 99% | 1% | 90% | 10% | 8478 | 5 | 1655 | 42.5 | 1908 | 5 | 1724 | 5 | 8471 | 34.5 | 1909 | 5 | 10043 | 1.5 | 10044 | 1.5 | | | | |
| veg_other_marg_poly_lowfat | 0% | 99% | 1% | 90% | 10% | 8478 | 5 | 1655 | 42.5 | 1908 | 5 | 1724 | 5 | 8471 | 34.5 | 1909 | 5 | 10049 | 3 | | | | | | |

| | | | | | | | | | | | | | | | | | | | | | | | | | | | |
|---|---|---|---|---|---|---|---|---|---|---|---|---|---|---|---|---|---|---|---|---|---|---|---|---|---|---|---|
| veg_other_marg_poly_vlowfat | 0% | 99% | 1% | 90% | 10% | 8478 | 5 | 1655 | 42.5 | 1908 | 5 | 1724 | 5 | 8471 | 34.5 | 1909 | 5 | 10049 | 3 | | | | | | | | |
| veg_other_marg_soya_chol | 0% | 99% | 1% | 99% | 1% | 8478 | 5 | 1655 | 42.5 | 1908 | 5 | 1724 | 5 | 8471 | 34.5 | 1909 | 5 | 3848 | 0.5 | 3243 | 0.5 | 2849 | 0.5 | 10786 | 1.5 | | |
| veg_other_marg_soya_dunno | 0% | 99% | 1% | 100% | 0% | 8478 | 5 | 1655 | 42.5 | 1908 | 5 | 1724 | 5 | 8471 | 34.5 | 1909 | 5 | 10980 | 1.5 | 10786 | 1.5 | | | | | | |
| veg_other_marg_soya_fat | 0% | 99% | 1% | 100% | 0% | 8478 | 5 | 1655 | 42.5 | 1908 | 5 | 1724 | 5 | 8471 | 34.5 | 1909 | 5 | 10786 | 3 | | | | | | | | |
| veg_other_marg_soya_lowfat | 0% | 99% | 1% | 100% | 0% | 8478 | 5 | 1655 | 42.5 | 1908 | 5 | 1724 | 5 | 8471 | 34.5 | 1909 | 5 | 10980 | 3 | | | | | | | | |
| veg_other_marg_soya_vlowfat | 0% | 99% | 1% | 100% | 0% | 8478 | 5 | 1655 | 42.5 | 1908 | 5 | 1724 | 5 | 8471 | 34.5 | 1909 | 5 | 10980 | 3 | | | | | | | | |
| veg_other_oil_olive | 0% | 100% | 0% | 100% | 0% | 8478 | 5 | 1655 | 42.5 | 1908 | 5 | 1724 | 5 | 8471 | 34.5 | 1909 | 5 | 874 | 3 | | | | | | | | |
| veg_other_oil_other | 0% | 100% | 0% | 100% | 0% | 8478 | 5 | 1655 | 42.5 | 1908 | 5 | 1724 | 5 | 8471 | 34.5 | 1909 | 5 | 870 | 0.9 | 870 | 0.6 | 10963 | 0.75 | 871 | 0.375 | 5000 | 0.375 |
| veg_other_oil_rapeseed | 0% | 100% | 0% | 100% | 0% | 8478 | 5 | 1655 | 42.5 | 1908 | 5 | 1724 | 5 | 8471 | 34.5 | 1909 | 5 | 7990 | 3 | | | | | | | | |
| veg_other_oil_sunflower | 0% | 100% | 0% | 100% | 0% | 8478 | 5 | 1655 | 42.5 | 1908 | 5 | 1724 | 5 | 8471 | 34.5 | 1909 | 5 | 873 | 3 | | | | | | | | |
| veg_other_oil_veg | 0% | 100% | 0% | 100% | 0% | 8478 | 5 | 1655 | 42.5 | 1908 | 5 | 1724 | 5 | 8471 | 34.5 | 1909 | 5 | 872 | 1.5 | 871 | 1.5 | | | | | | |
| veg_other_spread_dairy_chol | 0% | 99% | 1% | 99% | 1% | 8478 | 5 | 1655 | 42.5 | 1908 | 5 | 1724 | 5 | 8471 | 34.5 | 1909 | 5 | 3848 | 3 | | | | | | | | |
| veg_other_spread_dairy_dunno | 0% | 99% | 1% | 99% | 1% | 8478 | 5 | 1655 | 42.5 | 1908 | 5 | 1724 | 5 | 8471 | 34.5 | 1909 | 5 | 10047 | 1.5 | 7775 | 1.5 | | | | | | |
| veg_other_spread_dairy_fat | 0% | 99% | 1% | 99% | 1% | 8478 | 5 | 1655 | 42.5 | 1908 | 5 | 1724 | 5 | 8471 | 34.5 | 1909 | 5 | 7775 | 3 | | | | | | | | |
| veg_other_spread_dairy_lowfat | 0% | 99% | 1% | 99% | 1% | 8478 | 5 | 1655 | 42.5 | 1908 | 5 | 1724 | 5 | 8471 | 34.5 | 1909 | 5 | 10047 | 3 | | | | | | | | |
| veg_other_spread_dairy_vlowfat | 0% | 99% | 1% | 99% | 1% | 8478 | 5 | 1655 | 42.5 | 1908 | 5 | 1724 | 5 | 8471 | 34.5 | 1909 | 5 | 10047 | 3 | | | | | | | | |
| veg_other_spread_dunno_chol | 0% | 99% | 1% | 99% | 1% | 8478 | 5 | 1655 | 42.5 | 1908 | 5 | 1724 | 5 | 8471 | 34.5 | 1909 | 5 | 3848 | 1 | 3243 | 1 | 2849 | 1 | | | | |
| veg_other_spread_dunno_dunno | 0% | 99% | 1% | 99% | 1% | 8478 | 5 | 1655 | 42.5 | 1908 | 5 | 1724 | 5 | 8471 | 34.5 | 1909 | 5 | 10047 | 0.75 | 10049 | 0.75 | 10043 | 0.75 | 7775 | 0.75 | | |
| veg_other_spread_dunno_fat | 0% | 99% | 1% | 99% | 1% | 8478 | 5 | 1655 | 42.5 | 1908 | 5 | 1724 | 5 | 8471 | 34.5 | 1909 | 5 | 10043 | 1.5 | 7775 | 1.5 | | | | | | |
| veg_other_spread_dunno_lowfat | 0% | 99% | 1% | 99% | 1% | 8478 | 5 | 1655 | 42.5 | 1908 | 5 | 1724 | 5 | 8471 | 34.5 | 1909 | 5 | 10047 | 1.5 | 10049 | 1.5 | | | | | | |
| veg_other_spread_dunno_vlowfat | 0% | 99% | 1% | 99% | 1% | 8478 | 5 | 1655 | 42.5 | 1908 | 5 | 1724 | 5 | 8471 | 34.5 | 1909 | 5 | 10047 | 1.5 | 10049 | 1.5 | | | | | | |
| veg_other_spread_olive_chol | 0% | 99% | 1% | 99% | 1% | 8478 | 5 | 1655 | 42.5 | 1908 | 5 | 1724 | 5 | 8471 | 34.5 | 1909 | 5 | 3364 | 1.5 | 10053 | 1.5 | | | | | | |
| veg_other_spread_olive_dunno | 0% | 99% | 1% | 99% | 1% | 8478 | 5 | 1655 | 42.5 | 1908 | 5 | 1724 | 5 | 8471 | 34.5 | 1909 | 5 | 10048 | 1.5 | 10042 | 0.75 | 10131 | 0.75 | | | | |
| veg_other_spread_olive_fat | 0% | 99% | 1% | 99% | 1% | 8478 | 5 | 1655 | 42.5 | 1908 | 5 | 1724 | 5 | 8471 | 34.5 | 1909 | 5 | 10042 | 2.25 | 10131 | 0.75 | | | | | | |
| veg_other_spread_olive_lowfat | 0% | 99% | 1% | 99% | 1% | 8478 | 5 | 1655 | 42.5 | 1908 | 5 | 1724 | 5 | 8471 | 34.5 | 1909 | 5 | 10048 | 3 | | | | | | | | |
| veg_other_spread_olive_vlowfat | 0% | 99% | 1% | 99% | 1% | 8478 | 5 | 1655 | 42.5 | 1908 | 5 | 1724 | 5 | 8471 | 34.5 | 1909 | 5 | 10048 | 3 | | | | | | | | |
| veg_parsnip_butter_dunno | 0% | 99% | 1% | 10% | 90% | 1801 | 97.5 | 9407 | 0.63 | 10039 | 0.625 | 851 | 0.625 | 852 | 0.63 | | | | | | | | | | | | |
| veg_parsnip_butter_fat | 0% | 99% | 1% | 1% | 99% | 1801 | 97.5 | 851 | 1.25 | 852 | 1.25 | | | | | | | | | | | | | | | | |
| veg_parsnip_butter_lowfat | 0% | 99% | 1% | 40% | 60% | 1801 | 97.5 | 10140 | 2.5 | | | | | | | | | | | | | | | | | | |
| veg_parsnip_butter_spread_fat | 0% | 99% | 1% | 30% | 70% | 1801 | 97.5 | 9407 | 1.25 | 10039 | 1.25 | | | | | | | | | | | | | | | | |
| veg_parsnip_butter_spread_lowfat | 0% | 99% | 1% | 40% | 60% | 1801 | 97.5 | 10894 | 1.25 | 3891 | 1.25 | | | | | | | | | | | | | | | | |
| veg_parsnip_fat_dunno | 0% | 99% | 1% | 100% | 0% | 1801 | 97.5 | 875 | 2.5 | | | | | | | | | | | | | | | | | | |
| veg_parsnip_fat_none | 0% | 100% | 0% | 100% | 0% | 1801 | 100 | | | | | | | | | | | | | | | | | | | | |
| veg_parsnip_fat_other | 0% | 99% | 1% | 1% | 99% | 1801 | 97.5 | 856 | 1.25 | 855 | 1.25 | | | | | | | | | | | | | | | | |
| veg_parsnip_lard | 0% | 99% | 1% | 1% | 99% | 1801 | 97.5 | 858 | 2.5 | | | | | | | | | | | | | | | | | | |
| veg_parsnip_marg_hard | 0% | 99% | 1% | 99% | 1% | 1801 | 97.5 | 860 | 2.5 | | | | | | | | | | | | | | | | | | |
| veg_parsnip_marg_poly_chol | 0% | 99% | 1% | 90% | 10% | 1801 | 97.5 | 2849 | 0.84 | 3848 | 0.83 | 3243 | 0.83 | | | | | | | | | | | | | | |
| veg_parsnip_marg_poly_dunno | 0% | 99% | 1% | 90% | 10% | 1801 | 97.5 | 10049 | 1.25 | 10043 | 0.625 | 10044 | 0.625 | | | | | | | | | | | | | | |
| veg_parsnip_marg_poly_fat | 0% | 99% | 1% | 90% | 10% | 1801 | 97.5 | 10043 | 1.25 | 10044 | 1.25 | | | | | | | | | | | | | | | | |
| veg_parsnip_marg_poly_lowfat | 0% | 99% | 1% | 90% | 10% | 1801 | 97.5 | 10049 | 2.5 | | | | | | | | | | | | | | | | | | |
| veg_parsnip_marg_poly_vlowfat | 0% | 99% | 1% | 90% | 10% | 1801 | 97.5 | 10049 | 2.5 | | | | | | | | | | | | | | | | | | |
| veg_parsnip_marg_soya_chol | 0% | 99% | 1% | 99% | 1% | 1801 | 97.5 | 3848 | 0.42 | 3243 | 0.41 | 2849 | 0.42 | 10786 | 1.25 | | | | | | | | | | | | |
| veg_parsnip_marg_soya_dunno | 0% | 99% | 1% | 100% | 0% | 1801 | 97.5 | 10980 | 1.25 | 10786 | 1.25 | | | | | | | | | | | | | | | | |
| veg_parsnip_marg_soya_fat | 0% | 99% | 1% | 100% | 0% | 1801 | 97.5 | 10786 | 2.5 | | | | | | | | | | | | | | | | | | |
| veg_parsnip_marg_soya_lowfat | 0% | 99% | 1% | 100% | 0% | 1801 | 97.5 | 10980 | 2.5 | | | | | | | | | | | | | | | | | | |
| veg_parsnip_marg_soya_vlowfat | 0% | 99% | 1% | 100% | 0% | 1801 | 97.5 | 10980 | 2.5 | | | | | | | | | | | | | | | | | | |
| veg_parsnip_oil_olive | 0% | 100% | 0% | 100% | 0% | 1801 | 97.5 | 874 | 2.5 | | | | | | | | | | | | | | | | | | |
| veg_parsnip_oil_other | 0% | 100% | 0% | 100% | 0% | 1801 | 97.5 | 870 | 0.75 | 870 | 0.5 | 10963 | 0.625 | 871 | 0.31 | 5000 | 0.31 | | | | | | | | | | |
| veg_parsnip_oil_rapeseed | 0% | 100% | 0% | 100% | 0% | 1801 | 97.5 | 7990 | 2.5 | | | | | | | | | | | | | | | | | | |
| veg_parsnip_oil_sunflower | 0% | 100% | 0% | 100% | 0% | 1801 | 97.5 | 873 | 2.5 | | | | | | | | | | | | | | | | | | |
| veg_parsnip_oil_veg | 0% | 100% | 0% | 100% | 0% | 1801 | 97.5 | 872 | 1.25 | 871 | 1.25 | | | | | | | | | | | | | | | | |
| veg_parsnip_spread_dairy_chol | 0% | 99% | 1% | 99% | 1% | 1801 | 97.5 | 3848 | 2.5 | | | | | | | | | | | | | | | | | | |
| veg_parsnip_spread_dairy_dunno | 0% | 99% | 1% | 99% | 1% | 1801 | 97.5 | 10047 | 1.25 | 7775 | 1.25 | | | | | | | | | | | | | | | | |
| veg_parsnip_spread_dairy_fat | 0% | 99% | 1% | 99% | 1% | 1801 | 97.5 | 7775 | 2.5 | | | | | | | | | | | | | | | | | | |
| veg_parsnip_spread_dairy_lowfat | 0% | 99% | 1% | 99% | 1% | 1801 | 97.5 | 10047 | 2.5 | | | | | | | | | | | | | | | | | | |
| veg_parsnip_spread_dairy_vlowfat | 0% | 99% | 1% | 99% | 1% | 1801 | 97.5 | 10047 | 2.5 | | | | | | | | | | | | | | | | | | |
| veg_parsnip_spread_dunno_chol | 0% | 99% | 1% | 99% | 1% | 1801 | 97.5 | 3848 | 0.84 | 3243 | 0.83 | 2849 | 0.83 | | | | | | | | | | | | | | |
| veg_parsnip_spread_dunno_dunno | 0% | 99% | 1% | 99% | 1% | 1801 | 97.5 | 10047 | 0.63 | 10049 | 0.625 | 10043 | 0.625 | 7775 | 0.63 | | | | | | | | | | | | |
| veg_parsnip_spread_dunno_fat | 0% | 99% | 1% | 99% | 1% | 1801 | 97.5 | 10043 | 1.25 | 7775 | 1.25 | | | | | | | | | | | | | | | | |
| veg_parsnip_spread_dunno_lowfat | 0% | 99% | 1% | 99% | 1% | 1801 | 97.5 | 10047 | 1.25 | 10049 | 1.25 | | | | | | | | | | | | | | | | |
| veg_parsnip_spread_dunno_vlowfat | 0% | 99% | 1% | 99% | 1% | 1801 | 97.5 | 10047 | 1.25 | 10049 | 1.25 | | | | | | | | | | | | | | | | |
| veg_parsnip_spread_olive_chol | 0% | 99% | 1% | 99% | 1% | 1801 | 97.5 | 3364 | 1.25 | 10053 | 1.25 | | | | | | | | | | | | | | | | |
| veg_parsnip_spread_olive_dunno | 0% | 99% | 1% | 99% | 1% | 1801 | 97.5 | 10048 | 1.25 | 10042 | 0.625 | 10131 | 0.625 | | | | | | | | | | | | | | |
| veg_parsnip_spread_olive_fat | 0% | 99% | 1% | 99% | 1% | 1801 | 97.5 | 10042 | 1.88 | 10131 | 0.625 | | | | | | | | | | | | | | | | |
| veg_parsnip_spread_olive_lowfat | 0% | 99% | 1% | 99% | 1% | 1801 | 97.5 | 10048 | 2.5 | | | | | | | | | | | | | | | | | | |
| veg_parsnip_spread_olive_vlowfat | 0% | 99% | 1% | 99% | 1% | 1801 | 97.5 | 10048 | 2.5 | | | | | | | | | | | | | | | | | | |
| veg_peas | 0% | 100% | 0% | 100% | 0% | 11105 | 90 | 10796 | 10 | | | | | | | | | | | | | | | | | | |
| veg_pepper_bell_butter_dunno | 0% | 99% | 1% | 10% | 90% | 1824 | 19.5 | 7857 | 19.5 | 11102 | 19.5 | 7988 | 19.5 | 7989 | 19.5 | 9407 | 0.63 | 10039 | 0.625 | 851 | 0.63 | 852 | 0.63 | | | | |
| veg_pepper_bell_butter_fat | 0% | 99% | 1% | 1% | 99% | 1824 | 19.5 | 7857 | 19.5 | 11102 | 19.5 | 7988 | 19.5 | 7989 | 19.5 | 851 | 1.25 | 852 | 1.25 | | | | | | | | |
| veg_pepper_bell_butter_lowfat | 0% | 99% | 1% | 40% | 60% | 1824 | 19.5 | 7857 | 19.5 | 11102 | 19.5 | 7988 | 19.5 | 7989 | 19.5 | 10140 | 2.5 | | | | | | | | | | |
| veg_pepper_bell_butter_spread_fat | 0% | 99% | 1% | 30% | 70% | 1824 | 19.5 | 7857 | 19.5 | 11102 | 19.5 | 7988 | 19.5 | 7989 | 19.5 | 9407 | 1.25 | 10039 | 1.25 | | | | | | | | |
| veg_pepper_bell_butter_spread_lowfat | 0% | 99% | 1% | 40% | 60% | 1824 | 19.5 | 7857 | 19.5 | 11102 | 19.5 | 7988 | 19.5 | 7989 | 19.5 | 10894 | 1.25 | 3891 | 1.25 | | | | | | | | |
| veg_pepper_bell_fat_dunno | 0% | 99% | 1% | 100% | 0% | 1824 | 19.5 | 7857 | 19.5 | 11102 | 19.5 | 7988 | 19.5 | 7989 | 19.5 | 875 | 2.5 | | | | | | | | | | |
| veg_pepper_bell_fat_none | 0% | 100% | 0% | 100% | 0% | 1824 | 20 | 7857 | 20 | 11102 | 20 | 7988 | 20 | 7989 | 20 | | | | | | | | | | | | |
| veg_pepper_bell_fat_other | 0% | 99% | 1% | 1% | 99% | 1824 | 19.5 | 7857 | 19.5 | 11102 | 19.5 | 7988 | 19.5 | 7989 | 19.5 | 856 | 1.25 | 855 | 1.25 | | | | | | | | |
| veg_pepper_bell_lard | 0% | 99% | 1% | 1% | 99% | 1824 | 19.5 | 7857 | 19.5 | 11102 | 19.5 | 7988 | 19.5 | 7989 | 19.5 | 858 | 2.5 | | | | | | | | | | |
| veg_pepper_bell_marg_hard | 0% | 99% | 1% | 99% | 1% | 1824 | 19.5 | 7857 | 19.5 | 11102 | 19.5 | 7988 | 19.5 | 7989 | 19.5 | 860 | 2.5 | | | | | | | | | | |
| veg_pepper_bell_marg_poly_chol | 0% | 99% | 1% | 90% | 10% | 1824 | 19.5 | 7857 | 19.5 | 11102 | 19.5 | 7988 | 19.5 | 7989 | 19.5 | 2849 | 0.84 | 3848 | 0.83 | 3243 | 0.83 | | | | | | |
| veg_pepper_bell_marg_poly_dunno | 0% | 99% | 1% | 90% | 10% | 1824 | 19.5 | 7857 | 19.5 | 11102 | 19.5 | 7988 | 19.5 | 7989 | 19.5 | 10049 | 1.25 | 10043 | 0.625 | 10044 | 0.63 | | | | | | |
| veg_pepper_bell_marg_poly_fat | 0% | 99% | 1% | 90% | 10% | 1824 | 19.5 | 7857 | 19.5 | 11102 | 19.5 | 7988 | 19.5 | 7989 | 19.5 | 10043 | 1.25 | 10044 | 1.25 | | | | | | | | |
| veg_pepper_bell_marg_poly_lowfat | 0% | 99% | 1% | 90% | 10% | 1824 | 19.5 | 7857 | 19.5 | 11102 | 19.5 | 7988 | 19.5 | 7989 | 19.5 | 10049 | 2.5 | | | | | | | | | | |
| veg_pepper_bell_marg_poly_vlowfat | 0% | 99% | 1% | 90% | 10% | 1824 | 19.5 | 7857 | 19.5 | 11102 | 19.5 | 7988 | 19.5 | 7989 | 19.5 | 10049 | 2.5 | | | | | | | | | | |
| veg_pepper_bell_marg_soya_chol | 0% | 99% | 1% | 99% | 1% | 1824 | 19.5 | 7857 | 19.5 | 11102 | 19.5 | 7988 | 19.5 | 7989 | 19.5 | 3848 | 0.42 | 3243 | 0.41 | 2849 | 0.42 | 10786 | 1.25 | | | | |
| veg_pepper_bell_marg_soya_dunno | 0% | 99% | 1% | 100% | 0% | 1824 | 19.5 | 7857 | 19.5 | 11102 | 19.5 | 7988 | 19.5 | 7989 | 19.5 | 10980 | 1.25 | 10786 | 1.25 | | | | | | | | |
| veg_pepper_bell_marg_soya_fat | 0% | 99% | 1% | 100% | 0% | 1824 | 19.5 | 7857 | 19.5 | 11102 | 19.5 | 7988 | 19.5 | 7989 | 19.5 | 10786 | 2.5 | | | | | | | | | | |
| veg_pepper_bell_marg_soya_lowfat | 0% | 99% | 1% | 100% | 0% | 1824 | 19.5 | 7857 | 19.5 | 11102 | 19.5 | 7988 | 19.5 | 7989 | 19.5 | 10980 | 2.5 | | | | | | | | | | |
| veg_pepper_bell_marg_soya_vlowfat | 0% | 99% | 1% | 100% | 0% | 1824 | 19.5 | 7857 | 19.5 | 11102 | 19.5 | 7988 | 19.5 | 7989 | 19.5 | 10980 | 2.5 | | | | | | | | | | |
| veg_pepper_bell_oil_olive | 0% | 100% | 0% | 100% | 0% | 1824 | 19.5 | 7857 | 19.5 | 11102 | 19.5 | 7988 | 19.5 | 7989 | 19.5 | 874 | 2.5 | | | | | | | | | | |
| veg_pepper_bell_oil_other | 0% | 100% | 0% | 100% | 0% | 1824 | 19.5 | 7857 | 19.5 | 11102 | 19.5 | 7988 | 19.5 | 7989 | 19.5 | 870 | 0.75 | 870 | 0.5 | 10963 | 0.63 | 871 | 0.31 | 5000 | 0.313 | | |
| veg_pepper_bell_oil_rapeseed | 0% | 100% | 0% | 100% | 0% | 1824 | 19.5 | 7857 | 19.5 | 11102 | 19.5 | 7988 | 19.5 | 7989 | 19.5 | 7990 | 2.5 | | | | | | | | | | |
| veg_pepper_bell_oil_sunflower | 0% | 100% | 0% | 100% | 0% | 1824 | 19.5 | 7857 | 19.5 | 11102 | 19.5 | 7988 | 19.5 | 7989 | 19.5 | 873 | 2.5 | | | | | | | | | | |
| veg_pepper_bell_oil_veg | 0% | 100% | 0% | 100% | 0% | 1824 | 19.5 | 7857 | 19.5 | 11102 | 19.5 | 7988 | 19.5 | 7989 | 19.5 | 872 | 1.25 | 871 | 1.25 | | | | | | | | |
| veg_pepper_bell_spread_dairy_chol | 0% | 99% | 1% | 99% | 1% | 1824 | 19.5 | 7857 | 19.5 | 11102 | 19.5 | 7988 | 19.5 | 7989 | 19.5 | 3848 | 2.5 | | | | | | | | | | |
| veg_pepper_bell_spread_dairy_dunno | 0% | 99% | 1% | 99% | 1% | 1824 | 19.5 | 7857 | 19.5 | 11102 | 19.5 | 7988 | 19.5 | 7989 | 19.5 | 10047 | 1.25 | 7775 | 1.25 | | | | | | | | |
| veg_pepper_bell_spread_dairy_fat | 0% | 99% | 1% | 99% | 1% | 1824 | 19.5 | 7857 | 19.5 | 11102 | 19.5 | 7988 | 19.5 | 7989 | 19.5 | 7775 | 2.5 | | | | | | | | | | |
| veg_pepper_bell_spread_dairy_lowfat | 0% | 99% | 1% | 99% | 1% | 1824 | 19.5 | 7857 | 19.5 | 11102 | 19.5 | 7988 | 19.5 | 7989 | 19.5 | 10047 | 2.5 | | | | | | | | | | |
| veg_pepper_bell_spread_dairy_vlowfat | 0% | 99% | 1% | 99% | 1% | 1824 | 19.5 | 7857 | 19.5 | 11102 | 19.5 | 7988 | 19.5 | 7989 | 19.5 | 10047 | 2.5 | | | | | | | | | | |
| veg_pepper_bell_spread_dunno_chol | 0% | 99% | 1% | 99% | 1% | 1824 | 19.5 | 7857 | 19.5 | 11102 | 19.5 | 7988 | 19.5 | 7989 | 19.5 | 3848 | 0.84 | 3243 | 0.83 | 2849 | 0.83 | | | | | | |
| veg_pepper_bell_spread_dunno_dunno | 0% | 99% | 1% | 99% | 1% | 1824 | 19.5 | 7857 | 19.5 | 11102 | 19.5 | 7988 | 19.5 | 7989 | 19.5 | 10047 | 0.63 | 10049 | 0.625 | 10043 | 0.63 | 7775 | 0.63 | | | | |
| veg_pepper_bell_spread_dunno_fat | 0% | 99% | 1% | 99% | 1% | 1824 | 19.5 | 7857 | 19.5 | 11102 | 19.5 | 7988 | 19.5 | 7989 | 19.5 | 10043 | 1.25 | 7775 | 1.25 | | | | | | | | |
| veg_pepper_bell_spread_dunno_lowfat | 0% | 99% | 1% | 99% | 1% | 1824 | 19.5 | 7857 | 19.5 | 11102 | 19.5 | 7988 | 19.5 | 7989 | 19.5 | 10047 | 1.25 | 10049 | 1.25 | | | | | | | | |
| veg_pepper_bell_spread_dunno_vlowfat | 0% | 99% | 1% | 99% | 1% | 1824 | 19.5 | 7857 | 19.5 | 11102 | 19.5 | 7988 | 19.5 | 7989 | 19.5 | 10047 | 1.25 | 10049 | 1.25 | | | | | | | | |
| veg_pepper_bell_spread_olive_chol | 0% | 99% | 1% | 99% | 1% | 1824 | 19.5 | 7857 | 19.5 | 11102 | 19.5 | 7988 | 19.5 | 7989 | 19.5 | 3364 | 1.25 | 10053 | 1.25 | | | | | | | | |
| veg_pepper_bell_spread_olive_dunno | 0% | 99% | 1% | 99% | 1% | 1824 | 19.5 | 7857 | 19.5 | 11102 | 19.5 | 7988 | 19.5 | 7989 | 19.5 | 10048 | 1.25 | 10042 | 0.625 | 10131 | 0.63 | | | | | | |
| veg_pepper_bell_spread_olive_fat | 0% | 99% | 1% | 99% | 1% | 1824 | 19.5 | 7857 | 19.5 | 11102 | 19.5 | 7988 | 19.5 | 7989 | 19.5 | 10042 | 1.88 | 10131 | 0.625 | | | | | | | | |
| veg_pepper_bell_spread_olive_lowfat | 0% | 99% | 1% | 99% | 1% | 1824 | 19.5 | 7857 | 19.5 | 11102 | 19.5 | 7988 | 19.5 | 7989 | 19.5 | 10048 | 2.5 | | | | | | | | | | |
| veg_pepper_bell_spread_olive_vlowfat | 0% | 99% | 1% | 99% | 1% | 1824 | 19.5 | 7857 | 19.5 | 11102 | 19.5 | 7988 | 19.5 | 7989 | 19.5 | 10048 | 2.5 | | | | | | | | | | |
| veg_pulses | 0% | 100% | 0% | 100% | 0% | 1758 | 25 | 10788 | 12.5 | 10790 | 25 | 1669 | 25 | 10787 | 12.5 | | | | | | | | | | | | |
| veg_saladmayo | 59% | 20% | 80% | 50% | 50% | 8282 | 33.4 | 8079 | 33.3 | 8113 | 33.3 | | | | | | | | | | | | | | | | |
| veg_sidesalad | 0% | 100% | 0% | 100% | 0% | 1762 | 25 | 11103 | 50 | 1740 | 25 | | | | | | | | | | | | | | | | |
| veg_spinach | 0% | 100% | 0% | 100% | 0% | 1915 | 45 | 1916 | 45 | 11101 | 5 | 11100 | 5 | | | | | | | | | | | | | | |
| veg_sprouts | 0% | 100% | 0% | 100% | 0% | 1696 | 80 | 1698 | 20 | | | | | | | | | | | | | | | | | | |
| veg_sweetcorn | 0% | 100% | 0% | 100% | 0% | 1922 | 50 | 10794 | 25 | 10795 | 25 | | | | | | | | | | | | | | | | |
| veg_sweetpot | 0% | 100% | 0% | 100% | 0% | 10553 | 50 | 1930 | 50 | | | | | | | | | | | | | | | | | | |
| veg_tomato_fresh | 0% | 100% | 0% | 100% | 0% | 11103 | 50 | 11104 | 50 | | | | | | | | | | | | | | | | | | |
| veg_tomato_tinned | 0% | 100% | 0% | 100% | 0% | 1939 | 50 | 1935 | 25 | 1932 | 25 | | | | | | | | | | | | | | | | |
| veg_turnip | 0% | 100% | 0% | 100% | 0% | 1941 | 50 | 1921 | 50 | | | | | | | | | | | | | | | | | | |
| veg_watercress | 0% | 100% | 0% | 100% | 0% | 1947 | 100 | | | | | | | | | | | | | | | | | | | | |
| vegalt_burger | 0% | 100% | 0% | 100% | 0% | 9279 | 50 | 4203 | 50 | | | | | | | | | | | | | | | | | | |
| vegalt_other | 0% | 100% | 0% | 100% | 0% | 8587 | 75 | 8102 | 25 | | | | | | | | | | | | | | | | | | |
| vegalt_quorn | 0% | 100% | 0% | 100% | 0% | 7189 | 33.4 | 5677 | 33.3 | 8108 | 33.3 | | | | | | | | | | | | | | | | |
| vegalt_tofu | 0% | 100% | 0% | 100% | 0% | 1686 | 80 | 1371 | 10 | 5106 | 10 | | | | | | | | | | | | | | | | |
| yeast_extract | 0% | 100% | 0% | 100% | 0% | 2431 | 100 | | | | | | | | | | | | | | | | | | | | |
| yogurt_fullfat | 18% | 0% | 100% | 0% | 100% | 702 | 40 | 701 | 40 | 7741 | 20 | | | | | | | | | | | | | | | | |
| yogurt_lowfat | 28% | 0% | 100% | 0% | 100% | 705 | 25 | 704 | 50 | 2701 | 25 | | | | | | | | | | | | | | | | |
| yorkshirepud | 0% | 30% | 70% | 10% | 90% | 10711 | 25 | 10709 | 25 | 10712 | 50 | | | | | | | | | | | | | | | | |
