## Supplementary material for "Description of the updated nutrition calculation of the Oxford WebQ questionnaire and comparison with the previous version among 207,144 participants in UK Biobank": Sup table 3

**Supplementary table 3**. Nutrient calculation in the previous version (McCance and Widdowson).

| | Food codes from McCance and Widdowson and the % used from each food code | | | | | | | | | |
|---|---|---|---|---|---|---|---|---|---|---|
| **Item** | **Code1** | **% from code 1** | **Code2** | **% from code 2** | **Code3** | **% from code 3** | **Code4** | **% from code 4** | **Code5** | **% from code 5** |
| add_salt | 17367 | 100 | | | | | | | | |
| alcohol_beercider | 17211 | 35 | 17215 | 35 | 17222 | 15 | 17224 | 15 | | |
| alcohol_other | 17242 | 33.4 | 17245 | 33.3 | 17244 | 33.3 | | | | |
| alcohol_spirits | 17246 | 50 | 17247 | 50 | | | | | | |
| alcohol_wine_fort | 17235 | 16.7 | 17236 | 16.7 | 17237 | 16.6 | 17234 | 50 | | |
| alcohol_wine_red_large | 17228 | 100 | | | | | | | | |
| alcohol_wine_red_med | 17228 | 100 | | | | | | | | |
| alcohol_wine_red_small | 17228 | 100 | | | | | | | | |
| alcohol_wine_rose_large | 17229 | 100 | | | | | | | | |
| alcohol_wine_rose_med | 17229 | 100 | | | | | | | | |
| alcohol_wine_rose_small | 17229 | 100 | | | | | | | | |
| alcohol_wine_white_large | 17230 | 25 | 17231 | 25 | 17232 | 25 | 17233 | 25 | | |
| alcohol_wine_white_med | 17230 | 25 | 17231 | 25 | 17232 | 25 | 17233 | 25 | | |
| alcohol_wine_white_small | 17230 | 25 | 17231 | 25 | 17232 | 25 | 17233 | 25 | | |
| biscuit_choc | 11508 | 50 | 11512 | 50 | | | | | | |
| biscuit_choccov | 11507 | 50 | 11506 | 50 | | | | | | |
| biscuit_sweet | 11513 | 33.4 | 11523 | 33.3 | 11514 | 33.3 | | | | |
| bread_baguette_mixed | 11461 | 33.4 | 00033 | 33.3 | 11472 | 33.3 | | | | |
| bread_baguette_other | 00040 | 50 | 00046 | 50 | | | | | | |
| bread_baguette_seeded | 14844 | 50 | 14845 | 50 | | | | | | |
| bread_baguette_spread_butter_dunno_med | 17486 | 100 | | | | | | | | |
| bread_baguette_spread_butter_dunno_thick | 17486 | 100 | | | | | | | | |
| bread_baguette_spread_butter_dunno_thin | 17486 | 100 | | | | | | | | |
| bread_baguette_spread_butter_fat_med | 17485 | 100 | | | | | | | | |
| bread_baguette_spread_butter_fat_thick | 17485 | 100 | | | | | | | | |
| bread_baguette_spread_butter_fat_thin | 17485 | 100 | | | | | | | | |
| bread_baguette_spread_butter_lowfat_med | 17485 | 50 | 17017 | 50 | | | | | | |
| bread_baguette_spread_butter_lowfat_thick | 17485 | 50 | 17017 | 50 | | | | | | |
| bread_baguette_spread_butter_lowfat_thin | 17485 | 50 | 17017 | 50 | | | | | | |
| bread_baguette_spread_butter_spread_fat_med | 17486 | 100 | | | | | | | | |
| bread_baguette_spread_butter_spread_fat_thick | 17486 | 100 | | | | | | | | |
| bread_baguette_spread_butter_spread_fat_thin | 17486 | 100 | | | | | | | | |
| bread_baguette_spread_butter_spread_lowfat_med | 17486 | 50 | 17017 | 50 | | | | | | |
| bread_baguette_spread_butter_spread_lowfat_thick | 17486 | 50 | 17017 | 50 | | | | | | |
| bread_baguette_spread_butter_spread_lowfat_thin | 17486 | 50 | 17017 | 50 | | | | | | |
| bread_baguette_spread_dairy_chol_med | 17017 | 100 | | | | | | | | |
| bread_baguette_spread_dairy_chol_thick | 17017 | 100 | | | | | | | | |
| bread_baguette_spread_dairy_chol_thin | 17017 | 100 | | | | | | | | |
| bread_baguette_spread_dairy_dunno_med | 17017 | 30 | 12258 | 70 | | | | | | |
| bread_baguette_spread_dairy_dunno_thick | 17017 | 30 | 12258 | 70 | | | | | | |
| bread_baguette_spread_dairy_dunno_thin | 17017 | 30 | 12258 | 70 | | | | | | |
| bread_baguette_spread_dairy_fat_med | 12258 | 100 | | | | | | | | |
| bread_baguette_spread_dairy_fat_thick | 12258 | 100 | | | | | | | | |
| bread_baguette_spread_dairy_fat_thin | 12258 | 100 | | | | | | | | |
| bread_baguette_spread_dairy_lowfat_med | 17017 | 100 | | | | | | | | |
| bread_baguette_spread_dairy_lowfat_thick | 17017 | 100 | | | | | | | | |
| bread_baguette_spread_dairy_lowfat_thin | 17017 | 100 | | | | | | | | |
| bread_baguette_spread_dairy_vlowfat_med | 17028 | 100 | | | | | | | | |
| bread_baguette_spread_dairy_vlowfat_thick | 17028 | 100 | | | | | | | | |
| bread_baguette_spread_dairy_vlowfat_thin | 17028 | 100 | | | | | | | | |
| bread_baguette_spread_dunno_chol_med | 17552 | 50 | 17027 | 50 | | | | | | |
| bread_baguette_spread_dunno_chol_thick | 17552 | 50 | 17027 | 50 | | | | | | |
| bread_baguette_spread_dunno_chol_thin | 17552 | 50 | 17027 | 50 | | | | | | |
| bread_baguette_spread_dunno_dunno_med | 17552 | 25 | 17027 | 25 | 17025 | 25 | 17024 | 25 | | |
| bread_baguette_spread_dunno_dunno_thick | 17552 | 25 | 17027 | 25 | 17025 | 25 | 17024 | 25 | | |
| bread_baguette_spread_dunno_dunno_thin | 17552 | 25 | 17027 | 25 | 17025 | 25 | 17024 | 25 | | |
| bread_baguette_spread_dunno_fat_med | 12258 | 33.3 | 17025 | 33.3 | 17024 | 33.4 | | | | |
| bread_baguette_spread_dunno_fat_thick | 12258 | 33.3 | 17025 | 33.3 | 17024 | 33.4 | | | | |
| bread_baguette_spread_dunno_fat_thin | 12258 | 33.3 | 17025 | 33.3 | 17024 | 33.4 | | | | |
| bread_baguette_spread_dunno_lowfat_med | 17017 | 33.3 | 17552 | 33.3 | 17027 | 33.4 | | | | |
| bread_baguette_spread_dunno_lowfat_thick | 17017 | 33.3 | 17552 | 33.3 | 17027 | 33.4 | | | | |
| bread_baguette_spread_dunno_lowfat_thin | 17017 | 33.3 | 17552 | 33.3 | 17027 | 33.4 | | | | |
| bread_baguette_spread_dunno_vlowfat_med | 17028 | 50 | 17029 | 50 | | | | | | |
| bread_baguette_spread_dunno_vlowfat_thick | 17028 | 50 | 17029 | 50 | | | | | | |
| bread_baguette_spread_dunno_vlowfat_thin | 17028 | 50 | 17029 | 50 | | | | | | |
| bread_baguette_spread_hardmarg_med | 17018 | 50 | 17539 | 50 | | | | | | |
| bread_baguette_spread_hardmarg_thick | 17018 | 50 | 17539 | 50 | | | | | | |
| bread_baguette_spread_hardmarg_thin | 17018 | 50 | 17539 | 50 | | | | | | |
| bread_baguette_spread_olive_chol_med | 17552 | 50 | 17025 | 50 | | | | | | |
| bread_baguette_spread_olive_chol_thick | 17552 | 50 | 17025 | 50 | | | | | | |
| bread_baguette_spread_olive_chol_thin | 17552 | 50 | 17025 | 50 | | | | | | |
| bread_baguette_spread_olive_dunno_med | 17552 | 50 | 17025 | 50 | | | | | | |
| bread_baguette_spread_olive_dunno_thick | 17552 | 50 | 17025 | 50 | | | | | | |
| bread_baguette_spread_olive_dunno_thin | 17552 | 30 | 17025 | 70 | | | | | | |
| bread_baguette_spread_olive_fat_med | 17025 | 100 | | | | | | | | |
| bread_baguette_spread_olive_fat_thick | 17025 | 100 | | | | | | | | |
| bread_baguette_spread_olive_fat_thin | 17025 | 100 | | | | | | | | |
| bread_baguette_spread_olive_lowfat_med | 17552 | 100 | | | | | | | | |
| bread_baguette_spread_olive_lowfat_thick | 17552 | 100 | | | | | | | | |
| bread_baguette_spread_olive_lowfat_thin | 17552 | 100 | | | | | | | | |
| bread_baguette_spread_olive_vlowfat_med | 17028 | 100 | | | | | | | | |
| bread_baguette_spread_olive_vlowfat_thick | 17028 | 100 | | | | | | | | |
| bread_baguette_spread_olive_vlowfat_thin | 17028 | 100 | | | | | | | | |
| bread_baguette_spread_other_med | 17007 | 50 | 17487 | 50 | | | | | | |
| bread_baguette_spread_other_thick | 17007 | 50 | 17487 | 50 | | | | | | |
| bread_baguette_spread_other_thin | 17007 | 50 | 17487 | 50 | | | | | | |
| bread_baguette_spread_polymarg_chol_med | 17027 | 100 | | | | | | | | |
| bread_baguette_spread_polymarg_chol_thick | 17027 | 100 | | | | | | | | |
| bread_baguette_spread_polymarg_chol_thin | 17027 | 100 | | | | | | | | |
| bread_baguette_spread_polymarg_dunno_med | 17027 | 30 | 17024 | 70 | | | | | | |
| bread_baguette_spread_polymarg_dunno_thick | 17027 | 30 | 17024 | 70 | | | | | | |
| bread_baguette_spread_polymarg_dunno_thin | 17027 | 30 | 17024 | 70 | | | | | | |
| bread_baguette_spread_polymarg_fat_med | 17024 | 100 | | | | | | | | |
| bread_baguette_spread_polymarg_fat_thick | 17024 | 100 | | | | | | | | |
| bread_baguette_spread_polymarg_fat_thin | 17024 | 100 | | | | | | | | |
| bread_baguette_spread_polymarg_lowfat_med | 17027 | 100 | | | | | | | | |
| bread_baguette_spread_polymarg_lowfat_thick | 17027 | 100 | | | | | | | | |
| bread_baguette_spread_polymarg_lowfat_thin | 17027 | 100 | | | | | | | | |
| bread_baguette_spread_polymarg_vlowfat_med | 17029 | 100 | | | | | | | | |
| bread_baguette_spread_polymarg_vlowfat_thick | 17029 | 100 | | | | | | | | |
| bread_baguette_spread_polymarg_vlowfat_thin | 17029 | 100 | | | | | | | | |
| bread_baguette_spread_soya_chol_med | 17027 | 100 | | | | | | | | |
| bread_baguette_spread_soya_chol_thick | 17027 | 100 | | | | | | | | |
| bread_baguette_spread_soya_chol_thin | 17027 | 100 | | | | | | | | |
| bread_baguette_spread_soya_dunno_med | 17027 | 30 | 17024 | 70 | | | | | | |
| bread_baguette_spread_soya_dunno_thick | 17027 | 30 | 17024 | 70 | | | | | | |
| bread_baguette_spread_soya_dunno_thin | 17027 | 30 | 17024 | 70 | | | | | | |
| bread_baguette_spread_soya_fat_med | 17024 | 100 | | | | | | | | |
| bread_baguette_spread_soya_fat_thick | 17024 | 100 | | | | | | | | |

**Supplementary table 3**. Nutrient calculation in the previous version (McCance and Widdowson).

| | Food codes from McCance and Widdowson and the % used from each food code | | | | | | | | | |
|---|---|---|---|---|---|---|---|---|---|---|
| **Item** | **Code1** | **% from code 1** | **Code2** | **% from code 2** | **Code3** | **% from code 3** | **Code4** | **% from code 4** | **Code5** | **% from code 5** |
| bread_baguette_spread_soya_fat_thin | 17024 | 100 | | | | | | | | |
| bread_baguette_spread_soya_lowfat_med | 17027 | 100 | | | | | | | | |
| bread_baguette_spread_soya_lowfat_thick | 17027 | 100 | | | | | | | | |
| bread_baguette_spread_soya_lowfat_thin | 17027 | 100 | | | | | | | | |
| bread_baguette_spread_soya_vlowfat_med | 17029 | 100 | | | | | | | | |
| bread_baguette_spread_soya_vlowfat_thick | 17029 | 100 | | | | | | | | |
| bread_baguette_spread_soya_vlowfat_thin | 17029 | 100 | | | | | | | | |
| bread_baguette_unanswered | 00048 | 33.4 | 00056 | 33.3 | 00033 | 33.3 | | | | |
| bread_baguette_white | 11471 | 50 | 11609 | 50 | | | | | | |
| bread_baguette_wholemeal | 00056 | 100 | | | | | | | | |
| bread_crisp | 11511 | 33.4 | 11510 | 33.3 | 11572 | 33.3 | | | | |
| bread_crisp_spread_butter_dunno_med | 17486 | 100 | | | | | | | | |
| bread_crisp_spread_butter_dunno_thick | 17486 | 100 | | | | | | | | |
| bread_crisp_spread_butter_dunno_thin | 17486 | 100 | | | | | | | | |
| bread_crisp_spread_butter_fat_med | 17485 | 100 | | | | | | | | |
| bread_crisp_spread_butter_fat_thick | 17485 | 100 | | | | | | | | |
| bread_crisp_spread_butter_fat_thin | 17485 | 100 | | | | | | | | |
| bread_crisp_spread_butter_lowfat_med | 17485 | 50 | 17017 | 50 | | | | | | |
| bread_crisp_spread_butter_lowfat_thick | 17485 | 50 | 17017 | 50 | | | | | | |
| bread_crisp_spread_butter_lowfat_thin | 17485 | 50 | 17017 | 50 | | | | | | |
| bread_crisp_spread_butter_spread_fat_med | 17486 | 100 | | | | | | | | |
| bread_crisp_spread_butter_spread_fat_thick | 17486 | 100 | | | | | | | | |
| bread_crisp_spread_butter_spread_fat_thin | 17486 | 100 | | | | | | | | |
| bread_crisp_spread_butter_spread_lowfat_med | 17486 | 50 | 17017 | 50 | | | | | | |
| bread_crisp_spread_butter_spread_lowfat_thick | 17486 | 50 | 17017 | 50 | | | | | | |
| bread_crisp_spread_butter_spread_lowfat_thin | 17486 | 50 | 17017 | 50 | | | | | | |
| bread_crisp_spread_dairy_chol_med | 17017 | 100 | | | | | | | | |
| bread_crisp_spread_dairy_chol_thick | 17017 | 100 | | | | | | | | |
| bread_crisp_spread_dairy_chol_thin | 17017 | 100 | | | | | | | | |
| bread_crisp_spread_dairy_dunno_med | 17017 | 30 | 12258 | 70 | | | | | | |
| bread_crisp_spread_dairy_dunno_thick | 17017 | 30 | 12258 | 70 | | | | | | |
| bread_crisp_spread_dairy_dunno_thin | 17017 | 30 | 12258 | 70 | | | | | | |
| bread_crisp_spread_dairy_fat_med | 12258 | 100 | | | | | | | | |
| bread_crisp_spread_dairy_fat_thick | 12258 | 100 | | | | | | | | |
| bread_crisp_spread_dairy_fat_thin | 12258 | 100 | | | | | | | | |
| bread_crisp_spread_dairy_lowfat_med | 17017 | 100 | | | | | | | | |
| bread_crisp_spread_dairy_lowfat_thick | 17017 | 100 | | | | | | | | |
| bread_crisp_spread_dairy_lowfat_thin | 17017 | 100 | | | | | | | | |
| bread_crisp_spread_dairy_vlowfat_med | 17028 | 100 | | | | | | | | |
| bread_crisp_spread_dairy_vlowfat_thick | 17028 | 100 | | | | | | | | |
| bread_crisp_spread_dairy_vlowfat_thin | 17028 | 100 | | | | | | | | |
| bread_crisp_spread_dunno_chol_med | 17552 | 50 | 17027 | 50 | | | | | | |
| bread_crisp_spread_dunno_chol_thick | 17552 | 50 | 17027 | 50 | | | | | | |
| bread_crisp_spread_dunno_chol_thin | 17552 | 50 | 17027 | 50 | | | | | | |
| bread_crisp_spread_dunno_dunno_med | 17552 | 25 | 17027 | 25 | 17025 | 25 | 17024 | 25 | | |
| bread_crisp_spread_dunno_dunno_thick | 17552 | 25 | 17027 | 25 | 17025 | 25 | 17024 | 25 | | |
| bread_crisp_spread_dunno_dunno_thin | 17552 | 25 | 17027 | 25 | 17025 | 25 | 17024 | 25 | | |
| bread_crisp_spread_dunno_fat_med | 12258 | 33.3 | 17025 | 33.3 | 17024 | 33.4 | | | | |
| bread_crisp_spread_dunno_fat_thick | 12258 | 33.3 | 17025 | 33.3 | 17024 | 33.4 | | | | |
| bread_crisp_spread_dunno_fat_thin | 12258 | 33.3 | 17025 | 33.3 | 17024 | 33.4 | | | | |
| bread_crisp_spread_dunno_lowfat_med | 17017 | 33.3 | 17552 | 33.3 | 17027 | 33.4 | | | | |
| bread_crisp_spread_dunno_lowfat_thick | 17017 | 33.3 | 17552 | 33.3 | 17027 | 33.4 | | | | |
| bread_crisp_spread_dunno_lowfat_thin | 17017 | 33.3 | 17552 | 33.3 | 17027 | 33.4 | | | | |
| bread_crisp_spread_dunno_vlowfat_med | 17028 | 50 | 17029 | 50 | | | | | | |
| bread_crisp_spread_dunno_vlowfat_thick | 17028 | 50 | 17029 | 50 | | | | | | |
| bread_crisp_spread_dunno_vlowfat_thin | 17028 | 50 | 17029 | 50 | | | | | | |
| bread_crisp_spread_hardmarg_med | 17018 | 50 | 17539 | 50 | | | | | | |
| bread_crisp_spread_hardmarg_thick | 17018 | 50 | 17539 | 50 | | | | | | |
| bread_crisp_spread_hardmarg_thin | 17018 | 50 | 17539 | 50 | | | | | | |
| bread_crisp_spread_olive_chol_med | 17552 | 50 | 17025 | 50 | | | | | | |
| bread_crisp_spread_olive_chol_thick | 17552 | 50 | 17025 | 50 | | | | | | |
| bread_crisp_spread_olive_chol_thin | 17552 | 50 | 17025 | 50 | | | | | | |
| bread_crisp_spread_olive_dunno_med | 17552 | 50 | 17025 | 50 | | | | | | |
| bread_crisp_spread_olive_dunno_thick | 17552 | 50 | 17025 | 50 | | | | | | |
| bread_crisp_spread_olive_dunno_thin | 17552 | 30 | 17025 | 70 | | | | | | |
| bread_crisp_spread_olive_fat_med | 17025 | 100 | | | | | | | | |
| bread_crisp_spread_olive_fat_thick | 17025 | 100 | | | | | | | | |
| bread_crisp_spread_olive_fat_thin | 17025 | 100 | | | | | | | | |
| bread_crisp_spread_olive_lowfat_med | 17552 | 100 | | | | | | | | |
| bread_crisp_spread_olive_lowfat_thick | 17552 | 100 | | | | | | | | |
| bread_crisp_spread_olive_lowfat_thin | 17552 | 100 | | | | | | | | |
| bread_crisp_spread_olive_vlowfat_med | 17028 | 100 | | | | | | | | |
| bread_crisp_spread_olive_vlowfat_thick | 17028 | 100 | | | | | | | | |
| bread_crisp_spread_olive_vlowfat_thin | 17028 | 100 | | | | | | | | |
| bread_crisp_spread_other_med | 17007 | 50 | 17487 | 50 | | | | | | |
| bread_crisp_spread_other_thick | 17007 | 50 | 17487 | 50 | | | | | | |
| bread_crisp_spread_other_thin | 17007 | 50 | 17487 | 50 | | | | | | |
| bread_crisp_spread_polymarg_chol_med | 17027 | 100 | | | | | | | | |
| bread_crisp_spread_polymarg_chol_thick | 17027 | 100 | | | | | | | | |
| bread_crisp_spread_polymarg_chol_thin | 17027 | 100 | | | | | | | | |
| bread_crisp_spread_polymarg_dunno_med | 17027 | 30 | 17024 | 70 | | | | | | |
| bread_crisp_spread_polymarg_dunno_thick | 17027 | 30 | 17024 | 70 | | | | | | |
| bread_crisp_spread_polymarg_dunno_thin | 17027 | 30 | 17024 | 70 | | | | | | |
| bread_crisp_spread_polymarg_fat_med | 17024 | 100 | | | | | | | | |
| bread_crisp_spread_polymarg_fat_thick | 17024 | 100 | | | | | | | | |
| bread_crisp_spread_polymarg_fat_thin | 17024 | 100 | | | | | | | | |
| bread_crisp_spread_polymarg_lowfat_med | 17027 | 100 | | | | | | | | |
| bread_crisp_spread_polymarg_lowfat_thick | 17027 | 100 | | | | | | | | |
| bread_crisp_spread_polymarg_lowfat_thin | 17027 | 100 | | | | | | | | |
| bread_crisp_spread_polymarg_vlowfat_med | 17029 | 100 | | | | | | | | |
| bread_crisp_spread_polymarg_vlowfat_thick | 17029 | 100 | | | | | | | | |
| bread_crisp_spread_polymarg_vlowfat_thin | 17029 | 100 | | | | | | | | |
| bread_crisp_spread_soya_chol_med | 17027 | 100 | | | | | | | | |
| bread_crisp_spread_soya_chol_thick | 17027 | 100 | | | | | | | | |
| bread_crisp_spread_soya_chol_thin | 17027 | 100 | | | | | | | | |
| bread_crisp_spread_soya_dunno_med | 17027 | 30 | 17024 | 70 | | | | | | |
| bread_crisp_spread_soya_dunno_thick | 17027 | 30 | 17024 | 70 | | | | | | |
| bread_crisp_spread_soya_dunno_thin | 17027 | 30 | 17024 | 70 | | | | | | |
| bread_crisp_spread_soya_fat_med | 17024 | 100 | | | | | | | | |
| bread_crisp_spread_soya_fat_thick | 17024 | 100 | | | | | | | | |
| bread_crisp_spread_soya_fat_thin | 17024 | 100 | | | | | | | | |
| bread_crisp_spread_soya_lowfat_med | 17027 | 100 | | | | | | | | |
| bread_crisp_spread_soya_lowfat_thick | 17027 | 100 | | | | | | | | |
| bread_crisp_spread_soya_lowfat_thin | 17027 | 100 | | | | | | | | |
| bread_crisp_spread_soya_vlowfat_med | 17029 | 100 | | | | | | | | |
| bread_crisp_spread_soya_vlowfat_thick | 17029 | 100 | | | | | | | | |
| bread_crisp_spread_soya_vlowfat_thin | 17029 | 100 | | | | | | | | |
| bread_garlic | 11460 | 100 | | | | | | | | |
| bread_large_bap_mixed | 11461 | 33.4 | 00033 | 33.3 | 11472 | 33.3 | | | | |

**Supplementary table 3**. Nutrient calculation in the previous version (McCance and Widdowson).

| | Food codes from McCance and Widdowson and the % used from each food code | | | | | | | | | |
|---|---|---|---|---|---|---|---|---|---|---|
| **Item** | **Code1** | **% from code 1** | **Code2** | **% from code 2** | **Code3** | **% from code 3** | **Code4** | **% from code 4** | **Code5** | **% from code 5** |
| bread_large_bap_other | 00040 | 50 | 00046 | 50 | | | | | | |
| bread_large_bap_seeded | 14844 | 50 | 14845 | 50 | | | | | | |
| bread_large_bap_spread_butter_dunno_med | 17486 | 100 | | | | | | | | |
| bread_large_bap_spread_butter_dunno_thick | 17486 | 100 | | | | | | | | |
| bread_large_bap_spread_butter_dunno_thin | 17486 | 100 | | | | | | | | |
| bread_large_bap_spread_butter_fat_med | 17485 | 100 | | | | | | | | |
| bread_large_bap_spread_butter_fat_thick | 17485 | 100 | | | | | | | | |
| bread_large_bap_spread_butter_fat_thin | 17485 | 100 | | | | | | | | |
| bread_large_bap_spread_butter_lowfat_med | 17485 | 50 | 17017 | 50 | | | | | | |
| bread_large_bap_spread_butter_lowfat_thick | 17485 | 50 | 17017 | 50 | | | | | | |
| bread_large_bap_spread_butter_lowfat_thin | 17485 | 50 | 17017 | 50 | | | | | | |
| bread_large_bap_spread_butter_spread_fat_med | 17486 | 100 | | | | | | | | |
| bread_large_bap_spread_butter_spread_fat_thick | 17486 | 100 | | | | | | | | |
| bread_large_bap_spread_butter_spread_fat_thin | 17486 | 100 | | | | | | | | |
| bread_large_bap_spread_butter_spread_lowfat_med | 17486 | 50 | 17017 | 50 | | | | | | |
| bread_large_bap_spread_butter_spread_lowfat_thick | 17486 | 50 | 17017 | 50 | | | | | | |
| bread_large_bap_spread_butter_spread_lowfat_thin | 17486 | 50 | 17017 | 50 | | | | | | |
| bread_large_bap_spread_dairy_chol_med | 17017 | 100 | | | | | | | | |
| bread_large_bap_spread_dairy_chol_thick | 17017 | 100 | | | | | | | | |
| bread_large_bap_spread_dairy_chol_thin | 17017 | 100 | | | | | | | | |
| bread_large_bap_spread_dairy_dunno_med | 17017 | 30 | 12258 | 70 | | | | | | |
| bread_large_bap_spread_dairy_dunno_thick | 17017 | 30 | 12258 | 70 | | | | | | |
| bread_large_bap_spread_dairy_dunno_thin | 17017 | 30 | 12258 | 70 | | | | | | |
| bread_large_bap_spread_dairy_fat_med | 12258 | 100 | | | | | | | | |
| bread_large_bap_spread_dairy_fat_thick | 12258 | 100 | | | | | | | | |
| bread_large_bap_spread_dairy_fat_thin | 12258 | 100 | | | | | | | | |
| bread_large_bap_spread_dairy_lowfat_med | 17017 | 100 | | | | | | | | |
| bread_large_bap_spread_dairy_lowfat_thick | 17017 | 100 | | | | | | | | |
| bread_large_bap_spread_dairy_lowfat_thin | 17017 | 100 | | | | | | | | |
| bread_large_bap_spread_dairy_vlowfat_med | 17028 | 100 | | | | | | | | |
| bread_large_bap_spread_dairy_vlowfat_thick | 17028 | 100 | | | | | | | | |
| bread_large_bap_spread_dairy_vlowfat_thin | 17028 | 100 | | | | | | | | |
| bread_large_bap_spread_dunno_chol_med | 17552 | 50 | 17027 | 50 | | | | | | |
| bread_large_bap_spread_dunno_chol_thick | 17552 | 50 | 17027 | 50 | | | | | | |
| bread_large_bap_spread_dunno_chol_thin | 17552 | 50 | 17027 | 50 | | | | | | |
| bread_large_bap_spread_dunno_dunno_med | 17552 | 25 | 17027 | 25 | 17025 | 25 | 17024 | 25 | | |
| bread_large_bap_spread_dunno_dunno_thick | 17552 | 25 | 17027 | 25 | 17025 | 25 | 17024 | 25 | | |
| bread_large_bap_spread_dunno_dunno_thin | 17552 | 25 | 17027 | 25 | 17025 | 25 | 17024 | 25 | | |
| bread_large_bap_spread_dunno_fat_med | 12258 | 33.3 | 17025 | 33.3 | 17024 | 33.4 | | | | |
| bread_large_bap_spread_dunno_fat_thick | 12258 | 33.3 | 17025 | 33.3 | 17024 | 33.4 | | | | |
| bread_large_bap_spread_dunno_fat_thin | 12258 | 33.3 | 17025 | 33.3 | 17024 | 33.4 | | | | |
| bread_large_bap_spread_dunno_lowfat_med | 17017 | 33.3 | 17552 | 33.3 | 17027 | 33.4 | | | | |
| bread_large_bap_spread_dunno_lowfat_thick | 17017 | 33.3 | 17552 | 33.3 | 17027 | 33.4 | | | | |
| bread_large_bap_spread_dunno_lowfat_thin | 17017 | 33.3 | 17552 | 33.3 | 17027 | 33.4 | | | | |
| bread_large_bap_spread_dunno_vlowfat_med | 17028 | 50 | 17029 | 50 | | | | | | |
| bread_large_bap_spread_dunno_vlowfat_thick | 17028 | 50 | 17029 | 50 | | | | | | |
| bread_large_bap_spread_dunno_vlowfat_thin | 17028 | 50 | 17029 | 50 | | | | | | |
| bread_large_bap_spread_hardmarg_med | 17018 | 50 | 17539 | 50 | | | | | | |
| bread_large_bap_spread_hardmarg_thick | 17018 | 50 | 17539 | 50 | | | | | | |
| bread_large_bap_spread_hardmarg_thin | 17018 | 50 | 17539 | 50 | | | | | | |
| bread_large_bap_spread_olive_chol_med | 17552 | 50 | 17025 | 50 | | | | | | |
| bread_large_bap_spread_olive_chol_thick | 17552 | 50 | 17025 | 50 | | | | | | |
| bread_large_bap_spread_olive_chol_thin | 17552 | 50 | 17025 | 50 | | | | | | |
| bread_large_bap_spread_olive_dunno_med | 17552 | 50 | 17025 | 50 | | | | | | |
| bread_large_bap_spread_olive_dunno_thick | 17552 | 50 | 17025 | 50 | | | | | | |
| bread_large_bap_spread_olive_dunno_thin | 17552 | 30 | 17025 | 70 | | | | | | |
| bread_large_bap_spread_olive_fat_med | 17025 | 100 | | | | | | | | |
| bread_large_bap_spread_olive_fat_thick | 17025 | 100 | | | | | | | | |
| bread_large_bap_spread_olive_fat_thin | 17025 | 100 | | | | | | | | |
| bread_large_bap_spread_olive_lowfat_med | 17552 | 100 | | | | | | | | |
| bread_large_bap_spread_olive_lowfat_thick | 17552 | 100 | | | | | | | | |
| bread_large_bap_spread_olive_lowfat_thin | 17552 | 100 | | | | | | | | |
| bread_large_bap_spread_olive_vlowfat_med | 17028 | 100 | | | | | | | | |
| bread_large_bap_spread_olive_vlowfat_thick | 17028 | 100 | | | | | | | | |
| bread_large_bap_spread_olive_vlowfat_thin | 17028 | 100 | | | | | | | | |
| bread_large_bap_spread_other_med | 17007 | 50 | 17487 | 50 | | | | | | |
| bread_large_bap_spread_other_thick | 17007 | 50 | 17487 | 50 | | | | | | |
| bread_large_bap_spread_other_thin | 17007 | 50 | 17487 | 50 | | | | | | |
| bread_large_bap_spread_polymarg_chol_med | 17027 | 100 | | | | | | | | |
| bread_large_bap_spread_polymarg_chol_thick | 17027 | 100 | | | | | | | | |
| bread_large_bap_spread_polymarg_chol_thin | 17027 | 100 | | | | | | | | |
| bread_large_bap_spread_polymarg_dunno_med | 17027 | 30 | 17024 | 70 | | | | | | |
| bread_large_bap_spread_polymarg_dunno_thick | 17027 | 30 | 17024 | 70 | | | | | | |
| bread_large_bap_spread_polymarg_dunno_thin | 17027 | 30 | 17024 | 70 | | | | | | |
| bread_large_bap_spread_polymarg_fat_med | 17024 | 100 | | | | | | | | |
| bread_large_bap_spread_polymarg_fat_thick | 17024 | 100 | | | | | | | | |
| bread_large_bap_spread_polymarg_fat_thin | 17024 | 100 | | | | | | | | |
| bread_large_bap_spread_polymarg_lowfat_med | 17027 | 100 | | | | | | | | |
| bread_large_bap_spread_polymarg_lowfat_thick | 17027 | 100 | | | | | | | | |
| bread_large_bap_spread_polymarg_lowfat_thin | 17027 | 100 | | | | | | | | |
| bread_large_bap_spread_polymarg_vlowfat_med | 17029 | 100 | | | | | | | | |
| bread_large_bap_spread_polymarg_vlowfat_thick | 17029 | 100 | | | | | | | | |
| bread_large_bap_spread_polymarg_vlowfat_thin | 17029 | 100 | | | | | | | | |
| bread_large_bap_spread_soya_chol_med | 17027 | 100 | | | | | | | | |
| bread_large_bap_spread_soya_chol_thick | 17027 | 100 | | | | | | | | |
| bread_large_bap_spread_soya_chol_thin | 17027 | 100 | | | | | | | | |
| bread_large_bap_spread_soya_dunno_med | 17027 | 30 | 17024 | 70 | | | | | | |
| bread_large_bap_spread_soya_dunno_thick | 17027 | 30 | 17024 | 70 | | | | | | |
| bread_large_bap_spread_soya_dunno_thin | 17027 | 30 | 17024 | 70 | | | | | | |
| bread_large_bap_spread_soya_fat_med | 17024 | 100 | | | | | | | | |
| bread_large_bap_spread_soya_fat_thick | 17024 | 100 | | | | | | | | |
| bread_large_bap_spread_soya_fat_thin | 17024 | 100 | | | | | | | | |
| bread_large_bap_spread_soya_lowfat_med | 17027 | 100 | | | | | | | | |
| bread_large_bap_spread_soya_lowfat_thick | 17027 | 100 | | | | | | | | |
| bread_large_bap_spread_soya_lowfat_thin | 17027 | 100 | | | | | | | | |
| bread_large_bap_spread_soya_vlowfat_med | 17029 | 100 | | | | | | | | |
| bread_large_bap_spread_soya_vlowfat_thick | 17029 | 100 | | | | | | | | |
| bread_large_bap_spread_soya_vlowfat_thin | 17029 | 100 | | | | | | | | |
| bread_large_bap_unanswered | 00048 | 33.4 | 00056 | 33.3 | 00033 | 33.3 | | | | |
| bread_large_bap_white | 11465 | 50 | 00048 | 50 | | | | | | |
| bread_large_bap_wholemeal | 00056 | 100 | | | | | | | | |
| bread_naan | 11463 | 100 | | | | | | | | |
| bread_other | 11093 | 45 | 11535 | 45 | 17123 | 10 | | | | |
| bread_other_spread_butter_dunno_med | 17486 | 100 | | | | | | | | |
| bread_other_spread_butter_dunno_thick | 17486 | 100 | | | | | | | | |
| bread_other_spread_butter_dunno_thin | 17486 | 100 | | | | | | | | |
| bread_other_spread_butter_fat_med | 17485 | 100 | | | | | | | | |
| bread_other_spread_butter_fat_thick | 17485 | 100 | | | | | | | | |
| bread_other_spread_butter_fat_thin | 17485 | 100 | | | | | | | | |

**Supplementary table 3**. Nutrient calculation in the previous version (McCance and Widdowson).

| | Food codes from McCance and Widdowson and the % used from each food code | | | | | | | | | |
|---|---|---|---|---|---|---|---|---|---|---|
| **Item** | **Code1** | **% from code 1** | **Code2** | **% from code 2** | **Code3** | **% from code 3** | **Code4** | **% from code 4** | **Code5** | **% from code 5** |
| bread_other_spread_butter_lowfat_med | 17485 | 50 | 17017 | 50 | | | | | | |
| bread_other_spread_butter_lowfat_thick | 17485 | 50 | 17017 | 50 | | | | | | |
| bread_other_spread_butter_lowfat_thin | 17485 | 50 | 17017 | 50 | | | | | | |
| bread_other_spread_butter_spread_fat_med | 17486 | 100 | | | | | | | | |
| bread_other_spread_butter_spread_fat_thick | 17486 | 100 | | | | | | | | |
| bread_other_spread_butter_spread_fat_thin | 17486 | 100 | | | | | | | | |
| bread_other_spread_butter_spread_lowfat_med | 17486 | 50 | 17017 | 50 | | | | | | |
| bread_other_spread_butter_spread_lowfat_thick | 17486 | 50 | 17017 | 50 | | | | | | |
| bread_other_spread_butter_spread_lowfat_thin | 17486 | 50 | 17017 | 50 | | | | | | |
| bread_other_spread_dairy_chol_med | 17017 | 100 | | | | | | | | |
| bread_other_spread_dairy_chol_thick | 17017 | 100 | | | | | | | | |
| bread_other_spread_dairy_chol_thin | 17017 | 100 | | | | | | | | |
| bread_other_spread_dairy_dunno_med | 17017 | 30 | 12258 | 70 | | | | | | |
| bread_other_spread_dairy_dunno_thick | 17017 | 30 | 12258 | 70 | | | | | | |
| bread_other_spread_dairy_dunno_thin | 17017 | 30 | 12258 | 70 | | | | | | |
| bread_other_spread_dairy_fat_med | 12258 | 100 | | | | | | | | |
| bread_other_spread_dairy_fat_thick | 12258 | 100 | | | | | | | | |
| bread_other_spread_dairy_fat_thin | 12258 | 100 | | | | | | | | |
| bread_other_spread_dairy_lowfat_med | 17017 | 100 | | | | | | | | |
| bread_other_spread_dairy_lowfat_thick | 17017 | 100 | | | | | | | | |
| bread_other_spread_dairy_lowfat_thin | 17017 | 100 | | | | | | | | |
| bread_other_spread_dairy_vlowfat_med | 17028 | 100 | | | | | | | | |
| bread_other_spread_dairy_vlowfat_thick | 17028 | 100 | | | | | | | | |
| bread_other_spread_dairy_vlowfat_thin | 17028 | 100 | | | | | | | | |
| bread_other_spread_dunno_chol_med | 17552 | 50 | 17027 | 50 | | | | | | |
| bread_other_spread_dunno_chol_thick | 17552 | 50 | 17027 | 50 | | | | | | |
| bread_other_spread_dunno_chol_thin | 17552 | 50 | 17027 | 50 | | | | | | |
| bread_other_spread_dunno_dunno_med | 17552 | 25 | 17027 | 25 | 17025 | 25 | 17024 | 25 | | |
| bread_other_spread_dunno_dunno_thick | 17552 | 25 | 17027 | 25 | 17025 | 25 | 17024 | 25 | | |
| bread_other_spread_dunno_dunno_thin | 17552 | 25 | 17027 | 25 | 17025 | 25 | 17024 | 25 | | |
| bread_other_spread_dunno_fat_med | 12258 | 33.3 | 17025 | 33.3 | 17024 | 33.4 | | | | |
| bread_other_spread_dunno_fat_thick | 12258 | 33.3 | 17025 | 33.3 | 17024 | 33.4 | | | | |
| bread_other_spread_dunno_fat_thin | 12258 | 33.3 | 17025 | 33.3 | 17024 | 33.4 | | | | |
| bread_other_spread_dunno_lowfat_med | 17017 | 33.3 | 17552 | 33.3 | 17027 | 33.4 | | | | |
| bread_other_spread_dunno_lowfat_thick | 17017 | 33.3 | 17552 | 33.3 | 17027 | 33.4 | | | | |
| bread_other_spread_dunno_lowfat_thin | 17017 | 33.3 | 17552 | 33.3 | 17027 | 33.4 | | | | |
| bread_other_spread_dunno_vlowfat_med | 17028 | 50 | 17029 | 50 | | | | | | |
| bread_other_spread_dunno_vlowfat_thick | 17028 | 50 | 17029 | 50 | | | | | | |
| bread_other_spread_dunno_vlowfat_thin | 17028 | 50 | 17029 | 50 | | | | | | |
| bread_other_spread_hardmarg_med | 17018 | 50 | 17539 | 50 | | | | | | |
| bread_other_spread_hardmarg_thick | 17018 | 50 | 17539 | 50 | | | | | | |
| bread_other_spread_hardmarg_thin | 17018 | 50 | 17539 | 50 | | | | | | |
| bread_other_spread_olive_chol_med | 17552 | 50 | 17025 | 50 | | | | | | |
| bread_other_spread_olive_chol_thick | 17552 | 50 | 17025 | 50 | | | | | | |
| bread_other_spread_olive_chol_thin | 17552 | 50 | 17025 | 50 | | | | | | |
| bread_other_spread_olive_dunno_med | 17552 | 50 | 17025 | 50 | | | | | | |
| bread_other_spread_olive_dunno_thick | 17552 | 50 | 17025 | 50 | | | | | | |
| bread_other_spread_olive_dunno_thin | 17552 | 30 | 17025 | 70 | | | | | | |
| bread_other_spread_olive_fat_med | 17025 | 100 | | | | | | | | |
| bread_other_spread_olive_fat_thick | 17025 | 100 | | | | | | | | |
| bread_other_spread_olive_fat_thin | 17025 | 100 | | | | | | | | |
| bread_other_spread_olive_lowfat_med | 17552 | 100 | | | | | | | | |
| bread_other_spread_olive_lowfat_thick | 17552 | 100 | | | | | | | | |
| bread_other_spread_olive_lowfat_thin | 17552 | 100 | | | | | | | | |
| bread_other_spread_olive_vlowfat_med | 17028 | 100 | | | | | | | | |
| bread_other_spread_olive_vlowfat_thick | 17028 | 100 | | | | | | | | |
| bread_other_spread_olive_vlowfat_thin | 17028 | 100 | | | | | | | | |
| bread_other_spread_other_med | 17007 | 50 | 17487 | 50 | | | | | | |
| bread_other_spread_other_thick | 17007 | 50 | 17487 | 50 | | | | | | |
| bread_other_spread_other_thin | 17007 | 50 | 17487 | 50 | | | | | | |
| bread_other_spread_polymarg_chol_med | 17027 | 100 | | | | | | | | |
| bread_other_spread_polymarg_chol_thick | 17027 | 100 | | | | | | | | |
| bread_other_spread_polymarg_chol_thin | 17027 | 100 | | | | | | | | |
| bread_other_spread_polymarg_dunno_med | 17027 | 30 | 17024 | 70 | | | | | | |
| bread_other_spread_polymarg_dunno_thick | 17027 | 30 | 17024 | 70 | | | | | | |
| bread_other_spread_polymarg_dunno_thin | 17027 | 30 | 17024 | 70 | | | | | | |
| bread_other_spread_polymarg_fat_med | 17024 | 100 | | | | | | | | |
| bread_other_spread_polymarg_fat_thick | 17024 | 100 | | | | | | | | |
| bread_other_spread_polymarg_fat_thin | 17024 | 100 | | | | | | | | |
| bread_other_spread_polymarg_lowfat_med | 17027 | 100 | | | | | | | | |
| bread_other_spread_polymarg_lowfat_thick | 17027 | 100 | | | | | | | | |
| bread_other_spread_polymarg_lowfat_thin | 17027 | 100 | | | | | | | | |
| bread_other_spread_polymarg_vlowfat_med | 17029 | 100 | | | | | | | | |
| bread_other_spread_polymarg_vlowfat_thick | 17029 | 100 | | | | | | | | |
| bread_other_spread_polymarg_vlowfat_thin | 17029 | 100 | | | | | | | | |
| bread_other_spread_soya_chol_med | 17027 | 100 | | | | | | | | |
| bread_other_spread_soya_chol_thick | 17027 | 100 | | | | | | | | |
| bread_other_spread_soya_chol_thin | 17027 | 100 | | | | | | | | |
| bread_other_spread_soya_dunno_med | 17027 | 30 | 17024 | 70 | | | | | | |
| bread_other_spread_soya_dunno_thick | 17027 | 30 | 17024 | 70 | | | | | | |
| bread_other_spread_soya_dunno_thin | 17027 | 30 | 17024 | 70 | | | | | | |
| bread_other_spread_soya_fat_med | 17024 | 100 | | | | | | | | |
| bread_other_spread_soya_fat_thick | 17024 | 100 | | | | | | | | |
| bread_other_spread_soya_fat_thin | 17024 | 100 | | | | | | | | |
| bread_other_spread_soya_lowfat_med | 17027 | 100 | | | | | | | | |
| bread_other_spread_soya_lowfat_thick | 17027 | 100 | | | | | | | | |
| bread_other_spread_soya_lowfat_thin | 17027 | 100 | | | | | | | | |
| bread_other_spread_soya_vlowfat_med | 17029 | 100 | | | | | | | | |
| bread_other_spread_soya_vlowfat_thick | 17029 | 100 | | | | | | | | |
| bread_other_spread_soya_vlowfat_thin | 17029 | 100 | | | | | | | | |
| bread_roll_mixed | 11461 | 33.4 | 00033 | 33.3 | 11472 | 33.3 | | | | |
| bread_roll_other | 00040 | 50 | 00046 | 50 | | | | | | |
| bread_roll_seeded | 14844 | 50 | 14845 | 50 | | | | | | |
| bread_roll_spread_butter_dunno_med | 17486 | 100 | | | | | | | | |
| bread_roll_spread_butter_dunno_thick | 17486 | 100 | | | | | | | | |
| bread_roll_spread_butter_dunno_thin | 17486 | 100 | | | | | | | | |
| bread_roll_spread_butter_fat_med | 17485 | 100 | | | | | | | | |
| bread_roll_spread_butter_fat_thick | 17485 | 100 | | | | | | | | |
| bread_roll_spread_butter_fat_thin | 17485 | 100 | | | | | | | | |
| bread_roll_spread_butter_lowfat_med | 17485 | 50 | 17017 | 50 | | | | | | |
| bread_roll_spread_butter_lowfat_thick | 17485 | 50 | 17017 | 50 | | | | | | |
| bread_roll_spread_butter_lowfat_thin | 17485 | 50 | 17017 | 50 | | | | | | |
| bread_roll_spread_butter_spread_fat_med | 17486 | 100 | | | | | | | | |
| bread_roll_spread_butter_spread_fat_thick | 17486 | 100 | | | | | | | | |
| bread_roll_spread_butter_spread_fat_thin | 17486 | 100 | | | | | | | | |
| bread_roll_spread_butter_spread_lowfat_med | 17486 | 50 | 17017 | 50 | | | | | | |
| bread_roll_spread_butter_spread_lowfat_thick | 17486 | 50 | 17017 | 50 | | | | | | |
| bread_roll_spread_butter_spread_lowfat_thin | 17486 | 50 | 17017 | 50 | | | | | | |
| bread_roll_spread_dairy_chol_med | 17017 | 100 | | | | | | | | |

**Supplementary table 3**. Nutrient calculation in the previous version (McCance and Widdowson).

| | Food codes from McCance and Widdowson and the % used from each food code | | | | | | | | | |
|---|---|---|---|---|---|---|---|---|---|---|
| Item | Code1 | % from code 1 | Code2 | % from code 2 | Code3 | % from code 3 | Code4 | % from code 4 | Code5 | % from code 5 |
| bread_roll_spread_dairy_chol_thick | 17017 | 100 | | | | | | | | |
| bread_roll_spread_dairy_chol_thin | 17017 | 100 | | | | | | | | |
| bread_roll_spread_dairy_dunno_med | 17017 | 30 | 12258 | 70 | | | | | | |
| bread_roll_spread_dairy_dunno_thick | 17017 | 30 | 12258 | 70 | | | | | | |
| bread_roll_spread_dairy_dunno_thin | 17017 | 30 | 12258 | 70 | | | | | | |
| bread_roll_spread_dairy_fat_med | 12258 | 100 | | | | | | | | |
| bread_roll_spread_dairy_fat_thick | 12258 | 100 | | | | | | | | |
| bread_roll_spread_dairy_fat_thin | 12258 | 100 | | | | | | | | |
| bread_roll_spread_dairy_lowfat_med | 17017 | 100 | | | | | | | | |
| bread_roll_spread_dairy_lowfat_thick | 17017 | 100 | | | | | | | | |
| bread_roll_spread_dairy_lowfat_thin | 17017 | 100 | | | | | | | | |
| bread_roll_spread_dairy_vlowfat_med | 17028 | 100 | | | | | | | | |
| bread_roll_spread_dairy_vlowfat_thick | 17028 | 100 | | | | | | | | |
| bread_roll_spread_dairy_vlowfat_thin | 17028 | 100 | | | | | | | | |
| bread_roll_spread_dunno_chol_med | 17552 | 50 | 17027 | 50 | | | | | | |
| bread_roll_spread_dunno_chol_thick | 17552 | 50 | 17027 | 50 | | | | | | |
| bread_roll_spread_dunno_chol_thin | 17552 | 50 | 17027 | 50 | | | | | | |
| bread_roll_spread_dunno_dunno_med | 17552 | 25 | 17027 | 25 | 17025 | 25 | 17024 | 25 | | |
| bread_roll_spread_dunno_dunno_thick | 17552 | 25 | 17027 | 25 | 17025 | 25 | 17024 | 25 | | |
| bread_roll_spread_dunno_dunno_thin | 17552 | 25 | 17027 | 25 | 17025 | 25 | 17024 | 25 | | |
| bread_roll_spread_dunno_fat_med | 12258 | 33.3 | 17025 | 33.3 | 17024 | 33.4 | | | | |
| bread_roll_spread_dunno_fat_thick | 12258 | 33.3 | 17025 | 33.3 | 17024 | 33.4 | | | | |
| bread_roll_spread_dunno_fat_thin | 12258 | 33.3 | 17025 | 33.3 | 17024 | 33.4 | | | | |
| bread_roll_spread_dunno_lowfat_med | 17017 | 33.3 | 17552 | 33.3 | 17027 | 33.4 | | | | |
| bread_roll_spread_dunno_lowfat_thick | 17017 | 33.3 | 17552 | 33.3 | 17027 | 33.4 | | | | |
| bread_roll_spread_dunno_lowfat_thin | 17017 | 33.3 | 17552 | 33.3 | 17027 | 33.4 | | | | |
| bread_roll_spread_dunno_vlowfat_med | 17028 | 50 | 17029 | 50 | | | | | | |
| bread_roll_spread_dunno_vlowfat_thick | 17028 | 50 | 17029 | 50 | | | | | | |
| bread_roll_spread_dunno_vlowfat_thin | 17028 | 50 | 17029 | 50 | | | | | | |
| bread_roll_spread_hardmarg_med | 17018 | 50 | 17539 | 50 | | | | | | |
| bread_roll_spread_hardmarg_thick | 17018 | 50 | 17539 | 50 | | | | | | |
| bread_roll_spread_hardmarg_thin | 17018 | 50 | 17539 | 50 | | | | | | |
| bread_roll_spread_olive_chol_med | 17552 | 50 | 17025 | 50 | | | | | | |
| bread_roll_spread_olive_chol_thick | 17552 | 50 | 17025 | 50 | | | | | | |
| bread_roll_spread_olive_chol_thin | 17552 | 50 | 17025 | 50 | | | | | | |
| bread_roll_spread_olive_dunno_med | 17552 | 50 | 17025 | 50 | | | | | | |
| bread_roll_spread_olive_dunno_thick | 17552 | 50 | 17025 | 50 | | | | | | |
| bread_roll_spread_olive_dunno_thin | 17552 | 30 | 17025 | 70 | | | | | | |
| bread_roll_spread_olive_fat_med | 17025 | 100 | | | | | | | | |
| bread_roll_spread_olive_fat_thick | 17025 | 100 | | | | | | | | |
| bread_roll_spread_olive_fat_thin | 17025 | 100 | | | | | | | | |
| bread_roll_spread_olive_lowfat_med | 17552 | 100 | | | | | | | | |
| bread_roll_spread_olive_lowfat_thick | 17552 | 100 | | | | | | | | |
| bread_roll_spread_olive_lowfat_thin | 17552 | 100 | | | | | | | | |
| bread_roll_spread_olive_vlowfat_med | 17028 | 100 | | | | | | | | |
| bread_roll_spread_olive_vlowfat_thick | 17028 | 100 | | | | | | | | |
| bread_roll_spread_olive_vlowfat_thin | 17028 | 100 | | | | | | | | |
| bread_roll_spread_other_med | 17007 | 50 | 17487 | 50 | | | | | | |
| bread_roll_spread_other_thick | 17007 | 50 | 17487 | 50 | | | | | | |
| bread_roll_spread_other_thin | 17007 | 50 | 17487 | 50 | | | | | | |
| bread_roll_spread_polymarg_chol_med | 17027 | 100 | | | | | | | | |
| bread_roll_spread_polymarg_chol_thick | 17027 | 100 | | | | | | | | |
| bread_roll_spread_polymarg_chol_thin | 17027 | 100 | | | | | | | | |
| bread_roll_spread_polymarg_dunno_med | 17027 | 30 | 17024 | 70 | | | | | | |
| bread_roll_spread_polymarg_dunno_thick | 17027 | 30 | 17024 | 70 | | | | | | |
| bread_roll_spread_polymarg_dunno_thin | 17027 | 30 | 17024 | 70 | | | | | | |
| bread_roll_spread_polymarg_fat_med | 17024 | 100 | | | | | | | | |
| bread_roll_spread_polymarg_fat_thick | 17024 | 100 | | | | | | | | |
| bread_roll_spread_polymarg_fat_thin | 17024 | 100 | | | | | | | | |
| bread_roll_spread_polymarg_lowfat_med | 17027 | 100 | | | | | | | | |
| bread_roll_spread_polymarg_lowfat_thick | 17027 | 100 | | | | | | | | |
| bread_roll_spread_polymarg_lowfat_thin | 17027 | 100 | | | | | | | | |
| bread_roll_spread_polymarg_vlowfat_med | 17029 | 100 | | | | | | | | |
| bread_roll_spread_polymarg_vlowfat_thick | 17029 | 100 | | | | | | | | |
| bread_roll_spread_polymarg_vlowfat_thin | 17029 | 100 | | | | | | | | |
| bread_roll_spread_soya_chol_med | 17027 | 100 | | | | | | | | |
| bread_roll_spread_soya_chol_thick | 17027 | 100 | | | | | | | | |
| bread_roll_spread_soya_chol_thin | 17027 | 100 | | | | | | | | |
| bread_roll_spread_soya_dunno_med | 17027 | 30 | 17024 | 70 | | | | | | |
| bread_roll_spread_soya_dunno_thick | 17027 | 30 | 17024 | 70 | | | | | | |
| bread_roll_spread_soya_dunno_thin | 17027 | 30 | 17024 | 70 | | | | | | |
| bread_roll_spread_soya_fat_med | 17024 | 100 | | | | | | | | |
| bread_roll_spread_soya_fat_thick | 17024 | 100 | | | | | | | | |
| bread_roll_spread_soya_fat_thin | 17024 | 100 | | | | | | | | |
| bread_roll_spread_soya_lowfat_med | 17027 | 100 | | | | | | | | |
| bread_roll_spread_soya_lowfat_thick | 17027 | 100 | | | | | | | | |
| bread_roll_spread_soya_lowfat_thin | 17027 | 100 | | | | | | | | |
| bread_roll_spread_soya_vlowfat_med | 17029 | 100 | | | | | | | | |
| bread_roll_spread_soya_vlowfat_thick | 17029 | 100 | | | | | | | | |
| bread_roll_spread_soya_vlowfat_thin | 17029 | 100 | | | | | | | | |
| bread_roll_unanswered | 00048 | 33.4 | 00056 | 33.3 | 00033 | 33.3 | | | | |
| bread_roll_white | 11482 | 50 | 11481 | 50 | | | | | | |
| bread_roll_wholemeal | 11484 | 100 | | | | | | | | |
| bread_sliced_mixed | 11461 | 33.4 | 00033 | 33.3 | 11472 | 33.3 | | | | |
| bread_sliced_other | 00040 | 50 | 00046 | 50 | | | | | | |
| bread_sliced_seeded | 14844 | 50 | 14845 | 50 | | | | | | |
| bread_sliced_spread_butter_dunno_med | 17486 | 100 | | | | | | | | |
| bread_sliced_spread_butter_dunno_thick | 17486 | 100 | | | | | | | | |
| bread_sliced_spread_butter_dunno_thin | 17486 | 100 | | | | | | | | |
| bread_sliced_spread_butter_fat_med | 17485 | 100 | | | | | | | | |
| bread_sliced_spread_butter_fat_thick | 17485 | 100 | | | | | | | | |
| bread_sliced_spread_butter_fat_thin | 17485 | 100 | | | | | | | | |
| bread_sliced_spread_butter_lowfat_med | 17485 | 50 | 17017 | 50 | | | | | | |
| bread_sliced_spread_butter_lowfat_thick | 17485 | 50 | 17017 | 50 | | | | | | |
| bread_sliced_spread_butter_lowfat_thin | 17485 | 50 | 17017 | 50 | | | | | | |
| bread_sliced_spread_butter_spread_fat_med | 17486 | 100 | | | | | | | | |
| bread_sliced_spread_butter_spread_fat_thick | 17486 | 100 | | | | | | | | |
| bread_sliced_spread_butter_spread_fat_thin | 17486 | 100 | | | | | | | | |
| bread_sliced_spread_butter_spread_lowfat_med | 17486 | 50 | 17017 | 50 | | | | | | |
| bread_sliced_spread_butter_spread_lowfat_thick | 17486 | 50 | 17017 | 50 | | | | | | |
| bread_sliced_spread_butter_spread_lowfat_thin | 17486 | 50 | 17017 | 50 | | | | | | |
| bread_sliced_spread_dairy_chol_med | 17017 | 100 | | | | | | | | |
| bread_sliced_spread_dairy_chol_thick | 17017 | 100 | | | | | | | | |
| bread_sliced_spread_dairy_chol_thin | 17017 | 100 | | | | | | | | |
| bread_sliced_spread_dairy_dunno_med | 17017 | 30 | 12258 | 70 | | | | | | |
| bread_sliced_spread_dairy_dunno_thick | 17017 | 30 | 12258 | 70 | | | | | | |
| bread_sliced_spread_dairy_dunno_thin | 17017 | 30 | 12258 | 70 | | | | | | |
| bread_sliced_spread_dairy_fat_med | 12258 | 100 | | | | | | | | |
| bread_sliced_spread_dairy_fat_thick | 12258 | 100 | | | | | | | | |

**Supplementary table 3**. Nutrient calculation in the previous version (McCance and Widdowson).

| | Food codes from McCance and Widdowson and the % used from each food code | | | | | | | | | |
|---|---|---|---|---|---|---|---|---|---|---|
| Item | Code1 | % from code 1 | Code2 | % from code 2 | Code3 | % from code 3 | Code4 | % from code 4 | Code5 | % from code 5 |
| bread_sliced_spread_dairy_fat_thin | 12258 | 100 | | | | | | | | |
| bread_sliced_spread_dairy_lowfat_med | 17017 | 100 | | | | | | | | |
| bread_sliced_spread_dairy_lowfat_thick | 17017 | 100 | | | | | | | | |
| bread_sliced_spread_dairy_lowfat_thin | 17017 | 100 | | | | | | | | |
| bread_sliced_spread_dairy_vlowfat_med | 17028 | 100 | | | | | | | | |
| bread_sliced_spread_dairy_vlowfat_thick | 17028 | 100 | | | | | | | | |
| bread_sliced_spread_dairy_vlowfat_thin | 17028 | 100 | | | | | | | | |
| bread_sliced_spread_dunno_chol_med | 17552 | 50 | 17027 | 50 | | | | | | |
| bread_sliced_spread_dunno_chol_thick | 17552 | 50 | 17027 | 50 | | | | | | |
| bread_sliced_spread_dunno_chol_thin | 17552 | 50 | 17027 | 50 | | | | | | |
| bread_sliced_spread_dunno_dunno_med | 17552 | 25 | 17027 | 25 | 17025 | 25 | 17024 | 25 | | |
| bread_sliced_spread_dunno_dunno_thick | 17552 | 25 | 17027 | 25 | 17025 | 25 | 17024 | 25 | | |
| bread_sliced_spread_dunno_dunno_thin | 17552 | 25 | 17027 | 25 | 17025 | 25 | 17024 | 25 | | |
| bread_sliced_spread_dunno_fat_med | 12258 | 33.3 | 17025 | 33.3 | 17024 | 33.4 | | | | |
| bread_sliced_spread_dunno_fat_thick | 12258 | 33.3 | 17025 | 33.3 | 17024 | 33.4 | | | | |
| bread_sliced_spread_dunno_fat_thin | 12258 | 33.3 | 17025 | 33.3 | 17024 | 33.4 | | | | |
| bread_sliced_spread_dunno_lowfat_med | 17017 | 33.3 | 17552 | 33.3 | 17027 | 33.4 | | | | |
| bread_sliced_spread_dunno_lowfat_thick | 17017 | 33.3 | 17552 | 33.3 | 17027 | 33.4 | | | | |
| bread_sliced_spread_dunno_lowfat_thin | 17017 | 33.3 | 17552 | 33.3 | 17027 | 33.4 | | | | |
| bread_sliced_spread_dunno_vlowfat_med | 17028 | 50 | 17029 | 50 | | | | | | |
| bread_sliced_spread_dunno_vlowfat_thick | 17028 | 50 | 17029 | 50 | | | | | | |
| bread_sliced_spread_dunno_vlowfat_thin | 17028 | 50 | 17029 | 50 | | | | | | |
| bread_sliced_spread_hardmarg_med | 17018 | 50 | 17539 | 50 | | | | | | |
| bread_sliced_spread_hardmarg_thick | 17018 | 50 | 17539 | 50 | | | | | | |
| bread_sliced_spread_hardmarg_thin | 17018 | 50 | 17539 | 50 | | | | | | |
| bread_sliced_spread_olive_chol_med | 17552 | 50 | 17025 | 50 | | | | | | |
| bread_sliced_spread_olive_chol_thick | 17552 | 50 | 17025 | 50 | | | | | | |
| bread_sliced_spread_olive_chol_thin | 17552 | 50 | 17025 | 50 | | | | | | |
| bread_sliced_spread_olive_dunno_med | 17552 | 50 | 17025 | 50 | | | | | | |
| bread_sliced_spread_olive_dunno_thick | 17552 | 50 | 17025 | 50 | | | | | | |
| bread_sliced_spread_olive_dunno_thin | 17552 | 30 | 17025 | 70 | | | | | | |
| bread_sliced_spread_olive_fat_med | 17025 | 100 | | | | | | | | |
| bread_sliced_spread_olive_fat_thick | 17025 | 100 | | | | | | | | |
| bread_sliced_spread_olive_fat_thin | 17025 | 100 | | | | | | | | |
| bread_sliced_spread_olive_lowfat_med | 17552 | 100 | | | | | | | | |
| bread_sliced_spread_olive_lowfat_thick | 17552 | 100 | | | | | | | | |
| bread_sliced_spread_olive_lowfat_thin | 17552 | 100 | | | | | | | | |
| bread_sliced_spread_olive_vlowfat_med | 17028 | 100 | | | | | | | | |
| bread_sliced_spread_olive_vlowfat_thick | 17028 | 100 | | | | | | | | |
| bread_sliced_spread_olive_vlowfat_thin | 17028 | 100 | | | | | | | | |
| bread_sliced_spread_other_med | 17007 | 50 | 17487 | 50 | | | | | | |
| bread_sliced_spread_other_thick | 17007 | 50 | 17487 | 50 | | | | | | |
| bread_sliced_spread_other_thin | 17007 | 50 | 17487 | 50 | | | | | | |
| bread_sliced_spread_polymarg_chol_med | 17027 | 100 | | | | | | | | |
| bread_sliced_spread_polymarg_chol_thick | 17027 | 100 | | | | | | | | |
| bread_sliced_spread_polymarg_chol_thin | 17027 | 100 | | | | | | | | |
| bread_sliced_spread_polymarg_dunno_med | 17027 | 30 | 17024 | 70 | | | | | | |
| bread_sliced_spread_polymarg_dunno_thick | 17027 | 30 | 17024 | 70 | | | | | | |
| bread_sliced_spread_polymarg_dunno_thin | 17027 | 30 | 17024 | 70 | | | | | | |
| bread_sliced_spread_polymarg_fat_med | 17024 | 100 | | | | | | | | |
| bread_sliced_spread_polymarg_fat_thick | 17024 | 100 | | | | | | | | |
| bread_sliced_spread_polymarg_fat_thin | 17024 | 100 | | | | | | | | |
| bread_sliced_spread_polymarg_lowfat_med | 17027 | 100 | | | | | | | | |
| bread_sliced_spread_polymarg_lowfat_thick | 17027 | 100 | | | | | | | | |
| bread_sliced_spread_polymarg_lowfat_thin | 17027 | 100 | | | | | | | | |
| bread_sliced_spread_polymarg_vlowfat_med | 17029 | 100 | | | | | | | | |
| bread_sliced_spread_polymarg_vlowfat_thick | 17029 | 100 | | | | | | | | |
| bread_sliced_spread_polymarg_vlowfat_thin | 17029 | 100 | | | | | | | | |
| bread_sliced_spread_soya_chol_med | 17027 | 100 | | | | | | | | |
| bread_sliced_spread_soya_chol_thick | 17027 | 100 | | | | | | | | |
| bread_sliced_spread_soya_chol_thin | 17027 | 100 | | | | | | | | |
| bread_sliced_spread_soya_dunno_med | 17027 | 30 | 17024 | 70 | | | | | | |
| bread_sliced_spread_soya_dunno_thick | 17027 | 30 | 17024 | 70 | | | | | | |
| bread_sliced_spread_soya_dunno_thin | 17027 | 30 | 17024 | 70 | | | | | | |
| bread_sliced_spread_soya_fat_med | 17024 | 100 | | | | | | | | |
| bread_sliced_spread_soya_fat_thick | 17024 | 100 | | | | | | | | |
| bread_sliced_spread_soya_fat_thin | 17024 | 100 | | | | | | | | |
| bread_sliced_spread_soya_lowfat_med | 17027 | 100 | | | | | | | | |
| bread_sliced_spread_soya_lowfat_thick | 17027 | 100 | | | | | | | | |
| bread_sliced_spread_soya_lowfat_thin | 17027 | 100 | | | | | | | | |
| bread_sliced_spread_soya_vlowfat_med | 17029 | 100 | | | | | | | | |
| bread_sliced_spread_soya_vlowfat_thick | 17029 | 100 | | | | | | | | |
| bread_sliced_spread_soya_vlowfat_thin | 17029 | 100 | | | | | | | | |
| bread_sliced_unanswered | 00048 | 33.4 | 00056 | 33.3 | 00033 | 33.3 | | | | |
| bread_sliced_white | 00048 | 45 | 00054 | 10 | 11468 | 45 | | | | |
| bread_sliced_wholemeal | 00056 | 100 | | | | | | | | |
| cake | 11527 | 25 | 11571 | 25 | 11616 | 25 | 12394 | 25 | | |
| cereal_artf_swt | | | | | | | | | | |
| cereal_bran | 11485 | 50 | 11486 | 50 | | | | | | |
| cereal_bran_driedfruit | 11493 | 100 | | | | | | | | |
| cereal_muesli | 11494 | 50 | 11495 | 50 | | | | | | |
| cereal_muesli_driedfruit | 11138 | 100 | | | | | | | | |
| cereal_oatcrunch | 81361 | 50 | 11487 | 50 | | | | | | |
| cereal_oatcrunch_driedfruit | 81361 | 45 | 11487 | 45 | 888 | 10 | | | | |
| cereal_other | 81362 | 100 | | | | | | | | |
| cereal_other_driedfruit | 81362 | 80 | 888 | 20 | | | | | | |
| cereal_plain | 11490 | 33.4 | 11497 | 33.3 | 11501 | 33.3 | | | | |
| cereal_plain_driedfruit | 11490 | 26.6 | 11497 | 26.7 | 11501 | 26.7 | 888 | 20 | | |
| cereal_porridge_milk | 11570 | 100 | | | | | | | | |
| cereal_porridge_milk_driedfruit | 11570 | 91 | 888 | 9 | | | | | | |
| cereal_porridge_water | 11569 | 90 | 11496 | 10 | | | | | | |
| cereal_porridge_water_driedfruit | 11569 | 83 | 11496 | 8 | 888 | 9 | | | | |
| cereal_sugar | 17063 | 25 | 17074 | 25 | 17050 | 25 | 17065 | 25 | | |
| cereal_sweet | 11491 | 25 | 11498 | 25 | 11488 | 25 | 11612 | 25 | | |
| cereal_sweet_driedfruit | 11491 | 20 | 11498 | 20 | 11488 | 20 | 11612 | 20 | 888 | 20 |
| cereal_wwheat | 11499 | 33.4 | 11500 | 33.3 | 11505 | 33.3 | | | | |
| cereal_wwheat_driedfruit | 11500 | 26.6 | 11501 | 26.7 | 11506 | 26.7 | 888 | 20 | | |
| cerealbar | 17494 | 50 | 17103 | 50 | | | | | | |
| cheese_blue | 12177 | 33.4 | 12354 | 33.3 | 12367 | 33.3 | | | | |
| cheese_cottage | 12351 | 33.4 | 12352 | 33.3 | 12148 | 33.3 | | | | |
| cheese_feta | 12356 | 100 | | | | | | | | |
| cheese_goat | 12162 | 50 | 12357 | 50 | | | | | | |
| cheese_hard | 12348 | 50 | 12359 | 50 | | | | | | |
| cheese_hard_lof | 12155 | 33.4 | 12348 | 33.3 | 12355 | 33.3 | | | | |
| cheese_mozzarella | 12170 | 50 | 12360 | 50 | | | | | | |
| cheese_other | 12362 | 50 | 12368 | 50 | | | | | | |
| cheese_soft | 12168 | 33.4 | 12344 | 33.3 | 12345 | 33.3 | | | | |
| cheese_spread | 12143 | 25 | 12353 | 25 | 12364 | 25 | 12365 | 25 | | |
| cheese_spread_lof | 12366 | 100 | | | | | | | | |

**Supplementary table 3**. Nutrient calculation in the previous version (McCance and Widdowson).

| | Food codes from McCance and Widdowson and the % used from each food code | | | | | | | | | |
|---|---|---|---|---|---|---|---|---|---|---|
| **Item** | **Code1** | **% from code 1** | **Code2** | **% from code 2** | **Code3** | **% from code 3** | **Code4** | **% from code 4** | **Code5** | **% from code 5** |
| cheesecake | 12218 | 50 | 12395 | 50 | | | | | | |
| choc_bar | 17547 | 50 | 17549 | 50 | | | | | | |
| choc_dark | 17090 | 100 | | | | | | | | |
| choc_milk | 17089 | 100 | | | | | | | | |
| choc_sweets | 17088 | 100 | | | | | | | | |
| choc_white | 17091 | 100 | | | | | | | | |
| chocyog_raisin | 14835 | 100 | | | | | | | | |
| chutney | 17341 | 50 | 17352 | 50 | | | | | | |
| cof_artf_swt | | | | | | | | | | |
| cof_capp_decaf_other | 17153 | 33.3 | 82002 | 66.7 | | | | | | |
| cof_capp_decaf_semi | 17153 | 33.3 | 12313 | 66.7 | | | | | | |
| cof_capp_decaf_skimmed | 17153 | 33.3 | 12307 | 66.7 | | | | | | |
| cof_capp_decaf_whole | 17153 | 33.3 | 12316 | 66.7 | | | | | | |
| cof_capp_other | 17153 | 33.3 | 82002 | 66.7 | | | | | | |
| cof_capp_semi | 17153 | 33.3 | 12313 | 66.7 | | | | | | |
| cof_capp_skimmed | 17153 | 33.3 | 12307 | 66.7 | | | | | | |
| cof_capp_whole | 17153 | 33.3 | 12316 | 66.7 | | | | | | |
| cof_espresso | 17153 | 100 | | | | | | | | |
| cof_espresso_decaf | 17153 | 100 | | | | | | | | |
| cof_filter | 17152 | 100 | | | | | | | | |
| cof_filter_decaf | 17152 | 100 | | | | | | | | |
| cof_instant | 17159 | 100 | | | | | | | | |
| cof_instant_decaf | 17159 | 100 | | | | | | | | |
| cof_latte_decaf_other | 17153 | 33.3 | 82002 | 66.7 | | | | | | |
| cof_latte_decaf_semi | 17153 | 33.3 | 12313 | 66.7 | | | | | | |
| cof_latte_decaf_skimmed | 17153 | 33.3 | 12307 | 66.7 | | | | | | |
| cof_latte_decaf_whole | 17153 | 33.3 | 12316 | 66.7 | | | | | | |
| cof_latte_other | 17153 | 33.3 | 82002 | 66.7 | | | | | | |
| cof_latte_semi | 17153 | 33.3 | 12313 | 66.7 | | | | | | |
| cof_latte_skimmed | 17153 | 33.3 | 12307 | 66.7 | | | | | | |
| cof_latte_whole | 17153 | 33.3 | 12316 | 66.7 | | | | | | |
| cof_other | 17159 | 100 | | | | | | | | |
| cof_other_decaf | 17159 | 100 | | | | | | | | |
| cof_sugar | 17063 | 66.7 | 17061 | 33.3 | | | | | | |
| cream | 12332 | 20 | 12334 | 20 | 213 | 20 | 12335 | 20 | 12353 | 20 |
| croissant | 11480 | 100 | | | | | | | | |
| crumble | 11439 | 25 | 11018 | 25 | 17061 | 14 | 17485 | 22 | 12346 | 14 |
| danish_pastry | 11538 | 100 | | | | | | | | |
| dessert_milkbased | 12400 | 75 | 12397 | 25 | | | | | | |
| dessert_milkpuds | 12225 | 17 | 12413 | 17 | 12406 | 33 | 12217 | 33 | | |
| dessert_other | 12405 | 25 | 12830 | 25 | 12404 | 25 | 12252 | 25 | | |
| dessert_soya | 12196 | 100 | | | | | | | | |
| double_crust | 11585 | **90** | 11587 | **10** | | | | | | |
| doughnut | 11539 | 50 | 11241 | 25 | 11242 | 25 | | | | |
| drink_diethotchoc | 17500 | 10 | 12307 | 90 | | | | | | |
| drink_fizzy | 17175 | 25 | 17177 | 25 | 17178 | 25 | 17179 | 25 | | |
| drink_grapefruit | 14276 | 50 | 14275 | 50 | | | | | | |
| drink_hotchoc_other | 17498 | 9 | 82002 | 91 | | | | | | |
| drink_hotchoc_semi | 17532 | 100 | | | | | | | | |
| drink_hotchoc_skimmed | 12096 | 100 | | | | | | | | |
| drink_hotchoc_whole | 17533 | 100 | | | | | | | | |
| drink_lowcal | 17505 | 50 | 87001 | 50 | | | | | | |
| drink_milkbased | 12193 | 25 | 12326 | 25 | 12327 | 25 | 17203 | 25 | | |
| drink_orange | 14281 | 100 | | | | | | | | |
| drink_other | 17501 | 100 | | | | | | | | |
| drink_purejuice | 14271 | 60 | 17537 | 30 | 13382 | 10 | | | | |
| drink_squash | 17190 | 25 | 17195 | 25 | 17198 | 25 | 17201 | 25 | | |
| drink_water | 01186 | 80 | 17182 | 20 | | | | | | |
| drizzle_oil | 17038 | 100 | | | | | | | | |
| egg_omelet | 12812 | 25 | 12926 | 25 | 12921 | 25 | 12922 | 25 | | |
| egg_other | 12812 | 100 | | | | | | | | |
| egg_scotch | 12825 | 50 | 19320 | 50 | | | | | | |
| egg_swich | 293 | 90 | 17510 | 10 | | | | | | |
| egg_whole | 12806 | 33.4 | 12810 | 33.3 | 12919 | 33.3 | | | | |
| fish_battered | 16023 | 50 | 16054 | 50 | | | | | | |
| fish_breaded | 16288 | 50 | 16281 | 50 | | | | | | |
| fish_lobcrab | 16331 | 50 | 16332 | 50 | | | | | | |
| fish_oily | 16176 | 20 | 16188 | 20 | 16192 | 20 | 16327 | 20 | 16329 | 20 |
| fish_other | 16013 | 33.4 | 16327 | 33.3 | 16192 | 33.3 | | | | |
| fish_prawns | 16239 | 100 | | | | | | | | |
| fish_shell | 16262 | 30 | 16256 | 70 | | | | | | |
| fish_tinnedtuna | 16339 | 50 | 16230 | 50 | | | | | | |
| fish_white | 16013 | 50 | 16045 | 50 | | | | | | |
| fruit_apple | 14013 | 100 | | | | | | | | |
| fruit_banana | 14045 | 100 | | | | | | | | |
| fruit_berry | 14244 | 33.3 | 14260 | 33.4 | 14053 | 33.3 | | | | |
| fruit_cherry | 14061 | 100 | | | | | | | | |
| fruit_dried | 14031 | 33.4 | 14016 | 33.3 | 14242 | 33.3 | | | | |
| fruit_grapefruit | 14105 | 100 | | | | | | | | |
| fruit_grapes | 14109 | 100 | | | | | | | | |
| fruit_mango | 14148 | 100 | | | | | | | | |
| fruit_melon | 14153 | 100 | | | | | | | | |
| fruit_mixed | 14303 | 50 | 14096 | 50 | | | | | | |
| fruit_orange | 14176 | 100 | | | | | | | | |
| fruit_other | 14124 | 50 | 14208 | 50 | | | | | | |
| fruit_peach | 14183 | 50 | 14297 | 50 | | | | | | |
| fruit_pear | 14191 | 100 | | | | | | | | |
| fruit_pineapple | 14208 | 70 | 14211 | 30 | | | | | | |
| fruit_plum | 14213 | 100 | | | | | | | | |
| fruit_prunes | 14231 | 100 | | | | | | | | |
| fruit_satsuma | 14258 | 100 | | | | | | | | |
| fruit_stewed | 14005 | 33.4 | 14253 | 33.3 | 14215 | 33.3 | | | | |
| fruitcake | 11577 | 50 | 11529 | 50 | | | | | | |
| grains_couscous | 11339 | 100 | | | | | | | | |
| grains_other | 11003 | 33.4 | 11007 | 33.3 | 14843 | 33.3 | | | | |
| guacamole | 15180 | 100 | | | | | | | | |
| hummus | 13433 | 100 | | | | | | | | |
| icecream | 12387 | 33.4 | 12205 | 33.3 | 12200 | 33.3 | | | | |
| indian_snack | 15227 | 25 | 15231 | 25 | 19059 | 25 | 175 | 25 | | |
| jam_honey | 17074 | 33.4 | 17050 | 33.3 | 17065 | 33.3 | | | | |
| mayo | 17317 | 50 | 17510 | 50 | | | | | | |
| mayo_lowfat | 17511 | 100 | | | | | | | | |
| meat_bacon_nofat | 19008 | 100 | | | | | | | | |
| meat_bacon_withfat | 19003 | 50 | 19015 | 50 | | | | | | |
| meat_beef_nofat | 18049 | 100 | | | | | | | | |
| meat_beef_withfat | 373 | 100 | | | | | | | | |
| meat_ham_nofat | 19308 | 80 | 19025 | 10 | 19027 | 10 | | | | |
| meat_ham_withfat | 19308 | 60 | 19025 | 10 | 19027 | 10 | 19110 | 10 | 19142 | 10 |
| meat_lamb_nofat | 18141 | 100 | | | | | | | | |

**Supplementary table 3**. Nutrient calculation in the previous version (McCance and Widdowson).

| | Food codes from McCance and Widdowson and the % used from each food code | | | | | | | | | |
|---|---|---|---|---|---|---|---|---|---|---|
| **Item** | **Code1** | **% from code 1** | **Code2** | **% from code 2** | **Code3** | **% from code 3** | **Code4** | **% from code 4** | **Code5** | **% from code 5** |
| meat_lamb_withfat | 18477 | 100 | | | | | | | | |
| meat_liverpate | 488 | 20 | 18418 | 20 | 19317 | 60 | | | | |
| meat_other | 18405 | 20 | 18374 | 40 | 19154 | 40 | | | | 20 |
| meat_pork_nofat | 18251 | 100 | | | | | | | | |
| meat_pork_withfat | 18252 | 100 | | | | | | | | |
| meat_sausage | 19077 | 25 | 19080 | 75 | | | | | | |
| milk_chol_cereal | 12313 | 50 | 12307 | 50 | | | | | | |
| milk_chol_coffee | 12313 | 37.5 | 12307 | 37.5 | 12332 | 12.5 | 12027 | 12.5 | | |
| milk_chol_glass | 12313 | 50 | 12307 | 50 | | | | | | |
| milk_chol_tea | 12313 | 50 | 12307 | 50 | | | | | | |
| milk_dontknow_cereal | 82002 | 100 | | | | | | | | |
| milk_dontknow_coffee | 82002 | 75 | 12332 | 12.5 | 12027 | 12.5 | | | | |
| milk_dontknow_glass | 82002 | 100 | | | | | | | | |
| milk_dontknow_tea | 82002 | 100 | | | | | | | | |
| milk_goatsheep_cereal | 12328 | 50 | 12329 | 50 | | | | | | |
| milk_goatsheep_coffee | 12328 | 50 | 12329 | 50 | | | | | | |
| milk_goatsheep_glass | 12328 | 50 | 12329 | 50 | | | | | | |
| milk_goatsheep_tea | 12328 | 50 | 12329 | 50 | | | | | | |
| milk_other_cereal | 82002 | 100 | | | | | | | | |
| milk_other_coffee | 82002 | 75 | 12332 | 12.5 | 12027 | 12.5 | | | | |
| milk_other_glass | 82002 | 100 | | | | | | | | |
| milk_other_tea | 82002 | 100 | | | | | | | | |
| milk_powdered_cereal | 12030 | 70 | 12031 | 30 | | | | | | |
| milk_powdered_coffee | 12030 | 50 | 12031 | 25 | 12332 | 12.5 | 12027 | 12.5 | | |
| milk_powdered_glass | 12030 | 70 | 12031 | 30 | | | | | | |
| milk_powdered_tea | 12030 | 70 | 12031 | 30 | | | | | | |
| milk_riceoatveg_cereal | 12042 | 50 | 12331 | 50 | | | | | | |
| milk_riceoatveg_coffee | 12042 | 50 | 12331 | 50 | | | | | | |
| milk_riceoatveg_glass | 12042 | 50 | 12331 | 50 | | | | | | |
| milk_riceoatveg_tea | 12042 | 50 | 12331 | 50 | | | | | | |
| milk_semi_cereal | 12313 | 100 | | | | | | | | |
| milk_semi_coffee | 12313 | 75 | 12332 | 12.5 | 12027 | 12.5 | | | | |
| milk_semi_glass | 12313 | 100 | | | | | | | | |
| milk_semi_tea | 12313 | 100 | | | | | | | | |
| milk_skimmed_cereal | 12307 | 100 | | | | | | | | |
| milk_skimmed_coffee | 12307 | 75 | 12332 | 12.5 | 12027 | 12.5 | | | | |
| milk_skimmed_glass | 12307 | 100 | | | | | | | | |
| milk_skimmed_tea | 12307 | 100 | | | | | | | | |
| milk_soya_ca_cereal | 12331 | 100 | | | | | | | | |
| milk_soya_ca_coffee | 12331 | 100 | | | | | | | | |
| milk_soya_ca_glass | 12331 | 100 | | | | | | | | |
| milk_soya_ca_tea | 12331 | 100 | | | | | | | | |
| milk_soya_noca_cereal | 12042 | 50 | 12331 | 50 | | | | | | |
| milk_soya_noca_coffee | 12042 | 50 | 12331 | 50 | | | | | | |
| milk_soya_noca_glass | 12042 | 50 | 12331 | 50 | | | | | | |
| milk_soya_noca_tea | 12042 | 50 | 12331 | 50 | | | | | | |
| milk_whole_cereal | 12316 | 100 | | | | | | | | |
| milk_whole_coffee | 12316 | 75 | 12332 | 12.5 | 12027 | 12.5 | | | | |
| milk_whole_glass | 12316 | 100 | | | | | | | | |
| milk_whole_tea | 12316 | 100 | | | | | | | | |
| oatcakes | 11518 | 100 | | | | | | | | |
| oatcakes_spread_butter_dunno_med | 17486 | 100 | | | | | | | | |
| oatcakes_spread_butter_dunno_thick | 17486 | 100 | | | | | | | | |
| oatcakes_spread_butter_dunno_thin | 17486 | 100 | | | | | | | | |
| oatcakes_spread_butter_fat_med | 17485 | 100 | | | | | | | | |
| oatcakes_spread_butter_fat_thick | 17485 | 100 | | | | | | | | |
| oatcakes_spread_butter_fat_thin | 17485 | 100 | | | | | | | | |
| oatcakes_spread_butter_lowfat_med | 17485 | 50 | 17017 | 50 | | | | | | |
| oatcakes_spread_butter_lowfat_thick | 17485 | 50 | 17017 | 50 | | | | | | |
| oatcakes_spread_butter_lowfat_thin | 17485 | 50 | 17017 | 50 | | | | | | |
| oatcakes_spread_butter_spread_fat_med | 17486 | 100 | | | | | | | | |
| oatcakes_spread_butter_spread_fat_thick | 17486 | 100 | | | | | | | | |
| oatcakes_spread_butter_spread_fat_thin | 17486 | 100 | | | | | | | | |
| oatcakes_spread_butter_spread_lowfat_med | 17486 | 50 | 17017 | 50 | | | | | | |
| oatcakes_spread_butter_spread_lowfat_thick | 17486 | 50 | 17017 | 50 | | | | | | |
| oatcakes_spread_butter_spread_lowfat_thin | 17486 | 50 | 17017 | 50 | | | | | | |
| oatcakes_spread_dairy_chol_med | 17017 | 100 | | | | | | | | |
| oatcakes_spread_dairy_chol_thick | 17017 | 100 | | | | | | | | |
| oatcakes_spread_dairy_chol_thin | 17017 | 100 | | | | | | | | |
| oatcakes_spread_dairy_dunno_med | 17017 | 30 | 12258 | 70 | | | | | | |
| oatcakes_spread_dairy_dunno_thick | 17017 | 30 | 12258 | 70 | | | | | | |
| oatcakes_spread_dairy_dunno_thin | 17017 | 30 | 12258 | 70 | | | | | | |
| oatcakes_spread_dairy_fat_med | 12258 | 100 | | | | | | | | |
| oatcakes_spread_dairy_fat_thick | 12258 | 100 | | | | | | | | |
| oatcakes_spread_dairy_fat_thin | 12258 | 100 | | | | | | | | |
| oatcakes_spread_dairy_lowfat_med | 17017 | 100 | | | | | | | | |
| oatcakes_spread_dairy_lowfat_thick | 17017 | 100 | | | | | | | | |
| oatcakes_spread_dairy_lowfat_thin | 17017 | 100 | | | | | | | | |
| oatcakes_spread_dairy_vlowfat_med | 17028 | 100 | | | | | | | | |
| oatcakes_spread_dairy_vlowfat_thick | 17028 | 100 | | | | | | | | |
| oatcakes_spread_dairy_vlowfat_thin | 17028 | 100 | | | | | | | | |
| oatcakes_spread_dunno_chol_med | 17552 | 50 | 17027 | 50 | | | | | | |
| oatcakes_spread_dunno_chol_thick | 17552 | 50 | 17027 | 50 | | | | | | |
| oatcakes_spread_dunno_chol_thin | 17552 | 50 | 17027 | 50 | | | | | | |
| oatcakes_spread_dunno_dunno_med | 17552 | 25 | 17027 | 25 | 17025 | 25 | 17024 | 25 | | |
| oatcakes_spread_dunno_dunno_thick | 17552 | 25 | 17027 | 25 | 17025 | 25 | 17024 | 25 | | |
| oatcakes_spread_dunno_dunno_thin | 17552 | 25 | 17027 | 25 | 17025 | 25 | 17024 | 25 | | |
| oatcakes_spread_dunno_fat_med | 12258 | 33.3 | 17025 | 33.3 | 17024 | 33.4 | | | | |
| oatcakes_spread_dunno_fat_thick | 12258 | 33.3 | 17025 | 33.3 | 17024 | 33.4 | | | | |
| oatcakes_spread_dunno_fat_thin | 12258 | 33.3 | 17025 | 33.3 | 17024 | 33.4 | | | | |
| oatcakes_spread_dunno_lowfat_med | 17017 | 33.3 | 17552 | 33.3 | 17027 | 33.4 | | | | |
| oatcakes_spread_dunno_lowfat_thick | 17017 | 33.3 | 17552 | 33.3 | 17027 | 33.4 | | | | |
| oatcakes_spread_dunno_lowfat_thin | 17017 | 33.3 | 17552 | 33.3 | 17027 | 33.4 | | | | |
| oatcakes_spread_dunno_vlowfat_med | 17028 | 50 | 17029 | 50 | | | | | | |
| oatcakes_spread_dunno_vlowfat_thick | 17028 | 50 | 17029 | 50 | | | | | | |
| oatcakes_spread_dunno_vlowfat_thin | 17028 | 50 | 17029 | 50 | | | | | | |
| oatcakes_spread_hardmarg_med | 17018 | 50 | 17539 | 50 | | | | | | |
| oatcakes_spread_hardmarg_thick | 17018 | 50 | 17539 | 50 | | | | | | |
| oatcakes_spread_hardmarg_thin | 17018 | 50 | 17539 | 50 | | | | | | |
| oatcakes_spread_olive_chol_med | 17552 | 50 | 17025 | 50 | | | | | | |
| oatcakes_spread_olive_chol_thick | 17552 | 50 | 17025 | 50 | | | | | | |
| oatcakes_spread_olive_chol_thin | 17552 | 50 | 17025 | 50 | | | | | | |
| oatcakes_spread_olive_dunno_med | 17552 | 50 | 17025 | 50 | | | | | | |
| oatcakes_spread_olive_dunno_thick | 17552 | 50 | 17025 | 50 | | | | | | |
| oatcakes_spread_olive_dunno_thin | 17552 | 30 | 17025 | 70 | | | | | | |
| oatcakes_spread_olive_fat_med | 17025 | 100 | | | | | | | | |
| oatcakes_spread_olive_fat_thick | 17025 | 100 | | | | | | | | |
| oatcakes_spread_olive_fat_thin | 17025 | 100 | | | | | | | | |
| oatcakes_spread_olive_lowfat_med | 17552 | 100 | | | | | | | | |

**Supplementary table 3**. Nutrient calculation in the previous version (McCance and Widdowson).

| | Food codes from McCance and Widdowson and the % used from each food code | | | | | | | | | |
|---|---|---|---|---|---|---|---|---|---|---|
| **Item** | **Code1** | **% from code 1** | **Code2** | **% from code 2** | **Code3** | **% from code 3** | **Code4** | **% from code 4** | **Code5** | **% from code 5** |
| oatcakes_spread_olive_lowfat_thick | 17552 | 100 | | | | | | | | |
| oatcakes_spread_olive_lowfat_thin | 17552 | 100 | | | | | | | | |
| oatcakes_spread_olive_vlowfat_med | 17028 | 100 | | | | | | | | |
| oatcakes_spread_olive_vlowfat_thick | 17028 | 100 | | | | | | | | |
| oatcakes_spread_olive_vlowfat_thin | 17028 | 100 | | | | | | | | |
| oatcakes_spread_other_med | 17007 | 50 | 17487 | 50 | | | | | | |
| oatcakes_spread_other_thick | 17007 | 50 | 17487 | 50 | | | | | | |
| oatcakes_spread_other_thin | 17007 | 50 | 17487 | 50 | | | | | | |
| oatcakes_spread_polymarg_chol_med | 17027 | 100 | | | | | | | | |
| oatcakes_spread_polymarg_chol_thick | 17027 | 100 | | | | | | | | |
| oatcakes_spread_polymarg_chol_thin | 17027 | 100 | | | | | | | | |
| oatcakes_spread_polymarg_dunno_med | 17027 | 30 | 17024 | 70 | | | | | | |
| oatcakes_spread_polymarg_dunno_thick | 17027 | 30 | 17024 | 70 | | | | | | |
| oatcakes_spread_polymarg_dunno_thin | 17027 | 30 | 17024 | 70 | | | | | | |
| oatcakes_spread_polymarg_fat_med | 17024 | 100 | | | | | | | | |
| oatcakes_spread_polymarg_fat_thick | 17024 | 100 | | | | | | | | |
| oatcakes_spread_polymarg_fat_thin | 17024 | 100 | | | | | | | | |
| oatcakes_spread_polymarg_lowfat_med | 17027 | 100 | | | | | | | | |
| oatcakes_spread_polymarg_lowfat_thick | 17027 | 100 | | | | | | | | |
| oatcakes_spread_polymarg_lowfat_thin | 17027 | 100 | | | | | | | | |
| oatcakes_spread_polymarg_vlowfat_med | 17029 | 100 | | | | | | | | |
| oatcakes_spread_polymarg_vlowfat_thick | 17029 | 100 | | | | | | | | |
| oatcakes_spread_polymarg_vlowfat_thin | 17029 | 100 | | | | | | | | |
| oatcakes_spread_soya_chol_med | 17027 | 100 | | | | | | | | |
| oatcakes_spread_soya_chol_thick | 17027 | 100 | | | | | | | | |
| oatcakes_spread_soya_chol_thin | 17027 | 100 | | | | | | | | |
| oatcakes_spread_soya_dunno_med | 17027 | 30 | 17024 | 70 | | | | | | |
| oatcakes_spread_soya_dunno_thick | 17027 | 30 | 17024 | 70 | | | | | | |
| oatcakes_spread_soya_dunno_thin | 17027 | 30 | 17024 | 70 | | | | | | |
| oatcakes_spread_soya_fat_med | 17024 | 100 | | | | | | | | |
| oatcakes_spread_soya_fat_thick | 17024 | 100 | | | | | | | | |
| oatcakes_spread_soya_fat_thin | 17024 | 100 | | | | | | | | |
| oatcakes_spread_soya_lowfat_med | 17027 | 100 | | | | | | | | |
| oatcakes_spread_soya_lowfat_thick | 17027 | 100 | | | | | | | | |
| oatcakes_spread_soya_lowfat_thin | 17027 | 100 | | | | | | | | |
| oatcakes_spread_soya_vlowfat_med | 17029 | 100 | | | | | | | | |
| oatcakes_spread_soya_vlowfat_thick | 17029 | 100 | | | | | | | | |
| oatcakes_spread_soya_vlowfat_thin | 17029 | 100 | | | | | | | | |
| pancake_blini | 148 | 50 | 11544 | 50 | | | | | | |
| pancake_crepe | 11347 | 50 | 11604 | 50 | | | | | | |
| pasta_brown | 11455 | 100 | | | | | | | | |
| pasta_white | 11450 | 25 | 11453 | 50 | 28 | 25 | | | | |
| pesto | 15240 | 100 | | | | | | | | |
| pizza | 15252 | 100 | | | | | | | | |
| pnutbutter_nutella | 14876 | 70 | 17070 | 30 | | | | | | |
| potato_boil | 13002 | 33.4 | 13004 | 33.3 | 13013 | 33.3 | | | | |
| potato_boil_marg | 17485 | 50 | 12258 | 50 | | | | | | |
| potato_fried | 13411 | 75 | 13016 | 25 | | | | | | |
| potato_mashed | 13402 | 50 | 13015 | 50 | | | | | | |
| poultry_friedbcrumb_noskin | 19122 | 67 | 19124 | 33 | | | | | | |
| poultry_friedbcrumb_withskin | 19122 | 65 | 19124 | 30 | 18332 | 5 | | | | |
| poultry_noskin | 18331 | 90 | 18361 | 10 | | | | | | |
| poultry_withskin | 18341 | 90 | 463 | 10 | | | | | | |
| rice_brown | 11443 | 100 | | | | | | | | |
| rice_white | 11446 | 100 | | | | | | | | |
| salad_dressing | 17509 | 100 | | | | | | | | |
| sauce_brown | 1165 | 50 | 17289 | 50 | | | | | | |
| sauce_cheese | 17522 | 100 | | | | | | | | |
| sauce_gravy | 17311 | 100 | | | | | | | | |
| sauce_ketchup | 17513 | 100 | | | | | | | | |
| sauce_tomato | 17333 | 50 | 17516 | 50 | | | | | | |
| sauce_white | 17528 | 100 | | | | | | | | |
| scone | 11592 | 50 | 11543 | 50 | | | | | | |
| single_crust | 11583 | **40** | 11585 | **50** | 11587 | **10** | | | | |
| smoothie_dairy | 12193 | 100 | | | | | | | | |
| smoothie_fruit | 14271 | 55 | 14045 | 20 | 14244 | 12.5 | 14260 | 12.5 | | |
| snack_cheesybis | 11510 | 100 | | | | | | | | |
| snack_crisps | 17495 | 50 | 17142 | 25 | 17497 | 25 | | | | |
| snack_olives | 14173 | 100 | | | | | | | | |
| snack_saltednuts | 14823 | 25 | 14840 | 25 | 14812 | 50 | | | | |
| snack_saltedpeanuts | 14834 | 100 | | | | | | | | |
| snack_savourybis | 11511 | 50 | 11510 | 50 | | | | | | |
| snack_seeds | 14844 | 50 | 14845 | 50 | | | | | | |
| snack_svyother | 14827 | 50 | 17495 | 50 | | | | | | |
| snack_swtother | 17547 | 33.4 | 11480 | 33.3 | 11508 | 33.3 | | | | |
| snack_unsaltednuts | 14811 | 33.4 | 14850 | 33.3 | 14870 | 33.3 | | | | |
| snack_unsaltedpeanuts | 14831 | 100 | | | | | | | | |
| snackpot | 17508 | 70 | 11057 | 30 | | | | | | |
| soup_canned_fish | 17276 | 85 | 16256 | 5 | 16068 | 5 | 12332 | 5 | | |
| soup_canned_meat | 17250 | 33.4 | 17271 | 33.3 | 17272 | 33.3 | | | | |
| soup_canned_other | 17256 | 50 | 17275 | 50 | | | | | | |
| soup_canned_pasta | 17542 | 100 | | | | | | | | |
| soup_canned_pulse | 17264 | 100 | | | | | | | | |
| soup_canned_unanswered | 17264 | 33.3 | 17250 | 33.3 | 17283 | 33.4 | | | | |
| soup_canned_veg | 17270 | 25 | 17278 | 25 | 17283 | 25 | 17276 | 25 | | |
| soup_homemade_fish | 17276 | 85 | 16256 | 5 | 16068 | 5 | 12332 | 5 | | |
| soup_homemade_meat | 17250 | 33.4 | 17271 | 33.3 | 17272 | 33.3 | | | | |
| soup_homemade_other | 17256 | 50 | 17275 | 50 | | | | | | |
| soup_homemade_pasta | 17542 | 100 | | | | | | | | |
| soup_homemade_pulse | 17264 | 100 | | | | | | | | |
| soup_homemade_unanswered | 17264 | 33.3 | 17250 | 33.3 | 17283 | 33.4 | | | | |
| soup_homemade_veg | 17270 | 25 | 17278 | 25 | 17283 | 25 | 17276 | 25 | | |
| soup_powder | 17508 | 100 | | | | | | | | |
| spongepuds | 164 | 100 | | | | | | | | |
| spreadsauce_other | 17313 | 50 | 17364 | 25 | 17365 | 25 | | | | |
| sushi | 11446 | 54 | 13340 | 1 | 16207 | 14 | 14037 | 9 | 13233 | 22 |
| sweets | 17101 | 25 | 17120 | 25 | 17104 | 25 | 17117 | 25 | | |
| sweets_diet | 17101 | 50 | 17104 | 50 | | | | | | |
| tea_artf_swt | | | | | | | | | | |
| tea_black | 17165 | 100 | | | | | | | | |
| tea_black_decaf | 17165 | 100 | | | | | | | | |
| tea_green | 17171 | 100 | | | | | | | | |
| tea_herbal | 17172 | 100 | | | | | | | | |
| tea_other | 17170 | 100 | | | | | | | | |
| tea_rooibos | 17165 | 100 | | | | | | | | |
| tea_sugar | 17063 | 100 | | | | | | | | |
| veg_avocado | 14037 | 100 | | | | | | | | |
| veg_bakedbeans | 13044 | 100 | | | | | | | | |
| veg_beetroot | 13166 | 50 | 13165 | 50 | | | | | | |

**Supplementary table 3**. Nutrient calculation in the previous version (McCance and Widdowson).

| | Food codes from McCance and Widdowson and the % used from each food code | | | | | | | | | |
|---|---|---|---|---|---|---|---|---|---|---|
| **Item** | **Code1** | **% from code 1** | **Code2** | **% from code 2** | **Code3** | **% from code 3** | **Code4** | **% from code 4** | **Code5** | **% from code 5** |
| veg_broadbeans | 13065 | 100 | | | | | | | | |
| veg_broccoli | 13171 | 100 | | | | | | | | |
| veg_butternut | 13356 | 100 | | | | | | | | |
| veg_cabbagekale | 13349 | 33.4 | 13184 | 33.3 | 13235 | 33.3 | | | | |
| veg_carrots | 13201 | 53 | 13204 | 47 | | | | | | |
| veg_cauli | 13216 | 100 | | | | | | | | |
| veg_celery | 13221 | 100 | | | | | | | | |
| veg_courgette | 13231 | 100 | | | | | | | | |
| veg_cucumber | 13233 | 100 | | | | | | | | |
| veg_garlic | 13244 | 100 | | | | | | | | |
| veg_greenbeans | 13082 | 50 | 13113 | 50 | | | | | | |
| veg_leek | 13264 | 100 | | | | | | | | |
| veg_lettuce | 13266 | 100 | | | | | | | | |
| veg_mixed | 13281 | 100 | | | | | | | | |
| veg_mixtures | 13171 | 33.4 | 13201 | 33.3 | 13082 | 33.3 | | | | |
| veg_mushrooms | 13284 | 100 | | | | | | | | |
| veg_onion | 13304 | 100 | | | | | | | | |
| veg_other | 13123 | 25 | 13159 | 25 | 13327 | 25 | 13220 | 25 | | |
| veg_parsnip | 13313 | 100 | | | | | | | | |
| veg_peas | 13133 | 90 | 13135 | 10 | | | | | | |
| veg_pepper_bell | 13318 | 50 | 13320 | 50 | | | | | | |
| veg_pulses | 13092 | 25 | 13111 | 25 | 13078 | 25 | 13072 | 25 | | |
| veg_saladmayo | 15077 | 33.4 | 15078 | 33.3 | 15079 | 33.3 | | | | |
| veg_sidesalad | 13266 | 25 | 13384 | 50 | 13233 | 25 | | | | |
| veg_spinach | 13344 | 100 | | | | | | | | |
| veg_sprouts | 13178 | 100 | | | | | | | | |
| veg_sweetcorn | 13369 | 100 | | | | | | | | |
| veg_sweetpot | 13363 | 50 | 13364 | 50 | | | | | | |
| veg_tomato_fresh | 13384 | 100 | | | | | | | | |
| veg_tomato_tinned | 13387 | 100 | | | | | | | | |
| veg_turnip | 13390 | 50 | 13360 | 50 | | | | | | |
| veg_watercress | 13462 | 100 | | | | | | | | |
| vegalt_burger | 15330 | 100 | | | | | | | | |
| vegalt_other | 15213 | 50 | 15202 | 30 | 13281 | 20 | | | | |
| vegalt_quorn | 13328 | 100 | | | | | | | | |
| vegalt_tofu | 13119 | 80 | 15314 | 20 | | | | | | |
| yeast_extract | 17517 | 100 | | | | | | | | |
| yogurt_fullfat | 12184 | 40 | 12375 | 40 | 12376 | 20 | | | | |
| yogurt_lowfat | 12188 | 25 | 12189 | 25 | 12190 | 25 | 12382 | 25 | | |
| yorkshirepud | 11607 | 50 | 11360 | 50 | | | | | | |
